# Co-occurrence of neuropsychiatric symptoms in ADAMS, ADNI and NACC studies as assessed by Neuropsychiatric Inventory

**DOI:** 10.1101/2025.01.14.25320545

**Authors:** Timofey L. Galankin, Jina Swartz, Hans J. Moebius, Anton Y. Bespalov, the Alzheimer’s Disease Neuroimaging Initiative

## Abstract

**Objective:** Neuropsychiatric symptoms (NPS) are very common and associated with high levels of distress, both in dementia patients and their caregivers. Especially at more advanced dementia disease stages, NPS rarely occur in isolation, and the presence of two or more NPS may affect disease severity as well as the response to therapy. There is limited quantitative information on prevalence of specific symptom combinations in the general population, as well as in the populations recruited for symptom-specific investigations.

**Methods:** We performed cross-sectional analyses of publicly accessible Neuropsychiatric Inventory and Mini Mental State Examination (MMSE) data from three longitudinal studies (Aging, Demographics, and Memory Study (ADAMS), Alzheimer’s Disease Neuroimaging Initiative (ADNI) and the National Alzheimer’s Coordinating Center data (NACC)). Mean (with 95% confidence interval) prevalence was calculated for all possible pairs of symptoms (aberrant motor behavior, agitation/aggression, anxiety, apathy/indifference, appetite/eating changes, delusions; depression/dysphoria; disinhibition; elation/euphoria; hallucinations; irritability/lability and nighttime behavioral disturbances) in different MMSE strata. In addition, the conditional prevalence of one symptom given another symptom was provided for all possible combinations.

**Results:** In all three studies and MMSE strata, we observed every possible pair combination, from commonly recognized and discussed associations (e.g., hallucinations and delusions) to what might be seen as rather counter-intuitive patterns (e.g., apathy and agitation). Prevalence of symptom pairs cannot be readily predicted based on prevalence of individual symptoms. Presence of cognitive deficit and degree of cognitive impairment affected prevalence of all symptoms and symptom pairs, albeit to a different degree. For example, prevalence of the most common symptom, depression, in subjects without and with cognitive deficit, differed less than two-fold. In contrast, differences in the prevalence of psychotic symptoms (hallucinations and/or delusions) in subjects with and without cognitive deficit were much stronger (6- to 38-fold).

**Conclusions:** The present study illustrates that, while there is the possibility of any combination of neuropsychiatric symptoms presenting during the course of dementia, their co-occurrence cannot be readily predicted based on the prevalence of individual symptoms. Thus, our study results can serve as a source of reference information to inform the design and recruitment strategies for future clinical studies and epidemiological research on neuropsychiatric symptoms in people with dementia.

**Highlights:**

- *What is the primary question addressed by this study?—The question addressed by the study must limited to only one sentence*. There is very limited quantitative information on prevalence of neuropsychiatric symptom combinations despite the growing number of epidemiological and drug development studies in the field.
- *What is the main finding of this study?—The finding must be limited to two sentences*. All possible pair combinations frequently occur even in subjects with mild, minimal or no cognitive deficit in the general population, as well as in protocol-based dementia research studies. Co-occurrence of neuropsychiatric symptoms cannot be readily predicted based on the prevalence of individual symptoms.
- *What is the meaning of the finding?—The meaning of the finding must be limited to one sentence*. We provide reference information on neuropsychiatric symptom pair prevalence to inform the design and recruitment strategies for future clinical studies, as well as epidemiological research on neuropsychiatric symptoms.

## INTRODUCTION

During the natural course of dementia, a heterogeneous group of clinical phenomena is subjectively experienced by the patient and/or is observable by an examiner, caregiver or health professional, consisting of disturbed emotions, mood, perception, sleep patterns, behaviors and altered personality traits. These neuropsychiatric symptoms (NPS), as defined by the terminology most used in the US, or behavioral and psychological symptoms of dementia (BPSD), as designated by the International Psychogeriatric Association (IPA) ^1^, are very common and associated with high levels of distress, both in dementia patients and their caregivers.

The proportion of people developing NPS and the impact of NPS symptoms are almost ubiquitous in people with dementia, with the five-year period prevalence estimated to be as high as 97% in a population of elderly subjects with all-cause dementia ^2^. These non-cognitive symptoms are associated with poor outcomes in terms of function, quality of life, disease course, mortality and economic cost ^3^. Recent observation further associates p-Tau181 elevation with the emergence of one of the most frequent NPS in Alzheimer’s disease (AD), namely psychosis ^4^.

Expression of NPS depends on the disease severity stage and type of dementia ^5^. For example, agitation may be present in about 19% of subjects with a Clinical Dementia Rating (CDR) score of 0.5, reflective of mild cognitive impairment or questionable dementia, but it becomes more prevalent (up to 55%) at a CDR score of 3, indicative of severe cognitive impairment within the context of severe dementia. Delusions can be present in about 5% of patients with a CDR score of 0.5, but in 25% of patients with a CDR score of 3. As the disease progresses, both severity of the symptom and caregiver distress are increased.

In recent years, regulators have addressed the high medical need by being more permissive in their approvals of dementia therapies with syndromal and subsyndromal indication labels. Two drugs, pimavanserin (Nuplazid®) and brexpiprazole (Rexulti®) are currently approved in the US for treatment of hallucinations and delusions associated with Parkinson’s disease and agitation in AD ^3^, respectively. Although the underlying reasons are not the same, both drugs carry black box safety warnings, similar to all other members of the antipsychotic drug class, albeit for different mechanistic reasons. Antipsychotics are commonly used off-label for treatment of NPS despite significant risks of severe adverse effects, heightened morbidity and mortality as well as accelerated cognitive decline ^6–8^.

There are a number of advanced stage drug development programs focusing on novel therapies of NPS primarily in subjects with Alzheimer’s disease dementia ^9^. Positive outcomes of several of these efforts reveal not only useful clinical trial designs, outcome measures and analytic strategies, but also identify factors to be considered in developing safe and effective medications.

For example, there is a growing body of evidence that methylphenidate effectively reduces apathy in subjects with AD, but the effect size of methylphenidate is reduced in those with higher baseline anxiety and/or agitation ^10^. Further, novel agents such as pimavanserin may have no effect on agitation unless subjects also exhibit psychotic symptoms that are responsive to treatment with this drug ^11^.

It is well established that, especially at more advanced disease stages, NPS rarely occur in isolation. As a consequence of the use of the Neuropsychiatric Inventory (NPI) ^12^, a tool commonly utilized in clinical research, the presence of a broad range of NPS in the subject population, as well as efficacy of pharmacotherapies against these NPS, are reported for many clinical trials including those not employing the NPI as a primary study outcome measure ^13^.

The co-expression of different NPS has been the subject of numerous studies in an effort to understand better AD phenomenology and potentially identify further pathophysiological entities. Most of these studies aimed to identify clusters of symptoms using analytic tools such as principal component analysis. However, we have recently demonstrated ^14^ that such analysis risks revealing NPS clusters that are a statistical phenomenon rather than symptom associations that occur in real life (at least when studied using tools such as the NPI).

Using publicly accessible data from three large longitudinal studies (Aging, Demographics, and Memory Study (ADAMS), Alzheimer’s Disease Neuroimaging Initiative (ADNI) and the National Alzheimer’s Coordinating Center data (NACC)), the present study aims to address this apparent gap by reporting descriptive statistics of symptom pair prevalences that are simple, easy to describe, understand and employ (for example, to facilitate the planning of future clinical studies).

## METHODS

### Data sources

We were granted access to publicly accessible databases of three longitudinal studies: the Aging, Demographics, and Memory Study (ADAMS), Alzheimer’s Disease Neuroimaging Initiative (ADNI) and the National Alzheimer’s Coordinating Center data (NACC). We performed cross-sectional analyses of one visit with the maximum number of subjects having NPI and MMSE assessments for each study (Wave A for ADAMS, collected from August 2001 to January 2004; Month 12 for ADNI, collected from March 2011 to March 2020; Visit 1 for NACC, collected from June 2005 to November 2017).

### ADAMS dataset

The ADAMS ^15^ is a supplement to the Health and Retirement Study (HRS) that is sponsored by the National Institute on Aging (grant number NIA U01AG009740) and is conducted by the University of Michigan (https://hrs.isr.umich.edu/). The HRS is an ongoing biennial longitudinal survey of a nationally representative cohort of more than 20 000 US adults aged 51 and older who reside in the community and in nursing homes throughout the 48 contiguous United States. The ADAMS sample was a stratified random subsample of 1770 individuals aged 71 years and older from five cognitive strata based on scores for the 35-point HRS cognitive scale or proxy assessments of cognition from the 2000 or 2002 waves of the HRS. There were no specific eligibility criteria used and, by applying sampling weights, the estimates derived from ADAMS data can reflect the entire population aged 70 years and older in July 2001. Further details of the ADAMS sample design and selection procedures are described elsewhere ^16^.

All study procedures were approved by the Institutional Review Boards at Duke University Medical Center and the University of Michigan and informed consent was obtained from study participants or their surrogates.

The Wave A assessments were completed for 856 subjects, representing a 56% response rate among non-deceased sample members ^16^ (see Supplement section 1). Both NPI (original version) and Mini Mental Status Examination (MMSE) assessments were completed for 799 subjects of Wave A and were included in the present analysis.

### ADNI dataset

Data used in the preparation of this article were obtained from the Alzheimer’s Disease Neuroimaging Initiative (ADNI) database (adni.loni.usc.edu). The ADNI was launched in 2003 as a public-private partnership, led by Principal Investigator Michael W. Weiner, MD. The primary goal of ADNI has been to test whether serial magnetic resonance imaging (MRI), positron emission tomography (PET), other biological markers and clinical and neuropsychological assessment can be combined to measure the progression of mild cognitive impairment (MCI) and early Alzheimer’s disease (AD).

The ADNI study design is described as non-randomized natural history non-treatment study. Participants were enrolled in 54 American and five Canadian acquisition sites using centralized eligibility criteria that can be found on the ADNI website in the study protocols. In short, enrolled subjects were between 55 and 90 (inclusively) years of age, had a reliable study partner able to provide an independent evaluation of functioning, spoke either English or Spanish, were willing and able to undergo all test procedures including neuroimaging and lumbar puncture and had a baseline MMSE score of ≥20.

Written informed consent for the study was obtained from all participants and/or authorized representatives and the study partners before protocol-specific procedures were carried out.

ADNI has several recruitment protocols, all of which are still ongoing: ADNI1 was started in 2004, ADNIGO in 2009, ADNI2 in 2012 and ADNI3 in 2016. Starting from ADNI2, the NPI-12 questionnaire was introduced and is being applied on an annual basis. Participants may switch from one protocol to another while retaining the same ID number and the structure of subsequent annual visits. We used the data extracted on 16 May 2020. Both NPI-12 and MMSE assessments were completed for 889 subjects (across all protocols) at Month 12 (see Supplement section 1) and were included in the present analysis.

### NACC dataset

The NACC was established in 1999 and captures data of participants from 37 Alzheimer’s Disease Research Centers (ADRCs) across the USA that are supported by the National Institute on Aging ^17,18^. The NACC database is funded by NIA/NIH Grant U24 AG072122. The present study used publicly available data from the NACC Uniform Data Set (UDS)(https://naccdata.org/).

Each center enrolls its participants according to its own protocol — e.g., clinician referral, self-referral by participants or family members, active recruitment in community organizations, etc. Most centers also enroll volunteers with normal cognition. NACC participants are not a statistically based sample of the US population — with or without dementia. Rather, they are best regarded as a referral-based or volunteer case series. Some ADRCs require that participants agree to autopsy before being accepted for UDS participation; this may impose further selection pressures on the makeup of the NACC sample. Informants are required for all participants, including non-demented controls. Participants enrolled at each ADRC provide written consent as part of the Institutional Review Board-approved protocol at that center. This consent covers both the data collection procedures required by the respective center as well as the inclusion of the participant’s data in the larger NACC UDS database.

The NACC UDS is a comprehensive data repository for research on neurodegenerative disorders, including AD. The UDS contains longitudinal data that have been collected since 2005. Data elements and collection methods have been described previously ^17,18^. The NACC UDS includes neuropsychological, behavioral, medical and health history data that is used to accurately diagnose neurodegenerative disease and track its course. One of the variables that NACC tracks is the NPI-Q, a questionnaire version of the NPI (please see below for further information). For Visit 1, NACC UDS contains data from 41 170 subjects of which 26 171 completed assessments for both the NPI (original version) and MMSE and were included in the present analysis (see Supplement section 1).

### Neuropsychiatric Inventory (NPI)

The NPI is an instrument widely used to study neuropsychiatric symptoms in dementia, with psychometric properties previously reported ^12,19,20^. Three versions of the NPI were used in our analysis: NPI-10 for the ADAMS study, NPI-12 for ADNI and NPI-Q for NACC. In its initial form (NPI-10) ^12^, the NPI is an informant-based interview covering information on symptoms provided by informed caregivers during the past month across 10 domains – delusions, hallucinations, agitation, depression, anxiety, elation, apathy, disinhibition, irritability and aberrant motor behaviors – using a structured interview with a knowledgeable informant. For each symptom reported, information is obtained about frequency (on a scale from 0 to 4) and severity (on a scale from 0 to 3). NPI-12 ^19^ is a modification of NPI-10 with two new symptoms added: appetite/eating changes and nighttime behavioral disturbances. NPI-Q (NPI-Questionnaire) ^20^ is a self-administered questionnaire completed by informants that uses only the screening question of NPI-12 without sub questions. NPI-Q uses only the severity rating (not frequency). All three versions of the NPI assess and assign a “caregiver distress” score, but this is not used in analysis of symptom prevalence.

In addition to the 12 original NPI items, we introduced a composite item comprising both psychotic symptoms (hallucinations and delusions) for our analysis. Severity score for psychotic symptoms was entered as the greater value of severity scores for delusions or hallucinations. Where applicable, the product score (see Statistical analysis below) for psychotic symptoms is entered in the same way.

### Mini Mental State Examination (MMSE)

The MMSE is a 30-point questionnaire test used to briefly assess cognitive status and is commonly used in clinical medical practice to screen for cognitive deficits. It is also routinely used in the context of broadly characterizing the degree of cognitive impairment within the dementia spectrum. In so doing, it is used to estimate the severity of cognitive deficit at a specific time and to follow the course of cognitive changes in an individual over time. The MMSE scale comprises 11 questions or simple tasks concerning orientation, memory, attention and language to evaluate the patient’s cognitive state. The maximum total score is 30 ^21^. Standard cutoff points on the MMSE were used to classify the degree of cognitive deficit into no observable deficit (27-30), mild (20-26), moderate (10-19) or severe (0-9) cognitive deficit. These categories of cognitive impairment all imply the severity of the cognitive deficit, in the context of dementia.

### Statistical analysis

Prevalence of symptoms and symptom pairs was calculated as a percentage with Clopper-Pearson exact 95% binomial proportion confidence intervals estimated using “binom.test()” function in the R “base” package ^22^. Conditional prevalence of one symptom in a pair given another (S1|S2) and vice versa (S2|S1) was calculated as a percentage with 95% confidence intervals derived via bootstrapping procedure with 1000 samples. For the ADAMS data, in addition to these sample-based estimates, we derived population-based estimates using survey weights, survey clusters and survey strata with the help of the R “survey” package that was used to derive survey proportion and survey mean, and the R “jtools” package that was used to derive survey standard deviation ^23,24^. The 95% confidence intervals for survey proportions were based on the incomplete beta distribution similar to Clopper-Pearson exact confidence intervals ^25^. Please refer to Supplement section 1 for additional information about data management. Confidence intervals were used for descriptive purposes only, no inferences were made and no adjustments for multiplicity used.

Analysis was performed for three types of symptoms: NPS with severity scores greater than 0, NPS with severity scores greater than 1 (clinically significant symptoms), as well as NPS with product scores (severity x frequency) equal to or greater than 4 (another definition for clinically significant symptoms, product scores can be calculated only for ADAMS and ADNI studies).

Based on MMSE stratification, the following analysis sets were used: i) all subjects, ii) subjects with no cognitive deficit (MMSE >26), ii) subjects with cognitive deficit (MMSE ≤26), iii) subjects with mild cognitive deficit (MMSE score greater than 19 but less than or equal to 26), iv) subjects with moderate cognitive deficit (MMSE score greater than 9 but less than or equal to 19) and v) subjects with severe cognitive deficit (MMSE score is less than or equal to 9). The last three analysis sets were applied only to the NACC dataset, since ADNI and ADAMS studies did not have sufficient sample sizes.

The full analysis set from the NACC study was used to order symptoms and symptom pairs by their prevalence. This order was used as standard order throughout the article (i.e., was maintained for subsets of NACC data as well for presenting the data from the ADNI and ADAMS studies).

## RESULTS

### Demographic characteristics

Table 1 contains demographic characteristics of subjects meeting eligibility criteria of having completed MMSE and NPI assessments. Across all three studies (ADAMS, ADNI and NACC), there were data from a total of 27 859 subjects included in the analysis. Overall, 15 567 (55.9%) subjects were female, who were more frequently represented among those with no cognitive deficit (MMSE >26).

**Table 1.** Demographic characteristics.

| Study | ADAMS survey |  | ADAMS |  | ADNI |  | NACC |  |
| --- | --- | --- | --- | --- | --- | --- | --- | --- |
| MMSE stratum | > 26 | ≤ 26 | > 26 | ≤ 26 | > 26 | ≤ 26 | > 26 | ≤ 26 |
| MMSE mean score | 28.7 | 21.2 | 28.6 | 18.4 | 28.9 | 22.6 | 28.9 | 19.9 |
| N | 14,111,455 | 9,412,574 | 256 | 543 | 601 | 288 | 14,887 | 11,284 |
| Sex |  |  |  |  |  |  |  |  |
| Female | 9,242,112 (65.5) | 4,899,063 (52.0) | 150 (58.6) | 306 (56.4) | 280 (46.6) | 126 (43.8) | 8,783 (59.0) | 5,922 (52.5) |
| Male | 4,869,343 (34.5) | 4,513,511 (48.0) | 106 (41.4) | 237 (43.6) | 320 (53.2) | 162 (56.2) | 6,104 (41.0) | 5,362 (47.5) |
| Unknown | 0 ( 0.0) | 0 ( 0.0) | 0 ( 0.0) | 0 ( 0.0) | 1 ( 0.2) | 0 ( 0.0) | 0 ( 0.0) | 0 ( 0.0) |
| Race |  |  |  |  |  |  |  |  |
| White or Caucasian | 13,575,430 (96.2) | 7,614,460 (80.9) | 243 (94.9) | 376 (69.2) | 557 (92.7) | 265 (92.0) | 12,681 (85.2) | 8,897 (78.8) |
| Black or African American | 378,465 ( 2.7) | 1,341,768 (14.3) | 10 ( 3.9) | 138 (25.4) | 20 ( 3.3) | 11 ( 3.8) | 1,626 (10.9) | 1,582 (14.0) |
| Other | 157,560 ( 1.1) | 456,346 ( 4.8) | 3 ( 1.2) | 29 ( 5.3) | 10 ( 1.7) | 5 ( 1.7) | 159 ( 1.1) | 392 ( 3.5) |
| Asian | 0 ( 0.0) | 0 ( 0.0) | 0 ( 0.0) | 0 ( 0.0) | 7 ( 1.2) | 7 ( 2.4) | 254 ( 1.7) | 190 ( 1.7) |
| American Indian or Alaskan Native | 0 ( 0.0) | 0 ( 0.0) | 0 ( 0.0) | 0 ( 0.0) | 2 ( 0.3) | 0 ( 0.0) | 103 ( 0.7) | 130 ( 1.2) |
| Unknown | 0 ( 0.0) | 0 ( 0.0) | 0 ( 0.0) | 0 ( 0.0) | 4 ( 0.7) | 0 ( 0.0) | 53 ( 0.4) | 78 ( 0.7) |
| Native Hawaiian or Other Pacific Islander | 0 ( 0.0) | 0 ( 0.0) | 0 ( 0.0) | 0 ( 0.0) | 1 ( 0.2) | 0 ( 0.0) | 11 ( 0.1) | 15 ( 0.1) |
| Ethnicity |  |  |  |  |  |  |  |  |
| Not Hispanic or Latino | 13,610,899 (96.5) | 8,659,025 (92.0) | 242 (94.5) | 475 (87.5) | 572 (95.2) | 279 (96.9) | 13,834 (92.9) | 9,911 (87.8) |
| Hispanic or Latino | 500,556 ( 3.5) | 753,549 ( 8.0) | 14 ( 5.5) | 68 (12.5) | 25 ( 4.2) | 8 ( 2.8) | 998 ( 6.7) | 1,336 (11.8) |
| Unknown | 0 ( 0.0) | 0 ( 0.0) | 0 ( 0.0) | 0 ( 0.0) | 4 ( 0.7) | 1 ( 0.3) | 55 ( 0.4) | 37 ( 0.3) |
| Age group, years |  |  |  |  |  |  |  |  |
| > 75 | 8,279,895 (58.7) | 6,889,093 (73.2) | 166 (64.8) | 431 (79.4) | 197 (32.8) | 144 (50.0) | 4,840 (32.5) | 5,386 (47.7) |
| > 65 - ≤ 75 | 5,831,560 (41.3) | 2,523,481 (26.8) | 90 (35.2) | 112 (20.6) | 309 (51.4) | 105 (36.5) | 5,758 (38.7) | 3,517 (31.2) |
| > 40 - ≤ 65 | 0 ( 0.0) | 0 ( 0.0) | 0 ( 0.0) | 0 ( 0.0) | 93 (15.5) | 39 (13.5) | 4,151 (27.9) | 2,343 (20.8) |
| ≥ 18 - ≤ 40 | 0 ( 0.0) | 0 ( 0.0) | 0 ( 0.0) | 0 ( 0.0) | 0 ( 0.0) | 0 ( 0.0) | 138 ( 0.9) | 38 ( 0.3) |
| Unknown | 0 ( 0.0) | 0 ( 0.0) | 0 ( 0.0) | 0 ( 0.0) | 2 ( 0.3) | 0 ( 0.0) | 0 ( 0.0) | 0 ( 0.0) |

| Education group, years * |  |  |  |  |  |  |  |  |
| --- | --- | --- | --- | --- | --- | --- | --- | --- |
| ≤ 12 | 6,995,162 (49.6) | 7,647,878 (81.3) | 131 (51.2) | 461 (84.9) | 64 (10.6) | 49 (17.0) | 2,919 (19.6) | 4,706 (41.7) |
| > 12 - ≤ 18 | 7,116,293 (50.4) | 1,764,696 (18.7) | 125 (48.8) | 82 (15.1) | 392 (65.2) | 200 (69.4) | 9,926 (66.7) | 5,510 (48.8) |
| > 18 | 0 ( 0.0) | 0 ( 0.0) | 0 ( 0.0) | 0 ( 0.0) | 144 (24.0) | 39 (13.5) | 1,975 (13.3) | 966 ( 8.6) |
| Unknown | 0 ( 0.0) | 0 ( 0.0) | 0 ( 0.0) | 0 ( 0.0) | 1 ( 0.2) | 0 ( 0.0) | 67 ( 0.5) | 102 ( 0.9) |

Data are presented as absolute numbers of participants with percents from total population in brackets. Survey estimates for ADAMS are based on survey population weights. \* In general, 12 years is equivalent to high school or GRE (Graduate Record Examinations), 16 years to bachelor's degree, 18 years to master's degree, 20 years to doctorate.

Overall, 235 (0.8%) were American Indian or Alaskan Native; 458 (1.6%) Asian; 3 387 (12.2%) Black or African American; 27 (0.1%) Native Hawaiian or Other Pacific Islander; 23 019 (82.6%) White and 177 (2.6%) other/unknown race. In all three studies, whites were more frequently represented among those with no cognitive deficit (MMSE >26).

The mean [SD] years of education of all participants was 14.8 [3.7]. In all three studies, cognitively intact participants had, on average, more years of education. For example, in the NACC dataset, subjects with MMSE scores >26 had 15.7 [3.0] years of education on average, and subjects with MMSE scores ≤26 had 13.8 [3.9] years.

### Subjects without NPS

Subjects without any NPS were present across all three studies in every MMSE stratum, even among subjects with severe cognitive deficit (8.2% in the NACC dataset, see Supplemental Table 4).

We report symptom and symptom pair prevalences calculated for the entire population of eligible subjects including those with an NPI score of 0 for every symptom (note that, in some publications, prevalence is reported for populations with at least one NPS). The conversion to prevalence in a population with at least one NPS can be done with the help of Supplemental Table 4. For example, we report prevalence of depression in subjects without cognitive deficit to be 21.8% for the whole NACC dataset (Figure 1 and corresponding Supplemental Table 10). Prevalence of depression in subjects without cognitive deficit who had at least one NPS is then equal to (21.8% * 14 887) / (14 887 – 8 008) = 47.2% (14 887 and 8 008 are the total number of participants and the number of participants without NPS in the MMSE >26 stratum, respectively, taken from Supplemental Table 4).

**Figure 1.**
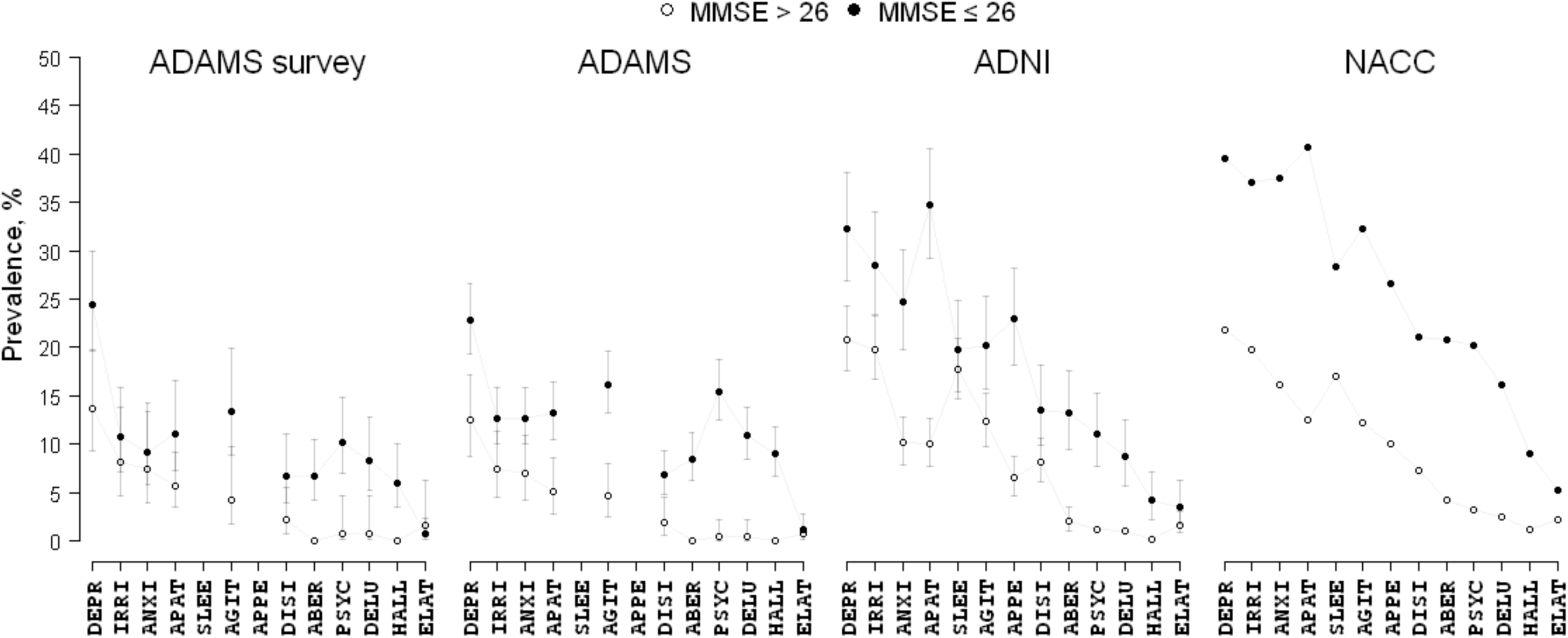
Prevalence of neuropsychiatric symptoms with the NPI severity score > 0 in subjects with MMSE total score >26 (open circles) or ≤26 (filled circles). Data are presented as prevalence (%) with 95% confidence intervals. Plotted values can be found in Supplemental Table 10. Survey estimates for ADAMS are based on survey population weights. ABER, Aberrant motor behavior; AGIT, Agitation/aggression; ANXI, Anxiety; APAT, Apathy/indifference; APPE, Appetite/eating changes; DELU, Delusions; DEPR, Depression/dysphoria; DISI, Disinhibition; ELAT, Elation/euphoria; HALU, Hallucinations; IRRI, Irritability/lability; PSYC, Psychotic symptoms (hallucinations and/or delusions); SLEE, nighttime behavioral disturbances.

The note above does not apply to conditional prevalence since conditionality implies the existence of at least two NPS.

### Prevalence of individual symptoms

Figure 1 presents the prevalence of NPS with severity score >0 for participants with and without cognitive deficit (see Supplemental Table 10 for the original values). Presence of cognitive deficit affected prevalence of all symptoms, albeit to a different degree. For example, prevalence of the most common symptom, depression, in subjects without and with cognitive deficit, differed less than two-fold (13.7% vs 24.4% in ADAMS survey population estimates, 12.5% versus 22.8% in ADAMS sample estimates, 20.8% vs 32.3% in ADNI and 21.8% versus 39.6% in NACC, respectively). In contrast, differences in the prevalence of psychotic symptoms (hallucinations and/or delusions) in subjects without and with cognitive deficit were much stronger: 15-fold in ADAMS survey population estimates (0.7% versus 10.2%), 38-fold in ADAMS sample estimates (0.4% versus 15.5%), 9-fold in ADNI (1.2% versus 11.1%) and 6-fold in NACC (3.2% versus 20.3%, respectively). Prevalence of NPS in the ADAMS dataset (both survey- and sample-based estimates) was lower than in the ADNI and NACC studies.

### Prevalence of symptom pairs

Table 2 contains estimated prevalence of pairs of symptoms with NPI severity score >0 in subjects with and without cognitive deficit (please refer to Supplemental Table 18 for prevalence of NPS pairs with severity score >1 and Supplemental Table 21 for prevalence of NPS with NPI product score ≥4). Overall, similar to what is described above for single symptoms, prevalence of most symptom pairs in the ADAMS dataset was lower than in the ADNI and NACC studies for both MMSE strata (≤26 and >26).

**Table 2.** Prevalence of pairs of neuropsychiatric symptoms with the NPI severity score > 0.

| Study |  | ADAMS survey |  | ADAMS |  | ADNI |  | NACC |  |
| --- | --- | --- | --- | --- | --- | --- | --- | --- | --- |
| MMSE stratum |  | > 26 | ≤ 26 | > 26 | ≤ 26 | > 26 | ≤ 26 | >26 | ≤26 |
| DEPR | IRRI | 3.9 (1.1-9.3) | 7.1 (3.3-13.0) | 3.1 (1.4-6.1) | 6.4 (4.5- 8.9) | 8.5 (6.4-11.0) | 15.3 (11.3-20.0) | 9.7 (9.2-10.2) | 20.1 (19.3-20.8) |
| DEPR | ANXI | 4.2 (1.4-9.6) | 6.1 (3.2-10.2) | 3.9 (1.9-7.1) | 7.9 (5.8-10.5) | 6.0 (4.2- 8.2) | 12.2 ( 8.6-16.5) | 9.6 (9.2-10.1) | 22.2 (21.5-23.0) |
| DEPR | APAT | 3.4 (1.6-6.1) | 8.4 (4.8-13.4) | 2.3 (0.9-5.0) | 8.8 (6.6-11.5) | 6.0 (4.2- 8.2) | 16.7 (12.6-21.5) | 7.5 (7.1- 7.9) | 22.0 (21.3-22.8) |
| DEPR | SLEE | NA | NA | NA | NA | 7.3 (5.4- 9.7) | 10.4 ( 7.1-14.5) | 7.9 (7.4- 8.3) | 15.2 (14.5-15.9) |
| DEPR | AGIT | 2.9 (0.7-7.8) | 8.7 (4.6-14.5) | 2.7 (1.1-5.6) | 7.6 (5.5-10.1) | 5.5 (3.8- 7.6) | 10.1 ( 6.8-14.1) | 6.3 (5.9- 6.7) | 17.5 (16.8-18.2) |
| DEPR | APPE | NA | NA | NA | NA | 3.3 (2.0- 5.1) | 11.5 ( 8.0-15.7) | 4.8 (4.5- 5.2) | 13.9 (13.2-14.5) |
| DEPR | DISI | 1.1 (0.1-5.0) | 3.6 (1.3- 8.0) | 1.2 (0.2-3.4) | 3.5 (2.1- 5.4) | 4.0 (2.6- 5.9) | 7.6 ( 4.8-11.3) | 3.6 (3.3- 4.0) | 10.7 (10.1-11.3) |
| DEPR | ABER | 0.0 (0.0- NE) | 3.4 (1.2- 7.3) | 0.0 (0.0-1.4) | 2.8 (1.6- 4.5) | 0.8 (0.3- 1.9) | 8.7 ( 5.7-12.5) | 2.3 (2.0- 2.5) | 10.8 (10.2-11.4) |
| DEPR | PSYC | 0.0 (0.0- NE) | 5.8 (3.0-10.2) | 0.0 (0.0-1.4) | 6.3 (4.4- 8.6) | 1.0 (0.4- 2.2) | 5.9 ( 3.5- 9.3) | 2.0 (1.8- 2.2) | 11.3 (10.7-11.9) |
| DEPR | DELU | 0.0 (0.0- NE) | 5.0 (2.2- 9.4) | 0.0 (0.0-1.4) | 4.2 (2.7- 6.3) | 1.0 (0.4- 2.2) | 5.2 ( 2.9- 8.4) | 1.6 (1.4- 1.8) | 9.3 ( 8.7- 9.8) |
| DEPR | HALL | 0.0 (0.0- NE) | 3.0 (1.3- 5.8) | 0.0 (0.0-1.4) | 4.1 (2.6- 6.1) | 0.0 (0.0- 0.6) | 2.1 ( 0.8- 4.5) | 0.7 (0.6- 0.9) | 4.9 ( 4.5- 5.3) |
| DEPR | ELAT | 0.8 (0.0-4.5) | 0.0 (0.0- NE) | 0.4 (0.0-2.2) | 0.0 (0.0- 0.7) | 1.0 (0.4- 2.2) | 1.4 ( 0.4- 3.5) | 1.0 (0.8- 1.2) | 2.8 ( 2.5- 3.1) |
| IRRI | ANXI | 2.8 (0.7-7.4) | 2.1 (1.1- 3.7) | 2.7 (1.1-5.6) | 5.2 (3.5- 7.4) | 5.0 (3.4- 7.0) | 12.8 ( 9.2-17.3) | 7.7 (7.3- 8.1) | 20.2 (19.5-21.0) |
| IRRI | APAT | 1.3 (0.2-4.6) | 4.3 (1.5- 9.6) | 1.2 (0.2-3.4) | 4.6 (3.0- 6.7) | 5.7 (3.9- 7.8) | 15.3 (11.3-20.0) | 6.8 (6.4- 7.2) | 20.3 (19.6-21.1) |
| IRRI | SLEE | NA | NA | NA | NA | 6.0 (4.2- 8.2) | 8.0 ( 5.1-11.7) | 7.0 (6.6- 7.5) | 14.7 (14.0-15.4) |
| IRRI | AGIT | 3.2 (1.0-7.5) | 8.0 (4.6-12.8) | 3.5 (1.6-6.6) | 9.0 (6.8-11.8) | 8.7 (6.5-11.2) | 11.5 ( 8.0-15.7) | 8.6 (8.2- 9.1) | 22.3 (21.5-23.0) |
| IRRI | APPE | NA | NA | NA | NA | 2.8 (1.7- 4.5) | 8.7 ( 5.7-12.5) | 4.5 (4.1- 4.8) | 13.3 (12.7-14.0) |
| IRRI | DISI | 1.2 (0.1-4.5) | 3.9 (1.6- 7.7) | 0.8 (0.1-2.8) | 4.2 (2.7- 6.3) | 5.0 (3.4- 7.0) | 7.6 ( 4.8-11.3) | 4.8 (4.5- 5.2) | 13.6 (12.9-14.2) |
| IRRI | ABER | 0.0 (0.0- NE) | 2.7 (1.0- 5.7) | 0.0 (0.0-1.4) | 3.5 (2.1- 5.4) | 0.8 (0.3- 1.9) | 5.6 ( 3.2- 8.9) | 2.5 (2.3- 2.8) | 11.8 (11.3-12.5) |
| IRRI | PSYC | 0.0 (0.0- NE) | 4.3 (1.9- 8.3) | 0.0 (0.0-1.4) | 6.1 (4.2- 8.4) | 1.2 (0.5- 2.4) | 4.9 ( 2.7- 8.0) | 2.0 (1.8- 2.3) | 11.4 (10.8-12.0) |
| IRRI | DELU | 0.0 (0.0- NE) | 3.5 (1.3- 7.7) | 0.0 (0.0-1.4) | 4.1 (2.6- 6.1) | 1.0 (0.4- 2.2) | 4.2 ( 2.2- 7.2) | 1.7 (1.5- 1.9) | 9.6 ( 9.1-10.2) |
| IRRI | HALL | 0.0 (0.0- NE) | 2.4 (0.8- 5.7) | 0.0 (0.0-1.4) | 4.1 (2.6- 6.1) | 0.2 (0.0- 0.9) | 1.4 ( 0.4- 3.5) | 0.6 (0.5- 0.8) | 4.7 ( 4.3- 5.1) |
| IRRI | ELAT | 0.0 (0.0- NE) | 0.1 (0.0- 0.6) | 0.0 (0.0-1.4) | 0.4 (0.0- 1.3) | 1.0 (0.4- 2.2) | 1.4 ( 0.4- 3.5) | 1.3 (1.1- 1.5) | 3.2 ( 2.9- 3.6) |
| ANXI | APAT | 2.4 (0.6-5.9) | 3.1 (1.5- 5.7) | 2.0 (0.6-4.5) | 5.2 (3.5- 7.4) | 3.0 (1.8- 4.7) | 13.5 ( 9.8-18.0) | 5.3 (5.0- 5.7) | 20.5 (19.8-21.3) |
| ANXI | SLEE | NA | NA | NA | NA | 4.7 (3.1- 6.7) | 7.6 ( 4.8-11.3) | 6.2 (5.8- 6.6) | 14.9 (14.2-15.5) |
| ANXI | AGIT | 2.0 (0.4-5.9) | 2.5 (1.5- 4.0) | 2.0 (0.6-4.5) | 5.5 (3.8- 7.8) | 4.0 (2.6- 5.9) | 10.8 ( 7.4-14.9) | 4.9 (4.5- 5.2) | 17.5 (16.8-18.2) |
| ANXI | APPE | NA | NA | NA | NA | 2.0 (1.0- 3.5) | 6.9 ( 4.3-10.5) | 3.7 (3.4- 4.0) | 13.4 (12.8-14.1) |
| ANXI | DISI | 0.6 (0.0-3.3) | 1.9 (0.9- 3.5) | 0.4 (0.0-2.2) | 3.7 (2.3- 5.6) | 3.0 (1.8- 4.7) | 5.9 ( 3.5- 9.3) | 3.3 (3.0- 3.6) | 11.6 (11.0-12.2) |
| ANXI | ABER | 0.0 (0.0- NE) | 1.9 (0.9- 3.3) | 0.0 (0.0-1.4) | 3.5 (2.1- 5.4) | 1.0 (0.4- 2.2) | 5.9 ( 3.5- 9.3) | 2.1 (1.9- 2.4) | 12.5 (11.9-13.2) |
| ANXI | PSYC | 0.0 (0.0- NE) | 2.4 (1.4- 3.7) | 0.0 (0.0-1.4) | 5.0 (3.3- 7.2) | 0.5 (0.1- 1.5) | 5.6 ( 3.2- 8.9) | 1.6 (1.4- 1.8) | 11.5 (10.9-12.1) |
| ANXI | DELU | 0.0 (0.0- NE) | 1.7 (1.1- 2.6) | 0.0 (0.0-1.4) | 3.5 (2.1- 5.4) | 0.5 (0.1- 1.5) | 3.8 ( 1.9- 6.7) | 1.2 (1.1- 1.4) | 9.3 ( 8.8- 9.9) |
| ANXI | HALL | 0.0 (0.0- NE) | 1.7 (1.0- 2.8) | 0.0 (0.0-1.4) | 3.5 (2.1- 5.4) | 0.0 (0.0- 0.6) | 2.4 ( 1.0- 4.9) | 0.6 (0.5- 0.8) | 5.4 ( 5.0- 5.8) |
| ANXI | ELAT | 0.8 (0.0-4.3) | 0.0 (0.0- NE) | 0.4 (0.0-2.2) | 0.0 (0.0- 0.7) | 0.5 (0.1- 1.5) | 1.0 ( 0.2- 3.0) | 1.0 (0.8- 1.2) | 3.1 ( 2.8- 3.5) |
| APAT | SLEE | NA | NA | NA | NA | 3.0 (1.8- 4.7) | 9.7 ( 6.6-13.7) | 5.6 (5.2- 6.0) | 16.3 (15.6-17.0) |
| APAT | AGIT | 1.0 (0.1-3.4) | 5.6 (2.4-10.9) | 1.2 (0.2-3.4) | 5.2 (3.5- 7.4) | 3.8 (2.4- 5.7) | 10.4 ( 7.1-14.5) | 4.7 (4.4- 5.1) | 18.6 (17.9-19.3) |
| APAT | APPE | NA | NA | NA | NA | 2.2 (1.2- 3.7) | 12.5 ( 8.9-16.9) | 4.3 (3.9- 4.6) | 16.0 (15.3-16.7) |
| APAT | DISI | 0.9 (0.1-3.5) | 2.6 (0.7- 6.7) | 0.8 (0.1-2.8) | 2.9 (1.7- 4.7) | 2.8 (1.7- 4.5) | 8.7 ( 5.7-12.5) | 3.6 (3.3- 3.9) | 13.3 (12.7-13.9) |
| APAT | ABER | 0.0 (0.0- NE) | 3.3 (1.0- 7.9) | 0.0 (0.0-1.4) | 3.3 (2.0- 5.2) | 1.0 (0.4- 2.2) | 9.0 ( 6.0-12.9) | 2.4 (2.1- 2.6) | 13.4 (12.8-14.1) |
| APAT | PSYC | 0.0 (0.0- NE) | 4.1 (1.5- 8.8) | 0.0 (0.0-1.4) | 5.2 (3.5- 7.4) | 0.8 (0.3- 1.9) | 5.2 ( 2.9- 8.4) | 1.5 (1.3- 1.7) | 11.9 (11.3-12.6) |
| APAT | DELU | 0.0 (0.0- NE) | 3.5 (1.0- 8.4) | 0.0 (0.0-1.4) | 4.1 (2.6- 6.1) | 0.7 (0.2- 1.7) | 3.8 ( 1.9- 6.7) | 1.2 (1.0- 1.4) | 9.5 ( 9.0-10.1) |
| APAT | HALL | 0.0 (0.0- NE) | 1.3 (0.5- 2.6) | 0.0 (0.0-1.4) | 2.9 (1.7- 4.7) | 0.2 (0.0- 0.9) | 2.8 ( 1.2- 5.4) | 0.6 (0.5- 0.7) | 5.6 ( 5.2- 6.0) |
| APAT | ELAT | 1.5 (0.2-5.6) | 0.0 (0.0- 0.2) | 0.8 (0.1-2.8) | 0.2 (0.0- 1.0) | 0.8 (0.3- 1.9) | 1.0 ( 0.2- 3.0) | 1.1 (1.0- 1.3) | 3.3 ( 3.0- 3.7) |
| SLEE | AGIT | NA | NA | NA | NA | 3.8 (2.4- 5.7) | 5.2 ( 2.9- 8.4) | 4.8 (4.5- 5.2) | 13.3 (12.7-14.0) |
| SLEE | APPE | NA | NA | NA | NA | 2.2 (1.2- 3.7) | 6.9 ( 4.3-10.5) | 4.6 (4.3- 4.9) | 12.0 (11.4-12.6) |
| SLEE | DISI | NA | NA | NA | NA | 2.3 (1.3- 3.9) | 4.9 ( 2.7- 8.0) | 3.1 (2.8- 3.4) | 9.3 ( 8.7- 9.8) |
| SLEE | ABER | NA | NA | NA | NA | 0.5 (0.1- 1.5) | 3.8 ( 1.9- 6.7) | 2.1 (1.8- 2.3) | 9.7 ( 9.1-10.2) |
| SLEE | PSYC | NA | NA | NA | NA | 0.5 (0.1- 1.5) | 2.8 ( 1.2- 5.4) | 1.6 (1.4- 1.8) | 9.3 ( 8.7- 9.8) |
| SLEE | DELU | NA | NA | NA | NA | 0.3 (0.0- 1.2) | 2.1 ( 0.8- 4.5) | 1.2 (1.0- 1.4) | 7.1 ( 6.6- 7.5) |
| SLEE | HALL | NA | NA | NA | NA | 0.2 (0.0- 0.9) | 1.0 ( 0.2- 3.0) | 0.7 (0.6- 0.8) | 5.1 ( 4.7- 5.6) |
| SLEE | ELAT | NA | NA | NA | NA | 0.3 (0.0- 1.2) | 1.4 ( 0.4- 3.5) | 0.9 (0.8- 1.1) | 2.5 ( 2.2- 2.8) |
| AGIT | APPE | NA | NA | NA | NA | 2.2 (1.2- 3.7) | 8.0 ( 5.1-11.7) | 3.3 (3.0- 3.6) | 12.3 (11.7-12.9) |
| AGIT | DISI | 0.6 (0.0-3.3) | 3.9 (1.7- 7.6) | 0.4 (0.0-2.2) | 4.2 (2.7- 6.3) | 4.0 (2.6- 5.9) | 7.0 ( 4.3-10.6) | 3.9 (3.6- 4.2) | 12.8 (12.2-13.5) |
| AGIT | ABER | 0.0 (0.0- NE) | 3.9 (1.5- 8.1) | 0.0 (0.0-1.4) | 4.6 (3.0- 6.7) | 1.0 (0.4- 2.2) | 5.2 ( 2.9- 8.4) | 2.1 (1.9- 2.4) | 11.6 (11.0-12.2) |
| AGIT | PSYC | 0.0 (0.0- NE) | 5.4 (2.6- 9.6) | 0.0 (0.0-1.4) | 7.0 (5.0- 9.5) | 0.8 (0.3- 1.9) | 4.2 ( 2.2- 7.2) | 1.8 (1.5- 2.0) | 12.2 (11.6-12.8) |
| AGIT | DELU | 0.0 (0.0- NE) | 4.8 (2.1- 9.2) | 0.0 (0.0-1.4) | 5.5 (3.8- 7.8) | 0.8 (0.3- 1.9) | 3.5 ( 1.7- 6.3) | 1.6 (1.4- 1.8) | 10.3 ( 9.7-10.8) |
| AGIT | HALL | 0.0 (0.0- NE) | 2.3 (0.8- 5.2) | 0.0 (0.0-1.4) | 3.9 (2.4- 5.9) | 0.0 (0.0- 0.6) | 1.4 ( 0.4- 3.5) | 0.5 (0.4- 0.6) | 5.1 ( 4.7- 5.6) |
| AGIT | ELAT | 0.0 (0.0- NE) | 0.2 (0.0- 0.6) | 0.0 (0.0-1.4) | 0.6 (0.1- 1.6) | 0.7 (0.2- 1.7) | 2.1 ( 0.8- 4.5) | 1.2 (1.0- 1.4) | 3.1 ( 2.8- 3.5) |
| APPE | DISI | NA | NA | NA | NA | 1.3 (0.6- 2.6) | 4.9 ( 2.7- 8.0) | 2.6 (2.3- 2.8) | 9.3 ( 8.8- 9.9) |
| APPE | ABER | NA | NA | NA | NA | 0.3 (0.0- 1.2) | 4.9 ( 2.7- 8.0) | 1.7 (1.5- 2.0) | 9.4 ( 8.9- 9.9) |
| APPE | PSYC | NA | NA | NA | NA | 0.3 (0.0- 1.2) | 4.2 ( 2.2- 7.2) | 1.1 (1.0- 1.3) | 7.7 ( 7.3- 8.2) |
| APPE | DELU | NA | NA | NA | NA | 0.3 (0.0- 1.2) | 3.1 ( 1.4- 5.8) | 0.9 (0.7- 1.0) | 6.2 ( 5.8- 6.7) |
| APPE | HALL | NA | NA | NA | NA | 0.0 (0.0- 0.6) | 1.7 ( 0.6- 4.0) | 0.5 (0.4- 0.6) | 3.5 ( 3.2- 3.8) |
| APPE | ELAT | NA | NA | NA | NA | 0.5 (0.1- 1.5) | 1.4 ( 0.4- 3.5) | 0.9 (0.8- 1.1) | 2.7 ( 2.5- 3.1) |
| DISI | ABER | 0.0 (0.0- NE) | 2.0 (0.4- 6.0) | 0.0 (0.0-1.4) | 2.2 (1.1- 3.8) | 0.8 (0.3- 1.9) | 6.2 ( 3.7- 9.7) | 2.0 (1.8- 2.2) | 9.3 ( 8.8- 9.9) |
| DISI | PSYC | 0.0 (0.0- NE) | 4.4 (1.7- 9.2) | 0.0 (0.0-1.4) | 3.5 (2.1- 5.4) | 0.5 (0.1- 1.5) | 2.4 ( 1.0- 4.9) | 1.3 (1.1- 1.5) | 7.8 ( 7.3- 8.3) |
| DISI | DELU | 0.0 (0.0- NE) | 4.1 (1.5- 8.8) | 0.0 (0.0-1.4) | 2.9 (1.7- 4.7) | 0.5 (0.1- 1.5) | 2.1 ( 0.8- 4.5) | 1.1 (0.9- 1.3) | 6.7 ( 6.2- 7.1) |
| DISI | HALL | 0.0 (0.0- NE) | 2.7 (0.6- 7.1) | 0.0 (0.0-1.4) | 2.0 (1.0- 3.6) | 0.0 (0.0- 0.6) | 0.7 ( 0.1- 2.5) | 0.4 (0.3- 0.5) | 3.2 ( 2.9- 3.6) |
| DISI | ELAT | 0.0 (0.0- NE) | 0.0 (0.0- NA) | 0.0 (0.0-1.4) | 0.0 (0.0- 0.7) | 0.7 (0.2- 1.7) | 2.8 ( 1.2- 5.4) | 1.4 (1.2- 1.6) | 3.3 ( 3.0- 3.7) |
| ABER | PSYC | 0.0 (0.0- NE) | 4.2 (1.9- 7.8) | 0.0 (0.0-1.4) | 5.0 (3.3- 7.2) | 0.2 (0.0- 0.9) | 2.8 ( 1.2- 5.4) | 0.8 (0.7- 1.0) | 8.3 ( 7.7- 8.8) |
| ABER | DELU | 0.0 (0.0- NE) | 3.6 (1.5- 7.0) | 0.0 (0.0-1.4) | 3.5 (2.1- 5.4) | 0.0 (0.0- 0.6) | 2.4 ( 1.0- 4.9) | 0.7 (0.5- 0.8) | 6.8 ( 6.3- 7.2) |
| ABER | HALL | 0.0 (0.0- NE) | 1.8 (0.9- 3.2) | 0.0 (0.0-1.4) | 3.3 (2.0- 5.2) | 0.2 (0.0- 0.9) | 1.0 ( 0.2- 3.0) | 0.3 (0.3- 0.5) | 4.0 ( 3.7- 4.4) |
| ABER | ELAT | 0.0 (0.0- NE) | 0.6 (0.1- 2.1) | 0.0 (0.0-1.4) | 0.7 (0.2- 1.9) | 0.3 (0.0- 1.2) | 0.7 ( 0.1- 2.5) | 0.8 (0.7- 1.0) | 2.7 ( 2.4- 3.0) |
| PSYC | ELAT | 0.0 (0.0- NE) | 0.6 (0.1- 2.1) | 0.0 (0.0-1.4) | 0.9 (0.3- 2.1) | 0.2 (0.0- 0.9) | 1.0 ( 0.2- 3.0) | 0.4 (0.3- 0.6) | 2.1 ( 1.8- 2.3) |
| DELU | HALL | 0.0 (0.0- NE) | 4.0 (1.7- 8.0) | 0.0 (0.0-1.4) | 4.4 (2.9- 6.5) | 0.0 (0.0- 0.6) | 1.7 ( 0.6- 4.0) | 0.4 (0.3- 0.6) | 4.8 ( 4.5- 5.3) |
| DELU | ELAT | 0.0 (0.0- NE) | 0.4 (0.0- 2.2) | 0.0 (0.0-1.4) | 0.4 (0.0- 1.3) | 0.2 (0.0- 0.9) | 0.7 ( 0.1- 2.5) | 0.4 (0.3- 0.5) | 1.8 ( 1.6- 2.1) |
| HALL | ELAT | 0.0 (0.0- NE) | 0.6 (0.1- 2.1) | 0.0 (0.0-1.4) | 0.9 (0.3- 2.1) | 0.0 (0.0- 0.6) | 0.7 ( 0.1- 2.5) | 0.2 (0.1- 0.2) | 0.9 ( 0.7- 1.1) |

Displayed is prevalence (%) with 95% confidence intervals. Survey estimates for ADAMS are based on survey population weights.
NA, not applicable; NE, not estimable.
ABER, Aberrant motor behavior; AGIT, Agitation/aggression; ANXI, Anxiety; APAT, Apathy/indifference; APPE, Appetite/eating changes; DELU, Delusions; DEPR, Depression/dysphoria; DISI, Disinhibition; ELAT, Elation/euphoria; HALU, Hallucinations; IRRI, Irritability/lability; PSYC, Psychotic symptoms (hallucinations and/or delusions); SLEE, nighttime behavioral disturbances.

In ADAMS subjects without cognitive deficit, the five most prevalent symptom pairs estimated for survey population were: depression-anxiety (4.2%), depression-irritability (3.9%), depression-apathy (3.4%), irritability-agitation (3.2%) and depression-agitation (2.9%); the five most prevalent symptom pairs in the cognitive deficit stratum were: depression-agitation (8.7%), depression-apathy (8.4%), irritability-agitation (8.0%), depression-irritability (7.1%) and depression-anxiety (6.1%).

In NACC subjects without cognitive deficit, the five most prevalent symptom pairs were: depression-irritability (9.7%), depression-anxiety (9.6%), irritability-agitation (8.6%), depression-sleep (7.9%) and irritability-anxiety (7.7%). In NACC subjects with cognitive deficit, the five most prevalent symptom pairs were: irritability-agitation (22.3%), depression-anxiety (22.2%), depression-apathy (22%), anxiety-apathy (20.5%) and irritability-apathy (20.3%).

Taking into account only pairs of symptoms with severity score >1, the picture looks similar (Supplemental Table 18). For example, the five most prevalent symptom pairs in NACC subjects without cognitive deficit were: depression-anxiety (2.7%), irritability-agitation (2.6%), depression-irritability (2.2%), depression-apathy (2.1%) and irritability-anxiety (2.1%). In NACC subjects with cognitive deficit, the five most prevalent symptom pairs were: irritability-agitation (8.1%), depression-anxiety (7.3%), depression-apathy (7%), irritability-apathy (6.9%) and apathy-agitation (6.8%).

### Conditional prevalence of symptoms

Tables 3 and 4 present the conditional prevalence of symptoms with the NPI severity score >0 in NACC study participants with MMSE >26 and MMSE ≤26. A lower MMSE score is generally associated with lower conditional prevalence similar to that described above for prevalence. For example, conditional prevalence of depression varies from 46.4 (44.5-48.3) to 63.0 (58.2-67.6) in subjects without cognitive deficit and from 50.9 (48.7-53.0) to 59.4 (57.9-60.9) in subjects with cognitive deficit. Similarly, conditional prevalence of psychotic symptoms varies from 9.1 (8.1-10.1) to 20.7 (16.3-25.2) in subjects without cognitive deficit and from 28.5 (27.1-29.7) to 39.7 (37.7-41.7) in subjects with cognitive deficit.

**Table 3.** Conditional prevalence of neuropsychiatric symptoms (S1 given S2), NPI severity score > 0 and MMSE score > 26 (NACC study)

| S2<br>S1 | DEPR | IRRI | ANXI | APAT | SLEE | AGIT | APPE | DISI | ABER | PSYC | DELU | HALL | ELAT |
| --- | --- | --- | --- | --- | --- | --- | --- | --- | --- | --- | --- | --- | --- |
| DEPR |  | 49.1<br>(47.2-50.8) | 59.8<br>(57.9-61.8) | 59.6<br>(57.5-61.8) | 46.4<br>(44.5-48.3) | 51.2<br>(49.0-53.4) | 48.6<br>(46.0-51.1) | 50.0<br>(46.9-52.6) | 54.9<br>(51.1-58.8) | 62.0<br>(57.5-66.2) | 63.0<br>(58.2-67.6) | 62.1<br>(55.3-69.2) | 47.1<br>(41.5-52.6) |
| IRRI | 44.6<br>(42.7-46.3) |  | 47.7<br>(45.6-49.9) | 54.5<br>(52.3-56.6) | 41.6<br>(39.6-43.7) | 70.6<br>(68.4-72.6) | 44.8<br>(42.3-47.4) | 66.5<br>(63.7-69.3) | 61.2<br>(57.2-64.7) | 63.2<br>(59.0-67.4) | 67.3<br>(62.7-72.0) | 54.4<br>(46.5-61.9) | 62.7<br>(57.5-68.1) |
| ANXI | 44.3<br>(42.7-46.0) | 38.8<br>(37.1-40.6) |  | 42.6<br>(40.4-44.8) | 36.7<br>(34.8-38.7) | 39.7<br>(37.5-42.0) | 37.4<br>(34.9-39.9) | 45.4<br>(42.6-48.2) | 51.2<br>(47.3-55.0) | 49.2<br>(44.6-53.4) | 49.3<br>(44.3-54.3) | 56.2<br>(48.6-63.7) | 46.5<br>(41.0-51.8) |
| APAT | 34.3<br>(32.6-35.9) | 34.5<br>(32.8-36.1) | 33.1<br>(31.2-35.1) |  | 32.9<br>(31.2-34.5) | 38.8<br>(36.6-40.9) | 42.8<br>(40.3-45.3) | 49.5<br>(46.6-52.6) | 57.4<br>(53.3-61.0) | 47.1<br>(42.8-51.7) | 47.5<br>(42.5-52.9) | 52.1<br>(44.4-59.6) | 53.2<br>(48.0-59.3) |
| SLEE | 36.1<br>(34.4-37.7) | 35.6<br>(33.9-37.4) | 38.6<br>(36.7-40.5) | 44.5<br>(42.5-46.6) |  | 39.4<br>(37.2-41.7) | 46.1<br>(43.4-48.6) | 42.5<br>(39.7-45.3) | 49.6<br>(45.9-53.3) | 50.4<br>(46.0-54.6) | 48.0<br>(43.1-52.6) | 61.5<br>(53.7-69.1) | 44.9<br>(39.4-50.0) |
| AGIT | 28.7<br>(27.2-30.3) | 43.6<br>(41.8-45.3) | 30.1<br>(28.2-31.9) | 37.9<br>(35.7-40.0) | 28.4<br>(26.7-30.2) |  | 33.1<br>(30.5-35.5) | 53.8<br>(50.7-56.6) | 51.2<br>(47.2-55.4) | 54.8<br>(50.1-59.3) | 61.9<br>(56.9-67.2) | 42.6<br>(35.0-50.0) | 55.4<br>(50.0-60.6) |
| APPE | 22.2<br>(20.8-23.6) | 22.6<br>(21.1-24.1) | 23.1<br>(21.4-24.8) | 34.0<br>(32.0-36.1) | 27.1<br>(25.3-28.7) | 27.0<br>(25.0-29.0) |  | 35.4<br>(32.7-38.2) | 42.0<br>(38.1-46.0) | 35.7<br>(31.3-40.0) | 34.9<br>(30.0-39.9) | 41.4<br>(34.1-48.3) | 43.9<br>(38.5-49.3) |
| DISI | 16.7<br>(15.4-18.0) | 24.5<br>(23.0-25.9) | 20.5<br>(18.9-22.2) | 28.8<br>(26.7-30.8) | 18.3<br>(16.7-19.8) | 32.0<br>(29.8-34.2) | 25.9<br>(23.7-28.1) |  | 47.5<br>(43.3-51.4) | 39.7<br>(35.4-44.1) | 43.7<br>(38.7-48.7) | 35.5<br>(28.3-42.9) | 65.9<br>(60.5-71.2) |
| ABER | 10.5<br>(9.3-11.6) | 12.9<br>(11.7-14.1) | 13.2<br>(11.9-14.5) | 19.0<br>(17.2-20.9) | 12.2<br>(10.9-13.5) | 17.4<br>(15.7-19.1) | 17.5<br>(15.6-19.4) | 27.1<br>(24.5-29.7) |  | 25.8<br>(22.0-29.9) | 26.5<br>(22.3-31.0) | 30.2<br>(23.2-37.7) | 39.5<br>(34.2-45.2) |
| PSYC | 9.1<br>(8.1-10.1) | 10.2<br>(9.2-11.3) | 9.7<br>(8.6-10.9) | 12.0<br>(10.6-13.6) | 9.5<br>(8.4-10.7) | 14.3<br>(12.7-16.0) | 11.5<br>(9.9-13.2) | 17.4<br>(15.3-19.7) | 19.9<br>(16.7-23.0) |  | -- | -- | 20.7<br>(16.3-25.2) |
| DELU | 7.2<br>(6.4-8.1) | 8.5<br>(7.6-9.6) | 7.7<br>(6.6-8.7) | 9.5<br>(8.2-10.9) | 7.1<br>(6.1-8.1) | 12.7<br>(11.2-14.2) | 8.8<br>(7.3-10.3) | 15.0<br>(13.0-17.2) | 16.0<br>(13.2-18.8) | -- |  | 39.1<br>(31.8-46.5) | 17.2<br>(13.2-21.3) |
| HALL | 3.2<br>(2.7-3.8) | 3.1<br>(2.5-3.8) | 4.0<br>(3.2-4.8) | 4.7<br>(3.8-5.7) | 4.1<br>(3.4-4.9) | 4.0<br>(3.1-4.9) | 4.7<br>(3.7-5.7) | 5.5<br>(4.3-7.0) | 8.2<br>(6.1-10.4) | -- | 17.7<br>(14.1-21.3) |  | 7.3<br>(4.6-10.2) |
| ELAT | 4.6<br>(3.8-5.3) | 6.7<br>(5.8-7.6) | 6.1<br>(5.1-7.2) | 8.9<br>(7.7-10.4) | 5.6<br>(4.6-6.5) | 9.6<br>(8.2-10.9) | 9.3<br>(7.9-10.9) | 19.1<br>(16.9-21.6) | 20.0<br>(16.9-23.3) | 13.7<br>(10.6-16.8) | 14.5<br>(10.8-18.1) | 13.6<br>(8.6-18.8) |  |

Displayed is conditional prevalence (%) with 95% confidence intervals.
ABER, Aberrant motor behavior; AGIT, Agitation/aggression; ANXI, Anxiety; APAT, Apathy/indifference; APPE, Appetite/eating changes; DELU, Delusions; DEPR, Depression/dysphoria; DISI, Disinhibition; ELAT, Elation/euphoria; HALU, Hallucinations; IRRI, Irritability/lability; PSYC, Psychotic symptoms (hallucinations and/or delusions); SLEE, nighttime behavioral disturbances.

**Table 4.** Conditional prevalence of neuropsychiatric symptoms (S1 given S2), NPI severity score > 0 and MMSE score ≤ 26 (NACC study)

| S2<br>S1 | DEPR | IRRI | ANXI | APAT | SLEE | AGIT | APPE | DISI | ABER | PSYC | DELU | HALL | ELAT |
| --- | --- | --- | --- | --- | --- | --- | --- | --- | --- | --- | --- | --- | --- |
| DEPR |  | 54.1<br>(52.6-55.7) | 59.4<br>(57.9-60.9) | 54.1<br>(52.6-55.5) | 53.5<br>(51.8-55.2) | 54.3<br>(52.6-55.9) | 52.2<br>(50.5-54.1) | 50.9<br>(48.7-53.0) | 52.0<br>(50.0-54.0) | 55.6<br>(53.6-57.6) | 57.4<br>(55.1-59.7) | 55.0<br>(52.0-58.1) | 52.5<br>(48.5-56.5) |
| IRRI | 50.6<br>(49.2-52.1) |  | 54.1<br>(52.6-55.6) | 49.9<br>(48.5-51.3) | 51.7<br>(50.0-53.5) | 69.1<br>(67.6-70.6) | 50.3<br>(48.6-52.2) | 64.4<br>(62.5-66.3) | 57.0<br>(55.0-59.1) | 56.0<br>(54.0-58.1) | 59.5<br>(57.3-61.8) | 52.5<br>(49.4-55.7) | 61.4<br>(57.2-65.2) |
| ANXI | 56.2<br>(54.7-57.7) | 54.6<br>(53.2-56.1) |  | 50.4<br>(49.0-51.9) | 52.4<br>(50.8-54.1) | 54.3<br>(52.7-56.0) | 50.5<br>(48.9-52.3) | 55.2<br>(53.1-57.3) | 60.3<br>(58.3-62.4) | 56.9<br>(54.9-58.8) | 57.9<br>(55.5-60.2) | 60.0<br>(57.0-63.2) | 59.1<br>(55.2-63.4) |
| APAT | 55.7<br>(54.2-57.1) | 54.9<br>(53.4-56.3) | 54.9<br>(53.3-56.3) |  | 57.5 (55.7-59.3) | 57.7<br>(56.1-59.4) | 60.3<br>(58.6-62.1) | 63.2<br>(61.4-65.2) | 64.7<br>(62.8-66.6) | 58.9<br>(56.9-61.0) | 58.8<br>(56.5-61.2) | 62.5<br>(59.5-65.5) | 63.1<br>(59.4-67.2) |
| SLEE | 38.4<br>(36.9-39.6) | 39.6<br>(38.2-41.0) | 39.7<br>(38.3-41.1) | 40.0<br>(38.6-41.5) |  | 41.4<br>(39.9-43.0) | 45.3<br>(43.4-47.0) | 44.1<br>(42.2-46.0) | 46.6<br>(44.8-48.5) | 45.8<br>(43.8-47.8) | 43.7<br>(41.4-45.9) | 57.3<br>(54.3-60.5) | 46.6<br>(42.6-50.5) |
| AGIT | 44.1<br>(42.7-45.5) | 60.1<br>(58.6-61.5) | 46.7<br>(45.2-48.1) | 45.6<br>(44.2-47.1) | 46.9<br>(45.3-48.6) |  | 46.3<br>(44.5-48.0) | 61.0<br>(59.0-63.0) | 55.8<br>(53.7-57.8) | 60.0<br>(58.1-61.9) | 63.6<br>(61.3-65.7) | 57.3<br>(54.4-60.5) | 59.1<br>(55.0-63.1) |
| APPE | 35.0<br>(33.7-36.3) | 36.0<br>(34.6-37.5) | 35.8<br>(34.4-37.3) | 39.3<br>(37.9-40.7) | 42.3<br>(40.5-44.0) | 38.1<br>(36.5-39.7) |  | 44.3<br>(42.3-46.3) | 45.2<br>(43.2-47.1) | 38.2<br>(36.0-40.1) | 38.5<br>(36.1-40.5) | 38.9<br>(35.8-42.0) | 52.0<br>(47.9-56.1) |
| DISI | 27.0<br>(25.8-28.3) | 36.6<br>(35.2-37.9) | 31.0<br>(29.7-32.4) | 32.7<br>(31.4-34.0) | 32.7<br>(31.1-34.3) | 39.9<br>(38.5-41.4) | 35.1<br>(33.5-36.8) |  | 44.8<br>(42.9-46.7) | 38.4<br>(36.5-40.5) | 41.3<br>(39.0-43.6) | 35.9<br>(32.7-38.8) | 62.6<br>(58.7-66.8) |
| ABER | 27.3<br>(26.0-28.5) | 32.0<br>(30.6-33.5) | 33.5<br>(32.2-34.9) | 33.0<br>(31.8-34.3) | 34.1<br>(32.7-35.7) | 36.0<br>(34.5-37.5) | 35.4<br>(33.7-37.1) | 44.3<br>(42.2-46.1) |  | 40.7<br>(38.7-42.7) | 41.8<br>(39.4-44.1) | 44.7<br>(41.9-48.0) | 50.5<br>(46.7-54.7) |
| PSYC | 28.5<br>(27.1- 29.7) | 30.7<br>(29.3- 32.0) | 30.8<br>(29.4- 32.1) | 29.3<br>(27.9- 30.7) | 32.7<br>(31.0- 34.2) | 37.8<br>(36.1- 39.3) | 29.2<br>(27.5- 30.7) | 37.0<br>(35.0- 39.0) | 39.7<br>(37.7- 41.7) |  | -- | -- | 39.1<br>(35.0- 42.9) |
| DELU | 23.4<br>(22.1-24.7) | 26.0<br>(24.6-27.3) | 25.0<br>(23.6-26.3) | 23.3<br>(22.1-24.6) | 24.8<br>(23.4-26.3) | 31.9<br>(30.3-33.4) | 23.4<br>(22.0-24.8) | 31.7<br>(29.8-33.5) | 32.5<br>(30.5-34.5) | -- |  | 54.0<br>(50.9-57.1) | 33.9<br>(30.1-37.8) |
| HALL | 12.4<br>(11.5-13.3) | 12.7<br>(11.7-13.8) | 14.4<br>(13.2-15.4) | 13.7<br>(12.7-14.7) | 18.1<br>(16.7-19.4) | 15.9<br>(14.7-17.2) | 13.1<br>(11.9-14.4) | 15.3<br>(13.8-16.7) | 19.3<br>(17.7-20.9) | -- | 30.0<br>(27.8-32.0) |  | 16.6<br>(13.6-19.6) |
| ELAT | 7.0<br>(6.3- 7.7) | 8.8<br>(7.9- 9.6) | 8.3<br>(7.5- 9.2) | 8.2<br>(7.4- 9.0) | 8.7<br>(7.7- 9.6) | 9.7<br>(8.7-10.7) | 10.4<br>(9.3-11.4) | 15.7<br>(14.2-17.3) | 12.8<br>(11.6-14.2) | 10.2 (8.9-11.4) | 11.1<br>(9.6-12.6) | 9.8<br>(8.0-11.6) |  |

Displayed is conditional prevalence (%) with 95% confidence intervals.
ABER, Aberrant motor behavior; AGIT, Agitation/aggression; ANXI, Anxiety; APAT, Apathy/indifference; APPE, Appetite/eating changes; DELU, Delusions; DEPR, Depression/dysphoria; DISI, Disinhibition; ELAT, Elation/euphoria; HALU, Hallucinations; IRRI, Irritability/lability; PSYC, Psychotic symptoms (hallucinations and/or delusions); SLEE, nighttime behavioral disturbances.

## DISCUSSION

The present study characterizes cross-sectional point prevalence of neuropsychiatric symptoms and symptom pairs in three longitudinal studies (ADAMS, ADNI and NACC). Prevalence of individual NPS has been repeatedly described in the literature for various populations of elderly subjects with or without dementia ^26–30^. In contrast, to the best of our knowledge, comprehensive analysis of prevalence of symptom pairs is reported here for the first time.

Co-expression of different NPS was previously discussed only in the studies that aimed to identify clusters of symptoms using methods such as principal component analysis, factor analysis or cluster analysis ^5,31–33^. However, such reports typically did not include quantitative information on prevalence of specific symptom pairs or combinations and certainly did not address prevalence of all possible symptom pairs.

NPS were found to be highly prevalent across all three studies. Considering all subjects with available NPI data, there were 28.9% or 36.7% with at least one symptom in the ADAMS study (survey- and sample-based estimates respectively), 55.1% and 61.3% in ADNI and NACC studies, respectively (Supplemental Table 4). Restricting this analysis to subjects with cognitive deficit only (MMSE ≤26), there were 39.1% or 44.0% with at least one symptom in the ADAMS study (survey- and sample-based estimates respectively), 72.2% and 81.2% in ADNI and NACC studies, respectively. These estimates are in agreement with previously reported values from other studies.

On average, in all three studies, subjects with cognitive deficit had more than one symptom. For example, in the NACC study, subjects with severe cognitive deficit (MMSE ≤9) had on average five neuropsychiatric symptoms (Supplemental Table 6). This does not mean, however, that all symptoms were of the same or similar clinical relevance. For example, if one takes into account only symptoms with severity ratings of 2 or 3, the average number of symptoms per NACC study subject with severe cognitive deficit is about 2.5 (Supplemental Table 7).

Overall, prevalence of all symptoms was lowest in the ADAMS study and, for most symptoms, was highest in the NACC study. Given the nature of these three studies, we did expect to see differences in NPS prevalence for the reasons cited below.

The ADAMS data originate from a stratified random subsample of individuals >70 years old at the time of selection from the larger Health and Retirement Study sample, a nationally representative survey, designed to investigate the health, social and economic implications of aging within the US population.

The primary goal of the ADNI is to test whether serial imaging, genetic, biochemical biomarkers as well clinical and neuropsychological assessment can be combined to characterize the entire spectrum of Alzheimer’s Disease (AD), as the pathology evolves from normal aging through very mild symptoms, to mild cognitive impairment (MCI) to dementia. Eligible participants with MMSE scores ≥20 were recruited by sites across the US and Canada. Accordingly, the average MMSE score in the ADNI sample was higher than in the ADAMS study.

The NACC is not a survey (like ADAMS) or a natural history study (like ADNI). NACC captures data of participants from Alzheimer’s Disease Research Centers (ADRCs) across the USA. Participants enrolled in the NACC reflect clinical referrals at ADRCs, self-referral, referral by family members, or active recruitment through community organizations. Most ADRCs also enroll volunteers with normal cognition.

The NACC sample is closer to the ADNI sample than to ADAMS in terms of years of education, age, MMSE score and data collection at clinical research centers (rather than in-home assessments). However, unlike ADAMS, neither ADNI nor NACC represent a weighted random sample of the US population. Study recruitment strategies likely contributed to the overall prevalence of NPS in the NACC and ADNI studies being similar and considerably higher than in the ADAMS study. At first glance, the latter observation appears to be at odds with the evidence of comparable degree of cognitive deficit in the ADAMS study. Indeed, within each of the three studies, greater cognitive deficit was paralleled by higher NPS prevalence. Between-study comparisons mislead to an opposite conclusion illustrating the relevance of various methodological differences between these three studies.

Having said that, one should nevertheless still acknowledge that there were also a number of similarities between studies. For example, disinhibition, aberrant behavior, delusions, hallucinations and elation were the least prevalent symptoms in all studies in participants with and without cognitive deficit. Apathy, while relatively rare in cognitively intact participants, was prominently prevalent in participants with cognitive deficit in all studies. Further, nighttime behavioral disturbances, relatively frequent in participants with MMSE score ≤26, are relatively less frequent in participants with MMSE score >26 in both ADNI and NACC studies (sleep data were not available for the ADAMS study).

The above-described observations for individual symptoms generally apply to prevalence of symptom pairs. Prevalence of most symptom pairs is highest in the NACC study and lowest in the ADAMS. Further, prevalence of all symptom pairs is elevated in subjects with a cognitive deficit, an observation seen across all three studies. Similar to what was observed for prevalence of individual symptoms, presence of a cognitive deficit (MMSE score ≤26) was associated with substantially greater prevalence of symptom pairs.

There are two conclusions that are specific to prevalence of symptom pairs.

First, in all three studies and MMSE strata, we observed all possible pair combinations, from those commonly recognized and discussed (e.g., hallucinations and delusions) to those that might be seen as rather counter-intuitive (e.g., apathy and agitation; Table 2).

Second, despite the above observation made for conditional prevalence, prevalence of symptom pairs cannot be readily predicted based on prevalence of individual symptoms. If, in a given pair, occurrence of each symptom was statistically a completely independent random event with equal probability for every subject, then prevalence of such a symptom pair would be equal to a product of multiplying prevalence of single symptoms. For example, in NACC subjects without cognitive impairment, prevalence of depression is 21.8% while prevalence of irritability is 19.8% and their product is 4.3% (Supplemental Table 10).

However, the observed prevalence for the depression-irritability pair is 9.7% (Table 2). The same conclusion can be made for all other symptoms pairs, the observed prevalences of which are consistently greater than what one would expect if symptoms were statistically independent random events.

Of note, differences between expected and observed prevalence of symptom pairs are even more pronounced for pairs of symptoms with severity scores >1 (can be calculated based on the data presented in Supplemental Table 11 and compared with Supplemental Table 18). However, this observation should not be taken as evidence for a shared mechanisms behind different symptoms or for symptoms not being independent from each other. For example, please refer to Supplement Section 7 for a hypothetical example of two populations within each of which symptoms A and B are statistically independent but, when combined, prevalence of a symptom pair can no longer be predicted by prevalence of individual symptoms A and B. In other words, prevalence of symptom pairs cannot be readily predicted because the real-world population is not homogenous and usually comprises a mixture of subpopulations with unique characteristics (such as prevalence of individual symptoms in subjects with different types of dementia ^5,34^).

## LIMITATIONS

One limitation of this study is that the analyses were performed using NPS data collected using one specific tool (NPI). Albeit the NPI is the most commonly used tool in clinical and epidemiological dementia research, it is unclear to what extent the conclusions of our study would hold if NPS data were collected using other tools. This limitation is particularly relevant given that the NPI collects information about symptom frequency and severity during a four-week period preceding the interview. As most symptoms are not constantly present, any conclusions about their co-occurrence are restricted to such specific short time windows. Each of the studies (ADAMS, ADNI and NACC) contained longitudinal data that could theoretically address this limitation. However, we performed cross-sectional analyses because there were substantially less observations available for subsequent years or visits.

Another potential limitation pertaining to the data collection methodology is that we cannot exclude the possibility that some subjects participated in more than one study. Thus, we cannot consider ADAMS, ADNI and NACC as three entirely independent sources of NPS profiles.

Further, ADAMS, ADNI and NACC utilized different versions of the NPI questionnaire (NPI-10, NPI-12 and NPI-Q, respectively) which adds to methodological differences between these studies.

Although this is not necessarily a limitation, one should acknowledge that the present report does not provide any descriptive measures of association between the symptoms (i.e., correlations). In our view, the use of such descriptive statistic is not justified since the datasets consist of participants with heterogeneous characteristics (e.g., low and high MMSE scores, presence of several, single or no NPS).

## CONCLUSION

Neuropsychiatric symptoms in people with dementia are increasingly recognized as a major unmet clinical and pharmaco-economic need. There are a growing number of clinical trials evaluating safety and therapeutic efficacy of a broad range of novel and re-purposed agents to address these disabling symptoms. Such studies typically focus on one symptom as a primary endpoint. However, it is evident that, in most study participants, other symptoms will be present too and may affect the study outcome ^10^. Further, a high degree of co-occurrence of different neuropsychiatric symptoms supports the consideration of basket-like trial designs that are common in oncology but have only recently been contextualized in the realm of neurodegenerative diseases ^35^.

The overall aim of such study designs is to increase development efficiency by reducing redundancy in trial implementation to enhance recruitment rates, share placebo groups and use biomarkers associated with the treatment mechanism of action across various associated, but non-identical, neurodegenerative conditions. A basket trial design simultaneously evaluates the effects of a therapy on multiple conditions with common underlying neuropathological features or shared clinical symptoms ^35^. Hence, the data presented by the analyses in this paper may help to identify symptom combinations that are highly prevalent and could stimulate further exploration of their prevalence under conditions of specific neurodegenerative diseases.

Alternatively, the present data may further allow a therapeutic agent to be assessed in a single condition in which there is intersectionality or coexistence of several NPS to determine the broader-based efficacy of such a drug’s impact on more than one neuropsychiatric symptom simultaneously. For example, when planning a study to estimate treatment-related changes in expression of one specific NPS as the primary outcome, one should keep in mind that recruited subjects will exhibit high rates of other NPS. Presence of some of these co-occurring symptoms may be prominent enough to allow assessment of treatment effects (e.g., as secondary or exploratory outcomes).

The present study clearly illustrates that, while there is the possibility of any combination of neuropsychiatric symptoms, their co-occurrence cannot be readily predicted based on the prevalence of individual symptoms. Thus, our study results can serve as a source of reference information to inform the design and recruitment strategies for future clinical and epidemiological research on neuropsychiatric symptoms.

## AUTHOR CONTRIBUTIONS

TLG and AYB conceived the study; HJM obtained permission to access the ADAMS and ADNI datasets; JS obtained permission to access the NACC UDS; TLG performed the data extraction and analysis; TLG and AYB wrote the initial manuscript draft; all authors contributed to finalization of the manuscript.

## ACKNOWLEDGMENTS

The ADAMS is a supplement to the Health and Retirement Study (HRS) data that is sponsored by the National Institute on Aging (U01-AG009740) and conducted by the University of Michigan with the specific aim of conducting a population-based study of dementia.

Data collection and sharing for the Alzheimer’s Disease Neuroimaging Initiative (ADNI) is funded by the National Institute on Aging (National Institutes of Health Grant U19 AG024904). The grantee organization is the Northern California Institute for Research and Education. In the past, ADNI has also received funding from the National Institute of Biomedical Imaging and Bioengineering, the Canadian Institutes of Health Research, and private sector contributions through the Foundation for the National Institutes of Health (FNIH) including generous contributions from the following: AbbVie, Alzheimer’s Association; Alzheimer’s Drug Discovery Foundation; Araclon Biotech; BioClinica, Inc.; Biogen; Bristol-Myers Squibb Company; CereSpir, Inc.; Cogstate; Eisai Inc.; Elan Pharmaceuticals, Inc.; Eli Lilly and Company; EuroImmun; F. Hoffmann-La Roche Ltd and its affiliated company Genentech, Inc.; Fujirebio; GE Healthcare; IXICO Ltd.; Janssen Alzheimer Immunotherapy Research & Development, LLC.; Johnson & Johnson Pharmaceutical Research &Development LLC.; Lumosity; Lundbeck; Merck & Co., Inc.; Meso Scale Diagnostics, LLC.; NeuroRx Research; Neurotrack Technologies; Novartis Pharmaceuticals Corporation; Pfizer Inc.; Piramal Imaging; Servier; Takeda Pharmaceutical Company; and Transition Therapeutics.

The NACC database is funded by NIA/NIH Grant U24 AG072122. NACC data are contributed by the NIA-funded ADRCs: P30 AG062429 (PI James Brewer, MD, PhD), P30 AG066468 (PI Oscar Lopez, MD), P30 AG062421 (PI Bradley Hyman, MD, PhD), P30 AG066509 (PI Thomas Grabowski, MD), P30 AG066514 (PI Mary Sano, PhD), P30 AG066530 (PI Helena Chui, MD), P30 AG066507 (PI Marilyn Albert, PhD), P30 AG066444 (PI John Morris, MD), P30 AG066518 (PI Jeffrey Kaye, MD), P30 AG066512 (PI Thomas Wisniewski, MD), P30 AG066462 (PI Scott Small, MD), P30 AG072979 (PI David Wolk, MD), P30 AG072972 (PI Charles DeCarli, MD), P30 AG072976 (PI Andrew Saykin, PsyD), P30 AG072975 (PI David Bennett, MD), P30 AG072978 (PI Neil Kowall, MD), P30 AG072977 (PI Robert Vassar, PhD), P30 AG066519 (PI Frank LaFerla, PhD), P30 AG062677 (PI Ronald Petersen, MD, PhD), P30 AG079280 (PI Eric Reiman, MD), P30 AG062422 (PI Gil Rabinovici, MD), P30 AG066511 (PI Allan Levey, MD, PhD), P30 AG072946 (PI Linda Van Eldik, PhD), P30 AG062715 (PI Sanjay Asthana, MD, FRCP), P30 AG072973 (PI Russell Swerdlow, MD), P30 AG066506 (PI Todd Golde, MD, PhD), P30 AG066508 (PI Stephen Strittmatter, MD, PhD), P30 AG066515 (PI Victor Henderson, MD, MS), P30 AG072947 (PI Suzanne Craft, PhD), P30 AG072931 (PI Henry Paulson, MD, PhD), P30 AG066546 (PI Sudha Seshadri, MD), P20 AG068024 (PI Erik Roberson, MD, PhD), P20 AG068053 (PI Justin Miller, PhD), P20 AG068077 (PI Gary Rosenberg, MD), P20 AG068082 (PI Angela Jefferson, PhD), P30 AG072958 (PI Heather Whitson, MD), P30 AG072959 (PI James Leverenz, MD).

## DATA STATEMENT

The ADAMS data are publicly available and access can be requested from the HRS Web site (https://hrs.isr.umich.edu/).

The ADNI data are publicly available and access can be requested from the ADNI project website (https://adni.loni.usc.edu/data-samples/access-data/).

The NACC data are publicly available and access can be requested from the NACC website (https://naccdata.org/requesting-data/submit-data-request).

