## Supplemental tables for "Co-occurrence of neuropsychiatric symptoms in ADAMS, ADNI and NACC studies as assessed by Neuropsychiatric Inventory"

### Supplement section 1. Data management

For ADAMS and ADNI analyses, we merged the original data tables as shown in Supplemental Table 1. We used ADNI data tables stored in ADNIMERGE R package (ADNIMERGE_0.0.1.tar.gz). For NACC analyses, all data was extracted from a single data table.

Supplemental Table 1. Data structure

| Data table | Parameter | Variables extracted |
|---|---|---|
| ADAMS study | | |
| ADAMS1AB_R.xlsx, ADAMS1BB_R.xlsx, ADAMS1CB_R.xlsx, ADAMS1DB_R.xlsx | NPI-10 | Subject ID, NPI-10 frequency and severity scores for waves A-D as per codelist |
| ADAMS1AN_R.xlsx, ADAMS1BN_R.xlsx, ADAMS1CN_R.xlsx, ADAMS1DN_R.xlsx | MMSE | Subject ID, MMSE total score for waves A-D as per code list |
| ADAMS1TRK_R.xlsx | Demographics, survey parameters | Subject ID, Sex, Age for waves A-D, Race, Ethnicity, Education, year and month of examination for waves A-D as per code list; survey individual weights, survey clusters, survey strata. |
| ADNI study | | |
| npi (ADNIMERGE R package v0.0.1) | NPI-12 | Subject ID, Visit, NPI-12 frequency and severity scores, date of examination as per code list, original protocol, follow-up protocol |
| mmse (ADNIMERGE R package v0.0.1) | MMSE | Subject ID, Visit, MMSE total score as per code list |
| adnimerge (ADNIMERGE R package v0.0.1) | Demographics | Subject ID, Visit, Age, Race, Ethnicity, Education as per code list |
| NACC study | | |
| commercial_nacc62.csv | NPI-Q, MMSE, demographics | Subject ID, Visit, NPI-Q severity scores, date of examination, MMSE total score, Age, Race, Ethnicity, Education as per code list, Alzheimer's Disease Research Center ID |

ADAMS dataset contained four biannual visits called waves (Wave A – Wave B). Wave A was used in our analysis; it contained 799 participants with NPI-10 severity and frequency scores and MMSE total score (Supplemental Table 2). One participant (female, age 81, MMSE total score 1) had a missing DELU domain.

ADNI dataset contained 17 annual visits, visit Month 12 was used for the analysis; it contained 889 participants with NPI-12 severity and frequency scores and MMSE total score (Supplemental Table 2). One participant (male, age 63, MMSE total score 23) had a missing DISI domain, another (male, age 73, MMSE total score 30) had a missing SLEE domain. Of the 889 participants used in our analysis, 112 were enrolled in ADNIGO protocol and then switched to ADNI2, 618 were enrolled in ADNI2, and 159 were enrolled in ADNI3.

NACC dataset contained 18 annual visits. Visit 1 with 26 171 participants that had NPI-Q assessments and MMSE total score was used for our analysis (Supplemental Table 2). Fifteen participants had partially missing NPI assessments: one female, 46 years old, MMSE total score 26, had missing DEPR and AGIT NPI domains, one male, 77 years old, MMSE total score

30, had missing DEPR, DELU, and HALL NPI domains, one female, 80 years old, MMSE total score 28, had missing IRRI NPI domain, one male, 79 years old, MMSE total score 30, had missing IRRI NPI domain, one male, 86 years old, MMSE total score 13, had missing IRRI NPI domain, one male, 72 years old, MMSE total score 24, had missing IRRI and APAT NPI domains, one female, 72 years old, MMSE total score 23, had missing ANXI NPI domain, one female, 66 years old, MMSE total score 21, had missing APAT NPI domain, one male, 64 years old, MMSE total score 24, had missing APAT NPI domain, one female, 77 years old, MMSE total score 29, had missing SLEE NPI domain, one male, 91 years old, MMSE total score 26, had missing SLEE and APPE NPI domains, one female, 73 years old, MMSE total score 29, had missing SLEE and ABER domains, one female, 81 years old, MMSE total score 28, had missing SLEE and ABER NPI domains, one female, 95 years old, MMSE total score 24, had missing AGIT NPI domain, one female, 68 years old, MMSE total score 21, had missing APPE NPI domain.

All NACC participants were enrolled in 37 Alzheimer's Disease Research Centers (ADRCs), but the 26 171 participants used in our analysis were enrolled in 27 ADRCs.

Supplemental Table 2. Sample sizes used in the analysis

| Study, visit | Number of participants | | |
|---|---|---|---|
| | Total | NPI assessments | NPI and MMSE assessments* |
| ADAMS survey, Wave A | 24,193,200 | 23,904,171 | 23,524,029 |
| ADAMS, Wave A | 856 | 840 | 799 |
| ADNI, Month 12 | 922 | 914 | 889 |
| NACC, Visit 1 | 43,185 | 41,169 | 26,171 |

Survey estimates for ADAMS are based on survey population weights. * Population used for the analysis.

Supplemental Table 3. Mean MMSE score in different MMSE strata

| MMSE stratum | Mean MMSE score [SD] | | | |
|---|---|---|---|---|
| | ADAMS survey | ADAMS | ADNI | NACC |
| Any | 25.7 [5.2] | 21.7 [7.1] | 26.9 [3.8] | 25.0 [6.1] |
| > 26 | 28.7 [1.0] | 28.6 [1.0] | 28.9 [1.1] | 28.9 [1.1] |
| ≤ 26 | 21.2 [5.6] | 18.4 [6.4] | 22.6 [4.1] | 19.9 [6.3] |
| > 19 - ≤ 26 | | | | 23.6 [2.0] |
| > 9 - ≤ 19 | | | | 15.7 [2.7] |
| ≤ 9 | | | | 4.5 [3.2] |

Survey estimates for ADAMS are based on survey population weights, SD is based on survey clusters and survey strata. SD, Standard deviation.

### Supplement section 2. Subjects without neuropsychiatric symptoms

Supplemental Table 4. Proportion of subjects without neuropsychiatric symptoms* with the NPI severity score > 0

| MMSE stratum | Number of participants with no symptoms / total number (%) | | | |
|---|---|---|---|---|
| | ADAMS survey | ADAMS | ADNI | NACC |
| > 26 | 10,982,507/14,111,455 (77.8%) | 202/256 (78.9%) | 319/601 (53.1%) | 8,008/14,887 (53.8%) |
| ≤ 26 | 5,736,194/ 9,412,574 (60.9%) | 304/543 (56.0%) | 80/288 (27.8%) | 2,126/11,284 (18.8%) |
| > 19 - ≤ 26 | | | | 1,712/ 7,317 (23.4%) |
| > 9 - ≤ 19 | | | | 334/ 2,993 (11.2%) |
| ≤ 9 | | | | 80/ 974 ( 8.2%) |

Survey estimates for ADAMS are based on survey population weights. * Severity scores of all assessed NPI domains are equal to 0.

Supplemental Table 5. Proportion of subjects without neuropsychiatric symptoms* with the NPI severity score > 1

| MMSE stratum | Number of participants with no symptoms / total number (%) | | | |
|---|---|---|---|---|
| | ADAMS survey | ADAMS | ADNI | NACC |
| > 26 | 12,830,524/14,111,455 (90.9%) | 235/256 (91.8%) | 485/601 (80.7%) | 11,851/14,887 (79.6%) |
| ≤ 26 | 7,753,671/ 9,412,574 (82.4%) | 425/543 (78.3%) | 180/288 (62.5%) | 5,525/11,284 (49.0%) |
| > 19 - ≤ 26 | | | | 4,030/ 7,317 (55.1%) |
| > 9 - ≤ 19 | | | | 1,200/ 2,993 (40.1%) |
| ≤ 9 | | | | 295/ 974 (30.3%) |

Survey estimates for ADAMS are based on survey population weights. * Severity scores of all assessed NPI domains are equal to or less than 1.

### Supplement section 3. Average number of neuropsychiatric symptoms per participant

Supplemental Table 6. Mean [standard deviation] number of neuropsychiatric symptoms with the NPI severity score > 0 per participant

| MMSE stratum | ADAMS survey | ADAMS | ADNI | NACC |
|---|---|---|---|---|
| Any | 0.7 [1.4] | 1.0 [1.8] | 1.5 [2.0] | 2.2 [2.6] |
| > 26 | 0.4 [1.0] | 0.4 [0.9] | 1.1 [1.7] | 1.3 [2.0] |
| ≤ 26 | 1.1 [1.9] | 1.3 [2.0] | 2.4 [2.3] | 3.3 [2.8] |
| > 19 - ≤ 26 | | | | 2.9 [2.6] |
| > 9 - ≤ 19 | | | | 4.0 [2.9] |
| ≤ 9 | | | | 5.0 [3.1] |

Survey estimates for ADAMS are based on survey population weights, standard deviation is based on survey clusters and survey strata.

Supplemental Table 7. Mean [standard deviation] number of neuropsychiatric symptoms with the NPI severity score > 1 per participant

| MMSE stratum | ADAMS survey | ADAMS | ADNI | NACC |
|---|---|---|---|---|
| Any | 0.3 [0.9] | 0.4 [1.0] | 0.5 [1.1] | 0.9 [1.8] |
| > 26 | 0.1 [0.4] | 0.1 [0.4] | 0.3 [0.9] | 0.5 [1.2] |
| ≤ 26 | 0.5 [1.4] | 0.6 [1.4] | 0.8 [1.4] | 1.5 [2.2] |
| > 19 - ≤ 26 | | | | 1.2 [1.9] |
| > 9 - ≤ 19 | | | | 1.9 [2.3] |
| ≤ 9 | | | | 2.5 [2.7] |

Survey estimates for ADAMS are based on survey population weights, standard deviation is based on survey clusters and survey strata.

### Supplement section 4. Prevalence of NPI domains

Supplemental Table 8. Prevalence of neuropsychiatric symptoms with the NPI severity score > 0

| NPS | ADAMS survey | ADAMS | ADNI | NACC |
|---|---|---|---|---|
| | N = 23,524,029 | N = 799 | N = 889 | N = 26,171 |
| DEPR | 18.0 (14.4-22.1) | 19.5 (16.8-22.4) | 24.5 (21.7-27.5) | 29.5 (28.9-30.0) |
| IRRI | 9.1 ( 5.8-13.6) | 11.0 ( 8.9-13.4) | 22.6 (19.9-25.5) | 27.2 (26.7-27.8) |
| ANXI | 8.1 ( 5.2-12.0) | 10.9 ( 8.8-13.3) | 14.8 (12.6-17.4) | 25.3 (24.8-25.9) |
| APAT | 7.9 ( 5.9-10.3) | 10.6 ( 8.6-13.0) | 18.0 (15.5-20.7) | 24.7 (24.2-25.2) |
| SLEE | NA | NA | 18.4 (15.9-21.1) | 21.9 (21.4-22.4) |
| AGIT | 7.9 ( 4.6-12.6) | 12.5 (10.3-15.0) | 14.8 (12.6-17.4) | 20.8 (20.3-21.3) |
| APPE | NA | NA | 11.8 ( 9.8-14.1) | 17.1 (16.7-17.6) |
| DISI | 3.9 ( 2.1- 6.6) | 5.3 ( 3.8- 7.0) | 9.9 ( 8.0-12.1) | 13.2 (12.8-13.6) |
| ABER | 2.7 ( 1.5- 4.3) | 5.8 ( 4.2- 7.6) | 5.6 ( 4.2- 7.3) | 11.3 (10.9-11.7) |
| PSYC | 4.5 ( 3.0- 6.4) | 10.6 ( 8.6-13.0) | 4.4 ( 3.1- 5.9) | 10.6 (10.2-10.9) |
| DELU | 3.7 ( 2.2- 5.8) | 7.5 ( 5.8- 9.6) | 3.5 ( 2.4- 4.9) | 8.4 ( 8.1- 8.7) |
| HALL | 2.4 ( 1.3- 4.1) | 6.1 ( 4.6- 8.0) | 1.5 ( 0.8- 2.5) | 4.5 ( 4.3- 4.8) |
| ELAT | 1.2 ( 0.2- 3.4) | 1.0 ( 0.4- 2.0) | 2.2 ( 1.4- 3.5) | 3.5 ( 3.3- 3.7) |

Displayed are prevalence (%) and 95% confidence intervals. Survey estimates for ADAMS are based on survey population weights, confidence intervals are based on survey clusters and survey strata.

NA, Not applicable; NPS, Neuropsychiatric symptom.

ABER, Aberrant motor behavior; AGIT, Agitation/aggression; ANXI, Anxiety; APAT, Apathy/indifference; APPE, Appetite/eating changes; DELU, Delusions; DEPR, Depression/dysphoria; DISI, Disinhibition; ELAT, Elation/euphoria; HALU, Hallucinations; IRRI, Irritability/lability; PSYC, Psychotic symptoms (hallucinations and/or delusions); SLEE, Nighttime behavioral disturbances.

Supplemental Table 9. Prevalence of neuropsychiatric symptoms with the NPI severity score > 1

| NPS | ADAMS, survey | ADAMS | ADNI | NACC |
|---|---|---|---|---|
| | N = 23,524,029 | N = 799 | N = 889 | N = 26,171 |
| DEPR | 4.4 (2.5-6.9) | 5.4 (3.9-7.2) | 4.9 (3.6-6.6) | 10.8 (10.4-11.2) |
| IRRI | 2.8 (1.2-5.7) | 4.1 (2.9-5.8) | 7.3 (5.7-9.2) | 10.1 ( 9.8-10.5) |
| ANXI | 3.2 (1.5-5.7) | 4.6 (3.3-6.3) | 4.2 (2.9-5.7) | 10.6 (10.2-11.0) |
| APAT | 5.1 (3.2-7.6) | 7.3 (5.6-9.3) | 6.5 (5.0-8.4) | 11.7 (11.3-12.1) |
| SLEE | NA | NA | 6.3 (4.8-8.1) | 10.2 ( 9.8-10.6) |
| AGIT | 2.9 (1.5-4.9) | 5.6 (4.1-7.5) | 3.8 (2.7-5.3) | 8.7 ( 8.3- 9.0) |
| APPE | NA | NA | 6.3 (4.8-8.1) | 7.2 ( 6.9- 7.6) |
| DISI | 1.4 (0.4-3.4) | 1.9 (1.1-3.1) | 2.7 (1.7-4.0) | 5.7 ( 5.5- 6.0) |
| ABER | 1.6 (0.7-3.1) | 3.5 (2.3-5.0) | 1.9 (1.1-3.0) | 5.3 ( 5.1- 5.6) |
| PSYC | 2.4 (1.2-4.4) | 4.3 (3.0-5.9) | 1.8 (1.0-2.9) | 4.8 ( 4.5- 5.0) |
| DELU | 2.3 (1.1-4.4) | 3.9 (2.7-5.5) | 1.7 (0.9-2.8) | 3.8 ( 3.6- 4.1) |
| HALL | 0.4 (0.1-0.8) | 1.5 (0.8-2.6) | 0.3 (0.1-1.0) | 1.8 ( 1.7- 2.0) |
| ELAT | 0.5 (0.0-2.6) | 0.1 (0.0-0.7) | 0.7 (0.2-1.5) | 1.3 ( 1.2- 1.5) |

Displayed are prevalence (%) and 95% confidence intervals. Survey estimates for ADAMS are based on survey population weights, confidence intervals are based on survey clusters and survey strata.

NA, Not applicable; NPS, Neuropsychiatric symptom.

ABER, Aberrant motor behavior; AGIT, Agitation/aggression; ANXI, Anxiety; APAT, Apathy/indifference; APPE, Appetite/eating changes; DELU, Delusions; DEPR, Depression/dysphoria; DISI, Disinhibition; ELAT, Elation/euphoria; HALU, Hallucinations; IRRI, Irritability/lability; PSYC, Psychotic symptoms (hallucinations and/or delusions); SLEE, Nighttime behavioral disturbances.

Supplemental Table 10. Prevalence of neuropsychiatric symptoms with the NPI severity score > 0 per MMSE stratum (ADAMS and ADNI studies only)

| NPS | ADAMS survey | | ADAMS | | ADNI | |
|---|---|---|---|---|---|---|
| | MMSE > 26 | MMSE ≤ 26 | MMSE > 26 | MMSE ≤ 26 | MMSE > 26 | MMSE ≤ 26 |
| | N = 14,111,455 | N = 9,412,574 | N = 256 | N = 543 | N = 601 | N = 288 |
| DEPR | 13.7 (8.9-19.7) | 24.4 (19.4-30.0) | 12.5 (8.7-17.2) | 22.8 (19.4-26.6) | 20.8 (17.6-24.3) | 32.3 (26.9-38.0) |
| IRRI | 8.1 (4.2-13.7) | 10.8 ( 6.8-15.9) | 7.4 (4.5-11.3) | 12.7 (10.0-15.8) | 19.8 (16.7-23.2) | 28.5 (23.3-34.1) |
| ANXI | 7.4 (3.5-13.4) | 9.2 ( 5.5-14.3) | 7.0 (4.2-10.9) | 12.7 (10.0-15.8) | 10.1 ( 7.9-12.8) | 24.7 (19.8-30.1) |
| APAT | 5.7 (3.3- 9.2) | 11.1 ( 7.0-16.6) | 5.1 (2.7- 8.5) | 13.3 (10.5-16.4) | 10.0 ( 7.7-12.7) | 34.7 (29.2-40.5) |
| SLEE | NA | NA | NA | NA | 17.7 (14.7-21.0) | 19.8 (15.3-24.9) |
| AGIT | 4.3 (1.4- 9.5) | 13.4 ( 8.4-20.0) | 4.7 (2.4- 8.0) | 16.2 (13.2-19.6) | 12.3 ( 9.8-15.2) | 20.1 (15.7-25.2) |
| APPE | NA | NA | NA | NA | 6.5 ( 4.7- 8.8) | 22.9 (18.2-28.2) |
| DISI | 2.1 (0.6- 5.3) | 6.7 ( 3.7-11.1) | 2.0 (0.6- 4.5) | 6.8 ( 4.8- 9.3) | 8.2 ( 6.1-10.6) | 13.6 ( 9.8-18.1) |
| ABER | 0.0 (0.0- NE) | 6.7 ( 3.9-10.5) | 0.0 (0.0- 1.4) | 8.5 ( 6.3-11.1) | 2.0 ( 1.0- 3.5) | 13.2 ( 9.5-17.7) |
| PSYC | 0.7 (0.0- 3.7) | 10.2 ( 6.7-14.8) | 0.4 (0.0- 2.2) | 15.5 (12.5-18.8) | 1.2 ( 0.5- 2.4) | 11.1 ( 7.7-15.3) |
| DELU | 0.7 (0.0- 3.7) | 8.2 ( 4.9-12.8) | 0.4 (0.0- 2.2) | 10.9 ( 8.4-13.8) | 1.0 ( 0.4- 2.2) | 8.7 ( 5.7-12.5) |
| HALL | 0.0 (0.0- NE) | 6.0 ( 3.3-10.0) | 0.0 (0.0- 1.4) | 9.0 ( 6.8-11.8) | 0.2 ( 0.0- 0.9) | 4.2 ( 2.2- 7.2) |
| ELAT | 1.5 (0.2- 5.6) | 0.7 ( 0.1- 2.1) | 0.8 (0.1- 2.8) | 1.1 ( 0.4- 2.4) | 1.7 ( 0.8- 3.0) | 3.5 ( 1.7- 6.3) |

Displayed are prevalence (%) and 95% confidence intervals. Survey estimates for ADAMS are based on survey population weights, confidence intervals are based on survey clusters and survey strata.

NA, Not applicable; NE, Not estimable; NPS, Neuropsychiatric symptom.

ABER, Aberrant motor behavior; AGIT, Agitation/aggression; ANXI, Anxiety; APAT, Apathy/indifference; APPE, Appetite/eating changes; DELU, Delusions; DEPR, Depression/dysphoria; DISI, Disinhibition; ELAT, Elation/euphoria; HALU, Hallucinations; IRRI, Irritability/lability; PSYC, Psychotic symptoms (hallucinations and/or delusions); SLEE, Nighttime behavioral disturbances.

Supplemental Table 10. Prevalence of neuropsychiatric symptoms with the NPI severity score > 1 per MMSE stratum (ADAMS and ADNI studies only)

| NPS | ADAMS survey | | ADAMS | | ADNI | |
|---|---|---|---|---|---|---|
| | MMSE > 26 | MMSE ≤ 26 | MMSE > 26 | MMSE ≤ 26 | MMSE > 26 | MMSE ≤ 26 |
| | N = 14,111,455 | N = 9,412,574 | N = 256 | N = 543 | N = 601 | N = 288 |
| DEPR | 1.3 (0.2-3.9) | 9.0 (5.3-14.2) | 2.0 (0.6-4.5) | 7.0 (5.0- 9.5) | 3.8 (2.4-5.7) | 7.3 (4.6-10.9) |
| IRRI | 1.3 (0.1-5.3) | 5.1 (2.2-10.0) | 0.8 (0.1-2.8) | 5.7 (3.9- 8.0) | 6.0 (4.2-8.2) | 10.1 (6.8-14.1) |
| ANXI | 3.0 (1.0-6.8) | 3.4 (1.7- 5.9) | 2.7 (1.1-5.6) | 5.5 (3.8- 7.8) | 3.3 (2.0-5.1) | 5.9 (3.5- 9.3) |
| APAT | 3.1 (1.3-6.3) | 8.0 (4.6-12.9) | 3.1 (1.4-6.1) | 9.2 (6.9-12.0) | 3.3 (2.0-5.1) | 13.2 (9.5-17.7) |
| SLEE | NA | NA | NA | NA | 5.2 (3.5-7.3) | 8.7 (5.7-12.5) |
| AGIT | 0.9 (0.1-3.2) | 5.8 (2.9-10.4) | 1.2 (0.2-3.4) | 7.7 (5.6-10.3) | 3.0 (1.8-4.7) | 5.6 (3.2- 8.9) |
| APPE | NA | NA | NA | NA | 3.2 (1.9-4.9) | 12.8 (9.2-17.3) |
| DISI | 0.3 (0.0-1.8) | 3.1 (0.9- 7.4) | 0.4 (0.0-2.2) | 2.6 (1.4- 4.3) | 2.7 (1.5-4.3) | 2.8 (1.2- 5.4) |
| ABER | 0.0 (0.0- NE) | 4.1 (1.9- 7.7) | 0.0 (0.0-1.4) | 5.2 (3.5- 7.4) | 1.2 (0.5-2.4) | 3.5 (1.7- 6.3) |
| PSYC | 0.7 (0.0-3.7) | 5.1 (2.3- 9.5) | 0.4 (0.0-2.2) | 6.1 (4.2- 8.4) | 1.0 (0.4-2.2) | 3.5 (1.7- 6.3) |
| DELU | 0.7 (0.0-3.7) | 4.8 (2.2- 9.2) | 0.4 (0.0-2.2) | 5.5 (3.8- 7.8) | 0.8 (0.3-1.9) | 3.5 (1.7- 6.3) |
| HALL | 0.0 (0.0- NE) | 0.9 (0.4- 2.0) | 0.0 (0.0-1.4) | 2.2 (1.1- 3.8) | 0.2 (0.0-0.9) | 0.7 (0.1- 2.5) |
| ELAT | 0.8 (0.0-4.3) | 0.0 (0.0- NE) | 0.4 (0.0-2.2) | 0.0 (0.0- 0.7) | 0.3 (0.0-1.2) | 1.4 (0.4- 3.5) |

Displayed are prevalence (%) and 95% confidence intervals. Survey estimates for ADAMS are based on survey population weights, confidence intervals are based on survey clusters and survey strata.

NA, Not applicable; NE, Not estimable; NPS, Neuropsychiatric symptom.

ABER, Aberrant motor behavior; AGIT, Agitation/aggression; ANXI, Anxiety; APAT, Apathy/indifference; APPE, Appetite/eating changes; DELU, Delusions; DEPR, Depression/dysphoria; DISI, Disinhibition; ELAT, Elation/euphoria; HALU, Hallucinations; IRRI, Irritability/lability; PSYC, Psychotic symptoms (hallucinations and/or delusions); SLEE, Nighttime behavioral disturbances.

Supplemental Table 11. Prevalence of neuropsychiatric symptoms with the NPI severity score > 0 per MMSE stratum (NACC study only)

| NPS | NACC | | | | |
|---|---|---|---|---|---|
| | MMSE > 26 | MMSE ≤ 26 | 19 < MMSE ≤ 26 | 9 < MMSE ≤ 19 | MMSE ≤ 9 |
| | N = 14,887 | N = 11,284 | N = 7,317 | N = 2,993 | N = 974 |
| DEPR | 21.8 (21.1-22.5) | 39.6 (38.7-40.5) | 37.8 (36.7-38.9) | 43.4 (41.6-45.2) | 41.7 (38.6-44.9) |
| IRRI | 19.8 (19.2-20.5) | 37.1 (36.2-38.0) | 34.9 (33.8-36.0) | 40.1 (38.3-41.9) | 44.4 (41.2-47.5) |
| ANXI | 16.1 (15.5-16.7) | 37.4 (36.6-38.3) | 32.7 (31.6-33.7) | 45.2 (43.4-47.0) | 49.6 (46.4-52.8) |
| APAT | 12.5 (12.0-13.1) | 40.7 (39.8-41.7) | 35.2 (34.1-36.3) | 47.8 (46.0-49.6) | 61.1 (57.9-64.2) |
| SLEE | 17.0 (16.4-17.6) | 28.4 (27.6-29.2) | 26.9 (25.9-28.0) | 29.7 (28.0-31.3) | 35.4 (32.4-38.5) |
| AGIT | 12.2 (11.7-12.8) | 32.2 (31.3-33.1) | 27.1 (26.1-28.2) | 38.6 (36.8-40.3) | 50.8 (47.6-54.0) |
| APPE | 10.0 ( 9.5-10.5) | 26.5 (25.7-27.4) | 24.5 (23.5-25.5) | 29.7 (28.1-31.4) | 32.1 (29.2-35.2) |
| DISI | 7.3 ( 6.9- 7.7) | 21.0 (20.3-21.8) | 17.9 (17.0-18.8) | 24.8 (23.3-26.4) | 33.3 (30.3-36.3) |
| ABER | 4.2 ( 3.8- 4.5) | 20.8 (20.0-21.5) | 14.0 (13.2-14.8) | 29.9 (28.2-31.5) | 43.9 (40.8-47.1) |
| PSYC | 3.2 ( 2.9- 3.5) | 20.3 (19.5-21.0) | 13.9 (13.2-14.8) | 28.7 (27.1-30.4) | 41.9 (38.8-45.1) |
| DELU | 2.5 ( 2.3- 2.8) | 16.2 (15.5-16.8) | 11.1 (10.4-11.9) | 23.1 (21.6-24.6) | 32.6 (29.7-35.7) |
| HALL | 1.1 ( 1.0- 1.3) | 9.0 ( 8.4- 9.5) | 5.2 ( 4.7- 5.7) | 13.1 (11.9-14.4) | 24.5 (21.9-27.4) |
| ELAT | 2.1 ( 1.9- 2.4) | 5.3 ( 4.9- 5.7) | 4.3 ( 3.8- 4.8) | 6.6 ( 5.7- 7.5) | 8.9 ( 7.2-10.9) |

Displayed are prevalence (%) and 95% confidence intervals.
NPS, Neuropsychiatric symptom.
ABER, Aberrant motor behavior; AGIT, Agitation/aggression; ANXI, Anxiety; APAT, Apathy/indifference; APPE, Appetite/eating changes; DELU, Delusions; DEPR, Depression/dysphoria; DISI, Disinhibition; ELAT, Elation/euphoria; HALU, Hallucinations; IRRI, Irritability/lability; PSYC, Psychotic symptoms (hallucinations and/or delusions); SLEE, Nighttime behavioral disturbances.

Supplemental Table 12. Prevalence of neuropsychiatric symptoms with the NPI severity score > 1 per MMSE stratum (NACC study only)

| NPS | NACC | | | | |
|---|---|---|---|---|---|
| | MMSE > 26 | MMSE ≤ 26 | 19 < MMSE ≤ 26 | 9 < MMSE ≤ 19 | MMSE ≤ 9 |
| | N = 14,887 | N = 11,284 | N = 7,317 | N = 2,993 | N = 974 |
| DEPR | 7.1 (6.7-7.5) | 15.7 (15.1-16.4) | 14.5 (13.7-15.3) | 18.0 (16.6-19.4) | 17.9 (15.5-20.4) |
| IRRI | 6.2 (5.8-6.6) | 15.3 (14.6-16.0) | 14.1 (13.3-14.9) | 16.9 (15.6-18.3) | 19.2 (16.8-21.8) |
| ANXI | 5.8 (5.4-6.1) | 16.9 (16.2-17.6) | 14.2 (13.4-15.1) | 20.3 (18.9-21.8) | 26.6 (23.8-29.5) |
| APAT | 4.7 (4.4-5.1) | 20.8 (20.1-21.6) | 16.4 (15.6-17.3) | 26.1 (24.5-27.7) | 38.2 (35.1-41.3) |
| SLEE | 6.9 (6.5-7.4) | 14.5 (13.9-15.2) | 13.0 (12.2-13.8) | 16.6 (15.3-18.0) | 19.8 (17.4-22.5) |
| AGIT | 4.3 (4.0-4.7) | 14.4 (13.7-15.0) | 11.8 (11.1-12.5) | 16.8 (15.5-18.2) | 26.4 (23.6-29.3) |
| APPE | 3.7 (3.4-4.0) | 12.0 (11.4-12.6) | 10.7 (10.0-11.5) | 13.8 (12.6-15.1) | 15.7 (13.5-18.1) |
| DISI | 2.9 (2.6-3.2) | 9.5 ( 9.0-10.1) | 8.1 ( 7.5- 8.8) | 10.9 ( 9.8-12.1) | 15.6 (13.4-18.0) |
| ABER | 1.9 (1.7-2.1) | 9.9 ( 9.4-10.5) | 6.0 ( 5.5- 6.6) | 14.7 (13.5-16.0) | 24.4 (21.8-27.3) |
| PSYC | 1.1 (1.0-1.3) | 9.6 ( 9.0-10.1) | 6.3 ( 5.7- 6.9) | 14.0 (12.8-15.3) | 20.6 (18.1-23.3) |
| DELU | 0.9 (0.8-1.1) | 7.7 ( 7.2- 8.2) | 5.0 ( 4.6- 5.6) | 11.5 (10.4-12.7) | 15.7 (13.5-18.1) |
| HALL | 0.3 (0.3-0.5) | 3.8 ( 3.5- 4.2) | 2.1 ( 1.8- 2.5) | 5.4 ( 4.6- 6.2) | 11.8 ( 9.8-14.0) |
| ELAT | 0.9 (0.8-1.1) | 1.9 ( 1.7- 2.2) | 1.7 ( 1.4- 2.0) | 2.4 ( 1.9- 3.0) | 2.4 ( 1.5- 3.5) |

Displayed are prevalence (%) and 95% confidence intervals.
NPS, Neuropsychiatric symptom.
ABER, Aberrant motor behavior; AGIT, Agitation/aggression; ANXI, Anxiety; APAT, Apathy/indifference; APPE, Appetite/eating changes; DELU, Delusions; DEPR, Depression/dysphoria; DISI, Disinhibition; ELAT, Elation/euphoria; HALU, Hallucinations; IRRI, Irritability/lability; PSYC, Psychotic symptoms (hallucinations and/or delusions); SLEE, Nighttime behavioral disturbances.

Supplemental Table 13. Prevalence of neuropsychiatric symptoms with the NPI product score ≥ 4 (ADAMS and ADNI studies only)

| NPS | ADAMS survey<br>N = 23,524,029 | ADAMS<br>N = 799 | ADNI<br>N = 889 |
|---|---|---|---|
| DEPR | 3.4 (2.0-5.9) | 4.5 (3.2-6.2) | 2.4 (1.5-3.6) |
| IRRI | 2.3 (1.0-5.0) | 3.4 (2.2-4.9) | 4.4 (3.1-5.9) |
| ANXI | 2.1 (1.2-3.6) | 3.5 (2.3-5.0) | 2.2 (1.4-3.5) |
| APAT | 4.0 (2.5-6.5) | 6.0 (4.5-7.9) | 5.6 (4.2-7.3) |
| SLEE | NA | NA | 5.3 (3.9-7.0) |
| AGIT | 1.9 (0.9-3.8) | 3.4 (2.2-4.9) | 1.9 (1.1-3.0) |
| APPE | NA | NA | 5.3 (3.9-7.0) |
| DISI | 1.1 (0.4-3.1) | 1.4 (0.7-2.4) | 1.4 (0.7-2.3) |
| ABER | 1.6 (0.8-3.1) | 3.3 (2.1-4.7) | 1.6 (0.9-2.6) |
| PSYC | 2.2 (1.1-4.2) | 3.4 (2.2-4.9) | 1.0 (0.5-1.9) |
| DELU | 2.1 (1.0-4.3) | 2.9 (1.8-4.3) | 1.0 (0.5-1.9) |
| HALL | 0.3 (0.1-0.6) | 1.3 (0.6-2.3) | 0.1 (0.0-0.6) |
| ELAT | 0.0 (0.0- NE) | 0.0 (0.0-0.5) | 0.7 (0.2-1.5) |

Displayed are prevalence (%) and 95% confidence intervals. Survey estimates for ADAMS are based on survey population weights, confidence intervals are based on survey clusters and survey strata.

NA Not applicable; NE Not estimable; NPS, Neuropsychiatric symptom.

Prevalence for NPI product scores greater than 0 is equivalent to prevalence with NPI severity scores > 0.

ABER, Aberrant motor behavior; AGIT, Agitation/aggression; ANXI, Anxiety; APAT, Apathy/indifference; APPE, Appetite/eating changes; DELU, Delusions; DEPR, Depression/dysphoria; DISI, Disinhibition; ELAT, Elation/euphoria; HALU, Hallucinations; IRRI, Irritability/lability; PSYC, Psychotic symptoms (hallucinations and/or delusions); SLEE, Nighttime behavioral disturbances.

Supplemental Table 14. Prevalence of neuropsychiatric symptoms with the NPI product score ≥ 4 per MMSE stratum (ADAMS and ADNI studies only)

| NPS | ADAMS survey | | ADAMS | | ADNI | |
|---|---|---|---|---|---|---|
| | MMSE > 26 | MMSE ≤ 26 | MMSE > 26 | MMSE ≤ 26 | MMSE > 26 | MMSE ≤ 26 |
| | N = 14,111,455 | N = 9,412,574 | N = 256 | N = 543 | N = 601 | N = 288 |
| DEPR | 1.2 (0.2-4.1) | 6.8 (3.8-11.2) | 1.6 (0.4-4.0) | 5.9 (4.1- 8.2) | 1.2 (0.5-2.4) | 4.9 (2.7- 8.0) |
| IRRI | 0.7 (0.0-4.3) | 4.6 (1.8- 9.5) | 0.4 (0.0-2.2) | 4.8 (3.2- 6.9) | 3.5 (2.2-5.3) | 6.2 (3.7- 9.7) |
| ANXI | 1.6 (0.3-4.6) | 2.8 (1.4- 5.2) | 1.6 (0.4-4.0) | 4.4 (2.9- 6.5) | 1.8 (0.9-3.3) | 3.1 (1.4- 5.8) |
| APAT | 2.4 (0.7-5.9) | 6.4 (3.6-10.6) | 2.3 (0.9-5.0) | 7.7 (5.6-10.3) | 3.0 (1.8-4.7) | 11.1 (7.7-15.3) |
| SLEE | NA | NA | NA | NA | 4.2 (2.7-6.1) | 7.6 (4.8-11.3) |
| AGIT | 0.0 (0.0-0.2) | 4.6 (2.0- 9.1) | 0.4 (0.0-2.2) | 4.8 (3.2- 6.9) | 1.2 (0.5-2.4) | 3.5 (1.7- 6.3) |
| APPE | NA | NA | NA | NA | 2.5 (1.4-4.1) | 11.1 (7.7-15.3) |
| DISI | 0.0 (0.0- NE) | 2.9 (0.8- 7.3) | 0.0 (0.0-1.4) | 2.0 (1.0- 3.6) | 1.3 (0.6-2.6) | 1.4 (0.4- 3.5) |
| ABER | 0.0 (0.0- NE) | 4.0 (1.8- 7.5) | 0.0 (0.0-1.4) | 4.8 (3.2- 6.9) | 0.8 (0.3-1.9) | 3.1 (1.4- 5.8) |
| PSYC | 0.7 (0.0-3.7) | 4.5 (1.9- 8.9) | 0.4 (0.0-2.2) | 4.8 (3.2- 6.9) | 0.5 (0.1-1.5) | 2.1 (0.8- 4.5) |
| DELU | 0.7 (0.0-3.7) | 4.2 (1.7- 8.5) | 0.4 (0.0-2.2) | 4.1 (2.6- 6.1) | 0.5 (0.1-1.5) | 2.1 (0.8- 4.5) |
| HALL | 0.0 (0.0- NE) | 0.7 (0.2- 1.6) | 0.0 (0.0-1.4) | 1.8 (0.9- 3.4) | 0.0 (0.0-0.6) | 0.3 (0.0- 1.9) |
| ELAT | 0.0 (0.0- NE) | 0.0 (0.0- NE) | 0.0 (0.0-1.4) | 0.0 (0.0- 0.7) | 0.3 (0.0-1.2) | 1.4 (0.4- 3.5) |

Displayed are prevalence (%) and 95% confidence intervals. Survey estimates for ADAMS are based on survey population weights, confidence intervals are based on survey clusters and survey strata.
NA, Not applicable; NE Not estimable; NPS, Neuropsychiatric symptom.
Prevalence for NPI product scores greater than 0 is equivalent to prevalence with NPI severity scores > 0.
ABER, Aberrant motor behavior; AGIT, Agitation/aggression; ANXI, Anxiety; APAT, Apathy/indifference; APPE, Appetite/eating changes; DELU, Delusions; DEPR, Depression/dysphoria; DISI, Disinhibition; ELAT, Elation/euphoria; HALU, Hallucinations; IRRI, Irritability/lability; PSYC, Psychotic symptoms (hallucinations and/or delusions); SLEE, Nighttime behavioral disturbances.

### Supplement section 5. Prevalence of pairs of NPI domains

Supplemental Table 15. Prevalence of pairs of neuropsychiatric symptoms with the NPI severity score > 0

| NPS pair | | ADAMS survey | ADAMS | ADNI | NACC |
|---|---|---|---|---|---|
| DEPR | IRRI | 5.2 (2.7-8.8) | 5.4 (3.9-7.2) | 10.7 (8.7-12.9) | 14.2 (13.8-14.6) |
| DEPR | ANXI | 4.9 (2.9-7.8) | 6.6 (5.0-8.6) | 8.0 (6.3-10.0) | 15.1 (14.6-15.5) |
| DEPR | APAT | 5.4 (3.8-7.3) | 6.8 (5.1-8.7) | 9.4 (7.6-11.6) | 13.8 (13.3-14.2) |
| DEPR | SLEE | NA | NA | 8.3 (6.6-10.3) | 11.0 (10.6-11.4) |
| DEPR | AGIT | 5.2 (2.5-9.3) | 6.0 (4.5-7.9) | 7.0 (5.4- 8.9) | 11.1 (10.7-11.5) |
| DEPR | APPE | NA | NA | 6.0 (4.5- 7.7) | 8.7 ( 8.4- 9.1) |
| DEPR | DISI | 2.1 (0.8-4.6) | 2.8 (1.7-4.1) | 5.2 (3.8- 6.8) | 6.7 ( 6.4- 7.0) |
| DEPR | ABER | 1.3 (0.5-3.0) | 1.9 (1.1-3.1) | 3.4 (2.3- 4.8) | 6.0 ( 5.7- 6.3) |
| DEPR | PSYC | 2.3 (1.1-4.2) | 4.3 (3.0-5.9) | 2.6 (1.6- 3.9) | 6.0 ( 5.7- 6.3) |
| DEPR | DELU | 2.0 (0.8-3.9) | 2.9 (1.8-4.3) | 2.4 (1.5- 3.6) | 4.9 ( 4.6- 5.2) |
| DEPR | HALL | 1.2 (0.5-2.4) | 2.8 (1.7-4.1) | 0.7 (0.2- 1.5) | 2.5 ( 2.3- 2.7) |
| DEPR | ELAT | 0.5 (0.0-2.7) | 0.1 (0.0-0.7) | 1.1 (0.5- 2.1) | 1.8 ( 1.6- 1.9) |
| IRRI | ANXI | 2.5 (1.0-5.2) | 4.4 (3.1-6.0) | 7.5 (5.9- 9.5) | 13.1 (12.7-13.5) |
| IRRI | APAT | 2.5 (1.2-4.6) | 3.5 (2.3-5.0) | 8.8 (7.0-10.8) | 12.7 (12.3-13.1) |
| IRRI | SLEE | NA | NA | 6.6 (5.1- 8.5) | 10.3 (10.0-10.7) |
| IRRI | AGIT | 5.1 (2.8-8.5) | 7.3 (5.6-9.3) | 9.6 (7.7-11.7) | 14.5 (14.1-14.9) |
| IRRI | APPE | NA | NA | 4.7 (3.4- 6.3) | 8.3 ( 8.0- 8.6) |
| IRRI | DISI | 2.2 (1.0-4.4) | 3.1 (2.0-4.6) | 5.8 (4.4- 7.6) | 8.6 ( 8.3- 8.9) |
| IRRI | ABER | 1.1 (0.4-2.3) | 2.4 (1.4-3.7) | 2.4 (1.5- 3.6) | 6.6 ( 6.3- 6.9) |
| IRRI | PSYC | 1.7 (0.7-3.4) | 4.1 (2.9-5.8) | 2.4 (1.5- 3.6) | 6.0 ( 5.8- 6.3) |
| IRRI | DELU | 1.4 (0.5-3.2) | 2.8 (1.7-4.1) | 2.0 (1.2- 3.2) | 5.1 ( 4.8- 5.4) |
| IRRI | HALL | 1.0 (0.3-2.3) | 2.8 (1.7-4.1) | 0.6 (0.2- 1.3) | 2.4 ( 2.2- 2.6) |
| IRRI | ELAT | 0.1 (0.0-0.2) | 0.3 (0.0-0.9) | 1.1 (0.5- 2.1) | 2.2 ( 2.0- 2.3) |
| ANXI | APAT | 2.7 (1.2-5.0) | 4.1 (2.9-5.8) | 6.4 (4.9- 8.2) | 11.9 (11.5-12.3) |
| ANXI | SLEE | NA | NA | 5.6 (4.2- 7.3) | 10.0 ( 9.6-10.3) |
| ANXI | AGIT | 2.2 (1.0-4.2) | 4.4 (3.1-6.0) | 6.2 (4.7- 8.0) | 10.3 ( 9.9-10.7) |
| ANXI | APPE | NA | NA | 3.6 (2.5- 5.0) | 7.9 ( 7.6- 8.2) |
| ANXI | DISI | 1.1 (0.4-2.3) | 2.6 (1.6-4.0) | 3.9 (2.8- 5.4) | 6.9 ( 6.6- 7.2) |
| ANXI | ABER | 0.7 (0.4-1.2) | 2.4 (1.4-3.7) | 2.6 (1.6- 3.9) | 6.6 ( 6.3- 6.9) |

| | | | | | |
|---|---|---|---|---|---|
| ANXI | PSYC | 0.9 (0.6-1.4) | 3.4 (2.2-4.9) | 2.1 (1.3- 3.3) | 5.9 ( 5.6- 6.2) |
| ANXI | DELU | 0.7 (0.5-1.0) | 2.4 (1.4-3.7) | 1.6 (0.9- 2.6) | 4.7 ( 4.5- 5.0) |
| ANXI | HALL | 0.7 (0.4-1.1) | 2.4 (1.4-3.7) | 0.8 (0.3- 1.6) | 2.7 ( 2.5- 2.9) |
| ANXI | ELAT | 0.5 (0.0-2.6) | 0.1 (0.0-0.7) | 0.7 (0.2- 1.5) | 1.9 ( 1.7- 2.1) |
| APAT | SLEE | NA | NA | 5.2 (3.8- 6.8) | 10.2 ( 9.8-10.6) |
| APAT | AGIT | 2.8 (1.3-5.2) | 3.9 (2.7-5.5) | 6.0 (4.5- 7.7) | 10.7 (10.3-11.1) |
| APAT | APPE | NA | NA | 5.5 (4.1- 7.2) | 9.3 ( 9.0- 9.7) |
| APAT | DISI | 1.6 (0.6-3.5) | 2.3 (1.3-3.5) | 4.7 (3.4- 6.3) | 7.8 ( 7.5- 8.1) |
| APAT | ABER | 1.3 (0.4-3.2) | 2.3 (1.3-3.5) | 3.6 (2.5- 5.0) | 7.2 ( 6.8- 7.5) |
| APAT | PSYC | 1.6 (0.6-3.6) | 3.5 (2.3-5.0) | 2.2 (1.4- 3.5) | 6.0 ( 5.7- 6.3) |
| APAT | DELU | 1.4 (0.4-3.4) | 2.8 (1.7-4.1) | 1.7 (0.9- 2.8) | 4.8 ( 4.5- 5.0) |
| APAT | HALL | 0.5 (0.2-1.0) | 2.0 (1.1-3.2) | 1.0 (0.5- 1.9) | 2.8 ( 2.6- 3.0) |
| APAT | ELAT | 0.9 (0.1-3.4) | 0.4 (0.1-1.1) | 0.9 (0.4- 1.8) | 2.1 ( 1.9- 2.3) |
| SLEE | AGIT | NA | NA | 4.3 (3.0- 5.8) | 8.5 ( 8.1- 8.8) |
| SLEE | APPE | NA | NA | 3.7 (2.6- 5.2) | 7.8 ( 7.5- 8.1) |
| SLEE | DISI | NA | NA | 3.1 (2.1- 4.5) | 5.8 ( 5.5- 6.1) |
| SLEE | ABER | NA | NA | 1.6 (0.9- 2.6) | 5.3 ( 5.1- 5.6) |
| SLEE | PSYC | NA | NA | 1.2 (0.6- 2.2) | 4.9 ( 4.7- 5.2) |
| SLEE | DELU | NA | NA | 0.9 (0.4- 1.8) | 3.7 ( 3.5- 4.0) |
| SLEE | HALL | NA | NA | 0.4 (0.1- 1.1) | 2.6 ( 2.4- 2.8) |
| SLEE | ELAT | NA | NA | 0.7 (0.2- 1.5) | 1.6 ( 1.5- 1.8) |
| AGIT | APPE | NA | NA | 4.0 (2.9- 5.6) | 7.2 ( 6.9- 7.5) |
| AGIT | DISI | 1.9 (0.9-3.6) | 3.0 (1.9-4.4) | 5.0 (3.6- 6.6) | 7.8 ( 7.4- 8.1) |
| AGIT | ABER | 1.5 (0.6-3.3) | 3.1 (2.0-4.6) | 2.4 (1.5- 3.6) | 6.2 ( 5.9- 6.5) |
| AGIT | PSYC | 2.1 (1.0-4.0) | 4.8 (3.4-6.5) | 1.9 (1.1- 3.0) | 6.2 ( 6.0- 6.5) |
| AGIT | DELU | 1.9 (0.8-3.8) | 3.8 (2.6-5.3) | 1.7 (0.9- 2.8) | 5.3 ( 5.0- 5.6) |
| AGIT | HALL | 0.9 (0.3-2.1) | 2.6 (1.6-4.0) | 0.4 (0.1- 1.1) | 2.5 ( 2.3- 2.7) |
| AGIT | ELAT | 0.1 (0.0-0.2) | 0.4 (0.1-1.1) | 1.1 (0.5- 2.1) | 2.0 ( 1.8- 2.2) |
| APPE | DISI | NA | NA | 2.5 (1.6- 3.7) | 5.5 ( 5.2- 5.8) |
| APPE | ABER | NA | NA | 1.8 (1.0- 2.9) | 5.0 ( 4.8- 5.3) |
| APPE | PSYC | NA | NA | 1.6 (0.9- 2.6) | 4.0 ( 3.8- 4.2) |
| APPE | DELU | NA | NA | 1.2 (0.6- 2.2) | 3.2 ( 3.0- 3.4) |
| APPE | HALL | NA | NA | 0.6 (0.2- 1.3) | 1.8 ( 1.6- 1.9) |
| APPE | ELAT | NA | NA | 0.8 (0.3- 1.6) | 1.7 ( 1.6- 1.9) |
| DISI | ABER | 0.8 (0.2-2.4) | 1.5 (0.8-2.6) | 2.6 (1.6- 3.9) | 5.1 ( 4.9- 5.4) |

| | | | | | |
|---|---|---|---|---|---|
| DISI | PSYC | 1.8 (0.6-3.9) | 2.4 (1.4-3.7) | 1.1 (0.5- 2.1) | 4.1 ( 3.8- 4.3) |
| DISI | DELU | 1.6 (0.6-3.7) | 2.0 (1.1-3.2) | 1.0 (0.5- 1.9) | 3.5 ( 3.3- 3.7) |
| DISI | HALL | 1.1 (0.2-2.9) | 1.4 (0.7-2.4) | 0.2 (0.0- 0.8) | 1.6 ( 1.5- 1.8) |
| DISI | ELAT | 0.0 (0.0- NE) | 0.0 (0.0-0.5) | 1.3 (0.7- 2.3) | 2.2 ( 2.0- 2.4) |
| ABER | PSYC | 1.7 (0.7-3.2) | 3.4 (2.2-4.9) | 1.0 (0.5- 1.9) | 4.0 ( 3.8- 4.3) |
| ABER | DELU | 1.4 (0.6-2.9) | 2.4 (1.4-3.7) | 0.8 (0.3- 1.6) | 3.3 ( 3.1- 3.5) |
| ABER | HALL | 0.7 (0.4-1.3) | 2.3 (1.3-3.5) | 0.4 (0.1- 1.1) | 1.9 ( 1.8- 2.1) |
| ABER | ELAT | 0.2 (0.0-0.8) | 0.5 (0.1-1.3) | 0.4 (0.1- 1.1) | 1.6 ( 1.5- 1.8) |
| PSYC | ELAT | 0.2 (0.0-0.8) | 0.6 (0.2-1.5) | 0.4 (0.1- 1.1) | 1.1 ( 1.0- 1.3) |
| DELU | HALL | 1.6 (0.6-3.3) | 3.0 (1.9-4.4) | 0.6 (0.2- 1.3) | 2.3 ( 2.2- 2.5) |
| DELU | ELAT | 0.2 (0.0-0.9) | 0.3 (0.0-0.9) | 0.3 (0.1- 1.0) | 1.0 ( 0.9- 1.1) |
| HALL | ELAT | 0.2 (0.0-0.8) | 0.6 (0.2-1.5) | 0.2 (0.0- 0.8) | 0.5 ( 0.4- 0.6) |

Displayed are prevalence (%) and 95% confidence intervals. Survey estimates for ADAMS are based on survey population weights, confidence intervals are based on survey clusters and survey strata.

NA, Not applicable; NE Not estimable; NPS, Neuropsychiatric symptom.

ABER, Aberrant motor behavior; AGIT, Agitation/aggression; ANXI, Anxiety; APAT, Apathy/indifference; APPE, Appetite/eating changes; DELU, Delusions; DEPR, Depression/dysphoria; DISI, Disinhibition; ELAT, Elation/euphoria; HALU, Hallucinations; IRRI, Irritability/lability; PSYC, Psychotic symptoms (hallucinations and/or delusions); SLEE, Nighttime behavioral disturbances.

Supplemental Table 16. Prevalence of pairs of neuropsychiatric symptoms with the NPI severity score > 1

| NPS pair | | ADAMS survey | ADAMS | ADNI | NACC |
|---|---|---|---|---|---|
| DEPR | IRRI | 1.2 (0.4-3.6) | 1.4 (0.7-2.4) | 2.0 (1.2-3.2) | 3.8 (3.5-4.0) |
| DEPR | ANXI | 1.1 (0.4-2.8) | 1.9 (1.1-3.1) | 1.3 (0.7-2.3) | 4.7 (4.4-4.9) |
| DEPR | APAT | 1.9 (0.9-4.1) | 2.6 (1.6-4.0) | 1.2 (0.6-2.2) | 4.2 (4.0-4.5) |
| DEPR | SLEE | NA | NA | 1.2 (0.6-2.2) | 3.2 (3.0-3.4) |
| DEPR | AGIT | 1.4 (0.6-3.6) | 1.5 (0.8-2.6) | 0.7 (0.2-1.5) | 3.3 (3.1-3.6) |
| DEPR | APPE | NA | NA | 1.0 (0.5-1.9) | 2.3 (2.1-2.4) |
| DEPR | DISI | 1.0 (0.3-3.2) | 0.6 (0.2-1.5) | 0.7 (0.2-1.5) | 1.9 (1.7-2.0) |
| DEPR | ABER | 0.9 (0.3-2.9) | 0.5 (0.1-1.3) | 0.2 (0.0-0.8) | 1.7 (1.5-1.9) |
| DEPR | PSYC | 1.1 (0.4-3.0) | 1.3 (0.6-2.3) | 0.6 (0.2-1.3) | 1.7 (1.6-1.9) |
| DEPR | DELU | 1.0 (0.3-3.0) | 1.1 (0.5-2.1) | 0.6 (0.2-1.3) | 1.4 (1.3-1.6) |
| DEPR | HALL | 0.1 (0.0-0.5) | 0.5 (0.1-1.3) | 0.0 (0.0-0.4) | 0.7 (0.6-0.8) |
| DEPR | ELAT | 0.0 (0.0- NE) | 0.0 (0.0-0.5) | 0.1 (0.0-0.6) | 0.4 (0.4-0.5) |
| IRRI | ANXI | 0.3 (0.1-0.7) | 1.3 (0.6-2.3) | 1.6 (0.9-2.6) | 4.1 (3.9-4.4) |
| IRRI | APAT | 1.0 (0.3-2.9) | 1.4 (0.7-2.4) | 1.7 (0.9-2.8) | 4.0 (3.8-4.2) |
| IRRI | SLEE | NA | NA | 1.2 (0.6-2.2) | 3.2 (3.0-3.4) |
| IRRI | AGIT | 1.8 (0.8-3.8) | 3.0 (1.9-4.4) | 1.7 (0.9-2.8) | 5.0 (4.7-5.2) |
| IRRI | APPE | NA | NA | 1.2 (0.6-2.2) | 2.5 (2.3-2.7) |
| IRRI | DISI | 0.7 (0.2-2.6) | 1.3 (0.6-2.3) | 1.3 (0.7-2.3) | 3.1 (2.9-3.3) |
| IRRI | ABER | 0.6 (0.1-2.7) | 0.8 (0.3-1.6) | 0.4 (0.1-1.1) | 2.4 (2.2-2.5) |
| IRRI | PSYC | 0.8 (0.3-2.5) | 1.6 (0.9-2.8) | 0.9 (0.4-1.8) | 2.0 (1.8-2.2) |
| IRRI | DELU | 0.7 (0.2-2.5) | 1.4 (0.7-2.4) | 0.8 (0.3-1.6) | 1.7 (1.6-1.9) |
| IRRI | HALL | 0.2 (0.1-0.5) | 0.8 (0.3-1.6) | 0.1 (0.0-0.6) | 0.7 (0.6-0.8) |
| IRRI | ELAT | 0.0 (0.0- NE) | 0.0 (0.0-0.5) | 0.2 (0.0-0.8) | 0.7 (0.6-0.8) |
| ANXI | APAT | 0.8 (0.4-1.9) | 1.9 (1.1-3.1) | 1.3 (0.7-2.3) | 3.9 (3.7-4.1) |
| ANXI | SLEE | NA | NA | 0.6 (0.2-1.3) | 3.2 (2.9-3.4) |
| ANXI | AGIT | 0.3 (0.2-0.5) | 1.3 (0.6-2.3) | 1.0 (0.5-1.9) | 3.4 (3.2-3.7) |
| ANXI | APPE | NA | NA | 0.9 (0.4-1.8) | 2.3 (2.1-2.5) |
| ANXI | DISI | 0.2 (0.1-0.6) | 0.8 (0.3-1.6) | 0.4 (0.1-1.1) | 2.3 (2.1-2.5) |
| ANXI | ABER | 0.3 (0.2-0.5) | 1.0 (0.4-2.0) | 0.3 (0.1-1.0) | 2.3 (2.1-2.5) |
| ANXI | PSYC | 0.3 (0.1-0.7) | 1.1 (0.5-2.1) | 0.4 (0.1-1.1) | 2.0 (1.8-2.1) |
| ANXI | DELU | 0.2 (0.1-0.6) | 1.0 (0.4-2.0) | 0.4 (0.1-1.1) | 1.6 (1.5-1.8) |
| ANXI | HALL | 0.2 (0.0-0.6) | 0.6 (0.2-1.5) | 0.1 (0.0-0.6) | 0.8 (0.7-1.0) |
| ANXI | ELAT | 0.5 (0.1-3.4) | 0.1 (0.0-0.7) | 0.1 (0.0-0.6) | 0.6 (0.5-0.7) |

| | | | | | |
|---|---|---|---|---|---|
| APAT | SLEE | NA | NA | 1.0 (0.5-1.9) | 3.8 (3.6-4.0) |
| APAT | AGIT | 1.3 (0.6-2.9) | 1.9 (1.1-3.1) | 1.7 (0.9-2.8) | 3.8 (3.6-4.0) |
| APAT | APPE | NA | NA | 2.0 (1.2-3.2) | 3.3 (3.1-3.5) |
| APAT | DISI | 0.6 (0.1-2.7) | 0.6 (0.2-1.5) | 0.7 (0.2-1.5) | 2.9 (2.7-3.1) |
| APAT | ABER | 0.9 (0.3-3.2) | 1.0 (0.4-2.0) | 0.9 (0.4-1.8) | 2.9 (2.7-3.1) |
| APAT | PSYC | 1.1 (0.3-3.3) | 1.3 (0.6-2.3) | 0.8 (0.3-1.6) | 2.0 (1.8-2.2) |
| APAT | DELU | 1.0 (0.3-3.3) | 1.1 (0.5-2.1) | 0.7 (0.2-1.5) | 1.7 (1.5-1.8) |
| APAT | HALL | 0.1 (0.0-0.5) | 0.4 (0.1-1.1) | 0.3 (0.1-1.0) | 0.8 (0.7-1.0) |
| APAT | ELAT | 0.0 (0.0- NE) | 0.0 (0.0-0.5) | 0.1 (0.0-0.6) | 0.7 (0.6-0.8) |
| SLEE | AGIT | NA | NA | 0.2 (0.0-0.8) | 2.8 (2.6-3.0) |
| SLEE | APPE | NA | NA | 0.8 (0.3-1.6) | 2.7 (2.5-2.9) |
| SLEE | DISI | NA | NA | 0.6 (0.2-1.3) | 2.0 (1.9-2.2) |
| SLEE | ABER | NA | NA | 0.6 (0.2-1.3) | 2.1 (1.9-2.3) |
| SLEE | PSYC | NA | NA | 0.2 (0.0-0.8) | 1.7 (1.6-1.9) |
| SLEE | DELU | NA | NA | 0.1 (0.0-0.6) | 1.3 (1.2-1.4) |
| SLEE | HALL | NA | NA | 0.1 (0.0-0.6) | 0.9 (0.8-1.1) |
| SLEE | ELAT | NA | NA | 0.1 (0.0-0.6) | 0.5 (0.4-0.6) |
| AGIT | APPE | NA | NA | 0.8 (0.3-1.6) | 2.2 (2.0-2.4) |
| AGIT | DISI | 0.7 (0.2-2.6) | 1.1 (0.5-2.1) | 0.9 (0.4-1.8) | 2.8 (2.6-3.0) |
| AGIT | ABER | 0.7 (0.2-2.5) | 1.4 (0.7-2.4) | 0.2 (0.0-0.8) | 2.2 (2.1-2.4) |
| AGIT | PSYC | 0.8 (0.2-2.5) | 1.8 (1.0-2.9) | 0.7 (0.2-1.5) | 2.2 (2.0-2.4) |
| AGIT | DELU | 0.7 (0.2-2.5) | 1.6 (0.9-2.8) | 0.7 (0.2-1.5) | 1.9 (1.8-2.1) |
| AGIT | HALL | 0.1 (0.0-0.3) | 0.6 (0.2-1.5) | 0.0 (0.0-0.4) | 0.7 (0.6-0.9) |
| AGIT | ELAT | 0.0 (0.0- NE) | 0.0 (0.0-0.5) | 0.2 (0.0-0.8) | 0.7 (0.6-0.8) |
| APPE | DISI | NA | NA | 0.7 (0.2-1.5) | 1.9 (1.8-2.1) |
| APPE | ABER | NA | NA | 0.6 (0.2-1.3) | 1.9 (1.7-2.0) |
| APPE | PSYC | NA | NA | 0.4 (0.1-1.1) | 1.2 (1.1-1.4) |
| APPE | DELU | NA | NA | 0.4 (0.1-1.1) | 1.0 (0.9-1.2) |
| APPE | HALL | NA | NA | 0.1 (0.0-0.6) | 0.5 (0.4-0.6) |
| APPE | ELAT | NA | NA | 0.2 (0.0-0.8) | 0.6 (0.6-0.8) |
| DISI | ABER | 0.6 (0.1-2.6) | 0.9 (0.4-1.8) | 0.3 (0.1-1.0) | 2.0 (1.8-2.2) |
| DISI | PSYC | 0.7 (0.2-2.6) | 0.9 (0.4-1.8) | 0.4 (0.1-1.1) | 1.3 (1.2-1.5) |
| DISI | DELU | 0.6 (0.1-2.7) | 0.8 (0.3-1.6) | 0.4 (0.1-1.1) | 1.2 (1.0-1.3) |
| DISI | HALL | 0.1 (0.0-0.5) | 0.4 (0.1-1.1) | 0.0 (0.0-0.4) | 0.5 (0.4-0.6) |
| DISI | ELAT | 0.0 (0.0- NE) | 0.0 (0.0-0.5) | 0.4 (0.1-1.1) | 0.8 (0.7-0.9) |

| | | | | | |
|---|---|---|---|---|---|
| ABER | PSYC | 1.0 (0.3-2.9) | 1.3 (0.6-2.3) | 0.3 (0.1-1.0) | 1.4 (1.3-1.6) |
| ABER | DELU | 1.0 (0.3-2.9) | 1.1 (0.5-2.1) | 0.2 (0.0-0.8) | 1.2 (1.1-1.4) |
| ABER | HALL | 0.1 (0.0-0.2) | 0.4 (0.1-1.1) | 0.2 (0.0-0.8) | 0.6 (0.5-0.7) |
| ABER | ELAT | 0.0 (0.0- NE) | 0.0 (0.0-0.5) | 0.2 (0.0-0.8) | 0.6 (0.5-0.7) |
| PSYC | ELAT | 0.0 (0.0- NE) | 0.0 (0.0-0.5) | 0.3 (0.1-1.0) | 0.3 (0.3-0.4) |
| DELU | HALL | 0.3 (0.1-0.6) | 1.1 (0.5-2.1) | 0.2 (0.0-0.8) | 0.9 (0.8-1.0) |
| DELU | ELAT | 0.0 (0.0- NE) | 0.0 (0.0-0.5) | 0.3 (0.1-1.0) | 0.3 (0.2-0.4) |
| HALL | ELAT | 0.0 (0.0- NE) | 0.0 (0.0-0.5) | 0.0 (0.0-0.4) | 0.1 (0.1-0.2) |

Displayed are prevalence (%) and 95% confidence intervals. Survey estimates for ADAMS are based on survey population weights, confidence intervals are based on survey clusters and survey strata.

NA, Not applicable, NE Not estimable; NPS, Neuropsychiatric symptom.

ABER, Aberrant motor behavior; AGIT, Agitation/aggression; ANXI, Anxiety; APAT, Apathy/indifference; APPE, Appetite/eating changes; DELU, Delusions; DEPR, Depression/dysphoria; DISI, Disinhibition; ELAT, Elation/euphoria; HALU, Hallucinations; IRRI, Irritability/lability; PSYC, Psychotic symptoms (hallucinations and/or delusions); SLEE, Nighttime behavioral disturbances.

Supplemental Table 17. Prevalence of pairs of neuropsychiatric symptoms with the NPI severity score > 1 per MMSE stratum (ADAMS and ADNI studies only)

| NPS pair | | ADAMS survey | | ADAMS | | ADNI | |
|---|---|---|---|---|---|---|---|
| | | MMSE > 26 | MMS ≤ 26 | MMSE > 26 | MMSE ≤ 26 | MMSE > 26 | MMSE ≤ 26 |
| DEPR | IRRI | 0.0 (0.0- NE) | 3.1 (1.1-8.6) | 0.0 (0.0-1.4) | 2.0 (1.0-3.6) | 1.2 (0.5-2.4) | 3.8 (1.9-6.7) |
| DEPR | ANXI | 0.7 (0.1-4.3) | 1.7 (0.6-4.5) | 0.8 (0.1-2.8) | 2.4 (1.3-4.1) | 0.8 (0.3-1.9) | 2.4 (1.0-4.9) |
| DEPR | APAT | 0.4 (0.0-2.6) | 4.3 (1.9-9.8) | 0.4 (0.0-2.2) | 3.7 (2.3-5.6) | 0.7 (0.2-1.7) | 2.4 (1.0-4.9) |
| DEPR | SLEE | NA | NA | NA | NA | 1.0 (0.4-2.2) | 1.7 (0.6-4.0) |
| DEPR | AGIT | 0.4 (0.1-2.3) | 3.0 (1.1-8.3) | 0.8 (0.1-2.8) | 1.8 (0.9-3.4) | 0.5 (0.1-1.5) | 1.0 (0.2-3.0) |
| DEPR | APPE | NA | NA | NA | NA | 0.7 (0.2-1.7) | 1.7 (0.6-4.0) |
| DEPR | DISI | 0.0 (0.0- NE) | 2.5 (0.8-7.5) | 0.0 (0.0-1.4) | 0.9 (0.3-2.1) | 0.3 (0.0-1.2) | 1.4 (0.4-3.5) |
| DEPR | ABER | 0.0 (0.0- NE) | 2.2 (0.7-6.9) | 0.0 (0.0-1.4) | 0.7 (0.2-1.9) | 0.2 (0.0-0.9) | 0.3 (0.0-1.9) |
| DEPR | PSYC | 0.0 (0.0- NE) | 2.7 (1.0-7.3) | 0.0 (0.0-1.4) | 1.8 (0.9-3.4) | 0.5 (0.1-1.5) | 0.7 (0.1-2.5) |
| DEPR | DELU | 0.0 (0.0- NE) | 2.6 (0.9-7.2) | 0.0 (0.0-1.4) | 1.7 (0.8-3.1) | 0.5 (0.1-1.5) | 0.7 (0.1-2.5) |
| DEPR | HALL | 0.0 (0.0- NE) | 0.3 (0.1-1.2) | 0.0 (0.0-1.4) | 0.7 (0.2-1.9) | 0.0 (0.0-0.6) | 0.0 (0.0-1.3) |
| DEPR | ELAT | 0.0 (0.0- NE) | 0.0 (0.0- NE) | 0.0 (0.0-1.4) | 0.0 (0.0-0.7) | 0.0 (0.0-0.6) | 0.3 (0.0-1.9) |
| IRRI | ANXI | 0.0 (0.0- NE) | 0.8 (0.3-1.9) | 0.0 (0.0-1.4) | 1.8 (0.9-3.4) | 1.3 (0.6-2.6) | 2.1 (0.8-4.5) |
| IRRI | APAT | 0.0 (0.0- NE) | 2.5 (0.8-7.2) | 0.0 (0.0-1.4) | 2.0 (1.0-3.6) | 1.3 (0.6-2.6) | 2.4 (1.0-4.9) |
| IRRI | SLEE | NA | NA | NA | NA | 1.0 (0.4-2.2) | 1.7 (0.6-4.0) |
| IRRI | AGIT | 0.0 (0.0- NE) | 4.4 (2.1-9.3) | 0.0 (0.0-1.4) | 4.4 (2.9-6.5) | 1.7 (0.8-3.0) | 1.7 (0.6-4.0) |
| IRRI | APPE | NA | NA | NA | NA | 0.5 (0.1-1.5) | 2.8 (1.2-5.4) |
| IRRI | DISI | 0.0 (0.0- NE) | 1.8 (0.5-6.2) | 0.0 (0.0-1.4) | 1.8 (0.9-3.4) | 1.0 (0.4-2.2) | 2.1 (0.8-4.5) |
| IRRI | ABER | 0.0 (0.0- NE) | 1.4 (0.3-6.5) | 0.0 (0.0-1.4) | 1.1 (0.4-2.4) | 0.2 (0.0-0.9) | 1.0 (0.2-3.0) |
| IRRI | PSYC | 0.0 (0.0- NE) | 2.0 (0.7-6.0) | 0.0 (0.0-1.4) | 2.4 (1.3-4.1) | 0.7 (0.2-1.7) | 1.4 (0.4-3.5) |
| IRRI | DELU | 0.0 (0.0- NE) | 1.8 (0.5-6.0) | 0.0 (0.0-1.4) | 2.0 (1.0-3.6) | 0.5 (0.1-1.5) | 1.4 (0.4-3.5) |
| IRRI | HALL | 0.0 (0.0- NE) | 0.4 (0.1-1.2) | 0.0 (0.0-1.4) | 1.1 (0.4-2.4) | 0.2 (0.0-0.9) | 0.0 (0.0-1.3) |
| IRRI | ELAT | 0.0 (0.0- NE) | 0.0 (0.0- NE) | 0.0 (0.0-1.4) | 0.0 (0.0-0.7) | 0.0 (0.0-0.6) | 0.7 (0.1-2.5) |
| ANXI | APAT | 0.1 (0.0-0.4) | 2.0 (0.9-4.6) | 0.4 (0.0-2.2) | 2.6 (1.4-4.3) | 0.3 (0.0-1.2) | 3.5 (1.7-6.3) |

| | | | | | | | |
|---|---|---|---|---|---|---|---|
| ANXI | SLEE | NA | NA | NA | NA | 0.5 (0.1-1.5) | 0.7 (0.1-2.5) |
| ANXI | AGIT | 0.0 (0.0- NE) | 0.8 (0.5-1.4) | 0.0 (0.0-1.4) | 1.8 (0.9-3.4) | 0.7 (0.2-1.7) | 1.7 (0.6-4.0) |
| ANXI | APPE | NA | NA | NA | NA | 0.5 (0.1-1.5) | 1.7 (0.6-4.0) |
| ANXI | DISI | 0.0 (0.0- NE) | 0.5 (0.2-1.5) | 0.0 (0.0-1.4) | 1.1 (0.4-2.4) | 0.5 (0.1-1.5) | 0.3 (0.0-1.9) |
| ANXI | ABER | 0.0 (0.0- NE) | 0.7 (0.4-1.4) | 0.0 (0.0-1.4) | 1.5 (0.6-2.9) | 0.2 (0.0-0.9) | 0.7 (0.1-2.5) |
| ANXI | PSYC | 0.0 (0.0- NE) | 0.8 (0.3-1.9) | 0.0 (0.0-1.4) | 1.7 (0.8-3.1) | 0.2 (0.0-0.9) | 1.0 (0.2-3.0) |
| ANXI | DELU | 0.0 (0.0- NE) | 0.6 (0.2-1.6) | 0.0 (0.0-1.4) | 1.5 (0.6-2.9) | 0.2 (0.0-0.9) | 1.0 (0.2-3.0) |
| ANXI | HALL | 0.0 (0.0- NE) | 0.4 (0.1-1.4) | 0.0 (0.0-1.4) | 0.9 (0.3-2.1) | 0.0 (0.0-0.6) | 0.3 (0.0-1.9) |
| ANXI | ELAT | 0.8 (0.1-5.5) | 0.0 (0.0- NE) | 0.4 (0.0-2.2) | 0.0 (0.0-0.7) | 0.2 (0.0-0.9) | 0.0 (0.0-1.3) |
| APAT | SLEE | NA | NA | NA | NA | 0.7 (0.2-1.7) | 1.7 (0.6-4.0) |
| APAT | AGIT | 0.4 (0.0-2.6) | 2.8 (1.1-6.6) | 0.4 (0.0-2.2) | 2.6 (1.4-4.3) | 1.0 (0.4-2.2) | 3.1 (1.4-5.8) |
| APAT | APPE | NA | NA | NA | NA | 0.2 (0.0-0.9) | 5.9 (3.5-9.3) |
| APAT | DISI | 0.0 (0.0- NE) | 1.6 (0.4-6.4) | 0.0 (0.0-1.4) | 0.9 (0.3-2.1) | 0.7 (0.2-1.7) | 0.7 (0.1-2.5) |
| APAT | ABER | 0.0 (0.0- NE) | 2.4 (0.7-7.6) | 0.0 (0.0-1.4) | 1.5 (0.6-2.9) | 0.5 (0.1-1.5) | 1.7 (0.6-4.0) |
| APAT | PSYC | 0.0 (0.0- NE) | 2.6 (0.8-8.1) | 0.0 (0.0-1.4) | 1.8 (0.9-3.4) | 0.3 (0.0-1.2) | 1.7 (0.6-4.0) |
| APAT | DELU | 0.0 (0.0- NE) | 2.5 (0.7-8.0) | 0.0 (0.0-1.4) | 1.7 (0.8-3.1) | 0.2 (0.0-0.9) | 1.7 (0.6-4.0) |
| APAT | HALL | 0.0 (0.0- NE) | 0.3 (0.1-1.2) | 0.0 (0.0-1.4) | 0.6 (0.1-1.6) | 0.2 (0.0-0.9) | 0.7 (0.1-2.5) |
| APAT | ELAT | 0.0 (0.0- NE) | 0.0 (0.0- NE) | 0.0 (0.0-1.4) | 0.0 (0.0-0.7) | 0.2 (0.0-0.9) | 0.0 (0.0-1.3) |
| SLEE | AGIT | NA | NA | NA | NA | 0.2 (0.0-0.9) | 0.3 (0.0-1.9) |
| SLEE | APPE | NA | NA | NA | NA | 0.3 (0.0-1.2) | 1.7 (0.6-4.0) |
| SLEE | DISI | NA | NA | NA | NA | 0.5 (0.1-1.5) | 0.7 (0.1-2.5) |
| SLEE | ABER | NA | NA | NA | NA | 0.3 (0.0-1.2) | 1.0 (0.2-3.0) |
| SLEE | PSYC | NA | NA | NA | NA | 0.3 (0.0-1.2) | 0.0 (0.0-1.3) |
| SLEE | DELU | NA | NA | NA | NA | 0.2 (0.0-0.9) | 0.0 (0.0-1.3) |
| SLEE | HALL | NA | NA | NA | NA | 0.2 (0.0-0.9) | 0.0 (0.0-1.3) |
| SLEE | ELAT | NA | NA | NA | NA | 0.0 (0.0-0.6) | 0.3 (0.0-1.9) |
| AGIT | APPE | NA | NA | NA | NA | 0.3 (0.0-1.2) | 1.7 (0.6-4.0) |
| AGIT | DISI | 0.0 (0.0- NE) | 1.6 (0.4-6.2) | 0.0 (0.0-1.4) | 1.7 (0.8-3.1) | 1.2 (0.5-2.4) | 0.3 (0.0-1.9) |
| AGIT | ABER | 0.0 (0.0- NE) | 1.8 (0.5-6.0) | 0.0 (0.0-1.4) | 2.0 (1.0-3.6) | 0.0 (0.0-0.6) | 0.7 (0.1-2.5) |

| | | | | | | | |
|---|---|---|---|---|---|---|---|
| AGIT | PSYC | 0.0 (0.0- NE) | 1.9 (0.6-6.1) | 0.0 (0.0-1.4) | 2.6 (1.4-4.3) | 0.5 (0.1-1.5) | 1.0 (0.2-3.0) |
| AGIT | DELU | 0.0 (0.0- NE) | 1.9 (0.6-6.1) | 0.0 (0.0-1.4) | 2.4 (1.3-4.1) | 0.5 (0.1-1.5) | 1.0 (0.2-3.0) |
| AGIT | HALL | 0.0 (0.0- NE) | 0.3 (0.1-0.8) | 0.0 (0.0-1.4) | 0.9 (0.3-2.1) | 0.0 (0.0-0.6) | 0.0 (0.0-1.3) |
| AGIT | ELAT | 0.0 (0.0- NE) | 0.0 (0.0- NE) | 0.0 (0.0-1.4) | 0.0 (0.0-0.7) | 0.2 (0.0-0.9) | 0.3 (0.0-1.9) |
| APPE | DISI | NA | NA | NA | NA | 0.8 (0.3-1.9) | 0.3 (0.0-1.9) |
| APPE | ABER | NA | NA | NA | NA | 0.3 (0.0-1.2) | 1.0 (0.2-3.0) |
| APPE | PSYC | NA | NA | NA | NA | 0.2 (0.0-0.9) | 1.0 (0.2-3.0) |
| APPE | DELU | NA | NA | NA | NA | 0.2 (0.0-0.9) | 1.0 (0.2-3.0) |
| APPE | HALL | NA | NA | NA | NA | 0.0 (0.0-0.6) | 0.3 (0.0-1.9) |
| APPE | ELAT | NA | NA | NA | NA | 0.2 (0.0-0.9) | 0.3 (0.0-1.9) |
| DISI | ABER | 0.0 (0.0- NE) | 1.5 (0.3-6.3) | 0.0 (0.0-1.4) | 1.3 (0.5-2.6) | 0.2 (0.0-0.9) | 0.7 (0.1-2.5) |
| DISI | PSYC | 0.0 (0.0- NE) | 1.6 (0.4-6.3) | 0.0 (0.0-1.4) | 1.3 (0.5-2.6) | 0.3 (0.0-1.2) | 0.7 (0.1-2.5) |
| DISI | DELU | 0.0 (0.0- NE) | 1.5 (0.3-6.4) | 0.0 (0.0-1.4) | 1.1 (0.4-2.4) | 0.3 (0.0-1.2) | 0.7 (0.1-2.5) |
| DISI | HALL | 0.0 (0.0- NE) | 0.3 (0.1-1.2) | 0.0 (0.0-1.4) | 0.6 (0.1-1.6) | 0.0 (0.0-0.6) | 0.0 (0.0-1.3) |
| DISI | ELAT | 0.0 (0.0- NE) | 0.0 (0.0- NE) | 0.0 (0.0-1.4) | 0.0 (0.0-0.7) | 0.3 (0.0-1.2) | 0.7 (0.1-2.5) |
| ABER | PSYC | 0.0 (0.0- NE) | 2.5 (0.9-6.9) | 0.0 (0.0-1.4) | 1.8 (0.9-3.4) | 0.2 (0.0-0.9) | 0.7 (0.1-2.5) |
| ABER | DELU | 0.0 (0.0- NE) | 2.4 (0.8-6.9) | 0.0 (0.0-1.4) | 1.7 (0.8-3.1) | 0.0 (0.0-0.6) | 0.7 (0.1-2.5) |
| ABER | HALL | 0.0 (0.0- NE) | 0.2 (0.0-0.6) | 0.0 (0.0-1.4) | 0.6 (0.1-1.6) | 0.2 (0.0-0.9) | 0.3 (0.0-1.9) |
| ABER | ELAT | 0.0 (0.0- NE) | 0.0 (0.0- NE) | 0.0 (0.0-1.4) | 0.0 (0.0-0.7) | 0.2 (0.0-0.9) | 0.3 (0.0-1.9) |
| PSYC | ELAT | 0.0 (0.0- NE) | 0.0 (0.0- NE) | 0.0 (0.0-1.4) | 0.0 (0.0-0.7) | 0.2 (0.0-0.9) | 0.7 (0.1-2.5) |
| DELU | HALL | 0.0 (0.0- NE) | 0.7 (0.3-1.6) | 0.0 (0.0-1.4) | 1.7 (0.8-3.1) | 0.0 (0.0-0.6) | 0.7 (0.1-2.5) |
| DELU | ELAT | 0.0 (0.0- NE) | 0.0 (0.0- NE) | 0.0 (0.0-1.4) | 0.0 (0.0-0.7) | 0.2 (0.0-0.9) | 0.7 (0.1-2.5) |
| HALL | ELAT | 0.0 (0.0- NE) | 0.0 (0.0- NE) | 0.0 (0.0-1.4) | 0.0 (0.0-0.7) | 0.0 (0.0-0.6) | 0.0 (0.0-1.3) |

Displayed are prevalence (%) and 95% confidence intervals. Survey estimates for ADAMS are based on survey population weights, confidence intervals are based on survey clusters and survey strata.

NA, Not applicable, NE Not estimable; NPS, Neuropsychiatric symptom.

Prevalence of pairs of neuropsychiatric symptoms with the NPI severity score > 0 is reported in Table 2 (main text).

ABER, Aberrant motor behavior; AGIT, Agitation/aggression; ANXI, Anxiety; APAT, Apathy/indifference; APPE, Appetite/eating changes; DELU, Delusions; DEPR, Depression/dysphoria; DISI, Disinhibition; ELAT, Elation/euphoria; HALU, Hallucinations; IRRI, Irritability/lability; PSYC, Psychotic symptoms (hallucinations and/or delusions); SLEE, Nighttime behavioral disturbances.

Supplemental Table 18. Prevalence of pairs of neuropsychiatric symptoms with the NPI severity score > 0 per MMSE stratum (NACC study only)

| NPS pair | | NACC | | |
|---|---|---|---|---|
| | | 19 < MMSE ≤ 26 | 9 < MMSE ≤ 19 | MMSE ≤ 9 |
| DEPR | IRRI | 18.5 (17.6-19.4) | 22.7 (21.2-24.2) | 24.0 (21.4-26.8) |
| DEPR | ANXI | 19.5 (18.6-20.4) | 27.1 (25.5-28.7) | 27.9 (25.1-30.9) |
| DEPR | APAT | 19.5 (18.6-20.5) | 25.2 (23.7-26.8) | 31.1 (28.2-34.1) |
| DEPR | SLEE | 14.5 (13.7-15.4) | 15.9 (14.6-17.2) | 18.2 (15.8-20.7) |
| DEPR | AGIT | 14.7 (13.9-15.6) | 21.6 (20.1-23.1) | 25.6 (22.9-28.4) |
| DEPR | APPE | 12.7 (12.0-13.5) | 16.1 (14.8-17.5) | 15.3 (13.1-17.7) |
| DEPR | DISI | 9.0 ( 8.3- 9.7) | 13.4 (12.2-14.7) | 15.2 (13.0-17.6) |
| DEPR | ABER | 7.5 ( 6.9- 8.2) | 15.7 (14.4-17.1) | 20.1 (17.6-22.8) |
| DEPR | PSYC | 7.9 ( 7.3- 8.6) | 15.9 (14.6-17.3) | 22.0 (19.4-24.7) |
| DEPR | DELU | 6.5 ( 6.0- 7.1) | 13.1 (11.9-14.3) | 18.1 (15.7-20.6) |
| DEPR | HALL | 3.0 ( 2.6- 3.4) | 7.4 ( 6.5- 8.4) | 11.8 ( 9.8-14.0) |
| DEPR | ELAT | 2.1 ( 1.8- 2.5) | 3.5 ( 2.9- 4.3) | 5.3 ( 4.0- 6.9) |
| IRRI | ANXI | 17.1 (16.3-18.0) | 24.6 (23.1-26.2) | 30.1 (27.2-33.1) |
| IRRI | APAT | 17.6 (16.8-18.5) | 23.7 (22.1-25.2) | 30.4 (27.5-33.4) |
| IRRI | SLEE | 13.5 (12.7-14.3) | 15.7 (14.4-17.0) | 20.6 (18.1-23.3) |
| IRRI | AGIT | 19.4 (18.5-20.3) | 25.8 (24.3-27.4) | 33.0 (30.0-36.0) |
| IRRI | APPE | 12.3 (11.6-13.1) | 14.8 (13.6-16.2) | 16.3 (14.1-18.8) |
| IRRI | DISI | 11.7 (11.0-12.5) | 15.6 (14.4-17.0) | 21.0 (18.5-23.7) |
| IRRI | ABER | 8.3 ( 7.7- 8.9) | 16.0 (14.7-17.4) | 25.8 (23.0-28.6) |
| IRRI | PSYC | 8.1 ( 7.4- 8.7) | 15.7 (14.4-17.0) | 22.9 (20.3-25.7) |
| IRRI | DELU | 6.9 ( 6.4- 7.5) | 13.2 (12.0-14.5) | 18.6 (16.2-21.2) |
| IRRI | HALL | 2.5 ( 2.1- 2.9) | 7.0 ( 6.1- 8.0) | 14.4 (12.2-16.7) |
| IRRI | ELAT | 2.6 ( 2.2- 3.0) | 4.0 ( 3.4- 4.8) | 5.6 ( 4.3- 7.3) |

| | | | | |
|---|---|---|---|---|
| ANXI | APAT | 16.3 (15.4-17.1) | 26.0 (24.4-27.6) | 35.9 (32.9-39.0) |
| ANXI | SLEE | 12.9 (12.1-13.7) | 17.7 (16.3-19.1) | 21.1 (18.6-23.9) |
| ANXI | AGIT | 13.7 (12.9-14.5) | 22.3 (20.8-23.8) | 31.4 (28.5-34.4) |
| ANXI | APPE | 11.5 (10.7-12.2) | 16.4 (15.1-17.8) | 18.8 (16.4-21.4) |
| ANXI | DISI | 9.2 ( 8.5- 9.9) | 14.8 (13.5-16.1) | 20.1 (17.6-22.8) |
| ANXI | ABER | 8.1 ( 7.5- 8.8) | 18.4 (17.1-19.9) | 27.5 (24.7-30.4) |
| ANXI | PSYC | 7.5 ( 6.9- 8.1) | 16.8 (15.5-18.2) | 25.7 (22.9-28.5) |
| ANXI | DELU | 6.1 ( 5.6- 6.7) | 13.7 (12.5-15.0) | 20.3 (17.8-23.0) |
| ANXI | HALL | 2.9 ( 2.5- 3.3) | 8.2 ( 7.3- 9.3) | 15.2 (13.0-17.6) |
| ANXI | ELAT | 2.3 ( 2.0- 2.7) | 4.2 ( 3.5- 5.0) | 5.6 ( 4.3- 7.3) |
| APAT | SLEE | 14.2 (13.4-15.0) | 18.9 (17.5-20.3) | 24.6 (22.0-27.5) |
| APAT | AGIT | 14.7 (13.9-15.5) | 23.1 (21.6-24.6) | 34.1 (31.1-37.2) |
| APAT | APPE | 14.2 (13.4-15.0) | 18.2 (16.8-19.6) | 22.9 (20.3-25.7) |
| APAT | DISI | 10.9 (10.2-11.7) | 15.8 (14.5-17.1) | 23.6 (21.0-26.4) |
| APAT | ABER | 8.7 ( 8.0- 9.3) | 19.3 (17.9-20.8) | 31.1 (28.2-34.1) |
| APAT | PSYC | 7.6 ( 7.0- 8.2) | 16.9 (15.5-18.3) | 29.6 (26.7-32.5) |
| APAT | DELU | 6.0 ( 5.5- 6.6) | 13.5 (12.3-14.7) | 23.4 (20.8-26.2) |
| APAT | HALL | 3.0 ( 2.6- 3.4) | 8.2 ( 7.3- 9.3) | 16.9 (14.6-19.4) |
| APAT | ELAT | 2.7 ( 2.3- 3.1) | 4.1 ( 3.4- 4.9) | 6.0 ( 4.6- 7.6) |
| SLEE | AGIT | 11.3 (10.6-12.0) | 15.0 (13.7-16.3) | 23.5 (20.9-26.3) |
| SLEE | APPE | 10.7 (10.0-11.4) | 13.4 (12.2-14.6) | 17.8 (15.4-20.3) |
| SLEE | DISI | 7.8 ( 7.2- 8.4) | 10.5 ( 9.4-11.7) | 16.5 (14.2-19.0) |
| SLEE | ABER | 6.6 ( 6.1- 7.2) | 13.1 (11.9-14.3) | 22.3 (19.7-25.0) |
| SLEE | PSYC | 6.4 ( 5.9- 7.0) | 12.6 (11.4-13.8) | 20.6 (18.1-23.3) |
| SLEE | DELU | 4.8 ( 4.3- 5.3) | 9.7 ( 8.7-10.8) | 16.0 (13.8-18.5) |
| SLEE | HALL | 3.0 ( 2.6- 3.4) | 7.2 ( 6.3- 8.1) | 14.9 (12.7-17.3) |
| SLEE | ELAT | 2.1 ( 1.8- 2.4) | 2.6 ( 2.1- 3.2) | 4.9 ( 3.7- 6.5) |
| AGIT | APPE | 10.5 ( 9.8-11.3) | 14.8 (13.5-16.1) | 17.8 (15.4-20.3) |
| AGIT | DISI | 10.3 ( 9.6-11.0) | 16.0 (14.7-17.3) | 22.7 (20.1-25.5) |

| | | | | |
|---|---|---|---|---|
| AGIT | ABER | 7.6 ( 7.0- 8.2) | 16.4 (15.1-17.7) | 26.8 (24.0-29.7) |
| AGIT | PSYC | 7.9 ( 7.3- 8.6) | 17.0 (15.7-18.4) | 29.3 (26.4-32.2) |
| AGIT | DELU | 6.7 ( 6.2- 7.3) | 14.5 (13.2-15.8) | 23.9 (21.3-26.7) |
| AGIT | HALL | 2.6 ( 2.2- 2.9) | 7.6 ( 6.6- 8.6) | 17.0 (14.7-19.6) |
| AGIT | ELAT | 2.5 ( 2.1- 2.8) | 4.0 ( 3.3- 4.7) | 5.4 ( 4.1- 7.1) |
| APPE | DISI | 8.0 ( 7.4- 8.7) | 11.1 (10.0-12.3) | 13.6 (11.5-15.9) |
| APPE | ABER | 6.7 ( 6.1- 7.3) | 12.8 (11.6-14.0) | 19.1 (16.7-21.7) |
| APPE | PSYC | 5.6 ( 5.1- 6.2) | 10.6 ( 9.5-11.7) | 14.7 (12.5-17.1) |
| APPE | DELU | 4.5 ( 4.1- 5.0) | 8.7 ( 7.7- 9.8) | 11.3 ( 9.4-13.5) |
| APPE | HALL | 2.1 ( 1.8- 2.5) | 5.0 ( 4.2- 5.8) | 9.1 ( 7.4-11.1) |
| APPE | ELAT | 2.3 ( 2.0- 2.7) | 3.2 ( 2.6- 3.9) | 4.4 ( 3.2- 5.9) |
| DISI | ABER | 6.3 ( 5.7- 6.9) | 12.7 (11.5-13.9) | 21.7 (19.1-24.4) |
| DISI | PSYC | 5.1 ( 4.6- 5.7) | 11.1 (10.0-12.2) | 17.6 (15.2-20.1) |
| DISI | DELU | 4.5 ( 4.0- 5.0) | 9.4 ( 8.4-10.5) | 14.7 (12.5-17.1) |
| DISI | HALL | 1.5 ( 1.2- 1.8) | 5.1 ( 4.3- 5.9) | 10.6 ( 8.7-12.7) |
| DISI | ELAT | 2.6 ( 2.2- 2.9) | 4.2 ( 3.5- 5.0) | 6.1 ( 4.6- 7.7) |
| ABER | PSYC | 4.2 ( 3.7- 4.7) | 13.0 (11.8-14.3) | 24.1 (21.5-26.9) |
| ABER | DELU | 3.4 ( 3.0- 3.9) | 11.0 ( 9.9-12.1) | 18.7 (16.3-21.3) |
| ABER | HALL | 1.6 ( 1.3- 1.9) | 6.1 ( 5.3- 7.0) | 15.6 (13.4-18.0) |
| ABER | ELAT | 1.8 ( 1.5- 2.2) | 3.6 ( 3.0- 4.4) | 5.9 ( 4.5- 7.5) |
| PSYC | ELAT | 1.3 ( 1.1- 1.6) | 3.0 ( 2.5- 3.7) | 4.7 ( 3.5- 6.2) |
| DELU | HALL | 2.4 ( 2.1- 2.8) | 7.4 ( 6.5- 8.4) | 15.3 (13.1-17.7) |
| DELU | ELAT | 1.1 ( 0.9- 1.4) | 2.6 ( 2.1- 3.3) | 4.1 ( 2.9- 5.6) |
| HALL | ELAT | 0.5 ( 0.3- 0.6) | 1.3 ( 0.9- 1.7) | 2.8 ( 1.8- 4.0) |

Displayed are prevalence (%) and 95% confidence intervals.
NA, Not applicable; NPS, Neuropsychiatric symptom.
ABER, Aberrant motor behavior; AGIT, Agitation/aggression; ANXI, Anxiety; APAT, Apathy/indifference; APPE, Appetite/eating changes; DELU, Delusions; DEPR, Depression/dysphoria; DISI, Disinhibition; ELAT, Elation/euphoria; HALU, Hallucinations; IRRI, Irritability/lability; PSYC, Psychotic symptoms (hallucinations and/or delusions); SLEE, Nighttime behavioral disturbances.
Prevalence of pairs of neuropsychiatric symptoms for MMSE stratum >26 and ≤26 is reported in Table 2 (main text).

Supplemental Table 19. Prevalence of pairs of neuropsychiatric symptoms with the NPI severity score > 1 per MMSE stratum (NACC study only)

| NPS pair | | NACC | | | | |
|---|---|---|---|---|---|---|
| | | MMSE >26 | MMSE ≤26 | 19 < MMSE ≤ 26 | 9 < MMSE ≤ 19 | MMSE ≤ 9 |
| DEPR | IRRI | 2.2 (2.0-2.4) | 5.8 (5.4-6.3) | 5.2 (4.7-5.8) | 7.0 ( 6.2- 8.0) | 6.7 ( 5.2- 8.4) |
| DEPR | ANXI | 2.7 (2.4-2.9) | 7.3 (6.9-7.8) | 6.1 (5.6-6.7) | 9.2 ( 8.1-10.2) | 10.9 ( 9.0-13.0) |
| DEPR | APAT | 2.1 (1.9-2.4) | 7.0 (6.5-7.4) | 5.9 (5.3-6.4) | 8.6 ( 7.6- 9.6) | 10.2 ( 8.3-12.2) |
| DEPR | SLEE | 2.1 (1.8-2.3) | 4.8 (4.4-5.2) | 4.2 (3.8-4.7) | 5.6 ( 4.8- 6.5) | 6.2 ( 4.7- 7.9) |
| DEPR | AGIT | 1.6 (1.4-1.8) | 5.6 (5.2-6.0) | 4.5 (4.1-5.0) | 7.3 ( 6.3- 8.2) | 8.5 ( 6.8-10.5) |
| DEPR | APPE | 1.1 (1.0-1.3) | 3.8 (3.4-4.1) | 3.3 (2.9-3.8) | 4.8 ( 4.0- 5.6) | 3.9 ( 2.8- 5.3) |
| DEPR | DISI | 1.0 (0.8-1.1) | 3.0 (2.7-3.4) | 2.5 (2.1-2.9) | 4.1 ( 3.4- 4.8) | 4.1 ( 2.9- 5.6) |
| DEPR | ABER | 0.6 (0.5-0.8) | 3.1 (2.8-3.4) | 1.9 (1.6-2.3) | 4.7 ( 3.9- 5.5) | 7.0 ( 5.5- 8.8) |
| DEPR | PSYC | 0.4 (0.3-0.5) | 3.4 (3.1-3.7) | 2.2 (1.9-2.5) | 5.1 ( 4.4- 6.0) | 7.2 ( 5.6- 9.0) |
| DEPR | DELU | 0.4 (0.3-0.5) | 2.8 (2.5-3.1) | 1.9 (1.6-2.2) | 4.3 ( 3.6- 5.1) | 5.5 ( 4.2- 7.2) |
| DEPR | HALL | 0.1 (0.1-0.2) | 1.4 (1.2-1.6) | 0.8 (0.6-1.0) | 2.0 ( 1.5- 2.5) | 4.3 ( 3.1- 5.8) |
| DEPR | ELAT | 0.3 (0.2-0.4) | 0.7 (0.5-0.8) | 0.5 (0.4-0.7) | 0.9 ( 0.6- 1.3) | 0.9 ( 0.4- 1.7) |
| IRRI | ANXI | 2.1 (1.9-2.4) | 6.7 (6.3-7.2) | 5.7 (5.1-6.2) | 8.0 ( 7.1- 9.0) | 10.9 ( 9.0-13.0) |
| IRRI | APAT | 1.8 (1.6-2.0) | 6.9 (6.4-7.4) | 5.9 (5.3-6.4) | 8.1 ( 7.2- 9.2) | 10.5 ( 8.6-12.6) |
| IRRI | SLEE | 1.7 (1.5-1.9) | 5.1 (4.7-5.5) | 4.4 (3.9-4.9) | 5.7 ( 4.9- 6.6) | 8.2 ( 6.6-10.1) |
| IRRI | AGIT | 2.6 (2.3-2.8) | 8.1 (7.6-8.7) | 7.0 (6.4-7.6) | 9.6 ( 8.5-10.7) | 12.6 (10.6-14.9) |
| IRRI | APPE | 1.1 (1.0-1.3) | 4.3 (3.9-4.7) | 3.9 (3.5-4.4) | 5.0 ( 4.3- 5.9) | 4.8 ( 3.6- 6.4) |
| IRRI | DISI | 1.6 (1.4-1.8) | 5.1 (4.7-5.5) | 4.4 (3.9-4.9) | 5.7 ( 4.9- 6.6) | 8.7 ( 7.0-10.7) |
| IRRI | ABER | 0.9 (0.8-1.1) | 4.3 (3.9-4.7) | 2.9 (2.5-3.3) | 5.7 ( 4.9- 6.6) | 10.4 ( 8.5-12.5) |
| IRRI | PSYC | 0.6 (0.4-0.7) | 3.9 (3.5-4.2) | 2.6 (2.3-3.0) | 5.3 ( 4.5- 6.2) | 8.7 ( 7.0-10.7) |
| IRRI | DELU | 0.5 (0.4-0.6) | 3.4 (3.1-3.7) | 2.3 (2.0-2.7) | 4.7 ( 3.9- 5.5) | 7.2 ( 5.6- 9.0) |
| IRRI | HALL | 0.1 (0.1-0.2) | 1.4 (1.2-1.6) | 0.7 (0.5-0.9) | 1.9 ( 1.4- 2.4) | 5.1 ( 3.8- 6.7) |
| IRRI | ELAT | 0.5 (0.4-0.6) | 1.0 (0.8-1.2) | 0.9 (0.7-1.2) | 1.0 ( 0.7- 1.5) | 1.3 ( 0.7- 2.3) |
| ANXI | APAT | 1.5 (1.3-1.7) | 7.1 (6.7-7.6) | 5.4 (4.9-5.9) | 9.3 ( 8.2-10.4) | 14.1 (11.9-16.4) |

| | | | | | | |
|---|---|---|---|---|---|---|
| ANXI | SLEE | 1.7 (1.5-1.9) | 5.1 (4.7-5.5) | 4.1 (3.6-4.5) | 6.2 ( 5.4- 7.1) | 8.9 ( 7.2-10.9) |
| ANXI | AGIT | 1.5 (1.3-1.7) | 6.1 (5.6-6.5) | 4.6 (4.1-5.1) | 7.5 ( 6.5- 8.5) | 12.7 (10.7-15.0) |
| ANXI | APPE | 1.0 (0.9-1.2) | 4.0 (3.7-4.4) | 3.3 (2.9-3.7) | 4.9 ( 4.2- 5.8) | 6.6 ( 5.1- 8.3) |
| ANXI | DISI | 1.0 (0.9-1.2) | 3.9 (3.6-4.3) | 3.2 (2.8-3.6) | 4.5 ( 3.8- 5.3) | 7.6 ( 6.0- 9.4) |
| ANXI | ABER | 0.7 (0.6-0.9) | 4.4 (4.0-4.8) | 2.7 (2.4-3.1) | 6.2 ( 5.4- 7.1) | 11.4 ( 9.5-13.6) |
| ANXI | PSYC | 0.5 (0.4-0.6) | 4.0 (3.6-4.3) | 2.4 (2.0-2.7) | 5.8 ( 5.0- 6.7) | 10.4 ( 8.5-12.5) |
| ANXI | DELU | 0.4 (0.3-0.5) | 3.3 (2.9-3.6) | 1.9 (1.6-2.3) | 4.7 ( 4.0- 5.6) | 8.6 ( 6.9-10.6) |
| ANXI | HALL | 0.2 (0.1-0.2) | 1.7 (1.5-2.0) | 0.8 (0.6-1.1) | 2.5 ( 2.0- 3.2) | 5.9 ( 4.5- 7.5) |
| ANXI | ELAT | 0.3 (0.2-0.4) | 0.9 (0.7-1.0) | 0.7 (0.5-0.9) | 1.2 ( 0.8- 1.7) | 1.1 ( 0.6- 2.0) |
| APAT | SLEE | 1.8 (1.6-2.0) | 6.4 (6.0-6.9) | 5.1 (4.6-5.7) | 8.0 ( 7.0- 9.0) | 11.7 ( 9.8-13.9) |
| APAT | AGIT | 1.5 (1.3-1.7) | 6.8 (6.3-7.3) | 5.4 (4.9-5.9) | 8.2 ( 7.2- 9.2) | 13.2 (11.2-15.5) |
| APAT | APPE | 1.3 (1.1-1.5) | 5.9 (5.4-6.3) | 4.9 (4.4-5.4) | 7.3 ( 6.4- 8.3) | 8.7 ( 7.0-10.7) |
| APAT | DISI | 1.3 (1.1-1.5) | 4.9 (4.5-5.4) | 4.2 (3.8-4.7) | 5.2 ( 4.4- 6.1) | 9.5 ( 7.8-11.6) |
| APAT | ABER | 0.9 (0.8-1.1) | 5.5 (5.0-5.9) | 3.3 (2.9-3.7) | 7.8 ( 6.8- 8.8) | 14.6 (12.4-17.0) |
| APAT | PSYC | 0.4 (0.3-0.5) | 4.1 (3.8-4.5) | 2.5 (2.2-2.9) | 5.9 ( 5.1- 6.8) | 10.6 ( 8.7-12.7) |
| APAT | DELU | 0.3 (0.3-0.5) | 3.4 (3.1-3.8) | 2.1 (1.8-2.4) | 4.9 ( 4.2- 5.8) | 8.7 ( 7.0-10.7) |
| APAT | HALL | 0.1 (0.1-0.2) | 1.7 (1.5-2.0) | 0.9 (0.7-1.2) | 2.4 ( 1.9- 3.1) | 5.7 ( 4.4- 7.4) |
| APAT | ELAT | 0.5 (0.4-0.6) | 1.1 (0.9-1.3) | 1.0 (0.8-1.2) | 1.2 ( 0.9- 1.7) | 1.5 ( 0.9- 2.5) |
| SLEE | AGIT | 1.3 (1.2-1.5) | 4.6 (4.2-5.0) | 3.6 (3.2-4.0) | 5.5 ( 4.7- 6.4) | 9.7 ( 7.9-11.7) |
| SLEE | APPE | 1.4 (1.2-1.6) | 4.5 (4.1-4.9) | 3.8 (3.4-4.3) | 5.3 ( 4.5- 6.1) | 6.9 ( 5.4- 8.7) |
| SLEE | DISI | 1.0 (0.9-1.2) | 3.3 (3.0-3.7) | 2.7 (2.4-3.1) | 3.8 ( 3.2- 4.6) | 6.6 ( 5.1- 8.3) |
| SLEE | ABER | 0.8 (0.6-0.9) | 3.9 (3.5-4.2) | 2.4 (2.1-2.8) | 5.4 ( 4.6- 6.2) | 10.1 ( 8.2-12.1) |
| SLEE | PSYC | 0.4 (0.3-0.5) | 3.5 (3.1-3.8) | 2.1 (1.8-2.5) | 4.9 ( 4.2- 5.8) | 9.0 ( 7.3-11.0) |
| SLEE | DELU | 0.3 (0.2-0.4) | 2.6 (2.3-2.9) | 1.5 (1.3-1.8) | 3.8 ( 3.1- 4.5) | 7.2 ( 5.6- 9.0) |
| SLEE | HALL | 0.2 (0.1-0.3) | 1.9 (1.7-2.2) | 1.1 (0.9-1.3) | 2.6 ( 2.1- 3.3) | 6.0 ( 4.6- 7.6) |
| SLEE | ELAT | 0.3 (0.3-0.5) | 0.8 (0.6-1.0) | 0.7 (0.5-0.9) | 0.9 ( 0.6- 1.3) | 1.3 ( 0.7- 2.3) |
| AGIT | APPE | 0.9 (0.8-1.1) | 3.8 (3.5-4.2) | 3.3 (2.9-3.7) | 4.6 ( 3.9- 5.5) | 5.4 ( 4.1- 7.1) |
| AGIT | DISI | 1.4 (1.2-1.5) | 4.6 (4.2-5.0) | 3.8 (3.4-4.3) | 5.4 ( 4.6- 6.3) | 8.1 ( 6.5-10.0) |
| AGIT | ABER | 0.8 (0.6-0.9) | 4.2 (3.8-4.6) | 2.7 (2.3-3.0) | 5.7 ( 4.9- 6.6) | 11.1 ( 9.2-13.2) |

| | | | | | | |
|---|---|---|---|---|---|---|
| AGIT | PSYC | 0.5 (0.4-0.7) | 4.4 (4.1-4.8) | 2.7 (2.3-3.1) | 6.2 ( 5.3- 7.1) | 12.2 (10.2-14.4) |
| AGIT | DELU | 0.5 (0.4-0.6) | 3.8 (3.5-4.2) | 2.4 (2.1-2.8) | 5.4 ( 4.6- 6.2) | 9.9 ( 8.1-11.9) |
| AGIT | HALL | 0.1 (0.1-0.2) | 1.6 (1.3-1.8) | 0.6 (0.4-0.8) | 2.2 ( 1.7- 2.8) | 6.6 ( 5.1- 8.3) |
| AGIT | ELAT | 0.4 (0.3-0.5) | 0.9 (0.8-1.1) | 0.8 (0.7-1.1) | 1.0 ( 0.7- 1.4) | 1.5 ( 0.9- 2.5) |
| APPE | DISI | 0.9 (0.7-1.0) | 3.3 (3.0-3.7) | 2.9 (2.5-3.3) | 3.8 ( 3.2- 4.6) | 5.0 ( 3.7- 6.6) |
| APPE | ABER | 0.7 (0.6-0.8) | 3.4 (3.1-3.8) | 2.5 (2.2-2.9) | 4.5 ( 3.8- 5.4) | 6.9 ( 5.4- 8.7) |
| APPE | PSYC | 0.4 (0.3-0.5) | 2.4 (2.1-2.7) | 1.7 (1.4-2.0) | 3.3 ( 2.7- 4.0) | 5.1 ( 3.8- 6.7) |
| APPE | DELU | 0.3 (0.2-0.4) | 2.0 (1.7-2.3) | 1.4 (1.1-1.7) | 2.8 ( 2.2- 3.5) | 4.1 ( 2.9- 5.6) |
| APPE | HALL | 0.1 (0.1-0.2) | 1.0 (0.8-1.2) | 0.6 (0.4-0.8) | 1.4 ( 1.0- 1.9) | 2.8 ( 1.8- 4.0) |
| APPE | ELAT | 0.4 (0.3-0.5) | 1.0 (0.8-1.2) | 0.9 (0.7-1.1) | 1.1 ( 0.8- 1.6) | 1.2 ( 0.6- 2.1) |
| DISI | ABER | 0.8 (0.6-0.9) | 3.6 (3.3-4.0) | 2.4 (2.1-2.8) | 4.7 ( 4.0- 5.6) | 8.9 ( 7.2-10.9) |
| DISI | PSYC | 0.4 (0.3-0.5) | 2.6 (2.4-3.0) | 1.8 (1.5-2.1) | 3.7 ( 3.1- 4.4) | 5.9 ( 4.5- 7.5) |
| DISI | DELU | 0.3 (0.2-0.4) | 2.3 (2.0-2.6) | 1.6 (1.3-1.9) | 3.2 ( 2.6- 3.9) | 4.8 ( 3.6- 6.4) |
| DISI | HALL | 0.1 (0.0-0.1) | 1.1 (0.9-1.3) | 0.5 (0.3-0.7) | 1.6 ( 1.2- 2.1) | 4.0 ( 2.9- 5.4) |
| DISI | ELAT | 0.6 (0.4-0.7) | 1.1 (1.0-1.4) | 1.0 (0.8-1.3) | 1.4 ( 1.0- 1.9) | 1.5 ( 0.9- 2.5) |
| ABER | PSYC | 0.3 (0.2-0.4) | 2.9 (2.6-3.2) | 1.3 (1.1-1.6) | 4.9 ( 4.1- 5.7) | 8.8 ( 7.1-10.8) |
| ABER | DELU | 0.2 (0.2-0.3) | 2.5 (2.2-2.8) | 1.1 (0.9-1.3) | 4.4 ( 3.7- 5.2) | 7.3 ( 5.7- 9.1) |
| ABER | HALL | 0.1 (0.1-0.2) | 1.3 (1.1-1.6) | 0.5 (0.4-0.7) | 1.9 ( 1.4- 2.5) | 5.4 ( 4.1- 7.1) |
| ABER | ELAT | 0.3 (0.2-0.4) | 0.9 (0.7-1.1) | 0.7 (0.5-0.9) | 1.4 ( 1.0- 1.9) | 1.1 ( 0.6- 2.0) |
| PSYC | ELAT | 0.1 (0.1-0.2) | 0.5 (0.4-0.7) | 0.4 (0.2-0.5) | 0.8 ( 0.5- 1.2) | 1.0 ( 0.5- 1.9) |
| DELU | HALL | 0.1 (0.1-0.2) | 1.9 (1.7-2.2) | 0.9 (0.7-1.1) | 2.9 ( 2.3- 3.6) | 6.9 ( 5.4- 8.7) |
| DELU | ELAT | 0.1 (0.1-0.2) | 0.5 (0.4-0.6) | 0.3 (0.2-0.5) | 0.8 ( 0.5- 1.2) | 0.7 ( 0.3- 1.5) |
| HALL | ELAT | 0.0 (0.0-0.1) | 0.2 (0.1-0.3) | 0.1 (0.1-0.3) | 0.3 ( 0.1- 0.6) | 0.6 ( 0.2- 1.3) |

Displayed are prevalence (%) and 95% confidence intervals.
NA, Not applicable; NPS, Neuropsychiatric symptom.
ABER, Aberrant motor behavior; AGIT, Agitation/aggression; ANXI, Anxiety; APAT, Apathy/indifference; APPE, Appetite/eating changes; DELU, Delusions; DEPR, Depression/dysphoria; DISI, Disinhibition; ELAT, Elation/euphoria; HALU, Hallucinations; IRRI, Irritability/lability; PSYC, Psychotic symptoms (hallucinations and/or delusions); SLEE, Nighttime behavioral disturbances.

Supplemental Table 20. Prevalence of pairs of neuropsychiatric symptoms with the NPI product score ≥ 4 (ADAMS and ADNI studies only)

| NPS pair | | ADAMS survey | ADAMS | ADNI |
|---|---|---|---|---|
| DEPR | IRRI | 0.8 (0.2-2.6) | 1.1 (0.5-2.1) | 0.9 (0.4-1.8) |
| DEPR | ANXI | 0.7 (0.2-2.2) | 1.1 (0.5-2.1) | 0.4 (0.1-1.1) |
| DEPR | APAT | 0.9 (0.4-2.2) | 1.9 (1.1-3.1) | 0.9 (0.4-1.8) |
| DEPR | SLEE | NA | NA | 0.8 (0.3-1.6) |
| DEPR | AGIT | 0.7 (0.2-2.6) | 1.0 (0.4-2.0) | 0.1 (0.0-0.6) |
| DEPR | APPE | NA | NA | 0.8 (0.3-1.6) |
| DEPR | DISI | 0.6 (0.1-2.2) | 0.5 (0.1-1.3) | 0.2 (0.0-0.8) |
| DEPR | ABER | 0.4 (0.1-1.5) | 0.4 (0.1-1.1) | 0.1 (0.0-0.6) |
| DEPR | PSYC | 0.6 (0.2-1.6) | 0.9 (0.4-1.8) | 0.1 (0.0-0.6) |
| DEPR | DELU | 0.6 (0.2-1.6) | 0.8 (0.3-1.6) | 0.1 (0.0-0.6) |
| DEPR | HALL | 0.1 (0.0-0.5) | 0.4 (0.1-1.1) | 0.0 (0.0-0.4) |
| DEPR | ELAT | 0.0 (0.0- NE) | 0.0 (0.0-0.5) | 0.1 (0.0-0.6) |
| IRRI | ANXI | 0.3 (0.1-0.7) | 1.1 (0.5-2.1) | 0.3 (0.1-1.0) |
| IRRI | APAT | 0.8 (0.2-2.6) | 1.0 (0.4-2.0) | 1.1 (0.5-2.1) |
| IRRI | SLEE | NA | NA | 0.6 (0.2-1.3) |
| IRRI | AGIT | 1.5 (0.6-3.6) | 2.0 (1.1-3.2) | 0.7 (0.2-1.5) |
| IRRI | APPE | NA | NA | 0.9 (0.4-1.8) |
| IRRI | DISI | 0.7 (0.2-2.6) | 1.0 (0.4-2.0) | 0.7 (0.2-1.5) |
| IRRI | ABER | 0.6 (0.1-2.7) | 0.8 (0.3-1.6) | 0.2 (0.0-0.8) |
| IRRI | PSYC | 0.7 (0.2-2.6) | 1.4 (0.7-2.4) | 0.6 (0.2-1.3) |
| IRRI | DELU | 0.6 (0.2-2.6) | 1.1 (0.5-2.1) | 0.6 (0.2-1.3) |
| IRRI | HALL | 0.2 (0.1-0.5) | 0.8 (0.3-1.6) | 0.0 (0.0-0.4) |
| IRRI | ELAT | 0.0 (0.0- NE) | 0.0 (0.0-0.5) | 0.2 (0.0-0.8) |
| ANXI | APAT | 0.5 (0.2-0.9) | 1.3 (0.6-2.3) | 0.8 (0.3-1.6) |
| ANXI | SLEE | NA | NA | 0.3 (0.1-1.0) |
| ANXI | AGIT | 0.2 (0.1-0.4) | 0.9 (0.4-1.8) | 0.2 (0.0-0.8) |
| ANXI | APPE | NA | NA | 0.3 (0.1-1.0) |
| ANXI | DISI | 0.2 (0.1-0.5) | 0.6 (0.2-1.5) | 0.1 (0.0-0.6) |
| ANXI | ABER | 0.3 (0.2-0.5) | 1.0 (0.4-2.0) | 0.1 (0.0-0.6) |
| ANXI | PSYC | 0.3 (0.1-0.7) | 1.0 (0.4-2.0) | 0.1 (0.0-0.6) |
| ANXI | DELU | 0.2 (0.1-0.6) | 0.9 (0.4-1.8) | 0.1 (0.0-0.6) |
| ANXI | HALL | 0.2 (0.0-0.6) | 0.6 (0.2-1.5) | 0.0 (0.0-0.4) |
| ANXI | ELAT | 0.0 (0.0- NE) | 0.0 (0.0-0.5) | 0.0 (0.0-0.4) |

| | | | | |
|---|---|---|---|---|
| APAT | SLEE | NA | NA | 0.9 (0.4-1.8) |
| APAT | AGIT | 0.8 (0.3-2.5) | 1.1 (0.5-2.1) | 0.8 (0.3-1.6) |
| APAT | APPE | NA | NA | 1.8 (1.0-2.9) |
| APAT | DISI | 0.6 (0.1-2.7) | 0.6 (0.2-1.5) | 0.3 (0.1-1.0) |
| APAT | ABER | 0.9 (0.2-3.2) | 0.8 (0.3-1.6) | 0.7 (0.2-1.5) |
| APAT | PSYC | 1.0 (0.3-3.2) | 1.0 (0.4-2.0) | 0.3 (0.1-1.0) |
| APAT | DELU | 0.9 (0.3-3.2) | 0.9 (0.4-1.8) | 0.3 (0.1-1.0) |
| APAT | HALL | 0.1 (0.0-0.5) | 0.3 (0.0-0.9) | 0.1 (0.0-0.6) |
| APAT | ELAT | 0.0 (0.0- NE) | 0.0 (0.0-0.5) | 0.1 (0.0-0.6) |
| SLEE | AGIT | NA | NA | 0.0 (0.0-0.4) |
| SLEE | APPE | NA | NA | 0.8 (0.3-1.6) |
| SLEE | DISI | NA | NA | 0.2 (0.0-0.8) |
| SLEE | ABER | NA | NA | 0.4 (0.1-1.1) |
| SLEE | PSYC | NA | NA | 0.0 (0.0-0.4) |
| SLEE | DELU | NA | NA | 0.0 (0.0-0.4) |
| SLEE | HALL | NA | NA | 0.0 (0.0-0.4) |
| SLEE | ELAT | NA | NA | 0.1 (0.0-0.6) |
| AGIT | APPE | NA | NA | 0.2 (0.0-0.8) |
| AGIT | DISI | 0.6 (0.1-2.6) | 0.9 (0.4-1.8) | 0.2 (0.0-0.8) |
| AGIT | ABER | 0.7 (0.2-2.5) | 1.1 (0.5-2.1) | 0.1 (0.0-0.6) |
| AGIT | PSYC | 0.7 (0.2-2.6) | 1.1 (0.5-2.1) | 0.3 (0.1-1.0) |
| AGIT | DELU | 0.7 (0.2-2.6) | 1.1 (0.5-2.1) | 0.3 (0.1-1.0) |
| AGIT | HALL | 0.1 (0.0-0.3) | 0.5 (0.1-1.3) | 0.0 (0.0-0.4) |
| AGIT | ELAT | 0.0 (0.0- NE) | 0.0 (0.0-0.5) | 0.1 (0.0-0.6) |
| APPE | DISI | NA | NA | 0.4 (0.1-1.1) |
| APPE | ABER | NA | NA | 0.6 (0.2-1.3) |
| APPE | PSYC | NA | NA | 0.3 (0.1-1.0) |
| APPE | DELU | NA | NA | 0.3 (0.1-1.0) |
| APPE | HALL | NA | NA | 0.1 (0.0-0.6) |
| APPE | ELAT | NA | NA | 0.1 (0.0-0.6) |
| DISI | ABER | 0.5 (0.1-2.8) | 0.6 (0.2-1.5) | 0.2 (0.0-0.8) |
| DISI | PSYC | 0.6 (0.1-2.7) | 0.8 (0.3-1.6) | 0.3 (0.1-1.0) |
| DISI | DELU | 0.6 (0.1-2.7) | 0.6 (0.2-1.5) | 0.3 (0.1-1.0) |
| DISI | HALL | 0.1 (0.0-0.5) | 0.4 (0.1-1.1) | 0.0 (0.0-0.4) |
| DISI | ELAT | 0.0 (0.0- NE) | 0.0 (0.0-0.5) | 0.3 (0.1-1.0) |

| | | | | |
|---|---|---|---|---|
| ABER | PSYC | 1.0 (0.3-2.9) | 1.1 (0.5-2.1) | 0.2 (0.0-0.8) |
| ABER | DELU | 0.9 (0.3-2.9) | 1.0 (0.4-2.0) | 0.2 (0.0-0.8) |
| ABER | HALL | 0.1 (0.0-0.2) | 0.4 (0.1-1.1) | 0.1 (0.0-0.6) |
| ABER | ELAT | 0.0 (0.0- NE) | 0.0 (0.0-0.5) | 0.2 (0.0-0.8) |
| PSYC | ELAT | 0.0 (0.0- NE) | 0.0 (0.0-0.5) | 0.2 (0.0-0.8) |
| DELU | HALL | 0.1 (0.0-0.4) | 0.8 (0.3-1.6) | 0.1 (0.0-0.6) |
| DELU | ELAT | 0.0 (0.0- NE) | 0.0 (0.0-0.5) | 0.2 (0.0-0.8) |
| HALL | ELAT | 0.0 (0.0- NE) | 0.0 (0.0-0.5) | 0.0 (0.0-0.4) |

Displayed are prevalence (%) and 95% confidence intervals. Survey estimates for ADAMS are based on survey population weights, confidence intervals are based on survey clusters and survey strata.

NA, Not applicable; NE, Not estimable; NPS, Neuropsychiatric symptom.

ABER, Aberrant motor behavior; AGIT, Agitation/aggression; ANXI, Anxiety; APAT, Apathy/indifference; APPE, Appetite/eating changes; DELU, Delusions; DEPR, Depression/dysphoria; DISI, Disinhibition; ELAT, Elation/euphoria; HALU, Hallucinations; IRRI, Irritability/lability; PSYC, Psychotic symptoms (hallucinations and/or delusions); SLEE, Nighttime behavioral disturbances.

Supplemental Table 21. Prevalence of pairs of neuropsychiatric symptoms with the NPI product score ≥ 4 per MMSE stratum (ADAMS and ADNI studies only)

| NPS pair | | ADAMS survey | | ADAMS | | ADNI | |
|---|---|---|---|---|---|---|---|
| | | MMSE > 26 | MMSE ≤ 26 | MMSE > 26 | MMSE ≤ 26 | MMSE > 26 | MMSE ≤ 26 |
| DEPR | IRRI | 0.0 (0.0- NE) | 1.9 (0.6-6.5) | 0.0 (0.0-1.4) | 1.7 (0.8-3.1) | 0.2 (0.0-0.9) | 2.4 (1.0-4.9) |
| DEPR | ANXI | 0.6 (0.1-4.6) | 0.8 (0.3-1.9) | 0.4 (0.0-2.2) | 1.5 (0.6-2.9) | 0.2 (0.0-0.9) | 1.0 (0.2-3.0) |
| DEPR | APAT | 0.0 (0.0- NE) | 2.3 (0.9-5.6) | 0.0 (0.0-1.4) | 2.8 (1.6-4.5) | 0.3 (0.0-1.2) | 2.1 (0.8-4.5) |
| DEPR | SLEE | NA | NA | NA | NA | 0.3 (0.0-1.2) | 1.7 (0.6-4.0) |
| DEPR | AGIT | 0.0 (0.0-0.3) | 1.8 (0.5-6.5) | 0.4 (0.0-2.2) | 1.3 (0.5-2.6) | 0.0 (0.0-0.6) | 0.3 (0.0-1.9) |
| DEPR | APPE | NA | NA | NA | NA | 0.5 (0.1-1.5) | 1.4 (0.4-3.5) |
| DEPR | DISI | 0.0 (0.0- NE) | 1.4 (0.3-5.4) | 0.0 (0.0-1.4) | 0.7 (0.2-1.9) | 0.2 (0.0-0.9) | 0.3 (0.0-1.9) |
| DEPR | ABER | 0.0 (0.0- NE) | 1.1 (0.3-3.7) | 0.0 (0.0-1.4) | 0.6 (0.1-1.6) | 0.2 (0.0-0.9) | 0.0 (0.0-1.3) |
| DEPR | PSYC | 0.0 (0.0- NE) | 1.5 (0.6-4.1) | 0.0 (0.0-1.4) | 1.3 (0.5-2.6) | 0.0 (0.0-0.6) | 0.3 (0.0-1.9) |
| DEPR | DELU | 0.0 (0.0- NE) | 1.4 (0.5-3.9) | 0.0 (0.0-1.4) | 1.1 (0.4-2.4) | 0.0 (0.0-0.6) | 0.3 (0.0-1.9) |
| DEPR | HALL | 0.0 (0.0- NE) | 0.3 (0.1-1.2) | 0.0 (0.0-1.4) | 0.6 (0.1-1.6) | 0.0 (0.0-0.6) | 0.0 (0.0-1.3) |
| DEPR | ELAT | 0.0 (0.0- NE) | 0.0 (0.0- NE) | 0.0 (0.0-1.4) | 0.0 (0.0-0.7) | 0.0 (0.0-0.6) | 0.3 (0.0-1.9) |
| IRRI | ANXI | 0.0 (0.0- NE) | 0.7 (0.3-1.8) | 0.0 (0.0-1.4) | 1.7 (0.8-3.1) | 0.2 (0.0-0.9) | 0.7 (0.1-2.5) |
| IRRI | APAT | 0.0 (0.0- NE) | 1.9 (0.5-6.4) | 0.0 (0.0-1.4) | 1.5 (0.6-2.9) | 0.8 (0.3-1.9) | 1.7 (0.6-4.0) |
| IRRI | SLEE | NA | NA | NA | NA | 0.3 (0.0-1.2) | 1.0 (0.2-3.0) |
| IRRI | AGIT | 0.0 (0.0- NE) | 3.7 (1.5-8.7) | 0.0 (0.0-1.4) | 2.9 (1.7-4.7) | 0.7 (0.2-1.7) | 0.7 (0.1-2.5) |
| IRRI | APPE | NA | NA | NA | NA | 0.3 (0.0-1.2) | 2.1 (0.8-4.5) |
| IRRI | DISI | 0.0 (0.0- NE) | 1.7 (0.4-6.3) | 0.0 (0.0-1.4) | 1.5 (0.6-2.9) | 0.5 (0.1-1.5) | 1.0 (0.2-3.0) |
| IRRI | ABER | 0.0 (0.0- NE) | 1.4 (0.3-6.5) | 0.0 (0.0-1.4) | 1.1 (0.4-2.4) | 0.0 (0.0-0.6) | 0.7 (0.1-2.5) |
| IRRI | PSYC | 0.0 (0.0- NE) | 1.8 (0.5-6.2) | 0.0 (0.0-1.4) | 2.0 (1.0-3.6) | 0.3 (0.0-1.2) | 1.0 (0.2-3.0) |
| IRRI | DELU | 0.0 (0.0- NE) | 1.6 (0.4-6.3) | 0.0 (0.0-1.4) | 1.7 (0.8-3.1) | 0.3 (0.0-1.2) | 1.0 (0.2-3.0) |
| IRRI | HALL | 0.0 (0.0- NE) | 0.4 (0.1-1.2) | 0.0 (0.0-1.4) | 1.1 (0.4-2.4) | 0.0 (0.0-0.6) | 0.0 (0.0-1.3) |
| IRRI | ELAT | 0.0 (0.0- NE) | 0.0 (0.0- NE) | 0.0 (0.0-1.4) | 0.0 (0.0-0.7) | 0.0 (0.0-0.6) | 0.7 (0.1-2.5) |
| ANXI | APAT | 0.0 (0.0- NE) | 1.2 (0.6-2.4) | 0.0 (0.0-1.4) | 1.8 (0.9-3.4) | 0.2 (0.0-0.9) | 2.1 (0.8-4.5) |

| | | | | | | | |
|---|---|---|---|---|---|---|---|
| ANXI | SLEE | NA | NA | NA | NA | 0.3 (0.0-1.2) | 0.3 (0.0-1.9) |
| ANXI | AGIT | 0.0 (0.0- NE) | 0.6 (0.3-1.1) | 0.0 (0.0-1.4) | 1.3 (0.5-2.6) | 0.0 (0.0-0.6) | 0.7 (0.1-2.5) |
| ANXI | APPE | NA | NA | NA | NA | 0.2 (0.0-0.9) | 0.7 (0.1-2.5) |
| ANXI | DISI | 0.0 (0.0- NE) | 0.4 (0.1-1.3) | 0.0 (0.0-1.4) | 0.9 (0.3-2.1) | 0.2 (0.0-0.9) | 0.0 (0.0-1.3) |
| ANXI | ABER | 0.0 (0.0- NE) | 0.7 (0.4-1.4) | 0.0 (0.0-1.4) | 1.5 (0.6-2.9) | 0.0 (0.0-0.6) | 0.3 (0.0-1.9) |
| ANXI | PSYC | 0.0 (0.0- NE) | 0.7 (0.3-1.9) | 0.0 (0.0-1.4) | 1.5 (0.6-2.9) | 0.0 (0.0-0.6) | 0.3 (0.0-1.9) |
| ANXI | DELU | 0.0 (0.0- NE) | 0.6 (0.2-1.6) | 0.0 (0.0-1.4) | 1.3 (0.5-2.6) | 0.0 (0.0-0.6) | 0.3 (0.0-1.9) |
| ANXI | HALL | 0.0 (0.0- NE) | 0.4 (0.1-1.4) | 0.0 (0.0-1.4) | 0.9 (0.3-2.1) | 0.0 (0.0-0.6) | 0.0 (0.0-1.3) |
| ANXI | ELAT | 0.0 (0.0- NE) | 0.0 (0.0- NE) | 0.0 (0.0-1.4) | 0.0 (0.0-0.7) | 0.0 (0.0-0.6) | 0.0 (0.0-1.3) |
| APAT | SLEE | NA | NA | NA | NA | 0.5 (0.1-1.5) | 1.7 (0.6-4.0) |
| APAT | AGIT | 0.0 (0.0- NE) | 2.1 (0.7-6.1) | 0.0 (0.0-1.4) | 1.7 (0.8-3.1) | 0.5 (0.1-1.5) | 1.4 (0.4-3.5) |
| APAT | APPE | NA | NA | NA | NA | 0.2 (0.0-0.9) | 5.2 (2.9-8.4) |
| APAT | DISI | 0.0 (0.0- NE) | 1.6 (0.4-6.4) | 0.0 (0.0-1.4) | 0.9 (0.3-2.1) | 0.5 (0.1-1.5) | 0.0 (0.0-1.3) |
| APAT | ABER | 0.0 (0.0- NE) | 2.2 (0.6-7.6) | 0.0 (0.0-1.4) | 1.1 (0.4-2.4) | 0.2 (0.0-0.9) | 1.7 (0.6-4.0) |
| APAT | PSYC | 0.0 (0.0- NE) | 2.5 (0.8-7.7) | 0.0 (0.0-1.4) | 1.5 (0.6-2.9) | 0.0 (0.0-0.6) | 1.0 (0.2-3.0) |
| APAT | DELU | 0.0 (0.0- NE) | 2.3 (0.7-7.7) | 0.0 (0.0-1.4) | 1.3 (0.5-2.6) | 0.0 (0.0-0.6) | 1.0 (0.2-3.0) |
| APAT | HALL | 0.0 (0.0- NE) | 0.2 (0.0-1.2) | 0.0 (0.0-1.4) | 0.4 (0.0-1.3) | 0.0 (0.0-0.6) | 0.3 (0.0-1.9) |
| APAT | ELAT | 0.0 (0.0- NE) | 0.0 (0.0- NE) | 0.0 (0.0-1.4) | 0.0 (0.0-0.7) | 0.2 (0.0-0.9) | 0.0 (0.0-1.3) |
| SLEE | AGIT | NA | NA | NA | NA | 0.0 (0.0-0.6) | 0.0 (0.0-1.3) |
| SLEE | APPE | NA | NA | NA | NA | 0.3 (0.0-1.2) | 1.7 (0.6-4.0) |
| SLEE | DISI | NA | NA | NA | NA | 0.3 (0.0-1.2) | 0.0 (0.0-1.3) |
| SLEE | ABER | NA | NA | NA | NA | 0.2 (0.0-0.9) | 1.0 (0.2-3.0) |
| SLEE | PSYC | NA | NA | NA | NA | 0.0 (0.0-0.6) | 0.0 (0.0-1.3) |
| SLEE | DELU | NA | NA | NA | NA | 0.0 (0.0-0.6) | 0.0 (0.0-1.3) |
| SLEE | HALL | NA | NA | NA | NA | 0.0 (0.0-0.6) | 0.0 (0.0-1.3) |
| SLEE | ELAT | NA | NA | NA | NA | 0.0 (0.0-0.6) | 0.3 (0.0-1.9) |
| AGIT | APPE | NA | NA | NA | NA | 0.0 (0.0-0.6) | 0.7 (0.1-2.5) |
| AGIT | DISI | 0.0 (0.0- NE) | 1.5 (0.3-6.3) | 0.0 (0.0-1.4) | 1.3 (0.5-2.6) | 0.3 (0.0-1.2) | 0.0 (0.0-1.3) |
| AGIT | ABER | 0.0 (0.0- NE) | 1.7 (0.5-6.0) | 0.0 (0.0-1.4) | 1.7 (0.8-3.1) | 0.0 (0.0-0.6) | 0.3 (0.0-1.9) |

| | | | | | | | |
|---|---|---|---|---|---|---|---|
| AGIT | PSYC | 0.0 (0.0- NE) | 1.6 (0.4-6.3) | 0.0 (0.0-1.4) | 1.7 (0.8-3.1) | 0.0 (0.0-0.6) | 1.0 (0.2-3.0) |
| AGIT | DELU | 0.0 (0.0- NE) | 1.6 (0.4-6.3) | 0.0 (0.0-1.4) | 1.7 (0.8-3.1) | 0.0 (0.0-0.6) | 1.0 (0.2-3.0) |
| AGIT | HALL | 0.0 (0.0- NE) | 0.2 (0.1-0.8) | 0.0 (0.0-1.4) | 0.7 (0.2-1.9) | 0.0 (0.0-0.6) | 0.0 (0.0-1.3) |
| AGIT | ELAT | 0.0 (0.0- NE) | 0.0 (0.0- NE) | 0.0 (0.0-1.4) | 0.0 (0.0-0.7) | 0.0 (0.0-0.6) | 0.3 (0.0-1.9) |
| APPE | DISI | NA | NA | NA | NA | 0.5 (0.1-1.5) | 0.3 (0.0-1.9) |
| APPE | ABER | NA | NA | NA | NA | 0.3 (0.0-1.2) | 1.0 (0.2-3.0) |
| APPE | PSYC | NA | NA | NA | NA | 0.2 (0.0-0.9) | 0.7 (0.1-2.5) |
| APPE | DELU | NA | NA | NA | NA | 0.2 (0.0-0.9) | 0.7 (0.1-2.5) |
| APPE | HALL | NA | NA | NA | NA | 0.0 (0.0-0.6) | 0.3 (0.0-1.9) |
| APPE | ELAT | NA | NA | NA | NA | 0.2 (0.0-0.9) | 0.0 (0.0-1.3) |
| DISI | ABER | 0.0 (0.0- NE) | 1.3 (0.2-6.7) | 0.0 (0.0-1.4) | 0.9 (0.3-2.1) | 0.2 (0.0-0.9) | 0.3 (0.0-1.9) |
| DISI | PSYC | 0.0 (0.0- NE) | 1.6 (0.4-6.4) | 0.0 (0.0-1.4) | 1.1 (0.4-2.4) | 0.2 (0.0-0.9) | 0.7 (0.1-2.5) |
| DISI | DELU | 0.0 (0.0- NE) | 1.4 (0.3-6.5) | 0.0 (0.0-1.4) | 0.9 (0.3-2.1) | 0.2 (0.0-0.9) | 0.7 (0.1-2.5) |
| DISI | HALL | 0.0 (0.0- NE) | 0.3 (0.1-1.2) | 0.0 (0.0-1.4) | 0.6 (0.1-1.6) | 0.0 (0.0-0.6) | 0.0 (0.0-1.3) |
| DISI | ELAT | 0.0 (0.0- NE) | 0.0 (0.0- NE) | 0.0 (0.0-1.4) | 0.0 (0.0-0.7) | 0.3 (0.0-1.2) | 0.3 (0.0-1.9) |
| ABER | PSYC | 0.0 (0.0- NE) | 2.4 (0.8-6.9) | 0.0 (0.0-1.4) | 1.7 (0.8-3.1) | 0.0 (0.0-0.6) | 0.7 (0.1-2.5) |
| ABER | DELU | 0.0 (0.0- NE) | 2.4 (0.8-6.9) | 0.0 (0.0-1.4) | 1.5 (0.6-2.9) | 0.0 (0.0-0.6) | 0.7 (0.1-2.5) |
| ABER | HALL | 0.0 (0.0- NE) | 0.2 (0.0-0.6) | 0.0 (0.0-1.4) | 0.6 (0.1-1.6) | 0.0 (0.0-0.6) | 0.3 (0.0-1.9) |
| ABER | ELAT | 0.0 (0.0- NE) | 0.0 (0.0- NE) | 0.0 (0.0-1.4) | 0.0 (0.0-0.7) | 0.2 (0.0-0.9) | 0.3 (0.0-1.9) |
| PSYC | ELAT | 0.0 (0.0- NE) | 0.0 (0.0- NE) | 0.0 (0.0-1.4) | 0.0 (0.0-0.7) | 0.0 (0.0-0.6) | 0.7 (0.1-2.5) |
| DELU | HALL | 0.0 (0.0- NE) | 0.4 (0.1-1.1) | 0.0 (0.0-1.4) | 1.1 (0.4-2.4) | 0.0 (0.0-0.6) | 0.3 (0.0-1.9) |
| DELU | ELAT | 0.0 (0.0- NE) | 0.0 (0.0- NE) | 0.0 (0.0-1.4) | 0.0 (0.0-0.7) | 0.0 (0.0-0.6) | 0.7 (0.1-2.5) |
| HALL | ELAT | 0.0 (0.0- NE) | 0.0 (0.0- NE) | 0.0 (0.0-1.4) | 0.0 (0.0-0.7) | 0.0 (0.0-0.6) | 0.0 (0.0-1.3) |

Displayed are prevalence (%) and 95% confidence intervals. Survey estimates for ADAMS are based on survey population weights, confidence intervals are based on survey clusters and survey strata.

NA, Not applicable; NE, Not estimable; NPS, Neuropsychiatric symptom.

ABER, Aberrant motor behavior; AGIT, Agitation/aggression; ANXI, Anxiety; APAT, Apathy/indifference; APPE, Appetite/eating changes; DELU, Delusions; DEPR, Depression/dysphoria; DISI, Disinhibition; ELAT, Elation/euphoria; HALU, Hallucinations; IRRI, Irritability/lability; PSYC, Psychotic symptoms (hallucinations and/or delusions); SLEE, Nighttime behavioral disturbances.

### Supplement section 6. Conditional prevalence of pairs of NPI domains

Supplemental Table 22. Conditional prevalence S1|S2 of neuropsychiatric symptoms with the NPI severity score > 0

| S1 | S2 | ADAMS survey | ADAMS | ADNI | NACC |
|---|---|---|---|---|---|
| DEPR | IRRI | 56.4 (NE-NE) | 48.9 (38.2- 60.0) | 47.3 (40.1-53.9) | 52.0 (50.8-53.1) |
| IRRI | DEPR | 28.7 (NE-NE) | 27.6 (20.9-34.3) | 43.6 (37.2-50.5) | 48.1 (46.9-49.2) |
| DEPR | ANXI | 60.8 (NE-NE) | 60.9 (50.7- 71.1) | 53.8 (44.6-62.2) | 59.6 (58.3-60.7) |
| ANXI | DEPR | 27.5 (NE-NE) | 34.0 (27.2-41.4) | 32.6 (26.3-39.1) | 51.2 (50.1-52.3) |
| DEPR | APAT | 68.4 (NE-NE) | 63.5 (53.2- 73.8) | 52.5 (44.6-60.3) | 55.7 (54.4-56.9) |
| APAT | DEPR | 30.0 (NE-NE) | 34.6 (27.7-42.2) | 38.5 (32.4-45.0) | 46.7 (45.6-47.8) |
| DEPR | SLEE | NA | NA | 45.4 (37.8-53.0) | 50.4 (49.1-51.5) |
| SLEE | DEPR | NA | NA | 33.9 (27.6-40.4) | 37.4 (36.3-38.4) |
| DEPR | AGIT | 65.3 (NE-NE) | 48.0 (37.8- 57.8) | 47.0 (38.1-55.3) | 53.3 (51.9-54.5) |
| AGIT | DEPR | 28.8 (NE-NE) | 30.8 (24.0-38.3) | 28.4 (23.0-34.8) | 37.7 (36.6-38.7) |
| DEPR | APPE | NA | NA | 50.5 (40.7-60.4) | 51.0 (49.6-52.5) |
| APPE | DEPR | NA | NA | 24.3 (18.9-30.2) | 29.6 (28.6-30.6) |
| DEPR | DISI | 54.2 (NE-NE) | 52.4 (36.4- 67.7) | 52.3 (42.2-62.9) | 50.6 (48.9-52.3) |
| DISI | DEPR | 11.9 (NE-NE) | 14.1 ( 8.8-20.1) | 21.1 (16.0-26.8) | 22.7 (21.8-23.5) |
| DEPR | ABER | 50.4 (NE-NE) | 32.6 (20.4- 45.5) | 60.0 (46.0-72.9) | 52.6 (50.8-54.5) |
| ABER | DEPR | 7.5 (NE-NE) | 9.6 ( 5.4-14.4) | 13.8 ( 9.1-18.3) | 20.2 (19.3-21.2) |
| DEPR | PSYC | 52.0 (NE-NE) | 40.0 (29.9- 51.1) | 59.0 (42.9-74.1) | 56.7 (54.9-58.5) |
| PSYC | DEPR | 13.0 (NE-NE) | 21.8 (15.7-28.3) | 10.6 ( 6.7-14.6) | 20.3 (19.4-21.2) |
| DEPR | DELU | 53.8 (NE-NE) | 38.3 (25.9- 50.9) | 67.7 (50.0-83.3) | 58.3 (56.3-60.2) |
| DELU | DEPR | 11.1 (NE-NE) | 14.7 ( 9.4-20.6) | 9.6 ( 6.0-13.5) | 16.6 (15.8-17.4) |
| DEPR | HALL | 50.1 (NE-NE) | 44.9 (30.8- 58.2) | 46.2 (16.7-75.0) | 56.0 (53.2-59.1) |
| HALL | DEPR | 6.7 (NE-NE) | 14.1 ( 8.8-19.9) | 2.8 ( 0.8- 5.2) | 8.6 ( 7.9- 9.1) |
| DEPR | ELAT | 38.6 (NE-NE) | 12.5 ( 0.0- 40.0) | 50.0 (28.6-72.7) | 50.7 (47.4-53.7) |
| ELAT | DEPR | 2.6 (NE-NE) | 0.6 ( 0.0- 2.1) | 4.6 ( 2.0- 7.5) | 6.0 ( 5.4- 6.5) |
| IRRI | ANXI | 31.0 (NE-NE) | 40.2 (30.3- 50.6) | 50.8 (42.5-59.5) | 51.8 (50.6-52.9) |
| ANXI | IRRI | 27.5 (NE-NE) | 39.8 (29.8-49.5) | 33.3 (27.1-40.4) | 48.1 (47.0-49.3) |
| IRRI | APAT | 32.2 (NE-NE) | 32.9 (23.0- 43.5) | 48.8 (41.0-57.0) | 51.2 (50.1-52.4) |
| APAT | IRRI | 27.8 (NE-NE) | 31.8 (21.8-42.3) | 38.8 (32.0-45.6) | 46.5 (45.3-47.5) |
| IRRI | SLEE | NA | NA | 36.2 (29.6-43.6) | 47.2 (45.9-48.6) |

| | | | | | |
|---|---|---|---|---|---|
| SLEE | IRRI | NA | NA | 29.4 (23.3-35.7) | 38.0 (36.9-39.1) |
| IRRI | AGIT | 64.7 (NE-NE) | 58.0 (48.1- 67.0) | 64.4 (56.4-72.7) | 69.6 (68.4-70.9) |
| AGIT | IRRI | 56.1 (NE-NE) | 65.9 (55.8-76.0) | 42.3 (35.4-49.3) | 53.3 (52.1-54.4) |
| IRRI | APPE | NA | NA | 40.0 (31.4-50.0) | 48.5 (47.0-50.0) |
| APPE | IRRI | NA | NA | 20.9 (15.5-26.7) | 30.5 (29.3-31.6) |
| IRRI | DISI | 56.9 (NE-NE) | 59.5 (43.9- 75.0) | 59.1 (48.8-68.8) | 65.1 (63.5-66.6) |
| DISI | IRRI | 24.6 (NE-NE) | 28.4 (19.7-37.9) | 25.9 (19.8-31.5) | 31.6 (30.6-32.6) |
| IRRI | ABER | 40.8 (NE-NE) | 41.3 (26.8- 54.3) | 42.0 (28.3-55.3) | 57.9 (56.1-59.6) |
| ABER | IRRI | 11.9 (NE-NE) | 21.6 (13.3-30.7) | 10.4 ( 6.1-14.4) | 24.1 (23.1-25.1) |
| IRRI | PSYC | 38.5 (NE-NE) | 38.8 (27.8- 49.4) | 53.8 (37.8-69.0) | 57.3 (55.5-59.1) |
| PSYC | IRRI | 18.9 (NE-NE) | 37.5 (27.2-47.8) | 10.4 ( 6.5-14.9) | 22.2 (21.3-23.2) |
| IRRI | DELU | 38.3 (NE-NE) | 36.7 (24.1- 49.1) | 58.1 (40.7-75.9) | 60.8 (58.8-62.8) |
| DELU | IRRI | 15.5 (NE-NE) | 25.0 (16.1-34.6) | 9.0 ( 5.3-13.2) | 18.7 (17.8-19.7) |
| IRRI | HALL | 40.6 (NE-NE) | 44.9 (31.2- 58.5) | 38.5 (12.5-69.2) | 52.8 (49.9-55.7) |
| HALL | IRRI | 10.7 (NE-NE) | 25.0 (16.7-34.5) | 2.5 ( 0.9- 4.7) | 8.7 ( 8.1- 9.4) |
| IRRI | ELAT | 4.5 (NE-NE) | 25.0 ( 0.0- 57.3) | 50.0 (28.6-72.2) | 61.9 (58.9-64.9) |
| ELAT | IRRI | 0.6 (NE-NE) | 2.3 ( 0.0- 5.4) | 5.0 ( 2.2- 8.0) | 7.9 ( 7.3- 8.5) |
| ANXI | APAT | 33.6 (NE-NE) | 38.8 (28.9- 50.0) | 35.6 (28.3-42.9) | 48.2 (47.0-49.4) |
| APAT | ANXI | 32.7 (NE-NE) | 37.9 (28.1-48.2) | 43.2 (34.9-51.1) | 47.0 (45.8-48.2) |
| ANXI | SLEE | NA | NA | 30.7 (23.5-38.2) | 45.5 (44.2-46.7) |
| SLEE | ANXI | NA | NA | 37.9 (29.5-46.0) | 39.3 (38.2-40.5) |
| ANXI | AGIT | 27.5 (NE-NE) | 35.0 (25.8- 44.3) | 41.7 (33.8-50.0) | 49.5 (48.2-50.7) |
| AGIT | ANXI | 26.9 (NE-NE) | 40.2 (30.0-50.6) | 41.7 (33.6-50.7) | 40.7 (39.5-41.8) |
| ANXI | APPE | NA | NA | 30.5 (21.9-40.5) | 46.2 (44.8-47.6) |
| APPE | ANXI | NA | NA | 24.2 (16.9-31.7) | 31.2 (30.1-32.3) |
| ANXI | DISI | 27.6 (NE-NE) | 50.0 (35.3- 65.6) | 39.8 (30.0-50.0) | 52.1 (50.5-53.8) |
| DISI | ANXI | 13.4 (NE-NE) | 24.1 (15.4-32.5) | 26.5 (19.4-34.1) | 27.2 (26.3-28.3) |
| ANXI | ABER | 27.9 (NE-NE) | 41.3 (26.5- 54.8) | 46.0 (31.6-58.7) | 58.4 (56.6-60.2) |
| ABER | ANXI | 9.2 (NE-NE) | 21.8 (13.5-30.3) | 17.4 (11.0-23.7) | 26.1 (25.1-27.2) |
| ANXI | PSYC | 21.1 (NE-NE) | 31.8 (21.6- 41.2) | 48.7 (32.4-63.6) | 55.5 (53.5-57.3) |
| PSYC | ANXI | 11.6 (NE-NE) | 31.0 (21.8-41.6) | 14.4 ( 8.8-20.4) | 23.2 (22.1-24.2) |
| ANXI | DELU | 18.5 (NE-NE) | 31.7 (20.3- 43.4) | 45.2 (27.6-62.2) | 56.4 (54.3-58.5) |
| DELU | ANXI | 8.4 (NE-NE) | 21.8 (13.4-30.8) | 10.6 ( 5.7-15.8) | 18.7 (17.7-19.7) |
| ANXI | HALL | 28.7 (NE-NE) | 38.8 (25.0- 51.9) | 53.8 (25.0-81.8) | 59.5 (56.5-62.4) |
| HALL | ANXI | 8.5 (NE-NE) | 21.8 (13.7-31.3) | 5.3 ( 1.7- 9.4) | 10.6 ( 9.8-11.3) |

| | | | | | |
|---|---|---|---|---|---|
| ANXI | ELAT | 38.6 (NE-NE) | 12.5 ( 0.0- 44.4) | 30.0 (10.5-50.0) | 54.7 (51.7-58.2) |
| ELAT | ANXI | 5.7 (NE-NE) | 1.1 ( 0.0- 4.2) | 4.5 ( 1.4- 8.2) | 7.5 ( 6.9- 8.2) |
| APAT | SLEE | NA | NA | 28.2 (21.2-35.4) | 46.6 (45.4-47.9) |
| SLEE | APAT | NA | NA | 28.7 (21.8-35.8) | 41.3 (40.1-42.5) |
| APAT | AGIT | 35.5 (NE-NE) | 31.0 (22.3- 40.8) | 40.2 (32.1-48.4) | 51.4 (50.2-52.8) |
| AGIT | APAT | 35.7 (NE-NE) | 36.5 (26.9-46.8) | 33.1 (26.2-40.7) | 43.4 (42.2-44.6) |
| APAT | APPE | NA | NA | 46.7 (36.9-56.0) | 54.5 (53.1-56.0) |
| APPE | APAT | NA | NA | 30.6 (23.5-37.1) | 37.8 (36.6-38.9) |
| APAT | DISI | 40.4 (NE-NE) | 42.9 (28.6- 58.2) | 47.7 (37.5-58.1) | 58.9 (57.3-60.6) |
| DISI | APAT | 20.2 (NE-NE) | 21.2 (12.7-30.7) | 26.2 (19.5-33.5) | 31.5 (30.5-32.6) |
| APAT | ABER | 49.8 (NE-NE) | 39.1 (25.0- 53.6) | 64.0 (50.0-77.1) | 63.2 (61.5-64.7) |
| ABER | APAT | 16.9 (NE-NE) | 21.2 (12.7-31.1) | 20.0 (13.8-26.3) | 29.0 (27.9-30.0) |
| APAT | PSYC | 36.2 (NE-NE) | 32.9 (23.0- 43.2) | 51.3 (34.2-66.7) | 56.8 (55.0-58.7) |
| PSYC | APAT | 20.6 (NE-NE) | 32.9 (23.3-43.8) | 12.5 ( 7.7-18.0) | 24.3 (23.3-25.4) |
| APAT | DELU | 37.6 (NE-NE) | 36.7 (24.6- 50.0) | 48.4 (29.6-66.7) | 56.9 (54.8-58.9) |
| DELU | APAT | 17.6 (NE-NE) | 25.9 (17.2-35.6) | 9.4 ( 5.1-14.4) | 19.3 (18.4-20.3) |
| APAT | HALL | 21.3 (NE-NE) | 32.7 (19.6- 46.2) | 69.2 (40.0-92.3) | 61.0 (58.2-63.8) |
| HALL | APAT | 6.5 (NE-NE) | 18.8 (10.3-27.5) | 5.6 ( 2.4- 9.6) | 11.1 (10.3-11.9) |
| APAT | ELAT | 78.3 (NE-NE) | 37.5 ( 0.0- 71.6) | 40.0 (18.7-61.9) | 59.7 (56.8-62.8) |
| ELAT | APAT | 11.8 (NE-NE) | 3.5 ( 0.0- 7.8) | 5.0 ( 1.8- 8.3) | 8.4 ( 7.8- 9.1) |
| SLEE | AGIT | NA | NA | 28.8 (20.8-36.4) | 40.7 (39.5-42.0) |
| AGIT | SLEE | NA | NA | 23.3 (17.0-29.8) | 38.8 (37.5-40.1) |
| SLEE | APPE | NA | NA | 31.4 (22.9-40.4) | 45.5 (44.1-47.0) |
| APPE | SLEE | NA | NA | 20.2 (14.5-26.3) | 35.6 (34.3-36.8) |
| SLEE | DISI | NA | NA | 31.8 (22.0-41.0) | 43.6 (42.0-45.2) |
| DISI | SLEE | NA | NA | 17.2 (11.5-23.0) | 26.3 (25.2-27.5) |
| SLEE | ABER | NA | NA | 28.0 (15.5-41.1) | 47.2 (45.5-49.0) |
| ABER | SLEE | NA | NA | 8.6 ( 4.5-13.3) | 24.4 (23.4-25.6) |
| SLEE | PSYC | NA | NA | 28.2 (14.0-43.2) | 46.6 (44.7-48.5) |
| PSYC | SLEE | NA | NA | 6.7 ( 3.3-10.9) | 22.5 (21.4-23.5) |
| SLEE | DELU | NA | NA | 25.8 (10.7-42.1) | 44.4 (42.2-46.5) |
| DELU | SLEE | NA | NA | 4.9 ( 1.9- 8.5) | 17.0 (16.1-18.0) |
| SLEE | HALL | NA | NA | 30.8 ( 5.4-61.5) | 57.9 (55.1-60.8) |
| HALL | SLEE | NA | NA | 2.5 ( 0.5- 5.1) | 11.9 (11.1-12.8) |
| SLEE | ELAT | NA | NA | 30.0 (10.5-52.2) | 46.0 (42.8-49.2) |

| | | | | | |
|---|---|---|---|---|---|
| ELAT | SLEE | NA | NA | 3.7 ( 1.2- 7.0) | 7.3 ( 6.6- 8.0) |
| AGIT | APPE | NA | NA | 34.3 (25.0-44.2) | 41.9 (40.4-43.3) |
| APPE | AGIT | NA | NA | 27.3 (20.0-35.0) | 34.4 (33.0-35.7) |
| AGIT | DISI | 48.5 (NE-NE) | 57.1 (42.2- 72.4) | 50.0 (39.0-60.5) | 58.8 (57.1-60.4) |
| DISI | AGIT | 24.1 (NE-NE) | 24.0 (16.5-32.6) | 33.3 (24.6-41.5) | 37.3 (36.0-38.5) |
| AGIT | ABER | 57.8 (NE-NE) | 54.3 (38.6- 68.0) | 42.0 (28.3-56.7) | 54.8 (53.0-56.5) |
| ABER | AGIT | 19.5 (NE-NE) | 25.0 (16.7-33.6) | 15.9 ( 9.6-22.2) | 29.8 (28.7-31.0) |
| AGIT | PSYC | 47.8 (NE-NE) | 44.7 (34.2- 55.2) | 43.6 (27.9-60.6) | 59.1 (57.3-60.9) |
| PSYC | AGIT | 27.1 (NE-NE) | 38.0 (28.8-47.9) | 12.9 ( 7.6-19.0) | 30.0 (28.6-31.2) |
| AGIT | DELU | 52.0 (NE-NE) | 50.0 (37.1- 62.7) | 48.4 (30.4-66.7) | 63.3 (61.2-65.3) |
| DELU | AGIT | 24.2 (NE-NE) | 30.0 (20.8-39.0) | 11.4 ( 6.2-17.3) | 25.5 (24.3-26.7) |
| AGIT | HALL | 38.0 (NE-NE) | 42.9 (28.3- 56.7) | 30.8 ( 7.1-58.3) | 55.2 (52.6-58.2) |
| HALL | AGIT | 11.5 (NE-NE) | 21.0 (13.0-29.0) | 3.0 ( 0.7- 6.3) | 11.9 (11.1-12.8) |
| AGIT | ELAT | 5.6 (NE-NE) | 37.5 ( 0.0- 75.1) | 50.0 (26.3-70.0) | 57.8 (54.5-61.0) |
| ELAT | AGIT | 0.8 (NE-NE) | 3.0 ( 0.0- 6.5) | 7.6 ( 3.3-12.1) | 9.6 ( 8.9-10.4) |
| APPE | DISI | NA | NA | 25.0 (16.3-34.4) | 41.5 (39.8-43.3) |
| DISI | APPE | NA | NA | 21.0 (13.3-28.9) | 32.1 (30.8-33.3) |
| APPE | ABER | NA | NA | 32.0 (19.1-45.5) | 44.5 (42.8-46.3) |
| ABER | APPE | NA | NA | 15.2 ( 8.7-22.5) | 29.5 (28.1-30.8) |
| APPE | PSYC | NA | NA | 35.9 (21.2-51.3) | 37.7 (35.9-39.4) |
| PSYC | APPE | NA | NA | 13.3 ( 7.1-20.7) | 23.3 (22.0-24.5) |
| APPE | DELU | NA | NA | 35.5 (19.2-53.3) | 37.9 (35.8-39.7) |
| DELU | APPE | NA | NA | 10.5 ( 5.1-16.9) | 18.6 (17.4-19.7) |
| APPE | HALL | NA | NA | 38.5 (11.1-64.7) | 39.2 (36.4-42.2) |
| HALL | APPE | NA | NA | 4.8 ( 1.0- 9.2) | 10.3 ( 9.5-11.3) |
| APPE | ELAT | NA | NA | 35.0 (15.8-57.9) | 49.2 (46.2-52.6) |
| ELAT | APPE | NA | NA | 6.7 ( 2.6-11.9) | 10.0 ( 9.2-10.9) |
| DISI | ABER | 30.2 (NE-NE) | 26.1 (13.2- 38.2) | 46.0 (32.0-60.0) | 45.4 (43.6-47.1) |
| ABER | DISI | 20.4 (NE-NE) | 28.6 (15.4-42.9) | 26.1 (17.1-35.1) | 38.9 (37.2-40.5) |
| DISI | PSYC | 39.5 (NE-NE) | 22.4 (13.5- 31.3) | 25.6 (12.1-40.0) | 38.6 (36.9-40.5) |
| PSYC | DISI | 44.9 (NE-NE) | 45.2 (30.0-60.0) | 11.4 ( 5.1-18.3) | 30.9 (29.4-32.4) |
| DISI | DELU | 44.6 (NE-NE) | 26.7 (15.7- 38.3) | 29.0 (13.5-46.4) | 41.7 (39.8-43.7) |
| DELU | DISI | 41.8 (NE-NE) | 38.1 (23.8-52.9) | 10.2 ( 4.3-17.0) | 26.5 (25.0-28.0) |
| DISI | HALL | 44.3 (NE-NE) | 22.4 (10.8- 34.6) | 15.4 ( 0.0-38.1) | 35.8 (32.9-38.5) |
| HALL | DISI | 27.0 (NE-NE) | 26.2 (13.9-40.0) | 2.3 ( 0.0- 5.8) | 12.2 (11.1-13.3) |

| | | | | | |
|---|---|---|---|---|---|
| DISI | ELAT | NE | NE | 60.0 (39.1-81.8) | 63.7 (60.7-67.0) |
| ELAT | DISI | NE | NE | 13.6 ( 6.8-20.7) | 16.8 (15.6-18.1) |
| ABER | PSYC | 37.2 (NE-NE) | 31.8 (21.8- 43.0) | 23.1 (10.2-36.1) | 38.1 (36.3-39.9) |
| PSYC | ABER | 62.5 (NE-NE) | 58.7 (43.7-72.9) | 18.0 ( 7.7-29.6) | 35.6 (33.9-37.3) |
| ABER | DELU | 38.4 (NE-NE) | 31.7 (20.4- 44.3) | 22.6 ( 8.0-37.1) | 39.2 (37.1-41.2) |
| DELU | ABER | 53.1 (NE-NE) | 41.3 (26.2-55.2) | 14.0 ( 4.5-24.5) | 29.0 (27.4-30.7) |
| ABER | HALL | 29.9 (NE-NE) | 36.7 (23.9- 51.3) | 30.8 ( 5.7-60.0) | 42.6 (40.0-45.5) |
| HALL | ABER | 26.9 (NE-NE) | 39.1 (24.4-53.7) | 8.0 ( 1.5-17.0) | 17.0 (15.5-18.4) |
| ABER | ELAT | 18.9 (NE-NE) | 50.0 (14.2- 88.9) | 20.0 ( 5.0-38.1) | 46.7 (43.5-49.9) |
| ELAT | ABER | 8.4 (NE-NE) | 8.7 ( 1.9-18.2) | 8.0 ( 1.9-15.9) | 14.3 (13.1-15.6) |
| PSYC | ELAT | 20.0 (NE-NE) | 62.5 (25.0-100.0) | 20.0 ( 4.3-38.5) | 32.7 (29.6-35.7) |
| ELAT | PSYC | 5.3 (NE-NE) | 5.9 ( 1.3-11.2) | 10.3 ( 2.2-20.8) | 10.8 ( 9.5-11.9) |
| DELU | HALL | 67.0 (NE-NE) | 49.0 (35.4- 63.3) | 38.5 (12.5-66.7) | 51.9 (48.9-54.6) |
| HALL | DELU | 43.6 (NE-NE) | 40.0 (27.9-52.3) | 16.1 ( 4.2-32.4) | 27.9 (26.0-29.7) |
| DELU | ELAT | 14.3 (NE-NE) | 25.0 ( 0.0- 66.7) | 15.0 ( 0.0-31.6) | 28.1 (25.2-31.1) |
| ELAT | DELU | 4.6 (NE-NE) | 3.3 ( 0.0- 8.6) | 9.7 ( 0.0-21.1) | 11.7 (10.3-13.0) |
| HALL | ELAT | 20.0 (NE-NE) | 62.5 (25.0-100.0) | 10.0 ( 0.0-26.1) | 13.4 (11.2-15.6) |
| ELAT | HALL | 9.9 (NE-NE) | 10.2 ( 2.4-19.2) | 15.4 ( 0.0-40.0) | 10.3 ( 8.6-12.1) |

S1|S2, conditional prevalence of symptom S1 given presence of symptom S2.

Displayed are conditional prevalence (%) and 95% confidence intervals. Survey estimates for ADAMS are based on survey population weights, confidence intervals are not estimated.

NA, Not applicable; NE, Not estimable.

ABER, Aberrant motor behavior; AGIT, Agitation/aggression; ANXI, Anxiety; APAT, Apathy/indifference; APPE, Appetite/eating changes; DELU, Delusions; DEPR, Depression/dysphoria; DISI, Disinhibition; ELAT, Elation/euphoria; HALU, Hallucinations; IRRI, Irritability/lability; PSYC, Psychotic symptoms (hallucinations and/or delusions); SLEE, Nighttime behavioral disturbances.

Supplemental Table 23. Conditional prevalence S1|S2 of neuropsychiatric symptoms with the NPI severity score > 1

| S1 | S2 | ADAMS survey | ADAMS | ADNI | NACC |
|---|---|---|---|---|---|
| DEPR | IRRI | 43.5 (NE-NE) | 33.3 ( 17.9- 48.7) | 27.7 ( 17.2- 38.3) | 37.2 (35.7-39.0) |
| IRRI | DEPR | 28.3 (NE-NE) | 25.6 (12.9-39.1) | 40.9 (26.2-55.6) | 34.9 (33.3-36.7) |
| DEPR | ANXI | 34.6 (NE-NE) | 40.5 ( 24.3- 56.4) | 32.4 ( 18.2- 47.8) | 44.2 (42.5-46.0) |
| ANXI | DEPR | 25.2 (NE-NE) | 34.9 (21.1-50.0) | 27.3 (14.3-41.3) | 43.2 (41.5-45.1) |
| DEPR | APAT | 38.4 (NE-NE) | 36.2 ( 24.4- 49.2) | 19.0 ( 9.3- 30.0) | 36.0 (34.5-37.8) |
| APAT | DEPR | 44.6 (NE-NE) | 48.8 (33.3-64.8) | 25.0 (12.5-37.5) | 39.0 (37.2-40.9) |
| DEPR | SLEE | NA | NA | 19.6 ( 9.6- 30.6) | 31.5 (29.9-33.4) |
| SLEE | DEPR | NA | NA | 25.0 (13.2-37.5) | 29.8 (28.2-31.6) |
| DEPR | AGIT | 50.0 (NE-NE) | 26.7 ( 15.1- 40.0) | 17.6 ( 6.1- 31.3) | 38.5 (36.6-40.4) |
| AGIT | DEPR | 32.9 (NE-NE) | 27.9 (15.9-42.6) | 13.6 ( 4.6-25.6) | 30.9 (29.3-32.6) |
| DEPR | APPE | NA | NA | 16.1 ( 6.4- 26.3) | 31.1 (29.1-33.2) |
| APPE | DEPR | NA | NA | 20.5 ( 8.3-34.1) | 20.9 (19.5-22.4) |
| DEPR | DISI | 70.4 (NE-NE) | 33.3 ( 8.3- 58.3) | 25.0 ( 8.7- 44.4) | 32.4 (30.2-34.6) |
| DISI | DEPR | 22.7 (NE-NE) | 11.6 ( 2.5-22.2) | 13.6 ( 4.5-24.4) | 17.2 (16.0-18.6) |
| DEPR | ABER | 53.6 (NE-NE) | 14.3 ( 3.2- 29.0) | 11.8 ( 0.0- 31.2) | 31.7 (29.1-34.2) |
| ABER | DEPR | 20.0 (NE-NE) | 9.3 ( 2.2-18.6) | 4.5 ( 0.0-12.1) | 15.7 (14.4-17.1) |
| DEPR | PSYC | 44.6 (NE-NE) | 29.4 ( 14.7- 46.7) | 31.2 ( 8.3- 56.3) | 35.8 (33.1-38.3) |
| PSYC | DEPR | 24.9 (NE-NE) | 23.3 (11.1-37.1) | 11.4 ( 2.5-22.2) | 15.8 (14.4-17.2) |
| DEPR | DELU | 44.0 (NE-NE) | 29.0 ( 14.3- 46.9) | 33.3 ( 9.1- 60.0) | 36.9 (33.9-39.7) |
| DELU | DEPR | 23.5 (NE-NE) | 20.9 ( 9.4-34.1) | 11.4 ( 2.5-22.2) | 13.1 (11.9-14.4) |
| DEPR | HALL | 35.5 (NE-NE) | 33.3 ( 0.0- 61.6) | NE | 36.6 (32.5-41.1) |
| HALL | DEPR | 3.1 (NE-NE) | 9.3 ( 0.0-19.0) | NE | 6.3 ( 5.4- 7.2) |
| DEPR | ELAT | NE | NE | 16.7 ( 0.0- 60.0) | 32.6 (28.2-37.8) |
| ELAT | DEPR | NE | NE | 2.3 ( 0.0- 7.7) | 4.1 ( 3.4- 4.9) |
| IRRI | ANXI | 9.5 (NE-NE) | 27.0 ( 12.9- 42.5) | 37.8 ( 23.7- 53.1) | 38.9 (37.0-40.8) |
| ANXI | IRRI | 10.7 (NE-NE) | 30.3 (13.8-46.2) | 21.5 (12.8-32.1) | 40.6 (38.7-42.5) |
| IRRI | APAT | 19.4 (NE-NE) | 19.0 ( 8.7- 30.1) | 25.9 ( 15.3- 38.3) | 34.1 (32.5-35.8) |
| APAT | IRRI | 34.6 (NE-NE) | 33.3 (16.7-51.3) | 23.1 (13.6-33.9) | 39.3 (37.7-41.2) |
| IRRI | SLEE | NA | NA | 19.6 ( 9.5- 30.8) | 31.1 (29.3-32.7) |
| SLEE | IRRI | NA | NA | 16.9 ( 8.6-26.4) | 31.3 (29.6-33.0) |
| IRRI | AGIT | 61.6 (NE-NE) | 53.3 ( 38.1- 68.0) | 44.1 ( 27.3- 60.0) | 57.3 (55.2-59.2) |
| AGIT | IRRI | 62.4 (NE-NE) | 72.7 (57.1-87.5) | 23.1 (13.6-34.2) | 49.0 (47.2-51.1) |

| | | | | | |
|---|---|---|---|---|---|
| IRRI | APPE | NA | NA | 19.6 ( 9.7- 31.5) | 34.3 (32.0-36.3) |
| APPE | IRRI | NA | NA | 16.9 ( 8.4-27.9) | 24.6 (22.8-26.2) |
| IRRI | DISI | 51.8 (NE-NE) | 66.7 ( 40.0- 90.9) | 50.0 ( 30.0- 70.6) | 53.9 (51.6-56.5) |
| DISI | IRRI | 25.7 (NE-NE) | 30.3 (15.4-46.7) | 18.5 (10.3-27.9) | 30.6 (28.8-32.3) |
| IRRI | ABER | 34.2 (NE-NE) | 21.4 ( 7.4- 37.5) | 23.5 ( 5.9- 45.5) | 44.2 (41.4-46.7) |
| ABER | IRRI | 19.7 (NE-NE) | 18.2 ( 6.1-32.5) | 6.2 ( 1.4-12.5) | 23.3 (21.7-24.9) |
| IRRI | PSYC | 33.2 (NE-NE) | 38.2 ( 20.7- 55.6) | 50.0 ( 26.7- 73.7) | 41.5 (38.8-44.1) |
| PSYC | IRRI | 28.6 (NE-NE) | 39.4 (22.7-58.1) | 12.3 ( 5.1-21.7) | 19.5 (18.1-21.1) |
| IRRI | DELU | 31.2 (NE-NE) | 35.5 ( 18.7- 53.1) | 46.7 ( 22.2- 72.7) | 45.0 (42.1-48.1) |
| DELU | IRRI | 25.7 (NE-NE) | 33.3 (17.2-50.0) | 10.8 ( 4.1-19.7) | 17.1 (15.7-18.6) |
| IRRI | HALL | 42.1 (NE-NE) | 50.0 ( 16.7- 77.8) | 33.3 ( 0.0-100.0) | 36.2 (32.0-40.4) |
| HALL | IRRI | 5.6 (NE-NE) | 18.2 ( 4.8-33.3) | 1.5 ( 0.0- 5.1) | 6.6 ( 5.7- 7.6) |
| IRRI | ELAT | NE | NE | 33.3 ( 0.0- 75.0) | 50.7 (46.0-56.4) |
| ELAT | IRRI | NE | NE | 3.1 ( 0.0- 7.1) | 6.8 ( 5.9- 7.7) |
| ANXI | APAT | 16.4 (NE-NE) | 25.9 ( 15.5- 37.8) | 20.7 ( 9.8- 31.8) | 33.4 (31.8-35.1) |
| APAT | ANXI | 26.2 (NE-NE) | 40.5 (26.2-57.1) | 32.4 (16.0-48.3) | 36.9 (35.1-38.8) |
| ANXI | SLEE | NA | NA | 8.9 ( 2.0- 17.2) | 30.9 (29.2-32.6) |
| SLEE | ANXI | NA | NA | 13.5 ( 2.9-25.0) | 29.8 (28.3-31.5) |
| ANXI | AGIT | 11.2 (NE-NE) | 22.2 ( 10.9- 35.7) | 26.5 ( 12.5- 41.7) | 39.8 (37.7-41.7) |
| AGIT | ANXI | 10.2 (NE-NE) | 27.0 (12.8-41.7) | 24.3 (12.1-40.6) | 32.6 (30.9-34.2) |
| ANXI | APPE | NA | NA | 14.3 ( 4.7- 24.0) | 31.8 (29.6-34.0) |
| APPE | ANXI | NA | NA | 21.6 ( 7.7-35.5) | 21.8 (20.2-23.4) |
| ANXI | DISI | 15.0 (NE-NE) | 40.0 ( 16.7- 63.6) | 16.7 ( 3.7- 32.1) | 39.6 (37.2-42.2) |
| DISI | ANXI | 6.7 (NE-NE) | 16.2 ( 5.7-29.4) | 10.8 ( 2.4-22.5) | 21.5 (20.1-23.0) |
| ANXI | ABER | 18.2 (NE-NE) | 28.6 ( 12.5- 47.4) | 17.6 ( 0.0- 38.5) | 43.5 (40.9-46.0) |
| ABER | ANXI | 9.4 (NE-NE) | 21.6 ( 9.1-35.5) | 8.1 ( 0.0-17.5) | 21.9 (20.4-23.5) |
| ANXI | PSYC | 12.4 (NE-NE) | 26.5 ( 12.0- 41.0) | 25.0 ( 6.2- 50.0) | 41.3 (38.6-44.0) |
| PSYC | ANXI | 9.5 (NE-NE) | 24.3 (10.4-38.7) | 10.8 ( 2.4-21.4) | 18.6 (17.2-20.1) |
| ANXI | DELU | 10.3 (NE-NE) | 25.8 ( 10.7- 41.7) | 26.7 ( 6.7- 50.0) | 41.9 (38.9-44.8) |
| DELU | ANXI | 7.6 (NE-NE) | 21.6 ( 8.8-35.3) | 10.8 ( 2.4-21.4) | 15.2 (13.9-16.6) |
| ANXI | HALL | 40.1 (NE-NE) | 41.7 ( 11.1- 70.0) | 33.3 ( 0.0-100.0) | 45.2 (40.8-49.7) |
| HALL | ANXI | 4.8 (NE-NE) | 13.5 ( 2.9-25.0) | 2.7 ( 0.0- 8.9) | 7.9 ( 6.9- 8.9) |
| ANXI | ELAT | NE | NE | 16.7 ( 0.0- 50.0) | 41.1 (36.3-46.5) |
| ELAT | ANXI | 14.5 (NE-NE) | 2.7 ( 0.0- 9.7) | 2.7 ( 0.0- 9.1) | 5.2 ( 4.4- 6.1) |
| APAT | SLEE | NA | NA | 16.1 ( 7.5- 26.1) | 37.1 (35.3-38.9) |

| | | | | | |
|---|---|---|---|---|---|
| SLEE | APAT | NA | NA | 15.5 ( 7.1-25.0) | 32.5 (30.8-34.2) |
| APAT | AGIT | 45.7 (NE-NE) | 33.3 ( 20.0- 47.5) | 44.1 ( 27.3- 61.1) | 43.8 (41.9-45.8) |
| AGIT | APAT | 25.9 (NE-NE) | 25.9 (15.6-37.9) | 25.9 (15.4-38.3) | 32.5 (31.0-34.2) |
| APAT | APPE | NA | NA | 32.1 ( 19.2- 44.8) | 45.0 (42.9-47.1) |
| APPE | APAT | NA | NA | 31.0 (18.5-42.9) | 27.9 (26.4-29.6) |
| APAT | DISI | 44.9 (NE-NE) | 33.3 ( 9.1- 58.3) | 25.0 ( 8.7- 45.0) | 50.0 (47.6-52.7) |
| DISI | APAT | 12.4 (NE-NE) | 8.6 ( 1.9-16.7) | 10.3 ( 3.4-19.6) | 24.6 (23.1-26.1) |
| APAT | ABER | 58.2 (NE-NE) | 28.6 ( 13.0- 46.2) | 47.1 ( 23.5- 72.7) | 53.8 (51.2-56.4) |
| ABER | APAT | 18.7 (NE-NE) | 13.8 ( 5.7-23.9) | 13.8 ( 5.9-23.7) | 24.6 (23.0-26.2) |
| APAT | PSYC | 43.2 (NE-NE) | 29.4 ( 15.6- 46.7) | 43.8 ( 20.0- 69.2) | 42.1 (39.5-44.8) |
| PSYC | APAT | 20.8 (NE-NE) | 17.2 ( 8.5-27.8) | 12.1 ( 4.3-21.4) | 17.2 (15.9-18.5) |
| APAT | DELU | 42.6 (NE-NE) | 29.0 ( 14.3- 46.4) | 40.0 ( 14.3- 64.3) | 43.4 (40.5-46.4) |
| DELU | APAT | 19.6 (NE-NE) | 15.5 ( 7.1-25.0) | 10.3 ( 3.3-19.0) | 14.3 (13.0-15.6) |
| APAT | HALL | 32.0 (NE-NE) | 25.0 ( 0.0- 52.9) | NE | 45.0 (40.8-49.4) |
| HALL | APAT | 2.4 (NE-NE) | 5.2 ( 0.0-12.1) | 5.2 ( 0.0-11.4) | 7.1 ( 6.2- 8.0) |
| APAT | ELAT | NE | NE | 16.7 ( 0.0- 50.0) | 54.4 (49.0-59.8) |
| ELAT | APAT | NE | NE | 1.7 ( 0.0- 5.7) | 6.3 ( 5.4- 7.2) |
| SLEE | AGIT | NA | NA | 5.9 ( 0.0- 14.8) | 31.8 (30.0-33.6) |
| AGIT | SLEE | NA | NA | 3.6 ( 0.0- 8.9) | 27.0 (25.4-28.6) |
| SLEE | APPE | NA | NA | 12.5 ( 4.1- 21.7) | 37.6 (35.5-39.8) |
| APPE | SLEE | NA | NA | 12.5 ( 4.0-21.7) | 26.7 (24.9-28.4) |
| SLEE | DISI | NA | NA | 20.8 ( 5.3- 38.1) | 35.2 (33.0-37.6) |
| DISI | SLEE | NA | NA | 8.9 ( 2.0-17.0) | 19.8 (18.4-21.2) |
| SLEE | ABER | NA | NA | 29.4 ( 7.7- 53.8) | 39.2 (36.7-41.6) |
| ABER | SLEE | NA | NA | 8.9 ( 1.9-16.4) | 20.5 (19.0-22.1) |
| SLEE | PSYC | NA | NA | 12.5 ( 0.0- 30.8) | 36.4 (33.7-39.3) |
| PSYC | SLEE | NA | NA | 3.6 ( 0.0- 9.2) | 17.0 (15.6-18.4) |
| SLEE | DELU | NA | NA | 6.7 ( 0.0- 22.2) | 33.9 (30.9-37.1) |
| DELU | SLEE | NA | NA | 1.8 ( 0.0- 5.8) | 12.8 (11.5-14.0) |
| SLEE | HALL | NA | NA | 33.3 ( 0.0-100.0) | 50.4 (46.0-54.9) |
| HALL | SLEE | NA | NA | 1.8 ( 0.0- 6.1) | 9.1 ( 8.0-10.1) |
| SLEE | ELAT | NA | NA | 16.7 ( 0.0- 60.0) | 39.1 (34.0-44.5) |
| ELAT | SLEE | NA | NA | 1.8 ( 0.0- 5.9) | 5.2 ( 4.4- 6.0) |
| AGIT | APPE | NA | NA | 12.5 ( 4.3- 22.0) | 30.0 (27.8-32.1) |
| APPE | AGIT | NA | NA | 20.6 ( 7.7-34.8) | 25.0 (23.2-26.8) |

| | | | | | |
|---|---|---|---|---|---|
| AGIT | DISI | 46.8 (NE-NE) | 60.0 ( 33.3- 85.7) | 33.3 ( 13.6- 55.0) | 48.1 (45.6-50.6) |
| DISI | AGIT | 22.9 (NE-NE) | 20.0 ( 9.3-31.6) | 23.5 ( 9.5-39.5) | 31.9 (30.0-33.7) |
| AGIT | ABER | 43.7 (NE-NE) | 39.3 ( 21.7- 59.3) | 11.8 ( 0.0- 30.0) | 41.9 (39.4-44.7) |
| ABER | AGIT | 24.8 (NE-NE) | 24.4 (12.5-37.5) | 5.9 ( 0.0-14.3) | 25.8 (24.1-27.7) |
| AGIT | PSYC | 31.5 (NE-NE) | 41.2 ( 23.8- 60.0) | 37.5 ( 14.3- 62.5) | 46.4 (43.4-49.2) |
| PSYC | AGIT | 26.8 (NE-NE) | 31.1 (17.8-45.2) | 17.6 ( 6.1-32.4) | 25.6 (23.6-27.5) |
| AGIT | DELU | 32.1 (NE-NE) | 41.9 ( 24.0- 60.7) | 40.0 ( 15.4- 65.0) | 50.3 (47.0-53.6) |
| DELU | AGIT | 26.1 (NE-NE) | 28.9 (16.3-42.1) | 17.6 ( 6.1-32.4) | 22.3 (20.5-24.1) |
| AGIT | HALL | 29.6 (NE-NE) | 41.7 ( 12.5- 72.7) | NE | 40.3 (35.8-44.9) |
| HALL | AGIT | 3.9 (NE-NE) | 11.1 ( 2.4-21.7) | NE | 8.6 ( 7.4- 9.8) |
| AGIT | ELAT | NE | NE | 33.3 ( 0.0- 75.0) | 48.4 (43.0-53.9) |
| ELAT | AGIT | NE | NE | 5.9 ( 0.0-15.4) | 7.5 ( 6.5- 8.7) |
| APPE | DISI | NA | NA | 25.0 ( 9.1- 44.4) | 33.6 (31.1-36.0) |
| DISI | APPE | NA | NA | 10.7 ( 3.4-19.3) | 26.6 (24.5-28.5) |
| APPE | ABER | NA | NA | 29.4 ( 7.1- 53.3) | 35.0 (32.5-37.8) |
| ABER | APPE | NA | NA | 8.9 ( 1.8-16.9) | 25.8 (23.7-27.6) |
| APPE | PSYC | NA | NA | 25.0 ( 5.9- 50.0) | 26.0 (23.7-28.5) |
| PSYC | APPE | NA | NA | 7.1 ( 1.6-15.0) | 17.1 (15.5-18.8) |
| APPE | DELU | NA | NA | 26.7 ( 6.2- 53.3) | 26.8 (24.1-29.6) |
| DELU | APPE | NA | NA | 7.1 ( 1.6-15.0) | 14.2 (12.6-15.8) |
| APPE | HALL | NA | NA | 33.3 ( 0.0-100.0) | 26.2 (22.4-30.1) |
| HALL | APPE | NA | NA | 1.8 ( 0.0- 6.0) | 6.7 ( 5.7- 7.8) |
| APPE | ELAT | NA | NA | 33.3 ( 0.0- 75.0) | 47.9 (41.8-52.9) |
| ELAT | APPE | NA | NA | 3.6 ( 0.0- 8.9) | 8.9 ( 7.5-10.2) |
| DISI | ABER | 37.0 (NE-NE) | 25.0 ( 9.5- 41.7) | 17.6 ( 0.0- 38.9) | 37.4 (35.0-40.0) |
| ABER | DISI | 42.9 (NE-NE) | 46.7 (21.1-75.0) | 12.5 ( 0.0-26.9) | 34.8 (32.4-37.1) |
| DISI | PSYC | 27.0 (NE-NE) | 20.6 ( 7.7- 36.1) | 25.0 ( 5.9- 50.0) | 28.1 (25.7-30.6) |
| PSYC | DISI | 46.9 (NE-NE) | 46.7 (21.4-72.7) | 16.7 ( 3.7-33.3) | 23.4 (21.2-25.5) |
| DISI | DELU | 25.6 (NE-NE) | 19.4 ( 6.2- 35.1) | 26.7 ( 5.9- 50.0) | 30.6 (28.0-33.5) |
| DELU | DISI | 42.5 (NE-NE) | 40.0 (15.4-66.7) | 16.7 ( 3.7-33.3) | 20.5 (18.4-22.6) |
| DISI | HALL | 27.4 (NE-NE) | 25.0 ( 0.0- 50.0) | NE | 27.9 (23.8-31.9) |
| HALL | DISI | 7.4 (NE-NE) | 20.0 ( 0.0-42.9) | NE | 9.0 ( 7.6-10.4) |
| DISI | ELAT | NE | NE | 66.7 ( 20.0-100.0) | 59.8 (54.7-64.9) |
| ELAT | DISI | NE | NE | 16.7 ( 3.7-32.4) | 14.0 (12.4-15.8) |
| ABER | PSYC | 40.9 (NE-NE) | 29.4 ( 14.3- 46.2) | 18.8 ( 0.0- 38.5) | 29.7 (27.2-32.3) |

| | | | | | |
|---|---|---|---|---|---|
| PSYC | ABER | 61.2 (NE-NE) | 35.7 (17.9-52.9) | 17.6 ( 0.0-37.5) | 26.6 (24.4-28.9) |
| ABER | DELU | 41.7 (NE-NE) | 29.0 ( 12.9- 46.2) | 13.3 ( 0.0- 33.3) | 31.6 (28.7-34.5) |
| DELU | ABER | 59.6 (NE-NE) | 32.1 (15.2-48.6) | 11.8 ( 0.0-27.3) | 22.8 (20.7-24.9) |
| ABER | HALL | 17.8 (NE-NE) | 25.0 ( 0.0- 53.9) | 66.7 ( 0.0-100.0) | 34.1 (30.0-38.1) |
| HALL | ABER | 4.1 (NE-NE) | 10.7 ( 0.0-22.2) | 11.8 ( 0.0-28.6) | 11.8 (10.2-13.4) |
| ABER | ELAT | NE | NE | 33.3 ( 0.0- 75.0) | 42.8 (37.6-48.1) |
| ELAT | ABER | NE | NE | 11.8 ( 0.0-30.0) | 10.8 ( 9.2-12.5) |
| PSYC | ELAT | NE | NE | 50.0 ( 0.0-100.0) | 23.8 (19.3-28.8) |
| ELAT | PSYC | NE | NE | 18.8 ( 0.0-37.5) | 6.7 ( 5.3- 8.3) |
| DELU | HALL | 71.4 (NE-NE) | 75.0 ( 50.0-100.0) | 66.7 ( 0.0-100.0) | 49.6 (45.3-53.9) |
| HALL | DELU | 11.6 (NE-NE) | 29.0 (13.2-46.2) | 13.3 ( 0.0-33.3) | 23.9 (21.0-26.5) |
| DELU | ELAT | NE | NE | 50.0 ( 0.0-100.0) | 21.2 (17.1-25.8) |
| ELAT | DELU | NE | NE | 20.0 ( 0.0-40.9) | 7.5 ( 5.8- 9.2) |
| HALL | ELAT | NE | NE | NE | 8.5 ( 5.7-11.4) |
| ELAT | HALL | NE | NE | NE | 6.2 ( 4.2- 8.4) |

S1|S2, conditional prevalence of symptom S1 given presence of symptom S2.
Displayed are conditional prevalence (%) and 95% confidence intervals. Survey estimates for ADAMS are based on survey population weights, confidence intervals are not estimated.
NA, Not applicable; NE, Not estimable.
ABER, Aberrant motor behavior; AGIT, Agitation/aggression; ANXI, Anxiety; APAT, Apathy/indifference; APPE, Appetite/eating changes; DELU, Delusions; DEPR, Depression/dysphoria; DISI, Disinhibition; ELAT, Elation/euphoria; HALU, Hallucinations; IRRI, Irritability/lability; PSYC, Psychotic symptoms (hallucinations and/or delusions); SLEE, Nighttime behavioral disturbances.

Supplemental Table 24. Conditional prevalence S1|S2 of neuropsychiatric symptoms with the NPI severity score > 0 per MMSE stratum (ADAMS and ADNI studies only)

| S1 | S2 | ADAMS survey | | ADAMS | | ADNI | |
|---|---|---|---|---|---|---|---|
| | | MMSE>26 | MMSE ≤26 | MMSE >26 | MMSE ≤26 | MMSE >26 | MMSE ≤26 |
| DEPR | IRRI | 47.8 (NE-NE) | 66.1 (NE-NE) | 42.1 ( 18.7- 64.7) | 50.7 (38.3- 62.3) | 42.9 ( 33.9- 51.2) | 53.7 (42.5- 63.6) |
| IRRI | DEPR | 28.2 (NE-NE) | 29.1 (NE-NE) | 25.0 (10.7-40.0) | 28.2 (20.5-36.4) | 40.8 (32.0-49.6) | 47.3 (37.0-58.0) |
| DEPR | ANXI | 56.6 (NE-NE) | 65.9 (NE-NE) | 55.6 ( 31.2- 80.0) | 62.3 (50.7- 74.2) | 59.0 ( 46.6- 71.4) | 49.3 (37.9- 60.3) |
| ANXI | DEPR | 30.7 (NE-NE) | 24.8 (NE-NE) | 31.2 (15.8-48.2) | 34.7 (27.1-43.8) | 28.8 (21.2-37.2) | 37.6 (27.3-48.0) |
| DEPR | APAT | 59.1 (NE-NE) | 75.5 (NE-NE) | 46.2 ( 17.6- 77.8) | 66.7 (55.9- 77.6) | 60.0 ( 47.7- 71.7) | 48.0 (37.9- 57.5) |
| APAT | DEPR | 24.8 (NE-NE) | 34.4 (NE-NE) | 18.8 ( 5.5-33.4) | 38.7 (31.0-48.0) | 28.8 (20.7-36.7) | 51.6 (41.5-61.5) |
| DEPR | SLEE | NA | NA | NA | NA | 41.5 ( 32.4- 50.5) | 52.6 (39.5- 65.9) |
| SLEE | DEPR | NA | NA | NA | NA | 35.2 (27.1-43.1) | 32.3 (23.2-41.8) |
| DEPR | AGIT | 67.0 (NE-NE) | 64.5 (NE-NE) | 58.3 ( 28.6- 88.9) | 46.6 (36.0- 56.6) | 44.6 ( 32.9- 55.6) | 50.0 (37.0- 62.7) |
| AGIT | DEPR | 20.9 (NE-NE) | 35.4 (NE-NE) | 21.9 ( 9.1-36.4) | 33.1 (25.7-41.7) | 26.4 (19.0-34.4) | 31.2 (22.0-41.2) |
| DEPR | APPE | NA | NA | NA | NA | 51.3 ( 35.5- 66.7) | 50.0 (38.2- 61.7) |
| APPE | DEPR | NA | NA | NA | NA | 16.0 ( 9.8-22.7) | 35.5 (26.4-45.6) |
| DEPR | DISI | 54.1 (NE-NE) | 54.3 (NE-NE) | 60.0 ( 0.0-100.0) | 51.4 (33.3- 68.6) | 49.0 ( 35.0- 62.5) | 56.4 (41.5- 71.4) |
| DISI | DEPR | 8.3 (NE-NE) | 14.9 (NE-NE) | 9.4 ( 0.0-21.9) | 15.3 ( 9.1-21.8) | 19.2 (12.6-26.8) | 23.7 (15.8-32.7) |
| DEPR | ABER | NE | 50.4 (NE-NE) | NE | 32.6 (20.4- 45.5) | 41.7 ( 11.1- 72.7) | 65.8 (50.0- 80.0) |
| ABER | DEPR | NE | 13.8 (NE-NE) | NE | 12.1 ( 7.0-17.9) | 4.0 ( 0.8- 7.6) | 26.9 (17.6-35.9) |
| DEPR | PSYC | NE | 57.1 (NE-NE) | NE | 40.5 (30.3- 51.5) | 85.7 ( 50.0-100.0) | 53.1 (35.3- 71.0) |
| PSYC | DEPR | NE | 23.9 (NE-NE) | NE | 27.4 (19.5-35.5) | 4.8 ( 1.6- 9.0) | 18.3 (11.0-26.2) |
| DEPR | DELU | NE | 60.3 (NE-NE) | NE | 39.0 (26.3- 51.5) | NE | 60.0 (40.0- 78.3) |
| DELU | DEPR | NE | 20.3 (NE-NE) | NE | 18.5 (12.0-26.0) | 4.8 ( 1.6- 9.0) | 16.1 ( 9.2-23.6) |
| DEPR | HALL | NE | 50.1 (NE-NE) | NE | 44.9 (30.8- 58.2) | NE | 50.0 (20.0- 80.0) |
| HALL | DEPR | NE | 12.3 (NE-NE) | NE | 17.7 (11.3-25.0) | NE | 6.5 ( 1.8-12.0) |
| DEPR | ELAT | 50.0 (NE-NE) | NE | 50.0 ( 0.0-100.0) | NE | 60.0 ( 25.0- 91.7) | 40.0 (11.1- 71.4) |
| ELAT | DEPR | 5.6 (NE-NE) | NE | 3.1 ( 0.0-10.7) | NE | 4.8 ( 1.5- 8.9) | 4.3 ( 1.0- 8.8) |
| IRRI | ANXI | 37.3 (NE-NE) | 23.3 (NE-NE) | 38.9 ( 15.8- 62.5) | 40.6 (29.7- 52.6) | 49.2 ( 36.8- 61.0) | 52.1 (40.5- 64.2) |
| ANXI | IRRI | 34.3 (NE-NE) | 19.9 (NE-NE) | 36.8 (14.3-61.5) | 40.6 (29.1-52.5) | 25.2 (17.9-33.1) | 45.1 (34.5-56.6) |
| IRRI | APAT | 23.4 (NE-NE) | 39.1 (NE-NE) | 23.1 ( 0.0- 50.0) | 34.7 (23.7- 46.3) | 56.7 ( 44.8- 69.5) | 44.0 (34.7- 55.0) |
| APAT | IRRI | 16.6 (NE-NE) | 40.4 (NE-NE) | 15.8 ( 0.0-35.3) | 36.2 (24.7-47.5) | 28.6 (20.7-37.0) | 53.7 (43.2-65.2) |

| | | | | | | | |
|---|---|---|---|---|---|---|---|
| IRRI | SLEE | NA | NA | NA | NA | 34.0 ( 26.0- 42.6) | 40.4 (28.1- 53.4) |
| SLEE | IRRI | NA | NA | NA | NA | 30.3 (23.1-38.7) | 28.0 (18.6-38.0) |
| IRRI | AGIT | 75.0 (NE-NE) | 59.8 (NE-NE) | 75.0 ( 50.0-100.0) | 55.7 (44.9- 65.3) | 70.3 ( 59.4- 80.0) | 56.9 (44.4- 70.0) |
| AGIT | IRRI | 39.6 (NE-NE) | 74.8 (NE-NE) | 47.4 (23.8-70.0) | 71.0 (60.0-82.2) | 43.7 (35.2-52.7) | 40.2 (29.6-51.9) |
| IRRI | APPE | NA | NA | NA | NA | 43.6 ( 27.8- 60.0) | 37.9 (26.4- 50.7) |
| APPE | IRRI | NA | NA | NA | NA | 14.3 ( 8.5-20.8) | 30.5 (20.2-40.9) |
| IRRI | DISI | 55.1 (NE-NE) | 57.7 (NE-NE) | 40.0 ( 0.0-100.0) | 62.2 (45.5- 78.1) | 61.2 ( 46.8- 74.6) | 56.4 (41.9- 72.2) |
| DISI | IRRI | 14.4 (NE-NE) | 36.0 (NE-NE) | 10.5 ( 0.0-26.7) | 33.3 (22.5-44.6) | 25.2 (17.2-32.7) | 26.8 (17.8-36.0) |
| IRRI | ABER | NE | 40.8 (NE-NE) | NE | 41.3 (26.8- 54.3) | 41.7 ( 12.5- 70.0) | 42.1 (25.7- 58.6) |
| ABER | IRRI | NE | 25.4 (NE-NE) | NE | 27.5 (17.3-38.5) | 4.2 ( 0.9- 8.2) | 19.5 (10.6-27.7) |
| IRRI | PSYC | NE | 42.3 (NE-NE) | NE | 39.3 (28.2- 49.5) | NE | 43.8 (25.9- 61.1) |
| PSYC | IRRI | NE | 40.2 (NE-NE) | NE | 47.8 (35.8-60.0) | 5.9 ( 1.9-10.5) | 17.1 ( 9.1-25.8) |
| IRRI | DELU | NE | 42.9 (NE-NE) | NE | 37.3 (24.5- 50.0) | NE | 48.0 (28.0- 69.2) |
| DELU | IRRI | NE | 32.9 (NE-NE) | NE | 31.9 (20.6-43.9) | 5.0 ( 1.6- 9.4) | 14.6 ( 7.4-22.7) |
| IRRI | HALL | NE | 40.6 (NE-NE) | NE | 44.9 (31.2- 58.5) | NE | 33.3 ( 7.7- 62.5) |
| HALL | IRRI | NE | 22.7 (NE-NE) | NE | 31.9 (21.3-42.9) | 0.8 ( 0.0- 2.8) | 4.9 ( 1.1-10.0) |
| IRRI | ELAT | NE | 19.6 (NE-NE) | NE | 33.3 ( 0.0- 75.0) | 60.0 ( 27.3- 90.0) | 40.0 (11.1- 72.8) |
| ELAT | IRRI | NE | 1.2 (NE-NE) | NE | 2.9 ( 0.0- 6.9) | 5.0 ( 1.6- 9.4) | 4.9 ( 1.2- 9.9) |
| ANXI | APAT | 41.0 (NE-NE) | 28.0 (NE-NE) | 38.5 ( 12.5- 66.7) | 38.9 (27.6- 50.7) | 30.0 ( 18.5- 40.7) | 39.0 (29.5- 48.9) |
| APAT | ANXI | 31.7 (NE-NE) | 33.9 (NE-NE) | 27.8 ( 7.7-50.0) | 40.6 (29.4-52.3) | 29.5 (17.6-40.9) | 54.9 (42.9-66.1) |
| ANXI | SLEE | NA | NA | NA | NA | 26.4 ( 18.1- 36.1) | 38.6 (26.1- 51.7) |
| SLEE | ANXI | NA | NA | NA | NA | 45.9 (33.9-58.6) | 31.0 (20.6-41.8) |
| ANXI | AGIT | 46.0 (NE-NE) | 18.8 (NE-NE) | 41.7 ( 14.3- 71.5) | 34.1 (24.4- 44.0) | 32.4 ( 22.2- 43.0) | 53.4 (40.7- 66.7) |
| AGIT | ANXI | 26.4 (NE-NE) | 27.4 (NE-NE) | 27.8 ( 7.1-50.0) | 43.5 (31.8-55.6) | 39.3 (27.7-51.0) | 43.7 (31.7-56.2) |
| ANXI | APPE | NA | NA | NA | NA | 30.8 ( 16.7- 45.7) | 30.3 (19.4- 42.3) |
| APPE | ANXI | NA | NA | NA | NA | 19.7 (10.3-30.0) | 28.2 (17.6-39.3) |
| ANXI | DISI | 26.5 (NE-NE) | 28.1 (NE-NE) | 20.0 ( 0.0- 66.7) | 54.1 (38.2- 69.4) | 36.7 ( 24.0- 51.0) | 43.6 (28.6- 59.5) |
| DISI | ANXI | 7.5 (NE-NE) | 20.6 (NE-NE) | 5.6 ( 0.0-20.0) | 29.0 (18.3-39.1) | 29.5 (18.2-40.7) | 23.9 (14.6-34.3) |
| ANXI | ABER | NE | 27.9 (NE-NE) | NE | 41.3 (26.5- 54.8) | 50.0 ( 20.0- 77.8) | 44.7 (29.2- 59.0) |
| ABER | ANXI | NE | 20.3 (NE-NE) | NE | 27.5 (16.9-38.2) | 9.8 ( 3.2-17.9) | 23.9 (13.9-33.8) |
| ANXI | PSYC | NE | 23.1 (NE-NE) | NE | 32.1 (21.9- 41.4) | 42.9 ( 0.0- 80.1) | 50.0 (32.4- 66.7) |
| PSYC | ANXI | NE | 25.8 (NE-NE) | NE | 39.1 (27.7-51.4) | 4.9 ( 0.0-11.1) | 22.5 (13.5-32.4) |
| ANXI | DELU | NE | 20.8 (NE-NE) | NE | 32.2 (20.4- 44.2) | 50.0 ( 0.0-100.0) | 44.0 (24.1- 64.3) |
| DELU | ANXI | NE | 18.6 (NE-NE) | NE | 27.5 (17.5-39.1) | 4.9 ( 0.0-11.1) | 15.5 ( 8.0-24.3) |
| ANXI | HALL | NE | 28.7 (NE-NE) | NE | 38.8 (25.0- 51.9) | NE | 58.3 (28.6- 87.5) |

| | | | | | | | |
|---|---|---|---|---|---|---|---|
| HALL | ANXI | NE | 18.8 (NE-NE) | NE | 27.5 (17.6-38.6) | NE | 9.9 ( 3.5-17.1) |
| ANXI | ELAT | 50.0 (NE-NE) | NE | 50.0 ( 0.0-100.0) | NE | 30.0 ( 0.0- 60.0) | 30.0 ( 0.0- 60.0) |
| ELAT | ANXI | 10.3 (NE-NE) | NE | 5.6 ( 0.0-20.0) | NE | 4.9 ( 0.0-10.5) | 4.2 ( 0.0- 9.4) |
| APAT | SLEE | NA | NA | NA | NA | 17.0 ( 10.2- 24.5) | 49.1 (36.0- 62.3) |
| SLEE | APAT | NA | NA | NA | NA | 30.0 (18.9-41.7) | 28.0 (19.2-36.8) |
| APAT | AGIT | 22.8 (NE-NE) | 41.6 (NE-NE) | 25.0 ( 0.0- 54.5) | 31.8 (22.5- 42.7) | 31.1 ( 20.9- 41.9) | 51.7 (39.3- 65.4) |
| AGIT | APAT | 17.0 (NE-NE) | 50.2 (NE-NE) | 23.1 ( 0.0-50.0) | 38.9 (27.8-50.7) | 38.3 (26.2-50.8) | 30.0 (21.6-39.0) |
| APAT | APPE | NA | NA | NA | NA | 33.3 ( 18.2- 48.5) | 54.5 (42.9- 65.6) |
| APPE | APAT | NA | NA | NA | NA | 21.7 (11.5-32.2) | 36.0 (27.2-45.6) |
| APAT | DISI | 43.9 (NE-NE) | 38.7 (NE-NE) | 40.0 ( 0.0-100.0) | 43.2 (27.0- 58.6) | 34.7 ( 22.0- 49.0) | 64.1 (48.8- 79.1) |
| DISI | APAT | 16.1 (NE-NE) | 23.4 (NE-NE) | 15.4 ( 0.0-38.9) | 22.2 (12.7-33.3) | 28.3 (17.3-40.7) | 25.0 (17.0-33.3) |
| APAT | ABER | NE | 49.8 (NE-NE) | NE | 39.1 (25.0- 53.6) | 50.0 ( 21.4- 80.0) | 68.4 (53.1- 83.9) |
| ABER | APAT | NE | 29.9 (NE-NE) | NE | 25.0 (15.2-36.1) | 10.0 ( 3.4-18.0) | 26.0 (17.5-34.8) |
| APAT | PSYC | NE | 39.8 (NE-NE) | NE | 33.3 (23.4- 43.4) | 71.4 ( 33.3-100.0) | 46.9 (28.1- 63.7) |
| PSYC | APAT | NE | 36.6 (NE-NE) | NE | 38.9 (27.6-50.9) | 8.3 ( 1.8-15.6) | 15.0 ( 8.3-22.8) |
| APAT | DELU | NE | 42.1 (NE-NE) | NE | 37.3 (25.0- 50.7) | 66.7 ( 16.7-100.0) | 44.0 (22.7- 63.6) |
| DELU | APAT | NE | 31.2 (NE-NE) | NE | 30.6 (20.3-41.5) | 6.7 ( 1.5-13.3) | 11.0 ( 5.2-17.8) |
| APAT | HALL | NE | 21.3 (NE-NE) | NE | 32.7 (19.6- 46.2) | NE | 66.7 (36.4- 90.9) |
| HALL | APAT | NE | 11.5 (NE-NE) | NE | 22.2 (12.5-32.3) | 1.7 ( 0.0- 5.5) | 8.0 ( 3.1-14.0) |
| APAT | ELAT | NE | 4.7 (NE-NE) | NE | 16.7 ( 0.0- 50.0) | 50.0 ( 16.7- 83.3) | 30.0 ( 0.0- 61.1) |
| ELAT | APAT | 26.7 (NE-NE) | 0.3 (NE-NE) | 15.4 ( 0.0-38.5) | 1.4 ( 0.0- 4.5) | 8.3 ( 1.8-16.0) | 3.0 ( 0.0- 6.7) |
| SLEE | AGIT | NA | NA | NA | NA | 31.1 ( 21.0- 41.8) | 25.9 (14.9- 37.3) |
| AGIT | SLEE | NA | NA | NA | NA | 21.7 (14.4-30.0) | 26.3 (15.1-37.9) |
| SLEE | APPE | NA | NA | NA | NA | 33.3 ( 18.2- 50.0) | 30.3 (19.6- 41.7) |
| APPE | SLEE | NA | NA | NA | NA | 12.3 ( 6.1-18.6) | 35.1 (22.6-48.3) |
| SLEE | DISI | NA | NA | NA | NA | 28.6 ( 16.2- 41.4) | 35.9 (21.0- 51.2) |
| DISI | SLEE | NA | NA | NA | NA | 13.2 ( 6.9-20.0) | 24.6 (14.3-35.5) |
| SLEE | ABER | NA | NA | NA | NA | 25.0 ( 0.0- 58.3) | 28.9 (15.1- 43.6) |
| ABER | SLEE | NA | NA | NA | NA | 2.8 ( 0.0- 6.2) | 19.3 ( 9.3-29.7) |
| SLEE | PSYC | NA | NA | NA | NA | 42.9 ( 0.0- 83.3) | 25.0 (10.7- 40.0) |
| PSYC | SLEE | NA | NA | NA | NA | 2.8 ( 0.0- 6.5) | 14.0 ( 5.9-23.4) |
| SLEE | DELU | NA | NA | NA | NA | 33.3 ( 0.0- 75.4) | 24.0 ( 8.7- 41.7) |
| DELU | SLEE | NA | NA | NA | NA | 1.9 ( 0.0- 5.0) | 10.5 ( 3.6-19.2) |
| SLEE | HALL | NA | NA | NA | NA | NE | 25.0 ( 0.0- 54.5) |
| HALL | SLEE | NA | NA | NA | NA | 0.9 ( 0.0- 3.1) | 5.3 ( 0.0-11.5) |

| | | | | | | | |
|---|---|---|---|---|---|---|---|
| SLEE | ELAT | NA | NA | NA | NA | 20.0 ( 0.0- 50.0) | 40.0 ( 7.1- 72.7) |
| ELAT | SLEE | NA | NA | NA | NA | 1.9 ( 0.0- 5.0) | 7.0 ( 1.4-14.8) |
| AGIT | APPE | NA | NA | NA | NA | 33.3 ( 18.2- 48.4) | 34.8 (23.2- 46.4) |
| APPE | AGIT | NA | NA | NA | NA | 17.6 ( 9.3-26.2) | 39.7 (27.3-52.4) |
| AGIT | DISI | 26.5 (NE-NE) | 58.8 (NE-NE) | 20.0 ( 0.0- 66.7) | 62.2 (45.2- 77.6) | 49.0 ( 34.0- 63.4) | 51.3 (35.1- 66.7) |
| DISI | AGIT | 13.1 (NE-NE) | 29.4 (NE-NE) | 8.3 ( 0.0-28.6) | 26.1 (17.7-35.1) | 32.4 (21.1-43.3) | 34.5 (22.7-46.9) |
| AGIT | ABER | NE | 57.8 (NE-NE) | NE | 54.3 (38.6- 68.0) | 50.0 ( 21.4- 81.9) | 39.5 (25.0- 55.9) |
| ABER | AGIT | NE | 28.7 (NE-NE) | NE | 28.4 (19.2-37.2) | 8.1 ( 2.7-14.6) | 25.9 (14.3-38.5) |
| AGIT | PSYC | NE | 52.5 (NE-NE) | NE | 45.2 (34.7- 55.7) | 71.4 ( 33.3-100.0) | 37.5 (21.2- 55.6) |
| PSYC | AGIT | NE | 39.9 (NE-NE) | NE | 43.2 (33.3-53.9) | 6.8 ( 1.4-13.2) | 20.7 (10.8-32.1) |
| AGIT | DELU | NE | 58.3 (NE-NE) | NE | 50.8 (37.9- 63.5) | 83.3 ( 42.6-100.0) | 40.0 (21.4- 60.0) |
| DELU | AGIT | NE | 35.7 (NE-NE) | NE | 34.1 (23.8-44.6) | 6.8 ( 1.4-13.2) | 17.2 ( 8.2-27.6) |
| AGIT | HALL | NE | 38.0 (NE-NE) | NE | 42.9 (28.3- 56.7) | NE | 33.3 ( 7.7- 63.6) |
| HALL | AGIT | NE | 17.0 (NE-NE) | NE | 23.9 (15.1-32.4) | NE | 6.9 ( 1.5-14.0) |
| AGIT | ELAT | NE | 24.4 (NE-NE) | NE | 50.0 ( 0.0-100.0) | 40.0 ( 0.0- 71.4) | 60.0 (28.6- 88.9) |
| ELAT | AGIT | NE | 1.2 (NE-NE) | NE | 3.4 ( 0.0- 7.3) | 5.4 ( 0.0-11.1) | 10.3 ( 3.5-18.2) |
| APPE | DISI | NA | NA | NA | NA | 16.3 ( 6.7- 27.5) | 35.9 (21.9- 51.1) |
| DISI | APPE | NA | NA | NA | NA | 20.5 ( 8.3-34.2) | 21.2 (12.0-31.5) |
| APPE | ABER | NA | NA | NA | NA | 16.7 ( 0.0- 42.9) | 36.8 (22.0- 52.4) |
| ABER | APPE | NA | NA | NA | NA | 5.1 ( 0.0-12.2) | 21.2 (11.9-31.3) |
| APPE | PSYC | NA | NA | NA | NA | 28.6 ( 0.0- 70.1) | 37.5 (20.8- 53.9) |
| PSYC | APPE | NA | NA | NA | NA | 5.1 ( 0.0-12.5) | 18.2 ( 9.7-28.4) |
| APPE | DELU | NA | NA | NA | NA | 33.3 ( 0.0- 83.3) | 36.0 (17.6- 55.0) |
| DELU | APPE | NA | NA | NA | NA | 5.1 ( 0.0-12.5) | 13.6 ( 5.9-23.3) |
| APPE | HALL | NA | NA | NA | NA | NE | 41.7 (13.3- 70.6) |
| HALL | APPE | NA | NA | NA | NA | NE | 7.6 ( 1.6-14.3) |
| APPE | ELAT | NA | NA | NA | NA | 30.0 ( 0.0- 61.6) | 40.0 (11.1- 71.4) |
| ELAT | APPE | NA | NA | NA | NA | 7.7 ( 0.0-17.1) | 6.1 ( 1.4-12.2) |
| DISI | ABER | NE | 30.2 (NE-NE) | NE | 26.1 (13.2- 38.2) | 41.7 ( 12.5- 71.4) | 47.4 (31.4- 62.8) |
| ABER | DISI | NE | 30.1 (NE-NE) | NE | 32.4 (17.1-48.7) | 10.2 ( 2.3-19.1) | 46.2 (30.6-61.8) |
| DISI | PSYC | NE | 43.4 (NE-NE) | NE | 22.6 (13.7- 31.7) | 42.9 ( 0.0- 83.3) | 21.9 ( 8.6- 37.0) |
| PSYC | DISI | NE | 66.1 (NE-NE) | NE | 51.4 (34.5-66.7) | 6.1 ( 0.0-14.3) | 17.9 ( 7.3-30.8) |
| DISI | DELU | NE | 50.0 (NE-NE) | NE | 27.1 (15.9- 38.8) | 50.0 ( 0.0-100.0) | 24.0 ( 8.0- 41.7) |
| DELU | DISI | NE | 61.5 (NE-NE) | NE | 43.2 (27.3-58.8) | 6.1 ( 0.0-14.3) | 15.4 ( 5.1-27.0) |
| DISI | HALL | NE | 44.3 (NE-NE) | NE | 22.4 (10.8- 34.6) | NE | 16.7 ( 0.0- 40.0) |

| | | | | | | | |
|---|---|---|---|---|---|---|---|
| HALL | DISI | NE | 39.7 (NE-NE) | NE | 29.7 (15.4-45.2) | NE | 5.1 ( 0.0-12.9) |
| DISI | ELAT | NE | NE | NE | NE | 40.0 ( 9.1- 71.4) | 80.0 (50.0-100.0) |
| ELAT | DISI | NE | NE | NE | NE | 8.2 ( 1.9-16.7) | 20.5 ( 8.8-34.2) |
| ABER | PSYC | NE | 40.9 (NE-NE) | NE | 32.1 (21.8- 43.2) | 14.3 ( 0.0- 50.0) | 25.0 (10.0- 40.6) |
| PSYC | ABER | NE | 62.5 (NE-NE) | NE | 58.7 (43.7-72.9) | 8.3 ( 0.0-28.6) | 21.1 ( 8.3-35.5) |
| ABER | DELU | NE | 43.1 (NE-NE) | NE | 32.2 (20.8- 44.4) | NE | 28.0 ( 9.5- 45.8) |
| DELU | ABER | NE | 53.1 (NE-NE) | NE | 41.3 (26.2-55.2) | NE | 18.4 ( 6.2-32.4) |
| ABER | HALL | NE | 29.9 (NE-NE) | NE | 36.7 (23.9- 51.3) | NE | 25.0 ( 0.0- 53.3) |
| HALL | ABER | NE | 26.9 (NE-NE) | NE | 39.1 (24.4-53.7) | 8.3 ( 0.0-28.6) | 7.9 ( 0.0-17.2) |
| ABER | ELAT | NE | 83.1 (NE-NE) | NE | 66.7 (24.8-100.0) | 20.0 ( 0.0- 50.0) | 20.0 ( 0.0- 50.0) |
| ELAT | ABER | NE | 8.4 (NE-NE) | NE | 8.7 ( 1.9-18.2) | 16.7 ( 0.0-41.7) | 5.3 ( 0.0-13.2) |
| PSYC | ELAT | NE | 87.6 (NE-NE) | NE | 83.3 (44.3-100.0) | 10.0 ( 0.0- 33.3) | 30.0 ( 0.0- 60.0) |
| ELAT | PSYC | NE | 5.8 (NE-NE) | NE | 6.0 ( 1.3-11.6) | 14.3 ( 0.0-50.0) | 9.4 ( 0.0-20.0) |
| DELU | HALL | NE | 67.0 (NE-NE) | NE | 49.0 (35.4- 63.3) | NE | 41.7 (12.5- 71.4) |
| HALL | DELU | NE | 48.9 (NE-NE) | NE | 40.7 (28.4-53.2) | NE | 20.0 ( 5.3-37.9) |
| DELU | ELAT | NE | 62.7 (NE-NE) | NE | 33.3 ( 0.0- 80.0) | 10.0 ( 0.0- 33.3) | 20.0 ( 0.0- 45.5) |
| ELAT | DELU | NE | 5.2 (NE-NE) | NE | 3.4 ( 0.0- 8.7) | 16.7 ( 0.0-57.1) | 8.0 ( 0.0-19.2) |
| HALL | ELAT | NE | 87.6 (NE-NE) | NE | 83.3 (44.3-100.0) | NE | 20.0 ( 0.0- 50.0) |
| ELAT | HALL | NE | 9.9 (NE-NE) | NE | 10.2 ( 2.4-19.2) | NE | 16.7 ( 0.0-41.7) |

S1|S2, conditional prevalence of symptom S1 given presence of symptom S2.
Displayed are conditional prevalence (%) and 95% confidence intervals. Survey estimates for ADAMS are based on survey population weights, confidence intervals are not estimated.
NA, Not applicable; NE, Not estimable.
ABER, Aberrant motor behavior; AGIT, Agitation/aggression; ANXI, Anxiety; APAT, Apathy/indifference; APPE, Appetite/eating changes; DELU, Delusions; DEPR, Depression/dysphoria; DISI, Disinhibition; ELAT, Elation/euphoria; HALU, Hallucinations; IRRI, Irritability/lability; PSYC, Psychotic symptoms (hallucinations and/or delusions); SLEE, Nighttime behavioral disturbances.

Supplemental Table 25. Conditional prevalence S1|S2 of neuropsychiatric symptoms with the NPI severity score > 1 per MMSE stratum (ADAMS and ADNI studies only)

| S1 | S2 | ADAMS survey | | ADAMS | | ADNI | |
|---|---|---|---|---|---|---|---|
| | | MMSE >26 | MMSE ≤26 | MMSE >26 | MMSE ≤26 | MMSE >26 | MMSE ≤26 |
| DEPR | IRRI | NE | 60.2 (NE-NE) | NE | 35.5 (18.8- 52.0) | 19.4 ( 8.1- 32.5) | 37.9 ( 21.7- 55.9) |
| IRRI | DEPR | NE | 34.3 (NE-NE) | NE | 28.9 (15.0-43.8) | 30.4 (13.3-50.0) | 52.4 (31.2-75.0) |
| DEPR | ANXI | 22.2 (NE-NE) | 51.6 (NE-NE) | 28.6 ( 0.0- 66.7) | 43.3 (25.0- 61.8) | 25.0 ( 6.2- 45.5) | 41.2 ( 16.7- 66.7) |
| ANXI | DEPR | 53.5 (NE-NE) | 19.2 (NE-NE) | 40.0 (0.0-100.0) | 34.2 (18.9-50.0) | 21.7 ( 5.3-40.9) | 33.3 (11.1-54.6) |
| DEPR | APAT | 11.5 (NE-NE) | 54.0 (NE-NE) | 12.5 ( 0.0- 40.0) | 40.0 (26.8- 53.7) | 20.0 ( 4.8- 38.5) | 18.4 ( 5.7- 31.4) |
| APAT | DEPR | 28.1 (NE-NE) | 48.0 (NE-NE) | 20.0 (0.0- 66.7) | 52.6 (36.4-69.5) | 17.4 ( 4.3-34.6) | 33.3 (11.8-54.5) |
| DEPR | SLEE | NA | NA | NA | NA | 19.4 ( 6.4- 33.3) | 20.0 ( 5.0- 36.0) |
| SLEE | DEPR | NA | NA | NA | NA | 26.1 ( 9.1-45.8) | 23.8 ( 6.7-42.9) |
| DEPR | AGIT | 44.3 (NE-NE) | 51.3 (NE-NE) | 66.7 ( 0.0-100.0) | 23.8 (12.1- 37.5) | 16.7 ( 0.0- 35.7) | 18.8 ( 0.0- 40.0) |
| AGIT | DEPR | 31.4 (NE-NE) | 33.3 (NE-NE) | 40.0 (0.0-100.0) | 26.3 (13.1-42.4) | 13.0 ( 0.0-28.0) | 14.3 ( 0.0-33.3) |
| DEPR | APPE | NA | NA | NA | NA | 21.1 ( 5.0- 42.9) | 13.5 ( 2.9- 25.5) |
| APPE | DEPR | NA | NA | NA | NA | 17.4 ( 4.3-35.0) | 23.8 ( 5.3-45.0) |
| DEPR | DISI | NE | 79.9 (NE-NE) | NE | 35.7 ( 9.1- 63.6) | 12.5 ( 0.0- 30.8) | 50.0 ( 14.3- 85.7) |
| DISI | DEPR | NE | 27.4 (NE-NE) | NE | 13.2 ( 2.9-25.0) | 8.7 ( 0.0-22.2) | 19.0 ( 4.8-40.0) |
| DEPR | ABER | NE | 53.6 (NE-NE) | NE | 14.3 ( 3.2- 29.0) | 14.3 ( 0.0- 50.0) | 10.0 ( 0.0- 33.3) |
| ABER | DEPR | NE | 24.2 (NE-NE) | NE | 10.5 ( 2.6-21.4) | 4.3 ( 0.0-15.0) | 4.8 ( 0.0-17.4) |
| DEPR | PSYC | NE | 53.3 (NE-NE) | NE | 30.3 (15.0- 48.2) | 50.0 ( 0.0-100.0) | 20.0 ( 0.0- 50.0) |
| PSYC | DEPR | NE | 30.2 (NE-NE) | NE | 26.3 (12.8-41.4) | 13.0 ( 0.0-28.6) | 9.5 ( 0.0-24.2) |
| DEPR | DELU | NE | 53.0 (NE-NE) | NE | 30.0 (14.3- 48.1) | 60.0 ( 0.0-100.0) | 20.0 ( 0.0- 50.0) |
| DELU | DEPR | NE | 28.5 (NE-NE) | NE | 23.7 (11.1-37.8) | 13.0 ( 0.0-28.6) | 9.5 ( 0.0-24.2) |
| DEPR | HALL | NE | 35.5 (NE-NE) | NE | 33.3 ( 0.0- 61.6) | NE | NE |
| HALL | DEPR | NE | 3.7 (NE-NE) | NE | 10.5 ( 0.0-21.1) | NE | NE |
| DEPR | ELAT | NE | NE | NE | NE | NE | 25.0 ( 0.0-100.0) |
| ELAT | DEPR | NE | NE | NE | NE | NE | 4.8 ( 0.0-15.8) |
| IRRI | ANXI | NE | 22.5 (NE-NE) | NE | 33.3 (16.0- 50.0) | 40.0 ( 18.8- 61.1) | 35.3 ( 12.5- 58.3) |
| ANXI | IRRI | NE | 14.7 (NE-NE) | NE | 32.3 (14.3-50.0) | 22.2 ( 9.7-36.6) | 20.7 ( 6.7-36.7) |
| IRRI | APAT | NE | 30.6 (NE-NE) | NE | 22.0 (10.3- 34.3) | 40.0 ( 20.0- 62.5) | 18.4 ( 7.1- 32.3) |
| APAT | IRRI | NE | 47.8 (NE-NE) | NE | 35.5 (18.2-53.1) | 22.2 ( 9.7-37.1) | 24.1 ( 9.7-41.4) |
| IRRI | SLEE | NA | NA | NA | NA | 19.4 ( 6.2- 34.6) | 20.0 ( 5.3- 36.8) |

| | | | | | | | |
|---|---|---|---|---|---|---|---|
| SLEE | IRRI | NA | NA | NA | NA | 16.7 ( 5.4-30.0) | 17.2 ( 4.5-31.8) |
| IRRI | AGIT | NE | 75.8 (NE-NE) | NE | 57.1 (41.2- 71.4) | 55.6 ( 33.3- 80.0) | 31.2 ( 9.1- 55.6) |
| AGIT | IRRI | NE | 86.3 (NE-NE) | NE | 77.4 (60.7-91.7) | 27.8 (14.3-43.2) | 17.2 ( 4.2-32.3) |
| IRRI | APPE | NA | NA | NA | NA | 15.8 ( 0.0- 34.8) | 21.6 ( 8.1- 35.9) |
| APPE | IRRI | NA | NA | NA | NA | 8.3 ( 0.0-18.2) | 27.6 (12.5-45.5) |
| IRRI | DISI | NE | 58.8 (NE-NE) | NE | 71.4 (44.4- 92.9) | 37.5 ( 13.3- 64.3) | 75.0 ( 37.5-100.0) |
| DISI | IRRI | NE | 35.5 (NE-NE) | NE | 32.3 (17.1-50.0) | 16.7 ( 5.4-30.0) | 20.7 ( 7.7-37.9) |
| IRRI | ABER | NE | 34.2 (NE-NE) | NE | 21.4 ( 7.4- 37.5) | 14.3 ( 0.0- 50.0) | 30.0 ( 0.0- 60.0) |
| ABER | IRRI | NE | 27.2 (NE-NE) | NE | 19.4 ( 6.5-34.6) | 2.8 ( 0.0- 9.1) | 10.3 ( 0.0-23.5) |
| IRRI | PSYC | NE | 39.7 (NE-NE) | NE | 39.4 (21.4- 57.7) | 66.7 ( 20.0-100.0) | 40.0 ( 11.1- 71.4) |
| PSYC | IRRI | NE | 39.5 (NE-NE) | NE | 41.9 (24.1-61.5) | 11.1 ( 2.1-24.1) | 13.8 ( 3.2-27.8) |
| IRRI | DELU | NE | 37.7 (NE-NE) | NE | 36.7 (19.2- 55.0) | 60.0 ( 0.0-100.0) | 40.0 ( 11.1- 71.4) |
| DELU | IRRI | NE | 35.5 (NE-NE) | NE | 35.5 (18.5-54.5) | 8.3 ( 0.0-19.0) | 13.8 ( 3.2-27.8) |
| IRRI | HALL | NE | 42.1 (NE-NE) | NE | 50.0 (16.7- 77.8) | NE | NE |
| HALL | IRRI | NE | 7.8 (NE-NE) | NE | 19.4 ( 5.1-35.7) | 2.8 ( 0.0- 9.1) | NE |
| IRRI | ELAT | NE | NE | NE | NE | NE | 50.0 ( 0.0-100.0) |
| ELAT | IRRI | NE | NE | NE | NE | NE | 6.9 ( 0.0-16.1) |
| ANXI | APAT | 1.7 (NE-NE) | 24.9 (NE-NE) | 12.5 ( 0.0- 50.0) | 28.0 (15.6- 41.0) | 10.0 ( 0.0- 25.0) | 26.3 ( 12.0- 41.0) |
| APAT | ANXI | 1.8 (NE-NE) | 59.5 (NE-NE) | 14.3 (0.0- 50.0) | 46.7 (29.7-65.0) | 10.0 ( 0.0-26.3) | 58.8 (33.3-81.3) |
| ANXI | SLEE | NA | NA | NA | NA | 9.7 ( 0.0- 20.7) | 8.0 ( 0.0- 20.0) |
| SLEE | ANXI | NA | NA | NA | NA | 15.0 ( 0.0-33.3) | 11.8 ( 0.0-28.6) |
| ANXI | AGIT | NE | 13.8 (NE-NE) | NE | 23.8 (11.6- 37.1) | 22.2 ( 4.5- 42.1) | 31.2 ( 9.1- 55.6) |
| AGIT | ANXI | NE | 24.0 (NE-NE) | NE | 33.3 (16.7-52.0) | 20.0 ( 4.3-39.3) | 29.4 ( 9.1-53.3) |
| ANXI | APPE | NA | NA | NA | NA | 15.8 ( 0.0- 33.3) | 13.5 ( 2.7- 25.0) |
| APPE | ANXI | NA | NA | NA | NA | 15.0 ( 0.0-31.6) | 29.4 ( 7.1-52.2) |
| ANXI | DISI | NE | 17.1 (NE-NE) | NE | 42.9 (17.6- 69.2) | 18.8 ( 0.0- 40.0) | 12.5 ( 0.0- 42.9) |
| DISI | ANXI | NE | 15.7 (NE-NE) | NE | 20.0 ( 7.1-36.4) | 15.0 ( 0.0-32.0) | 5.9 ( 0.0-21.1) |
| ANXI | ABER | NE | 18.2 (NE-NE) | NE | 28.6 (12.5- 47.4) | 14.3 ( 0.0- 50.0) | 20.0 ( 0.0- 50.0) |
| ABER | ANXI | NE | 22.1 (NE-NE) | NE | 26.7 (11.1-44.0) | 5.0 ( 0.0-17.2) | 11.8 ( 0.0-28.6) |
| ANXI | PSYC | NE | 14.8 (NE-NE) | NE | 27.3 (12.5- 41.7) | 16.7 ( 0.0- 57.6) | 30.0 ( 0.0- 62.5) |
| PSYC | ANXI | NE | 22.5 (NE-NE) | NE | 30.0 (13.3-48.2) | 5.0 ( 0.0-16.7) | 17.6 ( 0.0-37.5) |
| ANXI | DELU | NE | 12.4 (NE-NE) | NE | 26.7 (11.1- 42.9) | 20.0 ( 0.0- 75.0) | 30.0 ( 0.0- 62.5) |
| DELU | ANXI | NE | 17.9 (NE-NE) | NE | 26.7 (10.7-43.5) | 5.0 ( 0.0-16.7) | 17.6 ( 0.0-37.5) |
| ANXI | HALL | NE | 40.1 (NE-NE) | NE | 41.7 (11.1- 70.0) | NE | 50.0 ( 0.0-100.0) |
| HALL | ANXI | NE | 11.3 (NE-NE) | NE | 16.7 ( 3.7-30.6) | NE | 5.9 ( 0.0-20.0) |

| | | | | | | | |
|---|---|---|---|---|---|---|---|
| ANXI | ELAT | NE | NE | NE | NE | 50.0 ( 0.0-100.0) | NE |
| ELAT | ANXI | 25.1 (NE-NE) | NE | 14.3 (0.0- 50.0) | NE | 5.0 ( 0.0-17.2) | NE |
| APAT | SLEE | NA | NA | NA | NA | 12.9 ( 2.9- 28.0) | 20.0 ( 5.0- 36.8) |
| SLEE | APAT | NA | NA | NA | NA | 20.0 ( 4.8-38.5) | 13.2 ( 3.2-25.0) |
| APAT | AGIT | 39.7 (NE-NE) | 47.1 (NE-NE) | 33.3 ( 0.0-100.0) | 33.3 (19.6- 48.5) | 33.3 ( 12.5- 56.3) | 56.2 ( 30.0- 81.3) |
| AGIT | APAT | 11.5 (NE-NE) | 34.3 (NE-NE) | 12.5 (0.0- 40.0) | 28.0 (16.7-41.7) | 30.0 (10.7-52.4) | 23.7 (10.3-38.5) |
| APAT | APPE | NA | NA | NA | NA | 5.3 ( 0.0- 17.6) | 45.9 ( 28.6- 62.1) |
| APPE | APAT | NA | NA | NA | NA | 5.0 ( 0.0-15.8) | 44.7 (29.0-61.1) |
| APAT | DISI | NE | 50.9 (NE-NE) | NE | 35.7 ( 9.1- 63.7) | 25.0 ( 4.6- 47.4) | 25.0 ( 0.0- 60.0) |
| DISI | APAT | NE | 19.7 (NE-NE) | NE | 10.0 ( 2.1-19.1) | 20.0 ( 3.8-40.0) | 5.3 ( 0.0-14.6) |
| APAT | ABER | NE | 58.2 (NE-NE) | NE | 28.6 (13.0- 46.2) | 42.9 ( 0.0- 80.0) | 50.0 ( 18.1- 83.3) |
| ABER | APAT | NE | 29.6 (NE-NE) | NE | 16.0 ( 6.4-27.5) | 15.0 ( 0.0-31.2) | 13.2 ( 3.0-26.1) |
| APAT | PSYC | NE | 51.7 (NE-NE) | NE | 30.3 (16.1- 48.0) | 33.3 ( 0.0- 75.0) | 50.0 ( 16.7- 81.9) |
| PSYC | APAT | NE | 32.9 (NE-NE) | NE | 20.0 (10.0-31.4) | 10.0 ( 0.0-25.0) | 13.2 ( 3.1-25.0) |
| APAT | DELU | NE | 51.4 (NE-NE) | NE | 30.0 (14.8- 47.6) | 20.0 ( 0.0- 66.7) | 50.0 ( 16.7- 81.9) |
| DELU | APAT | NE | 31.0 (NE-NE) | NE | 18.0 ( 8.3-28.6) | 5.0 ( 0.0-16.7) | 13.2 ( 3.1-25.0) |
| APAT | HALL | NE | 32.0 (NE-NE) | NE | 25.0 ( 0.0- 52.9) | NE | NE |
| HALL | APAT | NE | 3.8 (NE-NE) | NE | 6.0 ( 0.0-14.0) | 5.0 ( 0.0-16.7) | 5.3 ( 0.0-13.5) |
| APAT | ELAT | NE | NE | NE | NE | 50.0 ( 0.0-100.0) | NE |
| ELAT | APAT | NE | NE | NE | NE | 5.0 ( 0.0-16.7) | NE |
| SLEE | AGIT | NA | NA | NA | NA | 5.6 ( 0.0- 16.7) | 6.2 ( 0.0- 21.4) |
| AGIT | SLEE | NA | NA | NA | NA | 3.2 ( 0.0-10.3) | 4.0 ( 0.0-13.3) |
| SLEE | APPE | NA | NA | NA | NA | 10.5 ( 0.0- 25.0) | 13.5 ( 3.1- 27.0) |
| APPE | SLEE | NA | NA | NA | NA | 6.5 ( 0.0-15.6) | 20.0 ( 5.0-36.4) |
| SLEE | DISI | NA | NA | NA | NA | 18.8 ( 0.0- 40.0) | 25.0 ( 0.0- 60.0) |
| DISI | SLEE | NA | NA | NA | NA | 9.7 ( 0.0-20.7) | 8.0 ( 0.0-20.8) |
| SLEE | ABER | NA | NA | NA | NA | 28.6 ( 0.0- 75.0) | 30.0 ( 0.0- 60.1) |
| ABER | SLEE | NA | NA | NA | NA | 6.5 ( 0.0-16.7) | 12.0 ( 0.0-25.0) |
| SLEE | PSYC | NA | NA | NA | NA | 33.3 ( 0.0- 83.3) | NE |
| PSYC | SLEE | NA | NA | NA | NA | 6.5 ( 0.0-16.1) | NE |
| SLEE | DELU | NA | NA | NA | NA | 20.0 ( 0.0- 75.0) | NE |
| DELU | SLEE | NA | NA | NA | NA | 3.2 ( 0.0-10.7) | NE |
| SLEE | HALL | NA | NA | NA | NA | NE | NE |
| HALL | SLEE | NA | NA | NA | NA | 3.2 ( 0.0-11.1) | NE |
| SLEE | ELAT | NA | NA | NA | NA | NE | 25.0 ( 0.0-100.0) |

| | | | | | | | |
|---|---|---|---|---|---|---|---|
| ELAT | SLEE | NA | NA | NA | NA | NE | 4.0 ( 0.0-13.3) |
| AGIT | APPE | NA | NA | NA | NA | 10.5 ( 0.0- 27.3) | 13.5 ( 3.1- 25.7) |
| APPE | AGIT | NA | NA | NA | NA | 11.1 ( 0.0-27.3) | 31.2 ( 9.1-54.5) |
| AGIT | DISI | NE | 53.1 (NE-NE) | NE | 64.3 (36.4- 90.0) | 43.8 ( 20.0- 69.2) | 12.5 ( 0.0- 42.9) |
| DISI | AGIT | NE | 28.1 (NE-NE) | NE | 21.4 (10.0-34.0) | 38.9 (14.3-63.2) | 6.2 ( 0.0-21.4) |
| AGIT | ABER | NE | 43.7 (NE-NE) | NE | 39.3 (21.7- 59.3) | NE | 20.0 ( 0.0- 50.0) |
| ABER | AGIT | NE | 30.5 (NE-NE) | NE | 26.2 (13.9-40.0) | NE | 12.5 ( 0.0-31.3) |
| AGIT | PSYC | NE | 37.7 (NE-NE) | NE | 42.4 (24.3- 61.9) | 50.0 ( 0.0-100.0) | 30.0 ( 0.0- 60.0) |
| PSYC | AGIT | NE | 32.9 (NE-NE) | NE | 33.3 (19.5-47.7) | 16.7 ( 0.0-37.5) | 18.8 ( 0.0-40.0) |
| AGIT | DELU | NE | 38.7 (NE-NE) | NE | 43.3 (25.0- 62.5) | 60.0 ( 0.0-100.0) | 30.0 ( 0.0- 60.0) |
| DELU | AGIT | NE | 32.0 (NE-NE) | NE | 31.0 (18.2-44.7) | 16.7 ( 0.0-37.5) | 18.8 ( 0.0-40.0) |
| AGIT | HALL | NE | 29.6 (NE-NE) | NE | 41.7 (12.5- 72.7) | NE | NE |
| HALL | AGIT | NE | 4.8 (NE-NE) | NE | 11.9 ( 2.6-22.9) | NE | NE |
| AGIT | ELAT | NE | NE | NE | NE | 50.0 ( 0.0-100.0) | 25.0 ( 0.0- 75.0) |
| ELAT | AGIT | NE | NE | NE | NE | 5.6 ( 0.0-18.8) | 6.2 ( 0.0-21.1) |
| APPE | DISI | NA | NA | NA | NA | 31.2 ( 9.1- 57.1) | 12.5 ( 0.0- 40.0) |
| DISI | APPE | NA | NA | NA | NA | 26.3 ( 6.7-47.4) | 2.7 ( 0.0- 9.3) |
| APPE | ABER | NA | NA | NA | NA | 28.6 ( 0.0- 66.7) | 30.0 ( 0.0- 62.5) |
| ABER | APPE | NA | NA | NA | NA | 10.5 ( 0.0-26.3) | 8.1 ( 0.0-18.4) |
| APPE | PSYC | NA | NA | NA | NA | 16.7 ( 0.0- 50.0) | 30.0 ( 0.0- 61.6) |
| PSYC | APPE | NA | NA | NA | NA | 5.3 ( 0.0-17.4) | 8.1 ( 0.0-18.8) |
| APPE | DELU | NA | NA | NA | NA | 20.0 ( 0.0- 66.7) | 30.0 ( 0.0- 61.6) |
| DELU | APPE | NA | NA | NA | NA | 5.3 ( 0.0-17.4) | 8.1 ( 0.0-18.8) |
| APPE | HALL | NA | NA | NA | NA | NE | 50.0 ( 0.0-100.0) |
| HALL | APPE | NA | NA | NA | NA | NE | 2.7 ( 0.0- 9.3) |
| APPE | ELAT | NA | NA | NA | NA | 50.0 ( 0.0-100.0) | 25.0 ( 0.0- 75.0) |
| ELAT | APPE | NA | NA | NA | NA | 5.3 ( 0.0-17.7) | 2.7 ( 0.0- 9.4) |
| DISI | ABER | NE | 37.0 (NE-NE) | NE | 25.0 ( 9.5- 41.7) | 14.3 ( 0.0- 50.0) | 20.0 ( 0.0- 50.0) |
| ABER | DISI | NE | 48.7 (NE-NE) | NE | 50.0 (22.2-78.6) | 6.2 ( 0.0-20.0) | 25.0 ( 0.0-60.0) |
| DISI | PSYC | NE | 32.3 (NE-NE) | NE | 21.2 ( 8.3- 37.0) | 33.3 ( 0.0- 75.0) | 20.0 ( 0.0- 50.0) |
| PSYC | DISI | NE | 53.2 (NE-NE) | NE | 50.0 (23.1-77.8) | 12.5 ( 0.0-31.6) | 25.0 ( 0.0-60.1) |
| DISI | DELU | NE | 30.9 (NE-NE) | NE | 20.0 ( 6.5- 36.8) | 40.0 ( 0.0-100.0) | 20.0 ( 0.0- 50.0) |
| DELU | DISI | NE | 48.2 (NE-NE) | NE | 42.9 (17.6-71.4) | 12.5 ( 0.0-31.6) | 25.0 ( 0.0-60.1) |
| DISI | HALL | NE | 27.4 (NE-NE) | NE | 25.0 ( 0.0- 50.0) | NE | NE |
| HALL | DISI | NE | 8.4 (NE-NE) | NE | 21.4 ( 0.0-45.5) | NE | NE |

| | | | | | | | |
|---|---|---|---|---|---|---|---|
| DISI | ELAT | NE | NE | NE | NE | NE | 50.0 ( 0.0-100.0) |
| ELAT | DISI | NE | NE | NE | NE | 12.5 ( 0.0-31.2) | 25.0 ( 0.0-57.1) |
| ABER | PSYC | NE | 48.9 (NE-NE) | NE | 30.3 (14.3- 47.5) | 16.7 ( 0.0- 50.0) | 20.0 ( 0.0- 50.0) |
| PSYC | ABER | NE | 61.2 (NE-NE) | NE | 35.7 (17.9-52.9) | 14.3 ( 0.0-50.0) | 20.0 ( 0.0-50.0) |
| ABER | DELU | NE | 50.3 (NE-NE) | NE | 30.0 (13.6- 47.8) | NE | 20.0 ( 0.0- 50.0) |
| DELU | ABER | NE | 59.6 (NE-NE) | NE | 32.1 (15.2-48.6) | NE | 20.0 ( 0.0-50.0) |
| ABER | HALL | NE | 17.8 (NE-NE) | NE | 25.0 ( 0.0- 53.9) | NE | 50.0 ( 0.0-100.0) |
| HALL | ABER | NE | 4.1 (NE-NE) | NE | 10.7 ( 0.0-22.2) | 14.3 ( 0.0-50.0) | 10.0 ( 0.0-33.3) |
| ABER | ELAT | NE | NE | NE | NE | 50.0 ( 0.0-100.0) | 25.0 ( 0.0-100.0) |
| ELAT | ABER | NE | NE | NE | NE | 14.3 ( 0.0-50.0) | 10.0 ( 0.0-33.3) |
| PSYC | ELAT | NE | NE | NE | NE | 50.0 ( 0.0-100.0) | 50.0 ( 0.0-100.0) |
| ELAT | PSYC | NE | NE | NE | NE | 16.7 ( 0.0-50.0) | 20.0 ( 0.0-44.4) |
| DELU | HALL | NE | 71.4 (NE-NE) | NE | 75.0 (50.0-100.0) | NE | NE |
| HALL | DELU | NE | 14.0 (NE-NE) | NE | 30.0 (13.5-48.1) | NE | 20.0 ( 0.0-50.0) |
| DELU | ELAT | NE | NE | NE | NE | 50.0 ( 0.0-100.0) | 50.0 ( 0.0-100.0) |
| ELAT | DELU | NE | NE | NE | NE | 20.0 ( 0.0-66.7) | 20.0 ( 0.0-44.4) |
| HALL | ELAT | NE | NE | NE | NE | NE | NE |
| ELAT | HALL | NE | NE | NE | NE | NE | NE |

S1|S2, conditional prevalence of symptom S1 given presence of symptom S2.
Displayed are conditional prevalence (%) and 95% confidence intervals. Survey estimates for ADAMS are based on survey population weights, confidence intervals are not estimated.
NA, Not applicable; NE, Not estimable.
ABER, Aberrant motor behavior; AGIT, Agitation/aggression; ANXI, Anxiety; APAT, Apathy/indifference; APPE, Appetite/eating changes; DELU, Delusions; DEPR, Depression/dysphoria; DISI, Disinhibition; ELAT, Elation/euphoria; HALU, Hallucinations; IRRI, Irritability/lability; PSYC, Psychotic symptoms (hallucinations and/or delusions); SLEE, Nighttime behavioral disturbances.

Supplemental Table 26. Conditional prevalence S1|S2 of neuropsychiatric symptoms with the NPI severity score > 0 per MMSE stratum (NACC study only)

| S1 | S2 | NACC | | | | |
|---|---|---|---|---|---|---|
| | | MMSE >26 | MMSE ≤26 | 19 < MMSE ≤ 26 | 9 < MMSE ≤ 19 | MMSE ≤ 9 |
| DEPR | IRRI | 49.1 (47.2-50.8) | 54.1 (52.6-55.7) | 53.0 (51.0-54.9) | 56.5 (53.9-59.4) | 54.2 (49.4-58.8) |
| IRRI | DEPR | 44.6 (42.7-46.3) | 50.6 (49.2-52.1) | 48.9 (47.0-50.7) | 52.2 (49.5-55.1) | 57.6 (52.5-62.6) |
| DEPR | ANXI | 59.8 (57.9-61.8) | 59.4 (57.9-60.9) | 59.7 (57.8-61.7) | 59.9 (57.3-62.6) | 56.3 (51.7-60.6) |
| ANXI | DEPR | 44.3 (42.7-46.0) | 56.2 (54.7-57.7) | 51.6 (49.7-53.5) | 62.5 (59.9-65.2) | 67.0 (62.2-71.6) |
| DEPR | APAT | 59.6 (57.5-61.8) | 54.1 (52.6-55.5) | 55.6 (53.7-57.4) | 52.8 (50.3-55.3) | 50.9 (46.7-55.1) |
| APAT | DEPR | 34.3 (32.6-35.9) | 55.7 (54.2-57.1) | 51.7 (49.7-53.4) | 58.2 (55.7-60.6) | 74.6 (70.4-78.8) |
| DEPR | SLEE | 46.4 (44.5-48.3) | 53.5 (51.8-55.2) | 53.9 (51.8-56.0) | 53.5 (50.1-56.7) | 51.3 (46.1-56.3) |
| SLEE | DEPR | 36.1 (34.4-37.7) | 38.4 (36.9-39.6) | 38.4 (36.6-40.1) | 36.6 (34.0-39.1) | 43.6 (39.0-48.3) |
| DEPR | AGIT | 51.2 (49.0-53.4) | 54.3 (52.6-55.9) | 54.3 (52.1-56.5) | 55.9 (53.2-58.7) | 50.3 (45.8-54.8) |
| AGIT | DEPR | 28.7 (27.2-30.3) | 44.1 (42.7-45.5) | 39.0 (37.1-40.9) | 49.7 (47.0-52.3) | 61.3 (56.6-66.0) |
| DEPR | APPE | 48.6 (46.0-51.1) | 52.2 (50.5-54.1) | 52.0 (49.8-54.3) | 54.2 (50.9-57.5) | 47.6 (42.4-53.0) |
| APPE | DEPR | 22.2 (20.8-23.6) | 35.0 (33.7-36.3) | 33.7 (32.1-35.4) | 37.1 (34.5-39.7) | 36.7 (32.1-41.2) |
| DEPR | DISI | 50.0 (46.9-52.6) | 50.9 (48.7-53.0) | 50.3 (47.6-53.2) | 54.1 (50.9-57.9) | 45.7 (40.2-50.9) |
| DISI | DEPR | 16.7 (15.4-18.0) | 27.0 (25.8-28.3) | 23.8 (22.3-25.5) | 31.0 (28.5-33.5) | 36.5 (31.8-41.1) |
| DEPR | ABER | 54.9 (51.1-58.8) | 52.0 (50.0-54.0) | 54.0 (50.8-57.1) | 52.7 (49.5-55.9) | 45.8 (40.8-50.6) |
| ABER | DEPR | 10.5 ( 9.3-11.6) | 27.3 (26.0-28.5) | 20.0 (18.5-21.4) | 36.3 (33.6-39.1) | 48.3 (43.5-53.2) |
| DEPR | PSYC | 62.0 (57.5-66.2) | 55.6 (53.6-57.6) | 57.0 (53.9-60.0) | 55.5 (52.2-58.8) | 52.5 (47.6-57.3) |
| PSYC | DEPR | 9.1 ( 8.1-10.1) | 28.5 (27.1-29.7) | 21.0 (19.5-22.5) | 36.7 (34.1-39.3) | 52.7 (48.0-57.6) |
| DEPR | DELU | 63.0 (58.2-67.6) | 57.4 (55.1-59.7) | 58.8 (55.3-62.1) | 56.7 (53.0-60.5) | 55.3 (50.0-60.7) |
| DELU | DEPR | 7.2 ( 6.4- 8.1) | 23.4 (22.1-24.7) | 17.3 (15.9-18.8) | 30.1 (27.6-32.5) | 43.3 (38.7-48.1) |
| DEPR | HALL | 62.1 (55.3-69.2) | 55.0 (52.0-58.1) | 57.6 (52.7-62.6) | 56.6 (51.7-61.3) | 48.1 (41.4-54.9) |
| HALL | DEPR | 3.2 ( 2.7- 3.8) | 12.4 (11.5-13.3) | 7.9 ( 6.9- 8.9) | 17.1 (15.1-19.1) | 28.3 (23.8-32.7) |
| DEPR | ELAT | 47.1 (41.5-52.6) | 52.5 (48.5-56.5) | 49.7 (44.3-55.0) | 53.8 (46.4-60.8) | 59.8 (49.4-70.2) |
| ELAT | DEPR | 4.6 ( 3.8- 5.3) | 7.0 ( 6.3- 7.7) | 5.6 ( 4.8- 6.4) | 8.2 ( 6.7- 9.7) | 12.8 ( 9.6-16.1) |

| | | | | | | |
|---|---|---|---|---|---|---|
| IRRI | ANXI | 47.7 (45.6-49.9) | 54.1 (52.6-55.6) | 52.5 (50.5-54.4) | 54.5 (51.8-57.2) | 60.7 (56.3-64.9) |
| ANXI | IRRI | 38.8 (37.1-40.6) | 54.6 (53.2-56.1) | 49.2 (47.4-51.2) | 61.5 (58.8-64.1) | 67.8 (63.1-72.3) |
| IRRI | APAT | 54.5 (52.3-56.6) | 49.9 (48.5-51.3) | 50.2 (48.1-52.1) | 49.5 (46.8-52.0) | 49.7 (45.9-53.6) |
| APAT | IRRI | 34.5 (32.8-36.1) | 54.9 (53.4-56.3) | 50.6 (48.7-52.5) | 59.0 (56.2-61.7) | 68.5 (63.7-72.9) |
| IRRI | SLEE | 41.6 (39.6-43.7) | 51.7 (50.0-53.5) | 50.1 (47.9-52.3) | 52.8 (49.6-56.3) | 58.3 (53.2-63.3) |
| SLEE | IRRI | 35.6 (33.9-37.4) | 39.6 (38.2-41.0) | 38.7 (36.8-40.4) | 39.1 (36.3-41.8) | 46.5 (41.7-51.3) |
| IRRI | AGIT | 70.6 (68.4-72.6) | 69.1 (67.6-70.6) | 71.5 (69.5-73.4) | 67.0 (64.3-69.6) | 64.8 (60.7-69.0) |
| AGIT | IRRI | 43.6 (41.8-45.3) | 60.1 (58.6-61.5) | 55.6 (53.7-57.6) | 64.5 (61.7-67.1) | 74.3 (70.1-78.5) |
| IRRI | APPE | 44.8 (42.3-47.4) | 50.3 (48.6-52.2) | 50.4 (48.0-52.8) | 49.9 (46.5-53.6) | 50.8 (45.3-56.1) |
| APPE | IRRI | 22.6 (21.1-24.1) | 36.0 (34.6-37.5) | 35.4 (33.6-37.3) | 37.0 (34.2-39.9) | 36.8 (32.4-41.4) |
| IRRI | DISI | 66.5 (63.7-69.3) | 64.4 (62.5-66.3) | 65.6 (63.1-68.0) | 63.0 (59.5-66.5) | 63.3 (57.9-68.7) |
| DISI | IRRI | 24.5 (23.0-25.9) | 36.6 (35.2-37.9) | 33.6 (31.9-35.4) | 39.0 (36.2-41.8) | 47.5 (42.6-52.3) |
| IRRI | ABER | 61.2 (57.2-64.7) | 57.0 (55.0-59.1) | 59.2 (56.1-62.1) | 53.7 (50.5-57.2) | 58.6 (54.1-63.3) |
| ABER | IRRI | 12.9 (11.7-14.1) | 32.0 (30.6-33.5) | 23.8 (22.1-25.4) | 40.0 (37.4-42.8) | 58.1 (53.5-62.6) |
| IRRI | PSYC | 63.2 (59.0-67.4) | 56.0 (54.0-58.1) | 57.8 (54.8-60.7) | 54.5 (51.2-57.6) | 54.7 (49.6-59.6) |
| PSYC | IRRI | 10.2 ( 9.2-11.3) | 30.7 (29.3-32.0) | 23.1 (21.5-24.7) | 39.1 (36.3-41.8) | 51.6 (46.5-56.1) |
| IRRI | DELU | 67.3 (62.7-72.0) | 59.5 (57.3-61.8) | 62.3 (59.0-65.6) | 57.4 (53.6-60.7) | 56.9 (51.4-62.4) |
| DELU | IRRI | 8.5 ( 7.6- 9.6) | 26.0 (24.6-27.3) | 19.9 (18.4-21.5) | 33.0 (30.4-35.6) | 41.9 (37.2-46.1) |
| IRRI | HALL | 54.4 (46.5-61.9) | 52.5 (49.4-55.7) | 47.9 (42.7-53.1) | 53.3 (48.5-58.2) | 58.6 (52.1-64.8) |
| HALL | IRRI | 3.1 ( 2.5- 3.8) | 12.7 (11.7-13.8) | 7.1 ( 6.2- 8.2) | 17.4 (15.1-19.6) | 32.4 (27.8-37.0) |
| IRRI | ELAT | 62.7 (57.5-68.1) | 61.4 (57.2-65.2) | 60.9 (55.3-66.1) | 61.4 (54.6-68.3) | 63.2 (53.4-72.2) |
| ELAT | IRRI | 6.7 ( 5.8- 7.6) | 8.8 ( 7.9- 9.6) | 7.5 ( 6.5- 8.5) | 10.1 ( 8.4-12.0) | 12.7 ( 9.8-16.1) |
| ANXI | APAT | 42.6 (40.4-44.8) | 50.4 (49.0-51.9) | 46.3 (44.4-48.2) | 54.3 (51.8-57.0) | 58.8 (54.9-62.9) |
| APAT | ANXI | 33.1 (31.2-35.1) | 54.9 (53.3-56.3) | 49.9 (47.9-51.9) | 57.4 (54.7-59.9) | 72.5 (68.6-76.3) |
| ANXI | SLEE | 36.7 (34.8-38.7) | 52.4 (50.8-54.1) | 47.9 (45.7-50.0) | 59.6 (56.5-62.8) | 59.7 (54.7-64.6) |
| SLEE | ANXI | 38.6 (36.7-40.5) | 39.7 (38.3-41.1) | 39.5 (37.5-41.4) | 39.1 (36.6-41.7) | 42.7 (38.0-47.1) |
| ANXI | AGIT | 39.7 (37.5-42.0) | 54.3 (52.7-56.0) | 50.5 (48.3-52.7) | 57.7 (54.7-60.4) | 61.8 (57.3-66.1) |
| AGIT | ANXI | 30.1 (28.2-31.9) | 46.7 (45.2-48.1) | 41.9 (39.9-43.9) | 49.2 (46.6-51.8) | 63.4 (59.5-67.6) |
| ANXI | APPE | 37.4 (34.9-39.9) | 50.5 (48.9-52.3) | 46.8 (44.7-49.1) | 55.2 (52.1-58.6) | 58.5 (53.2-63.8) |

| | | | | | | |
|---|---|---|---|---|---|---|
| APPE | ANXI | 23.1 (21.4-24.8) | 35.8 (34.4-37.3) | 35.1 (33.3-37.0) | 36.3 (33.7-39.0) | 37.9 (33.8-42.2) |
| ANXI | DISI | 45.4 (42.6-48.2) | 55.2 (53.1-57.3) | 51.4 (48.7-54.2) | 59.6 (56.0-63.4) | 60.5 (55.0-65.5) |
| DISI | ANXI | 20.5 (18.9-22.2) | 31.0 (29.7-32.4) | 28.1 (26.4-29.9) | 32.7 (30.0-35.4) | 40.6 (36.5-44.9) |
| ANXI | ABER | 51.2 (47.3-55.0) | 60.3 (58.3-62.4) | 58.2 (55.1-61.2) | 61.7 (58.4-64.8) | 62.6 (58.3-67.3) |
| ABER | ANXI | 13.2 (11.9-14.5) | 33.5 (32.2-34.9) | 24.9 (23.2-26.6) | 40.8 (38.2-43.3) | 55.5 (51.4-59.9) |
| ANXI | PSYC | 49.2 (44.6-53.4) | 56.9 (54.9-58.8) | 53.6 (50.5-56.7) | 58.6 (55.2-61.9) | 61.3 (56.7-65.9) |
| PSYC | ANXI | 9.7 ( 8.6-10.9) | 30.8 (29.4-32.1) | 22.9 (21.1-24.6) | 37.3 (34.7-39.8) | 51.8 (47.3-56.0) |
| ANXI | DELU | 49.3 (44.3-54.3) | 57.9 (55.5-60.2) | 54.7 (51.2-58.3) | 59.6 (55.6-63.2) | 62.3 (57.1-67.4) |
| DELU | ANXI | 7.7 ( 6.6- 8.7) | 25.0 (23.6-26.3) | 18.7 (17.0-20.2) | 30.4 (27.9-32.9) | 41.0 (36.8-45.3) |
| ANXI | HALL | 56.2 (48.6-63.7) | 60.0 (57.0-63.2) | 56.1 (50.7-60.9) | 62.8 (57.7-67.5) | 61.9 (55.8-68.7) |
| HALL | ANXI | 4.0 ( 3.2- 4.8) | 14.4 (13.2-15.4) | 8.9 ( 7.7-10.1) | 18.2 (16.1-20.2) | 30.6 (26.2-34.8) |
| ANXI | ELAT | 46.5 (41.0-51.8) | 59.1 (55.2-63.4) | 54.8 (49.4-60.7) | 64.0 (57.5-70.7) | 63.2 (52.7-73.2) |
| ELAT | ANXI | 6.1 ( 5.1- 7.2) | 8.3 ( 7.5- 9.2) | 7.2 ( 6.2- 8.2) | 9.3 ( 7.8-10.9) | 11.4 ( 8.6-14.4) |
| APAT | SLEE | 32.9 (31.2-34.5) | 57.5 (55.7-59.3) | 52.6 (50.3-54.8) | 63.6 (60.5-66.7) | 69.6 (64.8-74.1) |
| SLEE | APAT | 44.5 (42.5-46.6) | 40.0 (38.6-41.5) | 40.3 (38.5-42.2) | 39.5 (37.0-42.1) | 40.3 (36.5-44.4) |
| APAT | AGIT | 38.8 (36.6-40.9) | 57.7 (56.1-59.4) | 54.2 (52.0-56.3) | 59.9 (57.1-62.8) | 67.1 (63.0-71.2) |
| AGIT | APAT | 37.9 (35.7-40.0) | 45.6 (44.2-47.1) | 41.8 (39.9-43.7) | 48.3 (45.6-51.1) | 55.8 (52.1-60.0) |
| APAT | APPE | 42.8 (40.3-45.3) | 60.3 (58.6-62.1) | 58.0 (55.6-60.4) | 61.2 (58.2-64.2) | 71.2 (66.5-76.3) |
| APPE | APAT | 34.0 (32.0-36.1) | 39.3 (37.9-40.7) | 40.4 (38.6-42.3) | 38.0 (35.4-40.5) | 37.5 (33.8-41.3) |
| APAT | DISI | 49.5 (46.6-52.6) | 63.2 (61.4-65.2) | 61.1 (58.8-63.8) | 63.5 (60.1-66.9) | 71.0 (66.4-76.0) |
| DISI | APAT | 28.8 (26.7-30.8) | 32.7 (31.4-34.0) | 31.1 (29.4-32.8) | 33.0 (30.5-35.6) | 38.7 (35.0-42.5) |
| APAT | ABER | 57.4 (53.3-61.0) | 64.7 (62.8-66.6) | 62.1 (59.2-64.9) | 64.8 (61.6-67.9) | 70.8 (66.3-74.9) |
| ABER | APAT | 19.0 (17.2-20.9) | 33.0 (31.8-34.3) | 24.7 (23.1-26.3) | 40.5 (37.9-43.1) | 50.9 (46.9-54.9) |
| APAT | PSYC | 47.1 (42.8-51.7) | 58.9 (56.9-61.0) | 54.3 (51.5-57.2) | 58.7 (55.4-62.0) | 70.6 (66.4-74.9) |
| PSYC | APAT | 12.0 (10.6-13.6) | 29.3 (27.9-30.7) | 21.5 (19.9-23.2) | 35.3 (32.8-37.9) | 48.4 (44.1-52.1) |
| APAT | DELU | 47.5 (42.5-52.9) | 58.8 (56.5-61.2) | 54.1 (50.8-57.8) | 58.4 (54.7-62.0) | 71.7 (66.8-76.5) |
| DELU | APAT | 9.5 ( 8.2-10.9) | 23.3 (22.1-24.6) | 17.1 (15.6-18.6) | 28.2 (25.8-30.6) | 38.3 (34.4-42.0) |
| APAT | HALL | 52.1 (44.4-59.6) | 62.5 (59.5-65.5) | 58.2 (53.3-62.9) | 62.8 (58.0-67.3) | 69.0 (63.3-74.8) |
| HALL | APAT | 4.7 ( 3.8- 5.7) | 13.7 (12.7-14.7) | 8.6 ( 7.6- 9.7) | 17.2 (15.1-19.3) | 27.7 (24.0-31.2) |

| | | | | | | |
|---|---|---|---|---|---|---|
| APAT | ELAT | 53.2 (48.0-59.3) | 63.1 (59.4-67.2) | 62.5 (57.7-68.1) | 62.4 (55.4-69.0) | 66.7 (56.6-76.7) |
| ELAT | APAT | 8.9 ( 7.7-10.4) | 8.2 ( 7.4- 9.0) | 7.6 ( 6.6- 8.6) | 8.6 ( 7.2-10.0) | 9.7 ( 7.5-12.3) |
| SLEE | AGIT | 39.4 (37.2-41.7) | 41.4 (39.9-43.0) | 41.6 (39.6-43.8) | 38.8 (36.2-41.6) | 46.3 (41.3-50.8) |
| AGIT | SLEE | 28.4 (26.7-30.2) | 46.9 (45.3-48.6) | 41.9 (39.9-44.0) | 50.5 (47.2-53.7) | 66.4 (61.7-71.3) |
| SLEE | APPE | 46.1 (43.4-48.6) | 45.3 (43.4-47.0) | 43.6 (41.3-45.8) | 45.0 (41.8-48.2) | 55.3 (49.8-60.7) |
| APPE | SLEE | 27.1 (25.3-28.7) | 42.3 (40.5-44.0) | 39.7 (37.6-41.7) | 45.0 (41.8-48.2) | 50.1 (44.8-55.4) |
| SLEE | DISI | 42.5 (39.7-45.3) | 44.1 (42.2-46.0) | 43.7 (41.1-46.4) | 42.4 (38.9-46.0) | 49.7 (44.1-55.0) |
| DISI | SLEE | 18.3 (16.7-19.8) | 32.7 (31.1-34.3) | 29.0 (27.1-30.9) | 35.5 (32.2-38.7) | 46.7 (41.7-51.8) |
| SLEE | ABER | 49.6 (45.9-53.3) | 46.6 (44.8-48.5) | 47.4 (44.2-50.5) | 43.7 (40.8-47.0) | 50.7 (45.8-55.3) |
| ABER | SLEE | 12.2 (10.9-13.5) | 34.1 (32.7-35.7) | 24.6 (22.8-26.6) | 44.0 (40.8-47.6) | 62.9 (57.9-67.8) |
| SLEE | PSYC | 50.4 (46.0-54.6) | 45.8 (43.8-47.8) | 46.1 (42.9-49.0) | 43.7 (40.6-47.1) | 49.3 (44.1-54.1) |
| PSYC | SLEE | 9.5 ( 8.4-10.7) | 32.7 (31.0-34.2) | 23.8 (22.1-25.6) | 42.3 (39.4-45.4) | 58.3 (53.1-63.4) |
| SLEE | DELU | 48.0 (43.1-52.6) | 43.7 (41.4-45.9) | 42.8 (39.3-46.2) | 42.2 (38.8-45.9) | 49.1 (43.2-54.6) |
| DELU | SLEE | 7.1 ( 6.1- 8.1) | 24.8 (23.4-26.3) | 17.7 (16.1-19.3) | 32.8 (29.7-35.8) | 45.2 (40.2-50.4) |
| SLEE | HALL | 61.5 (53.7-69.1) | 57.3 (54.3-60.5) | 57.9 (52.8-62.9) | 54.6 (49.9-59.7) | 60.7 (54.5-66.8) |
| HALL | SLEE | 4.1 ( 3.4- 4.9) | 18.1 (16.7-19.4) | 11.2 ( 9.8-12.6) | 24.1 (21.2-26.7) | 42.0 (36.0-47.4) |
| SLEE | ELAT | 44.9 (39.4-50.0) | 46.6 (42.6-50.5) | 48.7 (42.9-54.4) | 39.6 (33.0-47.0) | 55.2 (44.8-65.6) |
| ELAT | SLEE | 5.6 ( 4.6- 6.5) | 8.7 ( 7.7- 9.6) | 7.7 ( 6.6- 8.9) | 8.8 ( 7.0-10.7) | 13.9 (10.6-17.6) |
| AGIT | APPE | 33.1 (30.5-35.5) | 46.3 (44.5-48.0) | 43.0 (40.7-45.3) | 49.7 (46.5-52.9) | 55.3 (49.4-60.9) |
| APPE | AGIT | 27.0 (25.0-29.0) | 38.1 (36.5-39.7) | 38.8 (36.6-40.9) | 38.3 (35.5-41.2) | 34.9 (30.8-39.4) |
| AGIT | DISI | 53.8 (50.7-56.6) | 61.0 (59.0-63.0) | 57.4 (54.7-60.0) | 64.3 (61.0-67.9) | 68.2 (62.7-73.4) |
| DISI | AGIT | 32.0 (29.8-34.2) | 39.9 (38.5-41.4) | 37.8 (35.8-39.9) | 41.4 (38.5-44.4) | 44.6 (40.4-49.1) |
| AGIT | ABER | 51.2 (47.2-55.4) | 55.8 (53.7-57.8) | 54.4 (51.3-57.8) | 54.8 (51.6-58.4) | 61.0 (56.4-65.4) |
| ABER | AGIT | 17.4 (15.7-19.1) | 36.0 (34.5-37.5) | 28.1 (26.1-30.1) | 42.5 (39.5-45.5) | 52.7 (48.4-57.3) |
| AGIT | PSYC | 54.8 (50.1-59.3) | 60.0 (58.1-61.9) | 56.8 (53.6-59.8) | 59.2 (55.9-62.4) | 69.9 (65.4-74.2) |
| PSYC | AGIT | 14.3 (12.7-16.0) | 37.8 (36.1-39.3) | 29.2 (27.1-31.1) | 44.1 (41.2-46.7) | 57.6 (53.1-61.6) |
| AGIT | DELU | 61.9 (56.9-67.2) | 63.6 (61.3-65.7) | 60.5 (56.8-63.7) | 62.8 (59.1-66.3) | 73.3 (68.3-78.4) |
| DELU | AGIT | 12.7 (11.2-14.2) | 31.9 (30.3-33.4) | 24.8 (22.9-26.8) | 37.5 (34.6-40.2) | 47.1 (42.7-51.3) |
| AGIT | HALL | 42.6 (35.0-50.0) | 57.3 (54.4-60.5) | 49.2 (44.4-54.3) | 57.7 (52.7-62.4) | 69.5 (63.8-75.1) |

| | | | | | | |
|---|---|---|---|---|---|---|
| HALL | AGIT | 4.0 ( 3.1- 4.9) | 15.9 (14.7-17.2) | 9.4 ( 8.2-10.7) | 19.6 (17.3-21.9) | 33.5 (28.8-37.6) |
| AGIT | ELAT | 55.4 (50.0-60.6) | 59.1 (55.0-63.1) | 57.7 (52.0-63.0) | 60.4 (53.9-67.4) | 60.9 (50.0-70.8) |
| ELAT | AGIT | 9.6 ( 8.2-10.9) | 9.7 ( 8.7-10.7) | 9.1 ( 7.8-10.4) | 10.3 ( 8.5-12.1) | 10.7 ( 8.1-13.5) |
| APPE | DISI | 35.4 (32.7-38.2) | 44.3 (42.3-46.3) | 44.9 (42.2-47.6) | 44.8 (41.2-48.4) | 40.7 (35.5-45.7) |
| DISI | APPE | 25.9 (23.7-28.1) | 35.1 (33.5-36.8) | 32.8 (30.6-34.8) | 37.5 (34.1-40.6) | 42.2 (36.6-47.3) |
| APPE | ABER | 42.0 (38.1-46.0) | 45.2 (43.2-47.1) | 48.0 (44.9-51.1) | 42.8 (39.6-46.2) | 43.5 (39.4-48.1) |
| ABER | APPE | 17.5 (15.6-19.4) | 35.4 (33.7-37.1) | 27.4 (25.3-29.5) | 43.1 (39.9-46.3) | 59.4 (54.1-64.6) |
| APPE | PSYC | 35.7 (31.3-40.0) | 38.2 (36.0-40.1) | 40.5 (37.4-43.4) | 36.9 (33.6-40.0) | 35.0 (30.5-39.6) |
| PSYC | APPE | 11.5 ( 9.9-13.2) | 29.2 (27.5-30.7) | 23.0 (21.1-25.0) | 35.7 (32.5-38.7) | 45.7 (40.1-51.1) |
| APPE | DELU | 34.9 (30.0-39.9) | 38.5 (36.1-40.5) | 40.6 (37.1-43.8) | 37.8 (34.1-41.4) | 34.6 (29.1-39.9) |
| DELU | APPE | 8.8 ( 7.3-10.3) | 23.4 (22.0-24.8) | 18.5 (16.7-20.2) | 29.4 (26.3-32.1) | 35.1 (30.0-40.7) |
| APPE | HALL | 41.4 (34.1-48.3) | 38.9 (35.8-42.0) | 40.8 (35.9-45.8) | 38.0 (33.1-42.5) | 37.2 (31.0-43.4) |
| HALL | APPE | 4.7 ( 3.7- 5.7) | 13.1 (11.9-14.4) | 8.6 ( 7.4- 9.9) | 16.8 (14.2-19.3) | 28.4 (23.4-33.3) |
| APPE | ELAT | 43.9 (38.5-49.3) | 52.0 (47.9-56.1) | 54.5 (49.5-60.1) | 49.2 (42.1-56.5) | 49.4 (39.6-59.3) |
| ELAT | APPE | 9.3 ( 7.9-10.9) | 10.4 ( 9.3-11.4) | 9.5 ( 8.2-10.8) | 10.9 ( 8.8-13.1) | 13.7 (10.2-17.7) |
| DISI | ABER | 47.5 (43.3-51.4) | 44.8 (42.9-46.7) | 45.0 (41.9-48.0) | 42.5 (39.4-45.6) | 49.3 (44.3-54.0) |
| ABER | DISI | 27.1 (24.5-29.7) | 44.3 (42.2-46.1) | 35.2 (32.6-38.0) | 51.1 (47.6-54.4) | 65.1 (59.6-69.8) |
| DISI | PSYC | 39.7 (35.4-44.1) | 38.4 (36.5-40.5) | 36.9 (34.0-39.8) | 38.5 (35.1-41.8) | 41.9 (37.1-46.3) |
| PSYC | DISI | 17.4 (15.3-19.7) | 37.0 (35.0-39.0) | 28.8 (26.4-31.3) | 44.5 (41.2-48.1) | 52.8 (47.3-58.0) |
| DISI | DELU | 43.7 (38.7-48.7) | 41.3 (39.0-43.6) | 40.2 (37.1-43.8) | 40.7 (37.1-44.6) | 45.0 (39.4-50.1) |
| DELU | DISI | 15.0 (13.0-17.2) | 31.7 (29.8-33.5) | 25.1 (22.8-27.6) | 37.8 (34.6-41.0) | 44.1 (38.5-49.3) |
| DISI | HALL | 35.5 (28.3-42.9) | 35.9 (32.7-38.8) | 28.4 (23.9-32.8) | 38.8 (33.7-43.3) | 43.1 (36.9-48.8) |
| HALL | DISI | 5.5 ( 4.3- 7.0) | 15.3 (13.8-16.7) | 8.3 ( 6.7- 9.7) | 20.5 (17.3-23.3) | 31.8 (26.4-36.6) |
| DISI | ELAT | 65.9 (60.5-71.2) | 62.6 (58.7-66.8) | 59.9 (54.3-65.7) | 64.5 (57.6-71.1) | 67.8 (58.3-77.3) |
| ELAT | DISI | 19.1 (16.9-21.6) | 15.7 (14.2-17.3) | 14.3 (12.4-16.2) | 17.1 (14.5-20.0) | 18.2 (14.2-22.7) |
| ABER | PSYC | 25.8 (22.0-29.9) | 40.7 (38.7-42.7) | 30.0 (27.2-32.8) | 45.3 (41.8-48.7) | 57.6 (52.8-62.3) |
| PSYC | ABER | 19.9 (16.7-23.0) | 39.7 (37.7-41.7) | 29.9 (26.9-32.7) | 43.6 (40.3-46.9) | 54.9 (50.1-59.5) |
| ABER | DELU | 26.5 (22.3-31.0) | 41.8 (39.4-44.1) | 30.9 (27.8-34.2) | 47.5 (43.8-51.3) | 57.2 (51.8-62.6) |
| DELU | ABER | 16.0 (13.2-18.8) | 32.5 (30.5-34.5) | 24.6 (21.9-27.2) | 36.7 (33.6-39.8) | 42.5 (37.9-47.2) |

| | | | | | | |
|---|---|---|---|---|---|---|
| ABER | HALL | 30.2 (23.2-37.7) | 44.7 (41.9-48.0) | 31.1 (26.8-36.1) | 46.4 (41.5-51.3) | 63.6 (57.4-69.9) |
| HALL | ABER | 8.2 ( 6.1-10.4) | 19.3 (17.7-20.9) | 11.5 ( 9.6-13.4) | 20.4 (17.7-23.0) | 35.5 (30.8-40.1) |
| ABER | ELAT | 39.5 (34.2-45.2) | 50.5 (46.7-54.7) | 43.3 (38.0-48.9) | 55.3 (48.1-61.9) | 65.5 (55.4-75.8) |
| ELAT | ABER | 20.0 (16.9-23.3) | 12.8 (11.6-14.2) | 13.2 (11.1-15.3) | 12.2 ( 9.9-14.3) | 13.3 (10.1-16.6) |
| PSYC | ELAT | 20.7 (16.3-25.2) | 39.1 (35.0-42.9) | 30.8 (25.7-35.7) | 46.2 (38.9-53.1) | 52.9 (42.5-63.6) |
| ELAT | PSYC | 13.7 (10.6-16.8) | 10.2 ( 8.9-11.4) | 9.4 ( 7.7-11.2) | 10.6 ( 8.6-12.6) | 11.3 ( 8.3-14.5) |
| DELU | HALL | 39.1 (31.8-46.5) | 54.0 (50.9-57.1) | 46.1 (41.1-50.8) | 56.6 (51.9-61.5) | 62.3 (57.0-68.5) |
| HALL | DELU | 17.7 (14.1-21.3) | 30.0 (27.8-32.0) | 21.5 (18.6-24.2) | 32.2 (28.6-35.7) | 46.9 (40.7-52.3) |
| DELU | ELAT | 17.2 (13.2-21.3) | 33.9 (30.1-37.8) | 26.6 (21.8-31.3) | 40.1 (33.1-46.6) | 46.0 (35.8-56.5) |
| ELAT | DELU | 14.5 (10.8-18.1) | 11.1 ( 9.6-12.6) | 10.2 ( 8.2-12.4) | 11.4 ( 9.1-13.7) | 12.6 ( 8.9-16.4) |
| HALL | ELAT | 7.3 ( 4.6-10.2) | 16.6 (13.6-19.6) | 10.9 ( 7.5-14.6) | 19.3 (13.9-24.9) | 31.0 (21.0-40.9) |
| ELAT | HALL | 13.6 ( 8.6-18.8) | 9.8 ( 8.0-11.6) | 8.9 ( 6.2-11.9) | 9.7 ( 6.8-12.5) | 11.3 ( 7.5-15.4) |

S1|S2, conditional prevalence of symptom S1 given presence of symptom S2.
Displayed are conditional prevalence (%) and 95% confidence intervals.
NA, Not applicable; NE, Not estimable.
ABER, Aberrant motor behavior; AGIT, Agitation/aggression; ANXI, Anxiety; APAT, Apathy/indifference; APPE, Appetite/eating changes; DELU, Delusions; DEPR, Depression/dysphoria; DISI, Disinhibition; ELAT, Elation/euphoria; HALU, Hallucinations; IRRI, Irritability/lability; PSYC, Psychotic symptoms (hallucinations and/or delusions); SLEE, Nighttime behavioral disturbances.

Supplemental Table 27. Conditional prevalence S1|S2 of neuropsychiatric symptoms with the NPI severity score > 1 per MMSE stratum (NACC study only)

| | | NACC | | | | |
|---|---|---|---|---|---|---|
| S1 | S2 | MMSE >26 | MMSE ≤26 | 19 < MMSE≤ 26 | 9 < MMSE ≤ 19 | MMSE ≤ 9 |
| DEPR | IRRI | 35.3 (32.2-38.2) | 38.3 (36.2-40.6) | 37.2 (34.5-40.1) | 41.7 (37.6-46.0) | 34.8 (27.6-41.8) |
| IRRI | DEPR | 31.1 (28.2-34.0) | 37.2 (35.0-39.4) | 36.2 (33.2-39.0) | 39.2 (35.1-43.2) | 37.4 (30.4-44.4) |
| DEPR | ANXI | 46.0 (42.6-49.2) | 43.3 (41.3-45.6) | 42.9 (40.1-46.1) | 45.1 (41.2-48.9) | 40.9 (35.2-47.2) |
| ANXI | DEPR | 37.5 (34.7-40.4) | 46.6 (44.3-48.9) | 42.1 (39.3-44.9) | 50.9 (46.6-55.1) | 60.9 (53.2-68.3) |
| DEPR | APAT | 44.8 (41.2-48.5) | 33.4 (31.6-35.4) | 35.8 (33.2-38.7) | 32.9 (29.7-36.3) | 26.6 (22.1-31.1) |
| APAT | DEPR | 30.0 (27.3-32.9) | 44.3 (41.8-46.7) | 40.4 (37.3-43.6) | 47.8 (43.8-52.1) | 56.9 (49.7-63.9) |
| DEPR | SLEE | 29.7 (26.9-32.4) | 32.7 (30.6-35.0) | 32.5 (29.8-35.6) | 33.7 (29.5-37.7) | 31.1 (24.9-37.2) |
| SLEE | DEPR | 29.2 (26.5-32.0) | 30.2 (28.2-32.4) | 29.1 (26.4-31.8) | 31.0 (27.3-35.0) | 34.5 (27.5-42.2) |
| DEPR | AGIT | 37.5 (33.7-41.0) | 38.9 (36.6-41.2) | 38.5 (35.5-41.8) | 43.1 (38.7-47.5) | 32.3 (27.0-37.8) |
| AGIT | DEPR | 23.0 (20.4-25.6) | 35.6 (33.4-37.9) | 31.3 (28.5-34.2) | 40.3 (36.1-44.7) | 47.7 (40.2-55.1) |
| DEPR | APPE | 30.5 (26.6-34.6) | 31.4 (28.9-33.9) | 30.9 (27.9-34.3) | 34.6 (30.2-39.1) | 24.8 (18.5-32.3) |
| APPE | DEPR | 15.8 (13.7-18.2) | 23.9 (22.1-26.0) | 22.9 (20.6-25.4) | 26.6 (23.0-30.2) | 21.8 (16.2-28.4) |
| DEPR | DISI | 33.4 (29.3-37.9) | 31.9 (29.1-34.8) | 30.4 (26.9-34.3) | 37.4 (32.6-42.7) | 26.3 (19.5-33.3) |
| DISI | DEPR | 13.6 (11.6-15.7) | 19.3 (17.6-21.2) | 17.0 (14.9-19.3) | 22.7 (19.3-25.9) | 23.0 (17.2-29.3) |
| DEPR | ABER | 33.2 (27.8-39.1) | 31.3 (28.6-34.0) | 32.3 (27.7-36.8) | 31.8 (27.4-36.0) | 28.6 (22.7-34.4) |
| ABER | DEPR | 8.8 ( 7.1-10.5) | 19.7 (17.9-21.6) | 13.4 (11.5-15.6) | 26.0 (22.3-29.7) | 39.1 (31.7-46.4) |
| DEPR | PSYC | 37.6 (30.7-44.8) | 35.5 (32.7-38.1) | 34.6 (30.3-39.0) | 36.8 (32.3-41.1) | 34.8 (28.6-41.6) |
| PSYC | DEPR | 6.1 ( 4.7- 7.6) | 21.6 (19.6-23.4) | 15.0 (12.8-17.1) | 28.6 (25.1-32.4) | 40.2 (33.3-47.8) |
| DEPR | DELU | 38.4 (30.3-46.4) | 36.7 (33.5-39.7) | 36.9 (32.0-41.6) | 37.1 (32.1-42.2) | 35.3 (28.0-43.3) |
| DELU | DEPR | 5.0 ( 3.7- 6.3) | 17.9 (16.2-19.7) | 12.8 (10.9-14.8) | 23.8 (20.3-27.6) | 31.0 (24.6-38.1) |
| DEPR | HALL | 40.4 (27.3-53.9) | 36.1 (31.8-40.6) | 35.3 (27.6-43.1) | 36.6 (29.8-44.0) | 36.5 (28.0-45.5) |
| HALL | DEPR | 2.0 ( 1.3- 3.0) | 8.8 ( 7.5-10.0) | 5.2 ( 3.9- 6.6) | 11.0 ( 8.6-13.6) | 24.1 (17.8-30.7) |
| DEPR | ELAT | 29.0 (21.3-37.0) | 34.9 (29.0-41.2) | 31.4 (23.8-39.8) | 39.4 (27.8-51.3) | 39.1 (19.2-60.0) |
| ELAT | DEPR | 3.8 ( 2.6- 5.0) | 4.2 ( 3.4- 5.2) | 3.6 ( 2.6- 4.7) | 5.2 ( 3.5- 7.2) | 5.2 ( 2.2- 8.9) |

| | | | | | | |
|---|---|---|---|---|---|---|
| IRRI | ANXI | 36.8 (33.6-40.2) | 39.8 (37.7-42.0) | 39.7 (36.8-42.6) | 39.5 (35.6-43.2) | 40.9 (34.9-46.6) |
| ANXI | IRRI | 34.1 (30.8-37.1) | 44.1 (41.8-46.7) | 40.2 (37.4-43.3) | 47.4 (43.3-51.6) | 56.7 (49.5-64.2) |
| IRRI | APAT | 38.0 (34.2-41.4) | 33.0 (31.1-34.9) | 35.9 (33.2-38.6) | 31.2 (27.9-34.2) | 27.4 (23.1-32.2) |
| APAT | IRRI | 28.9 (26.1-31.6) | 45.0 (42.8-47.2) | 41.7 (38.7-44.6) | 48.0 (43.6-52.5) | 54.5 (47.3-62.0) |
| IRRI | SLEE | 24.9 (22.2-27.4) | 35.0 (32.7-37.2) | 33.8 (30.8-36.6) | 34.7 (30.5-38.8) | 41.5 (34.9-48.6) |
| SLEE | IRRI | 27.7 (24.8-30.5) | 33.2 (31.2-35.3) | 31.1 (28.4-33.7) | 34.0 (29.8-38.2) | 42.8 (35.6-50.3) |
| IRRI | AGIT | 59.1 (55.3-63.0) | 56.6 (54.3-58.9) | 59.0 (55.9-62.3) | 56.7 (52.7-60.9) | 47.9 (41.7-54.1) |
| AGIT | IRRI | 41.2 (38.2-44.5) | 53.2 (51.0-55.6) | 49.4 (46.5-52.3) | 56.5 (52.4-60.6) | 65.8 (59.0-72.1) |
| IRRI | APPE | 31.2 (27.1-35.2) | 35.6 (32.9-38.0) | 36.1 (32.6-39.4) | 36.3 (31.3-40.6) | 30.7 (23.5-37.6) |
| APPE | IRRI | 18.3 (15.7-20.9) | 27.9 (25.8-29.9) | 27.5 (24.9-30.3) | 29.6 (25.7-33.3) | 25.1 (18.6-31.4) |
| IRRI | DISI | 54.4 (49.9-58.9) | 53.7 (50.8-56.8) | 54.0 (50.2-58.0) | 52.1 (46.6-57.8) | 55.9 (48.5-64.2) |
| DISI | IRRI | 25.1 (22.3-27.9) | 33.5 (31.3-35.7) | 31.2 (28.5-34.0) | 33.6 (29.1-38.0) | 45.5 (38.3-52.5) |
| IRRI | ABER | 47.9 (42.2-54.0) | 43.2 (40.3-46.1) | 47.8 (43.2-52.8) | 39.1 (34.3-43.8) | 42.4 (36.4-48.6) |
| ABER | IRRI | 14.5 (12.2-16.8) | 28.0 (25.8-30.2) | 20.4 (17.9-23.0) | 34.0 (29.9-38.4) | 54.0 (46.7-61.0) |
| IRRI | PSYC | 48.2 (40.7-55.5) | 40.4 (37.6-43.3) | 41.8 (37.6-46.4) | 37.9 (32.8-42.6) | 42.3 (35.5-49.5) |
| PSYC | IRRI | 8.8 ( 7.2-10.7) | 25.3 (23.3-27.4) | 18.6 (16.4-20.9) | 31.4 (27.6-35.5) | 45.5 (38.6-52.4) |
| IRRI | DELU | 51.4 (43.0-59.2) | 43.9 (40.7-47.3) | 46.3 (41.3-51.5) | 40.6 (35.0-45.6) | 45.8 (37.6-53.6) |
| DELU | IRRI | 7.7 ( 6.0- 9.4) | 22.1 (20.2-24.1) | 16.6 (14.5-18.7) | 27.7 (23.8-31.6) | 37.4 (30.8-44.4) |
| IRRI | HALL | 40.4 (27.5-54.5) | 35.6 (31.0-40.0) | 30.8 (23.6-39.2) | 34.8 (27.2-42.0) | 43.5 (34.0-52.5) |
| HALL | IRRI | 2.3 ( 1.4- 3.3) | 8.9 ( 7.5-10.3) | 4.7 ( 3.5- 6.1) | 11.1 ( 8.5-13.7) | 26.7 (20.1-32.9) |
| IRRI | ELAT | 49.3 (41.6-58.0) | 51.6 (45.3-58.7) | 55.4 (47.1-64.2) | 43.7 (32.3-55.4) | 56.5 (36.7-76.9) |
| ELAT | IRRI | 7.3 ( 5.8- 9.0) | 6.4 ( 5.3- 7.7) | 6.5 ( 5.1- 8.0) | 6.1 ( 4.2- 8.2) | 7.0 ( 3.7-10.8) |
| ANXI | APAT | 30.6 (27.3-33.9) | 34.3 (32.5-36.2) | 32.7 (30.1-35.3) | 35.5 (32.0-38.8) | 36.8 (32.0-42.0) |
| APAT | ANXI | 25.2 (22.1-28.0) | 42.2 (40.0-44.4) | 37.6 (34.9-40.7) | 45.6 (41.6-49.4) | 52.9 (47.0-58.9) |
| ANXI | SLEE | 24.7 (22.1-27.2) | 34.8 (32.8-37.2) | 31.3 (28.5-34.4) | 37.5 (33.2-41.8) | 45.1 (38.2-51.8) |
| SLEE | ANXI | 29.7 (26.8-32.8) | 29.9 (28.0-32.0) | 28.5 (25.8-31.2) | 30.6 (27.1-34.3) | 33.6 (27.7-39.4) |
| ANXI | AGIT | 33.9 (30.3-37.4) | 42.1 (39.7-44.3) | 39.0 (35.6-42.2) | 44.2 (39.9-48.3) | 48.2 (42.3-54.1) |
| AGIT | ANXI | 25.5 (22.4-28.5) | 35.8 (33.7-37.9) | 32.2 (29.3-35.0) | 36.7 (32.9-40.6) | 47.9 (41.9-53.8) |
| ANXI | APPE | 27.8 (24.0-31.7) | 33.4 (30.9-36.0) | 30.5 (27.4-33.9) | 35.8 (31.2-40.6) | 41.8 (34.2-49.4) |

| | | | | | | |
|---|---|---|---|---|---|---|
| APPE | ANXI | 17.6 (15.2-20.1) | 23.7 (21.8-25.5) | 23.0 (20.6-25.7) | 24.3 (20.9-27.7) | 24.7 (19.6-30.4) |
| ANXI | DISI | 35.5 (31.2-40.0) | 41.2 (38.4-44.1) | 39.3 (35.4-43.4) | 41.4 (36.1-46.9) | 48.7 (40.6-56.4) |
| DISI | ANXI | 17.7 (15.1-20.2) | 23.2 (21.3-25.0) | 22.5 (20.1-25.0) | 22.2 (19.1-25.4) | 28.6 (23.0-34.0) |
| ANXI | ABER | 39.6 (34.1-45.7) | 44.4 (41.5-47.4) | 45.3 (40.9-50.3) | 42.3 (37.6-46.9) | 46.6 (40.3-52.9) |
| ABER | ANXI | 12.9 (10.7-15.1) | 26.0 (24.0-28.0) | 19.1 (16.7-21.8) | 30.6 (26.9-34.2) | 42.9 (37.1-49.0) |
| ANXI | PSYC | 40.0 (32.4-47.7) | 41.5 (38.5-44.5) | 37.5 (33.0-41.9) | 41.8 (36.9-46.2) | 50.2 (43.5-57.4) |
| PSYC | ANXI | 7.9 ( 6.1- 9.7) | 23.5 (21.6-25.3) | 16.5 (14.1-18.7) | 28.8 (25.1-32.3) | 39.0 (33.2-45.0) |
| ANXI | DELU | 38.4 (30.6-47.0) | 42.4 (39.2-45.6) | 38.5 (33.7-43.4) | 41.2 (35.8-46.2) | 54.9 (47.1-62.6) |
| DELU | ANXI | 6.2 ( 4.6- 7.9) | 19.3 (17.5-21.0) | 13.6 (11.6-15.6) | 23.4 (20.1-26.8) | 32.4 (27.1-38.1) |
| ANXI | HALL | 48.1 (34.5-62.2) | 44.9 (40.1-49.5) | 39.1 (31.3-46.6) | 47.2 (39.7-55.0) | 49.6 (40.3-58.9) |
| HALL | ANXI | 2.9 ( 1.8- 4.2) | 10.2 ( 8.7-11.5) | 5.9 ( 4.3- 7.4) | 12.5 (10.0-15.0) | 22.0 (16.9-27.2) |
| ANXI | ELAT | 35.5 (27.2-43.7) | 44.7 (38.5-51.8) | 40.5 (31.7-49.6) | 50.7 (38.9-62.8) | 47.8 (26.1-68.2) |
| ELAT | ANXI | 5.7 ( 4.2- 7.3) | 5.0 ( 4.1- 6.0) | 4.7 ( 3.5- 6.0) | 5.9 ( 4.1- 7.9) | 4.2 ( 2.0- 7.1) |
| APAT | SLEE | 25.7 (22.9-28.1) | 44.4 (42.0-46.8) | 39.5 (36.4-42.6) | 48.0 (43.7-52.3) | 59.1 (51.9-65.9) |
| SLEE | APAT | 37.6 (34.2-41.2) | 30.9 (29.0-32.8) | 31.3 (28.7-33.8) | 30.5 (27.2-33.8) | 30.6 (26.0-35.4) |
| APAT | AGIT | 35.0 (31.2-38.3) | 47.3 (45.0-49.7) | 45.8 (42.6-49.1) | 48.4 (43.9-52.7) | 50.2 (44.0-56.1) |
| AGIT | APAT | 32.1 (28.4-35.4) | 32.7 (30.9-34.7) | 32.9 (30.3-35.6) | 31.3 (28.0-34.7) | 34.7 (30.1-39.6) |
| APAT | APPE | 35.3 (31.3-39.1) | 49.0 (46.3-51.6) | 45.5 (42.1-49.1) | 53.0 (48.2-57.9) | 55.6 (48.7-63.5) |
| APPE | APAT | 27.2 (24.1-30.3) | 28.2 (26.5-30.0) | 29.9 (27.4-32.5) | 28.1 (25.2-31.0) | 22.8 (18.7-27.3) |
| APAT | DISI | 45.3 (40.8-50.1) | 51.9 (49.1-54.9) | 51.7 (47.8-56.0) | 47.9 (42.5-53.0) | 61.2 (53.5-68.7) |
| DISI | APAT | 27.5 (24.3-30.9) | 23.7 (22.0-25.4) | 25.7 (23.6-28.1) | 20.0 (17.2-22.8) | 25.0 (20.6-29.6) |
| APAT | ABER | 48.6 (42.6-54.1) | 55.1 (52.3-58.0) | 54.9 (50.5-59.7) | 53.0 (48.4-57.3) | 59.7 (53.8-66.1) |
| ABER | APAT | 19.3 (16.5-22.1) | 26.2 (24.5-28.0) | 20.1 (18.0-22.5) | 29.9 (26.7-32.9) | 38.2 (33.6-43.5) |
| APAT | PSYC | 35.9 (28.6-42.5) | 43.1 (40.3-45.9) | 40.3 (36.0-44.8) | 42.2 (37.5-46.9) | 51.2 (44.6-58.3) |
| PSYC | APAT | 8.7 ( 6.7-10.7) | 19.8 (18.1-21.3) | 15.4 (13.4-17.4) | 22.7 (19.9-25.7) | 27.7 (23.0-32.3) |
| APAT | DELU | 37.7 (29.6-45.1) | 44.3 (41.2-47.6) | 40.9 (36.2-45.8) | 42.9 (37.5-48.2) | 55.6 (48.4-63.3) |
| DELU | APAT | 7.4 ( 5.5- 9.4) | 16.3 (14.9-17.8) | 12.6 (10.8-14.5) | 19.0 (16.2-21.9) | 22.8 (19.1-26.8) |
| APAT | HALL | 40.4 (26.8-54.4) | 45.6 (40.8-50.1) | 43.6 (35.9-51.2) | 45.3 (38.0-53.2) | 48.7 (39.0-58.0) |
| HALL | APAT | 3.0 ( 1.8- 4.4) | 8.4 ( 7.2- 9.4) | 5.7 ( 4.3- 7.0) | 9.4 ( 7.4-11.3) | 15.1 (11.4-18.7) |

| | | | | | | |
|---|---|---|---|---|---|---|
| APAT | ELAT | 50.0 (41.3-58.5) | 57.2 (50.7-63.9) | 58.7 (49.2-67.8) | 52.1 (40.9-64.0) | 65.2 (44.0-85.7) |
| ELAT | APAT | 9.8 ( 7.7-11.9) | 5.2 ( 4.4- 6.2) | 5.9 ( 4.6- 7.4) | 4.7 ( 3.4- 6.4) | 4.0 ( 2.2- 6.2) |
| SLEE | AGIT | 31.0 (27.5-34.6) | 32.2 (30.0-34.4) | 30.5 (27.6-33.6) | 32.7 (28.7-36.7) | 36.6 (30.2-42.8) |
| AGIT | SLEE | 19.4 (16.9-21.7) | 31.8 (29.7-34.0) | 27.7 (25.0-30.5) | 33.3 (29.0-37.3) | 48.7 (41.9-55.7) |
| SLEE | APPE | 38.4 (34.2-42.5) | 37.3 (34.8-40.1) | 35.5 (32.3-38.8) | 38.3 (33.7-43.3) | 43.8 (35.4-52.0) |
| APPE | SLEE | 20.2 (17.8-22.7) | 30.8 (28.6-33.0) | 29.4 (26.6-32.2) | 31.9 (27.8-36.1) | 34.7 (27.7-41.9) |
| SLEE | DISI | 35.3 (30.8-39.9) | 35.2 (32.4-38.1) | 33.4 (29.8-37.2) | 35.3 (30.4-40.5) | 42.1 (33.8-49.7) |
| DISI | SLEE | 14.6 (12.5-16.8) | 23.1 (21.1-25.0) | 20.9 (18.5-23.5) | 23.2 (19.5-26.9) | 33.2 (26.8-39.5) |
| SLEE | ABER | 40.4 (34.7-45.9) | 38.9 (36.2-41.7) | 40.1 (35.6-44.7) | 36.6 (32.7-40.7) | 41.2 (34.3-47.5) |
| ABER | SLEE | 10.9 ( 9.1-12.8) | 26.5 (24.4-28.7) | 18.5 (16.1-21.0) | 32.5 (28.6-36.5) | 50.8 (43.2-57.8) |
| SLEE | PSYC | 37.1 (30.1-44.6) | 36.3 (33.5-39.4) | 34.0 (29.8-38.5) | 35.3 (30.8-39.9) | 43.8 (36.5-51.0) |
| PSYC | SLEE | 6.1 ( 4.6- 7.6) | 23.9 (21.9-25.9) | 16.4 (14.1-18.8) | 29.8 (25.9-33.7) | 45.6 (37.9-52.8) |
| SLEE | DELU | 33.3 (25.3-41.7) | 34.0 (31.0-37.4) | 30.4 (25.8-35.1) | 32.8 (27.9-37.8) | 45.8 (37.4-53.8) |
| DELU | SLEE | 4.5 ( 3.2- 5.8) | 18.0 (16.3-19.9) | 11.8 ( 9.9-13.8) | 22.8 (19.3-26.5) | 36.3 (29.5-43.0) |
| SLEE | HALL | 53.8 (39.1-66.7) | 50.0 (45.1-54.8) | 50.6 (42.8-58.7) | 49.1 (41.7-57.1) | 50.4 (40.5-59.2) |
| HALL | SLEE | 2.7 ( 1.7- 3.8) | 13.2 (11.4-14.7) | 8.3 ( 6.6-10.0) | 15.9 (12.9-18.8) | 30.1 (23.2-36.5) |
| SLEE | ELAT | 37.0 (28.9-45.0) | 40.5 (34.2-47.3) | 39.7 (31.0-48.3) | 36.6 (25.5-48.3) | 56.5 (35.0-76.9) |
| ELAT | SLEE | 4.9 ( 3.7- 6.2) | 5.3 ( 4.3- 6.4) | 5.1 ( 3.7- 6.5) | 5.2 ( 3.4- 7.2) | 6.7 ( 3.5-10.5) |
| AGIT | APPE | 25.0 (21.4-28.6) | 32.0 (29.5-34.4) | 30.5 (27.3-33.6) | 33.7 (29.0-38.1) | 34.6 (27.5-41.9) |
| APPE | AGIT | 21.1 (18.1-24.4) | 26.6 (24.4-28.7) | 27.8 (24.9-30.7) | 27.6 (23.6-31.4) | 20.6 (15.6-25.8) |
| AGIT | DISI | 47.0 (41.8-51.7) | 48.6 (45.6-51.7) | 47.1 (43.3-51.2) | 49.7 (44.6-55.1) | 52.0 (43.4-60.0) |
| DISI | AGIT | 31.1 (27.5-34.8) | 32.2 (29.9-34.4) | 32.6 (29.6-35.7) | 32.1 (28.2-36.1) | 30.7 (25.0-36.3) |
| AGIT | ABER | 40.0 (33.9-45.8) | 42.4 (39.6-45.3) | 44.2 (39.4-48.9) | 39.1 (34.4-43.5) | 45.4 (39.3-52.2) |
| ABER | AGIT | 17.3 (14.2-20.2) | 29.2 (27.0-31.4) | 22.5 (19.7-25.4) | 34.1 (30.0-38.2) | 42.0 (35.6-48.7) |
| AGIT | PSYC | 47.6 (40.0-55.2) | 46.2 (43.2-49.3) | 42.5 (38.3-47.1) | 44.2 (39.5-48.8) | 59.2 (52.6-66.0) |
| PSYC | AGIT | 12.5 (10.1-15.2) | 30.7 (28.4-32.9) | 22.6 (19.9-25.3) | 36.7 (32.5-40.8) | 46.3 (39.8-52.2) |
| AGIT | DELU | 53.6 (45.4-62.1) | 49.8 (46.4-53.3) | 47.4 (42.4-52.5) | 46.7 (41.4-51.4) | 62.7 (55.5-70.4) |
| DELU | AGIT | 11.5 ( 9.3-13.9) | 26.6 (24.3-28.7) | 20.3 (17.8-22.8) | 31.9 (27.8-35.8) | 37.4 (31.4-43.0) |
| AGIT | HALL | 38.5 (26.1-52.5) | 40.5 (35.7-45.5) | 28.2 (21.2-35.4) | 41.6 (33.5-49.7) | 55.7 (46.8-64.6) |

| | | | | | | |
|---|---|---|---|---|---|---|
| HALL | AGIT | 3.1 ( 1.8- 4.6) | 10.8 ( 9.2-12.3) | 5.1 ( 3.7- 6.7) | 13.3 (10.3-16.3) | 24.9 (19.3-30.3) |
| AGIT | ELAT | 46.4 (38.1-55.8) | 49.8 (42.9-56.7) | 51.2 (42.9-59.6) | 42.3 (30.9-53.7) | 65.2 (44.4-84.2) |
| ELAT | AGIT | 9.9 ( 7.7-12.2) | 6.6 ( 5.4- 7.9) | 7.2 ( 5.6- 9.0) | 6.0 ( 4.1- 8.1) | 5.8 ( 3.2- 9.2) |
| APPE | DISI | 30.6 (26.3-35.0) | 34.8 (31.9-37.7) | 35.4 (31.5-39.3) | 35.0 (29.8-39.9) | 32.2 (24.8-39.2) |
| DISI | APPE | 24.1 (20.3-27.6) | 27.7 (25.2-29.9) | 26.8 (23.6-29.8) | 27.6 (23.0-31.7) | 32.0 (24.3-39.1) |
| APPE | ABER | 36.8 (30.8-42.4) | 34.6 (31.7-37.3) | 41.7 (37.0-46.2) | 30.9 (26.6-35.2) | 28.2 (22.7-34.1) |
| ABER | APPE | 18.9 (15.6-22.3) | 28.6 (26.1-30.8) | 23.3 (20.3-26.1) | 32.9 (28.5-37.4) | 43.8 (37.0-51.9) |
| APPE | PSYC | 32.4 (25.1-39.8) | 25.0 (22.4-27.7) | 26.6 (22.9-30.7) | 23.4 (19.4-27.3) | 24.9 (19.2-31.3) |
| PSYC | APPE | 10.1 ( 7.7-12.9) | 20.0 (17.9-22.0) | 15.5 (13.0-18.1) | 23.7 (19.8-27.7) | 32.7 (25.7-40.3) |
| APPE | DELU | 31.9 (24.2-39.7) | 26.0 (23.0-29.0) | 27.4 (23.2-32.1) | 24.3 (19.9-28.7) | 26.1 (19.4-33.3) |
| DELU | APPE | 8.1 ( 5.8-10.5) | 16.6 (14.7-18.6) | 12.8 (10.5-15.1) | 20.3 (16.6-24.1) | 26.1 (19.5-33.6) |
| APPE | HALL | 32.7 (20.0-47.2) | 25.5 (21.2-29.6) | 26.9 (20.4-34.2) | 25.5 (19.1-32.0) | 23.5 (16.0-31.5) |
| HALL | APPE | 3.1 ( 1.7- 4.7) | 8.1 ( 6.7- 9.5) | 5.3 ( 3.9- 7.0) | 9.9 ( 7.2-12.6) | 17.6 (12.1-23.7) |
| APPE | ELAT | 42.0 (33.3-50.0) | 51.6 (44.6-58.2) | 53.7 (45.1-62.4) | 47.9 (35.9-60.0) | 52.2 (30.0-73.3) |
| ELAT | APPE | 10.7 ( 8.1-13.2) | 8.2 ( 6.7- 9.7) | 8.3 ( 6.3-10.1) | 8.2 ( 5.6-11.0) | 7.8 ( 3.8-12.7) |
| DISI | ABER | 41.4 (35.8-47.3) | 36.4 (33.7-38.9) | 40.5 (35.8-45.0) | 32.3 (27.9-36.3) | 36.6 (30.7-42.7) |
| ABER | DISI | 27.1 (22.8-31.5) | 37.9 (34.8-40.7) | 29.9 (26.2-33.7) | 43.6 (38.2-49.1) | 57.2 (49.7-64.8) |
| DISI | PSYC | 31.2 (25.0-37.6) | 27.6 (25.0-30.2) | 28.3 (24.2-32.7) | 26.5 (22.4-30.6) | 28.4 (22.4-34.8) |
| PSYC | DISI | 12.4 ( 9.5-15.7) | 27.7 (24.9-30.5) | 21.8 (18.5-25.1) | 34.0 (29.2-39.4) | 37.5 (29.8-45.2) |
| DISI | DELU | 34.8 (27.2-42.1) | 30.0 (27.1-33.1) | 31.4 (26.7-36.4) | 28.1 (23.5-32.8) | 30.7 (23.7-38.2) |
| DELU | DISI | 11.2 ( 8.5-14.5) | 24.2 (21.7-27.0) | 19.5 (16.1-22.6) | 29.8 (25.1-34.9) | 30.9 (24.1-38.7) |
| DISI | HALL | 25.0 (13.3-37.9) | 28.2 (24.0-32.4) | 23.1 (16.8-30.1) | 29.2 (22.2-36.2) | 33.9 (24.8-42.9) |
| HALL | DISI | 3.0 ( 1.6- 4.8) | 11.4 ( 9.3-13.2) | 6.0 ( 4.2- 8.0) | 14.4 (10.6-18.2) | 25.7 (18.3-32.4) |
| DISI | ELAT | 59.4 (51.5-67.5) | 60.0 (53.6-66.5) | 60.3 (51.6-68.5) | 57.7 (45.7-69.0) | 65.2 (44.4-85.0) |
| ELAT | DISI | 19.2 (15.4-22.9) | 12.0 (10.2-14.0) | 12.2 ( 9.8-14.8) | 12.6 ( 9.2-16.3) | 9.9 ( 5.5-15.0) |
| ABER | PSYC | 25.3 (18.8-31.9) | 30.4 (27.6-33.1) | 20.9 (17.4-24.7) | 34.8 (30.2-39.2) | 42.8 (35.8-49.7) |
| PSYC | ABER | 15.4 (11.1-19.9) | 29.4 (26.8-32.1) | 21.9 (18.0-25.7) | 33.2 (28.9-37.1) | 36.1 (29.6-42.1) |
| ABER | DELU | 26.1 (18.4-33.3) | 32.5 (29.4-35.7) | 21.4 (17.3-25.7) | 38.3 (33.4-43.2) | 46.4 (38.7-54.2) |
| DELU | ABER | 12.9 ( 8.8-17.0) | 25.2 (22.9-27.8) | 18.0 (14.5-21.5) | 30.0 (26.0-34.2) | 29.8 (24.0-35.4) |

| | | | | | | |
|---|---|---|---|---|---|---|
| ABER | HALL | 28.8 (16.7-42.6) | 34.7 (30.1-39.0) | 25.6 (19.3-32.7) | 35.4 (28.2-43.1) | 46.1 (36.4-55.1) |
| HALL | ABER | 5.4 ( 2.7- 8.3) | 13.4 (11.4-15.3) | 9.1 ( 6.5-11.8) | 13.0 (10.0-15.9) | 22.3 (16.8-27.3) |
| ABER | ELAT | 35.5 (28.2-43.5) | 47.4 (40.8-53.9) | 41.3 (32.0-50.5) | 57.7 (46.3-69.0) | 47.8 (26.7-69.6) |
| ELAT | ABER | 17.5 (13.4-21.8) | 9.1 ( 7.5-10.9) | 11.4 ( 8.7-14.4) | 9.3 ( 6.7-12.3) | 4.6 ( 2.1- 7.4) |
| PSYC | ELAT | 15.9 ( 9.7-22.2) | 28.8 (22.4-35.4) | 22.3 (15.5-30.4) | 35.2 (24.4-46.8) | 43.5 (23.1-64.7) |
| ELAT | PSYC | 12.9 ( 7.8-18.7) | 5.7 ( 4.4- 7.2) | 5.9 ( 3.9- 8.2) | 6.0 ( 3.9- 8.3) | 5.0 ( 2.1- 8.3) |
| DELU | HALL | 38.5 (25.4-52.6) | 50.9 (46.0-55.6) | 42.3 (34.9-50.0) | 54.0 (46.8-61.9) | 58.3 (48.8-67.5) |
| HALL | DELU | 14.5 ( 9.0-20.4) | 25.4 (22.2-28.3) | 17.9 (13.8-22.0) | 25.2 (20.9-29.5) | 43.8 (35.3-51.8) |
| DELU | ELAT | 15.2 ( 9.3-21.4) | 25.1 (19.3-31.6) | 19.8 (13.2-27.3) | 32.4 (21.4-43.8) | 30.4 (13.0-52.6) |
| ELAT | DELU | 15.2 ( 9.0-22.1) | 6.2 ( 4.7- 7.9) | 6.5 ( 4.2- 9.1) | 6.7 ( 4.2- 9.4) | 4.6 ( 1.8- 8.4) |
| HALL | ELAT | 3.6 ( 0.7- 7.2) | 11.6 ( 7.5-16.2) | 8.3 ( 3.8-13.8) | 12.7 ( 5.3-21.3) | 26.1 ( 8.3-44.4) |
| ELAT | HALL | 9.6 ( 2.1-18.3) | 5.8 ( 3.7- 8.2) | 6.4 ( 3.0-10.9) | 5.6 ( 2.3- 9.6) | 5.2 ( 1.7- 9.4) |

S1|S2, conditional prevalence of symptom S1 given presence of symptom S2.
Displayed are conditional prevalence (%) and 95% confidence intervals.
NA, Not applicable; NE, Not estimable.
ABER, Aberrant motor behavior; AGIT, Agitation/aggression; ANXI, Anxiety; APAT, Apathy/indifference; APPE, Appetite/eating changes; DELU, Delusions; DEPR, Depression/dysphoria; DISI, Disinhibition; ELAT, Elation/euphoria; HALU, Hallucinations; IRRI, Irritability/lability; PSYC, Psychotic symptoms (hallucinations and/or delusions); SLEE, Nighttime behavioral disturbances.

Supplemental Table 28. Conditional prevalence S1|S2 of neuropsychiatric symptoms with the NPI product score ≥ 4 (ADAMS and ADNI studies only), all participants

| S1 | S2 | ADAMS survey | ADAMS | ADNI |
|---|---|---|---|---|
| DEPR | IRRI | 34.1 (NE-NE) | 33.3 (16.0- 52.2) | 20.5 ( 7.5- 34.4) |
| IRRI | DEPR | 22.4 (NE-NE) | 25.0 (10.8-40.0) | 38.1 (15.0-61.1) |
| DEPR | ANXI | 32.7 (NE-NE) | 32.1 (14.8- 50.0) | 20.0 ( 5.3- 40.9) |
| ANXI | DEPR | 19.9 (NE-NE) | 25.0 (10.3-39.5) | 19.0 ( 4.5-36.8) |
| DEPR | APAT | 22.5 (NE-NE) | 31.2 (18.4- 44.8) | 16.0 ( 7.3- 26.8) |
| APAT | DEPR | 26.2 (NE-NE) | 41.7 (25.8-59.3) | 38.1 (20.0-59.1) |
| DEPR | SLEE | NA | NE | 14.9 ( 5.0- 26.7) |
| SLEE | DEPR | NA | NE | 33.3 (12.5-53.6) |
| DEPR | AGIT | 39.0 (NE-NE) | 29.6 (13.6- 47.8) | 5.9 ( 0.0- 20.0) |
| AGIT | DEPR | 21.2 (NE-NE) | 22.2 ( 9.7-35.6) | 4.8 ( 0.0-16.7) |
| DEPR | APPE | NA | NE | 14.9 ( 5.0- 26.7) |
| APPE | DEPR | NA | NE | 33.3 (13.0-53.8) |
| DEPR | DISI | 48.7 (NE-NE) | 36.4 ( 8.3- 66.7) | 16.7 ( 0.0- 40.0) |
| DISI | DEPR | 16.1 (NE-NE) | 11.1 ( 2.3-22.6) | 9.5 ( 0.0-25.0) |
| DEPR | ABER | 27.8 (NE-NE) | 11.5 ( 0.0- 25.9) | 7.1 ( 0.0- 25.0) |
| ABER | DEPR | 12.8 (NE-NE) | 8.3 ( 0.0-18.5) | 4.8 ( 0.0-16.7) |
| DEPR | PSYC | 27.7 (NE-NE) | 25.9 ( 9.5- 43.5) | 11.1 ( 0.0- 36.4) |
| PSYC | DEPR | 17.8 (NE-NE) | 19.4 ( 6.7-33.3) | 4.8 ( 0.0-15.4) |
| DEPR | DELU | 26.4 (NE-NE) | 26.1 ( 9.5- 44.8) | 11.1 ( 0.0- 36.4) |
| DELU | DEPR | 16.0 (NE-NE) | 16.7 ( 5.3-29.7) | 4.8 ( 0.0-15.4) |
| DEPR | HALL | 40.9 (NE-NE) | 30.0 ( 0.0- 60.0) | NE |
| HALL | DEPR | 3.1 (NE-NE) | 8.3 ( 0.0-19.0) | NE |
| DEPR | ELAT | NE | NE | 16.7 ( 0.0- 50.0) |
| ELAT | DEPR | NE | NE | 4.8 ( 0.0-15.4) |
| IRRI | ANXI | 13.7 (NE-NE) | 32.1 (14.8- 51.9) | 15.0 ( 0.0- 34.8) |
| ANXI | IRRI | 12.7 (NE-NE) | 33.3 (15.8-52.2) | 7.7 ( 0.0-17.8) |
| IRRI | APAT | 18.9 (NE-NE) | 16.7 ( 6.7- 28.1) | 20.0 ( 9.5- 32.1) |
| APAT | IRRI | 33.6 (NE-NE) | 29.6 (12.5-48.0) | 25.6 (12.2-39.0) |
| IRRI | SLEE | NA | NE | 10.6 ( 2.2- 20.0) |

| | | | | |
|---|---|---|---|---|
| SLEE | IRRI | NA | NE | 12.8 ( 2.7-24.4) |
| IRRI | AGIT | 77.9 (NE-NE) | 59.3 (40.9- 76.9) | 35.3 ( 13.0- 57.1) |
| AGIT | IRRI | 64.6 (NE-NE) | 59.3 (40.9-79.2) | 15.4 ( 5.4-27.5) |
| IRRI | APPE | NA | NE | 17.0 ( 6.5- 27.5) |
| APPE | IRRI | NA | NE | 20.5 ( 8.3-34.1) |
| IRRI | DISI | 58.9 (NE-NE) | 72.7 (42.9-100.0) | 50.0 ( 20.0- 80.0) |
| DISI | IRRI | 29.7 (NE-NE) | 29.6 (13.3-48.0) | 15.4 ( 5.3-27.3) |
| IRRI | ABER | 35.2 (NE-NE) | 23.1 ( 7.7- 42.9) | 14.3 ( 0.0- 35.7) |
| ABER | IRRI | 24.7 (NE-NE) | 22.2 ( 7.5-40.0) | 5.1 ( 0.0-13.3) |
| IRRI | PSYC | 32.7 (NE-NE) | 40.7 (22.6- 60.0) | 55.6 ( 20.0- 87.5) |
| PSYC | IRRI | 32.0 (NE-NE) | 40.7 (23.5-60.0) | 12.8 ( 2.9-23.4) |
| IRRI | DELU | 30.7 (NE-NE) | 39.1 (19.0- 61.1) | 55.6 ( 20.0- 87.5) |
| DELU | IRRI | 28.4 (NE-NE) | 33.3 (16.1-52.4) | 12.8 ( 2.9-23.4) |
| IRRI | HALL | 60.5 (NE-NE) | 60.0 (30.0- 92.3) | NE |
| HALL | IRRI | 7.1 (NE-NE) | 22.2 ( 8.3-38.1) | NE |
| IRRI | ELAT | NE | NE | 33.3 ( 0.0- 80.0) |
| ELAT | IRRI | NE | NE | 5.1 ( 0.0-13.3) |
| ANXI | APAT | 11.7 (NE-NE) | 20.8 ( 9.4- 32.6) | 14.0 ( 5.5- 24.5) |
| APAT | ANXI | 22.3 (NE-NE) | 35.7 (18.2-53.8) | 35.0 (15.8-57.7) |
| ANXI | SLEE | NA | NE | 6.4 ( 0.0- 13.3) |
| SLEE | ANXI | NA | NE | 15.0 ( 0.0-31.6) |
| ANXI | AGIT | 12.3 (NE-NE) | 25.9 (10.3- 43.3) | 11.8 ( 0.0- 30.8) |
| AGIT | ANXI | 11.0 (NE-NE) | 25.0 ( 9.4-43.3) | 10.0 ( 0.0-26.1) |
| ANXI | APPE | NA | NE | 6.4 ( 0.0- 14.6) |
| APPE | ANXI | NA | NE | 15.0 ( 0.0-31.9) |
| ANXI | DISI | 14.2 (NE-NE) | 45.5 (12.5- 75.0) | 8.3 ( 0.0- 28.6) |
| DISI | ANXI | 7.7 (NE-NE) | 17.9 ( 3.7-34.5) | 5.0 ( 0.0-16.0) |
| ANXI | ABER | 18.7 (NE-NE) | 30.8 (12.0- 50.0) | 7.1 ( 0.0- 22.2) |
| ABER | ANXI | 14.1 (NE-NE) | 28.6 (11.1-46.4) | 5.0 ( 0.0-16.7) |
| ANXI | PSYC | 13.0 (NE-NE) | 29.6 (12.0- 48.0) | 11.1 ( 0.0- 38.5) |
| PSYC | ANXI | 13.7 (NE-NE) | 28.6 (11.8-46.2) | 5.0 ( 0.0-18.2) |
| ANXI | DELU | 10.8 (NE-NE) | 30.4 (10.5- 52.2) | 11.1 ( 0.0- 38.5) |
| DELU | ANXI | 10.7 (NE-NE) | 25.0 ( 9.1-42.1) | 5.0 ( 0.0-18.2) |
| ANXI | HALL | 57.5 (NE-NE) | 50.0 (14.3- 81.8) | NE |
| HALL | ANXI | 7.3 (NE-NE) | 17.9 ( 4.0-34.6) | NE |

| | | | | |
|---|---|---|---|---|
| ANXI | ELAT | NE | NE | NE |
| ELAT | ANXI | NE | NE | NE |
| APAT | SLEE | NA | NE | 17.0 ( 6.9- 29.6) |
| SLEE | APAT | NA | NE | 16.0 ( 6.2-26.8) |
| APAT | AGIT | 44.9 (NE-NE) | 33.3 (16.1- 52.8) | 41.2 ( 16.7- 64.8) |
| AGIT | APAT | 21.0 (NE-NE) | 18.8 ( 8.3-30.4) | 14.0 ( 4.9-23.3) |
| APAT | APPE | NA | NE | 34.0 ( 21.3- 49.0) |
| APPE | APAT | NA | NE | 32.0 (19.6-45.5) |
| APAT | DISI | 55.3 (NE-NE) | 45.5 (14.2- 75.0) | 25.0 ( 0.0- 50.0) |
| DISI | APAT | 15.7 (NE-NE) | 10.4 ( 2.2-20.4) | 6.0 ( 0.0-14.0) |
| APAT | ABER | 55.3 (NE-NE) | 23.1 ( 8.3- 41.4) | 42.9 ( 14.3- 66.7) |
| ABER | APAT | 21.8 (NE-NE) | 12.5 ( 4.2-23.1) | 12.0 ( 3.4-22.2) |
| APAT | PSYC | 44.8 (NE-NE) | 29.6 (12.5- 48.3) | 33.3 ( 0.0- 70.0) |
| PSYC | APAT | 24.7 (NE-NE) | 16.7 ( 6.5-28.3) | 6.0 ( 0.0-13.6) |
| APAT | DELU | 44.4 (NE-NE) | 30.4 (12.5- 52.0) | 33.3 ( 0.0- 70.0) |
| DELU | APAT | 23.2 (NE-NE) | 14.6 ( 5.8-25.5) | 6.0 ( 0.0-13.6) |
| APAT | HALL | 36.0 (NE-NE) | 20.0 ( 0.0- 50.0) | NE |
| HALL | APAT | 2.4 (NE-NE) | 4.2 ( 0.0-10.3) | 2.0 ( 0.0- 6.9) |
| APAT | ELAT | NE | NE | 16.7 ( 0.0- 50.0) |
| ELAT | APAT | NE | NE | 2.0 ( 0.0- 6.7) |
| SLEE | AGIT | NA | NE | NE |
| AGIT | SLEE | NA | NE | NE |
| SLEE | APPE | NA | NE | 14.9 ( 5.3- 25.6) |
| APPE | SLEE | NA | NE | 14.9 ( 5.3-25.9) |
| SLEE | DISI | NA | NE | 16.7 ( 0.0- 41.7) |
| DISI | SLEE | NA | NE | 4.3 ( 0.0-10.9) |
| SLEE | ABER | NA | NE | 28.6 ( 7.7- 52.6) |
| ABER | SLEE | NA | NE | 8.5 ( 2.0-16.7) |
| SLEE | PSYC | NA | NE | NE |
| PSYC | SLEE | NA | NE | NE |
| SLEE | DELU | NA | NE | NE |
| DELU | SLEE | NA | NE | NE |
| SLEE | HALL | NA | NE | NE |
| HALL | SLEE | NA | NE | NE |
| SLEE | ELAT | NA | NE | 16.7 ( 0.0- 50.0) |

| | | | | |
|---|---|---|---|---|
| ELAT | SLEE | NA | NE | 2.1 ( 0.0- 7.3) |
| AGIT | APPE | NA | NE | 4.3 ( 0.0- 10.3) |
| APPE | AGIT | NA | NE | 11.8 ( 0.0-31.2) |
| AGIT | DISI | 52.8 (NE-NE) | 63.6 (35.7- 91.7) | 16.7 ( 0.0- 40.0) |
| DISI | AGIT | 32.1 (NE-NE) | 25.9 (10.7-43.8) | 11.8 ( 0.0-29.2) |
| AGIT | ABER | 42.2 (NE-NE) | 34.6 (17.2- 54.8) | 7.1 ( 0.0- 22.2) |
| ABER | AGIT | 35.7 (NE-NE) | 33.3 (17.4-52.0) | 5.9 ( 0.0-20.0) |
| AGIT | PSYC | 29.6 (NE-NE) | 33.3 (15.4- 50.0) | 33.3 ( 0.0- 71.4) |
| PSYC | AGIT | 34.9 (NE-NE) | 33.3 (15.6-52.6) | 17.6 ( 0.0-36.9) |
| AGIT | DELU | 31.3 (NE-NE) | 39.1 (20.0- 58.9) | 33.3 ( 0.0- 71.4) |
| DELU | AGIT | 34.9 (NE-NE) | 33.3 (15.6-52.6) | 17.6 ( 0.0-36.9) |
| AGIT | HALL | 34.7 (NE-NE) | 40.0 (10.0- 75.0) | NE |
| HALL | AGIT | 4.9 (NE-NE) | 14.8 ( 3.4-29.2) | NE |
| AGIT | ELAT | NE | NE | 16.7 ( 0.0- 50.0) |
| ELAT | AGIT | NE | NE | 5.9 ( 0.0-20.0) |
| APPE | DISI | NA | NE | 33.3 ( 7.7- 62.5) |
| DISI | APPE | NA | NE | 8.5 ( 1.9-16.7) |
| APPE | ABER | NA | NE | 35.7 ( 10.5- 63.6) |
| ABER | APPE | NA | NE | 10.6 ( 2.3-20.4) |
| APPE | PSYC | NA | NE | 33.3 ( 0.0- 66.7) |
| PSYC | APPE | NA | NE | 6.4 ( 0.0-14.3) |
| APPE | DELU | NA | NE | 33.3 ( 0.0- 66.7) |
| DELU | APPE | NA | NE | 6.4 ( 0.0-14.3) |
| APPE | HALL | NA | NE | NE |
| HALL | APPE | NA | NE | 2.1 ( 0.0- 7.5) |
| APPE | ELAT | NA | NE | 16.7 ( 0.0- 50.8) |
| ELAT | APPE | NA | NE | 2.1 ( 0.0- 7.1) |
| DISI | ABER | 33.2 (NE-NE) | 19.2 ( 4.8- 36.4) | 14.3 ( 0.0- 36.4) |
| ABER | DISI | 46.1 (NE-NE) | 45.5 (15.4-75.0) | 16.7 ( 0.0-40.0) |
| DISI | PSYC | 28.1 (NE-NE) | 22.2 ( 6.7- 40.0) | 33.3 ( 0.0- 66.7) |
| PSYC | DISI | 54.5 (NE-NE) | 54.5 (22.2-84.6) | 25.0 ( 0.0-50.0) |
| DISI | DELU | 26.8 (NE-NE) | 21.7 ( 5.6- 40.6) | 33.3 ( 0.0- 66.7) |
| DELU | DISI | 49.1 (NE-NE) | 45.5 (16.6-75.0) | 25.0 ( 0.0-50.0) |
| DISI | HALL | 39.3 (NE-NE) | 30.0 ( 0.0- 62.5) | NE |
| HALL | DISI | 9.1 (NE-NE) | 27.3 ( 0.0-57.1) | NE |

| | | | | |
|---|---|---|---|---|
| DISI | ELAT | NE | NE | 50.0 ( 0.0-100.0) |
| ELAT | DISI | NE | NE | 25.0 ( 0.0-50.0) |
| ABER | PSYC | 43.9 (NE-NE) | 33.3 (16.0- 51.4) | 22.2 ( 0.0- 55.6) |
| PSYC | ABER | 61.4 (NE-NE) | 34.6 (16.7-53.6) | 14.3 ( 0.0-37.5) |
| ABER | DELU | 45.3 (NE-NE) | 34.8 (15.8- 54.6) | 22.2 ( 0.0- 55.6) |
| DELU | ABER | 59.8 (NE-NE) | 30.8 (13.6-50.0) | 14.3 ( 0.0-37.5) |
| ABER | HALL | 25.5 (NE-NE) | 30.0 ( 0.0- 60.0) | NE |
| HALL | ABER | 4.3 (NE-NE) | 11.5 ( 0.0-25.0) | 7.1 ( 0.0-25.0) |
| ABER | ELAT | NE | NE | 33.3 ( 0.0- 80.0) |
| ELAT | ABER | NE | NE | 14.3 ( 0.0-36.4) |
| PSYC | ELAT | NE | NE | 33.3 ( 0.0- 75.0) |
| ELAT | PSYC | NE | NE | 22.2 ( 0.0-50.1) |
| DELU | HALL | 53.0 (NE-NE) | 60.0 (25.0- 90.0) | NE |
| HALL | DELU | 6.7 (NE-NE) | 26.1 ( 8.0-46.4) | 11.1 ( 0.0-37.5) |
| DELU | ELAT | NE | NE | 33.3 ( 0.0- 75.0) |
| ELAT | DELU | NE | NE | 22.2 ( 0.0-50.1) |
| HALL | ELAT | NE | NE | NE |
| ELAT | HALL | NE | NE | NE |

S1|S2, conditional prevalence of symptom S1 given presence of symptom S2.
Displayed are conditional prevalence (%) and 95% confidence intervals. Survey estimates for ADAMS are based on survey population weights, confidence intervals are not estimated.
NA, Not applicable; NE, Not estimable.
ABER, Aberrant motor behavior; AGIT, Agitation/aggression; ANXI, Anxiety; APAT, Apathy/indifference; APPE, Appetite/eating changes; DELU, Delusions; DEPR, Depression/dysphoria; DISI, Disinhibition; ELAT, Elation/euphoria; HALU, Hallucinations; IRRI, Irritability/lability; PSYC, Psychotic symptoms (hallucinations and/or delusions); SLEE, Nighttime behavioral disturbances.

Supplemental Table 29. Conditional prevalence S1|S2 of neuropsychiatric symptoms with the NPI product score ≥ 4 per MMSE stratum (ADAMS and ADNI studies only)

| S1 | S2 | ADAMS, survey | | ADAMS | | ADNI | |
|---|---|---|---|---|---|---|---|
| | | MMSE >26 | MMSE ≤26 | MMSE >26 | MMSE ≤26 | MMSE >26 | MMSE ≤26 |
| DEPR | IRRI | NE | 42.0 (NE-NE) | NE | 34.6 (16.1- 53.2) | 4.8 ( 0.0- 15.8) | 38.9 ( 13.3- 63.6) |
| IRRI | DEPR | NE | 28.3 (NE-NE) | NE | 28.1 (12.1-44.0) | 14.3 (0.0-50.0) | 50.0 (21.0- 76.9) |
| DEPR | ANXI | 38.3 (NE-NE) | 27.9 (NE-NE) | 25.0 ( 0.0-100.0) | 33.3 (13.6- 53.9) | 9.1 ( 0.0- 30.0) | 33.3 ( 0.0- 66.7) |
| ANXI | DEPR | 51.3 (NE-NE) | 11.6 (NE-NE) | 25.0 (0.0-100.0) | 25.0 ( 9.7-40.6) | 14.3 (0.0-50.0) | 21.4 ( 0.0- 45.5) |
| DEPR | APAT | NE | 35.0 (NE-NE) | NE | 35.7 (21.3- 51.4) | 11.1 ( 0.0- 28.0) | 18.8 ( 6.2- 33.4) |
| APAT | DEPR | NE | 33.1 (NE-NE) | NE | 46.9 (30.0-65.1) | 28.6 (0.0-71.4) | 42.9 (16.7- 70.0) |
| DEPR | SLEE | NA | NA | NE | NE | 8.0 ( 0.0- 20.0) | 22.7 ( 5.3- 42.3) |
| SLEE | DEPR | NA | NA | NE | NE | 28.6 (0.0-66.8) | 35.7 ( 9.1- 62.5) |
| DEPR | AGIT | NE | 38.2 (NE-NE) | NE | 26.9 (10.5- 45.0) | NE | 10.0 ( 0.0- 30.8) |
| AGIT | DEPR | 3.4 (NE-NE) | 26.0 (NE-NE) | 25.0 (0.0-100.0) | 21.9 ( 7.9-36.4) | NE | 7.1 ( 0.0- 23.8) |
| DEPR | APPE | NA | NA | NE | NE | 20.0 ( 0.0- 42.9) | 12.5 ( 2.8- 25.0) |
| APPE | DEPR | NA | NA | NE | NE | 42.9 (0.0-83.3) | 28.6 ( 6.7- 55.6) |
| DEPR | DISI | NE | 48.7 (NE-NE) | NE | 36.4 ( 8.3- 66.7) | 12.5 ( 0.0- 44.4) | 25.0 ( 0.0-100.0) |
| DISI | DEPR | NE | 20.4 (NE-NE) | NE | 12.5 ( 2.6-25.7) | 14.3 (0.0-50.0) | 7.1 ( 0.0- 23.1) |
| DEPR | ABER | NE | 27.8 (NE-NE) | NE | 11.5 ( 0.0- 25.9) | 20.0 ( 0.0- 66.7) | NE |
| ABER | DEPR | NE | 16.2 (NE-NE) | NE | 9.4 ( 0.0-20.7) | 14.3 (0.0-50.0) | NE |
| DEPR | PSYC | NE | 33.8 (NE-NE) | NE | 26.9 ( 9.7- 44.8) | NE | 16.7 ( 0.0- 50.0) |
| PSYC | DEPR | NE | 22.6 (NE-NE) | NE | 21.9 ( 7.7-36.4) | NE | 7.1 ( 0.0- 23.1) |
| DEPR | DELU | NE | 32.6 (NE-NE) | NE | 27.3 (10.0- 46.7) | NE | 16.7 ( 0.0- 50.0) |
| DELU | DEPR | NE | 20.3 (NE-NE) | NE | 18.8 ( 5.9-33.3) | NE | 7.1 ( 0.0- 23.1) |
| DEPR | HALL | NE | 40.9 (NE-NE) | NE | 30.0 ( 0.0- 60.0) | NE | NE |
| HALL | DEPR | NE | 4.0 (NE-NE) | NE | 9.4 ( 0.0-21.2) | NE | NE |
| DEPR | ELAT | NE | NE | NE | NE | NE | 25.0 ( 0.0-100.0) |
| ELAT | DEPR | NE | NE | NE | NE | NE | 7.1 ( 0.0- 23.1) |
| IRRI | ANXI | NE | 25.4 (NE-NE) | NE | 37.5 (17.4- 58.8) | 9.1 ( 0.0- 30.0) | 22.2 ( 0.0- 50.1) |
| ANXI | IRRI | NE | 15.6 (NE-NE) | NE | 34.6 (16.7-54.2) | 4.8 (0.0-15.8) | 11.1 ( 0.0- 28.6) |
| IRRI | APAT | NE | 29.5 (NE-NE) | NE | 19.0 ( 7.5- 32.4) | 27.8 ( 7.7- 50.0) | 15.6 ( 3.7- 29.2) |
| APAT | IRRI | NE | 41.3 (NE-NE) | NE | 30.8 (12.5-50.0) | 23.8 (5.6-43.8) | 27.8 ( 7.7- 50.0) |

| | | | | | | | |
|---|---|---|---|---|---|---|---|
| IRRI | SLEE | NA | NA | NE | NE | 8.0 ( 0.0- 20.8) | 13.6 ( 0.0- 31.6) |
| SLEE | IRRI | NA | NA | NE | NE | 9.5 (0.0-25.0) | 16.7 ( 0.0- 36.4) |
| IRRI | AGIT | NE | 78.9 (NE-NE) | NE | 61.5 (42.9- 79.4) | 57.1 ( 16.7-100.0) | 20.0 ( 0.0- 50.0) |
| AGIT | IRRI | NE | 79.5 (NE-NE) | NE | 61.5 (42.9-81.0) | 19.0 (4.3-38.1) | 11.1 ( 0.0- 28.6) |
| IRRI | APPE | NA | NA | NE | NE | 13.3 ( 0.0- 33.3) | 18.8 ( 6.5- 32.4) |
| APPE | IRRI | NA | NA | NE | NE | 9.5 (0.0-23.8) | 33.3 (12.5- 56.2) |
| IRRI | DISI | NE | 58.9 (NE-NE) | NE | 72.7 (42.9-100.0) | 37.5 ( 0.0- 75.0) | 75.0 ( 20.0-100.0) |
| DISI | IRRI | NE | 36.5 (NE-NE) | NE | 30.8 (14.3-50.0) | 14.3 (0.0-30.8) | 16.7 ( 0.0- 35.7) |
| IRRI | ABER | NE | 35.2 (NE-NE) | NE | 23.1 ( 7.7- 42.9) | NE | 22.2 ( 0.0- 55.6) |
| ABER | IRRI | NE | 30.4 (NE-NE) | NE | 23.1 ( 8.0-40.7) | NE | 11.1 ( 0.0- 27.8) |
| IRRI | PSYC | NE | 39.9 (NE-NE) | NE | 42.3 (24.0- 61.9) | 66.7 ( 0.0-100.0) | 50.0 ( 0.0-100.0) |
| PSYC | IRRI | NE | 39.4 (NE-NE) | NE | 42.3 (25.0-62.1) | 9.5 (0.0-23.8) | 16.7 ( 0.0- 35.7) |
| IRRI | DELU | NE | 37.9 (NE-NE) | NE | 40.9 (21.0- 63.6) | 66.7 ( 0.0-100.0) | 50.0 ( 0.0-100.0) |
| DELU | IRRI | NE | 34.9 (NE-NE) | NE | 34.6 (17.4-54.2) | 9.5 (0.0-23.8) | 16.7 ( 0.0- 35.7) |
| IRRI | HALL | NE | 60.5 (NE-NE) | NE | 60.0 (30.0- 92.3) | NE | NE |
| HALL | IRRI | NE | 8.7 (NE-NE) | NE | 23.1 ( 8.6-40.0) | NE | NE |
| IRRI | ELAT | NE | NE | NE | NE | NE | 50.0 ( 0.0-100.0) |
| ELAT | IRRI | NE | NE | NE | NE | NE | 11.1 ( 0.0- 28.6) |
| ANXI | APAT | NE | 18.1 (NE-NE) | NE | 23.8 (10.8- 36.8) | 5.6 ( 0.0- 18.8) | 18.8 ( 6.1- 34.5) |
| APAT | ANXI | NE | 41.3 (NE-NE) | NE | 41.7 (22.2-61.1) | 9.1 (0.0-30.0) | 66.7 (30.0-100.0) |
| ANXI | SLEE | NA | NA | NE | NE | 8.0 ( 0.0- 19.2) | 4.5 ( 0.0- 14.7) |
| SLEE | ANXI | NA | NA | NE | NE | 18.2 (0.0-45.5) | 11.1 ( 0.0- 33.4) |
| ANXI | AGIT | NE | 12.5 (NE-NE) | NE | 26.9 (10.7- 45.5) | NE | 20.0 ( 0.0- 50.0) |
| AGIT | ANXI | NE | 20.3 (NE-NE) | NE | 29.2 (11.5-50.0) | NE | 22.2 ( 0.0- 50.0) |
| ANXI | APPE | NA | NA | NE | NE | 6.7 ( 0.0- 22.2) | 6.2 ( 0.0- 16.0) |
| APPE | ANXI | NA | NA | NE | NE | 9.1 (0.0-30.0) | 22.2 ( 0.0- 58.3) |
| ANXI | DISI | NE | 14.2 (NE-NE) | NE | 45.5 (12.5- 75.0) | 12.5 ( 0.0- 44.4) | NE |
| DISI | ANXI | NE | 14.3 (NE-NE) | NE | 20.8 ( 4.5-40.0) | 9.1 (0.0-30.0) | NE |
| ANXI | ABER | NE | 18.7 (NE-NE) | NE | 30.8 (12.0- 50.0) | NE | 11.1 ( 0.0- 37.5) |
| ABER | ANXI | NE | 26.2 (NE-NE) | NE | 33.3 (14.3-52.4) | NE | 11.1 ( 0.0- 33.3) |
| ANXI | PSYC | NE | 15.8 (NE-NE) | NE | 30.8 (13.0- 50.0) | NE | 16.7 ( 0.0- 55.6) |
| PSYC | ANXI | NE | 25.4 (NE-NE) | NE | 33.3 (13.8-52.6) | NE | 11.1 ( 0.0- 37.5) |
| ANXI | DELU | NE | 13.3 (NE-NE) | NE | 31.8 (11.7- 53.0) | NE | 16.7 ( 0.0- 55.6) |
| DELU | ANXI | NE | 19.9 (NE-NE) | NE | 29.2 (11.1-47.4) | NE | 11.1 ( 0.0- 37.5) |
| ANXI | HALL | NE | 57.5 (NE-NE) | NE | 50.0 (14.3- 81.8) | NE | NE |

| | | | | | | | |
|---|---|---|---|---|---|---|---|
| HALL | ANXI | NE | 13.4 (NE-NE) | NE | 20.8 ( 4.5-39.2) | NE | NE |
| ANXI | ELAT | NE | NE | NE | NE | NE | NE |
| ELAT | ANXI | NE | NE | NE | NE | NE | NE |
| APAT | SLEE | NA | NA | NE | NE | 12.0 ( 0.0- 25.9) | 22.7 ( 5.3- 41.2) |
| SLEE | APAT | NA | NA | NE | NE | 16.7 (0.0-35.0) | 15.6 ( 3.6- 29.4) |
| APAT | AGIT | NE | 45.5 (NE-NE) | NE | 34.6 (17.4- 55.0) | 42.9 ( 0.0- 83.3) | 40.0 ( 10.0- 72.8) |
| AGIT | APAT | NE | 32.6 (NE-NE) | NE | 21.4 (10.0-33.9) | 16.7 (0.0-36.0) | 12.5 ( 2.7- 24.2) |
| APAT | APPE | NA | NA | NE | NE | 6.7 ( 0.0- 22.2) | 46.9 ( 30.8- 65.0) |
| APPE | APAT | NA | NA | NE | NE | 5.6 (0.0-18.8) | 46.9 (30.8- 65.7) |
| APAT | DISI | NE | 55.3 (NE-NE) | NE | 45.5 (14.2- 75.0) | 37.5 ( 0.0- 75.0) | NE |
| DISI | APAT | NE | 24.4 (NE-NE) | NE | 11.9 ( 2.4-22.5) | 16.7 (0.0-37.5) | NE |
| APAT | ABER | NE | 55.3 (NE-NE) | NE | 23.1 ( 8.3- 41.4) | 20.0 ( 0.0- 66.7) | 55.6 ( 20.0- 90.0) |
| ABER | APAT | NE | 34.0 (NE-NE) | NE | 14.3 ( 4.9-26.7) | 5.6 (0.0-18.8) | 15.6 ( 3.4- 29.6) |
| APAT | PSYC | NE | 54.6 (NE-NE) | NE | 30.8 (13.3- 50.0) | NE | 50.0 ( 0.0-100.0) |
| PSYC | APAT | NE | 38.5 (NE-NE) | NE | 19.0 ( 7.4-31.1) | NE | 9.4 ( 0.0- 21.1) |
| APAT | DELU | NE | 54.9 (NE-NE) | NE | 31.8 (13.3- 54.2) | NE | 50.0 ( 0.0-100.0) |
| DELU | APAT | NE | 36.0 (NE-NE) | NE | 16.7 ( 6.4-28.6) | NE | 9.4 ( 0.0- 21.1) |
| APAT | HALL | NE | 36.0 (NE-NE) | NE | 20.0 ( 0.0- 50.0) | NE | NE |
| HALL | APAT | NE | 3.7 (NE-NE) | NE | 4.8 ( 0.0-11.6) | NE | 3.1 ( 0.0- 10.7) |
| APAT | ELAT | NE | NE | NE | NE | 50.0 ( 0.0-100.0) | NE |
| ELAT | APAT | NE | NE | NE | NE | 5.6 (0.0-19.1) | NE |
| SLEE | AGIT | NA | NA | NE | NE | NE | NE |
| AGIT | SLEE | NA | NA | NE | NE | NE | NE |
| SLEE | APPE | NA | NA | NE | NE | 13.3 ( 0.0- 33.3) | 15.6 ( 3.6- 29.2) |
| APPE | SLEE | NA | NA | NE | NE | 8.0 (0.0-20.0) | 22.7 ( 5.6- 40.9) |
| SLEE | DISI | NA | NA | NE | NE | 25.0 ( 0.0- 60.0) | NE |
| DISI | SLEE | NA | NA | NE | NE | 8.0 (0.0-20.7) | NE |
| SLEE | ABER | NA | NA | NE | NE | 20.0 ( 0.0- 66.7) | 33.3 ( 0.0- 66.7) |
| ABER | SLEE | NA | NA | NE | NE | 4.0 (0.0-14.3) | 13.6 ( 0.0- 28.6) |
| SLEE | PSYC | NA | NA | NE | NE | NE | NE |
| PSYC | SLEE | NA | NA | NE | NE | NE | NE |
| SLEE | DELU | NA | NA | NE | NE | NE | NE |
| DELU | SLEE | NA | NA | NE | NE | NE | NE |
| SLEE | HALL | NA | NA | NE | NE | NE | NE |
| HALL | SLEE | NA | NA | NE | NE | NE | NE |

| | | | | | | | |
|---|---|---|---|---|---|---|---|
| SLEE | ELAT | NA | NA | NE | NE | NE | 25.0 ( 0.0-100.0) |
| ELAT | SLEE | NA | NA | NE | NE | NE | 4.5 ( 0.0- 15.8) |
| AGIT | APPE | NA | NA | NE | NE | NE | 6.2 ( 0.0- 15.4) |
| APPE | AGIT | NA | NA | NE | NE | NE | 20.0 ( 0.0- 50.0) |
| AGIT | DISI | NE | 52.8 (NE-NE) | NE | 63.6 (35.7- 91.7) | 25.0 ( 0.0- 60.0) | NE |
| DISI | AGIT | NE | 32.5 (NE-NE) | NE | 26.9 (11.1-45.5) | 28.6 (0.0-66.7) | NE |
| AGIT | ABER | NE | 42.2 (NE-NE) | NE | 34.6 (17.2- 54.8) | NE | 11.1 ( 0.0- 37.5) |
| ABER | AGIT | NE | 36.1 (NE-NE) | NE | 34.6 (18.2-53.6) | NE | 10.0 ( 0.0- 33.3) |
| AGIT | PSYC | NE | 36.0 (NE-NE) | NE | 34.6 (16.7- 53.6) | NE | 50.0 ( 0.0-100.0) |
| PSYC | AGIT | NE | 35.4 (NE-NE) | NE | 34.6 (16.7-53.9) | NE | 30.0 ( 0.0- 60.0) |
| AGIT | DELU | NE | 38.7 (NE-NE) | NE | 40.9 (21.1- 61.9) | NE | 50.0 ( 0.0-100.0) |
| DELU | AGIT | NE | 35.4 (NE-NE) | NE | 34.6 (16.7-53.9) | NE | 30.0 ( 0.0- 60.0) |
| AGIT | HALL | NE | 34.7 (NE-NE) | NE | 40.0 (10.0- 75.0) | NE | NE |
| HALL | AGIT | NE | 5.0 (NE-NE) | NE | 15.4 ( 3.4-30.4) | NE | NE |
| AGIT | ELAT | NE | NE | NE | NE | NE | 25.0 ( 0.0-100.0) |
| ELAT | AGIT | NE | NE | NE | NE | NE | 10.0 ( 0.0- 33.3) |
| APPE | DISI | NA | NA | NE | NE | 37.5 ( 0.0- 75.0) | 25.0 ( 0.0-100.0) |
| DISI | APPE | NA | NA | NE | NE | 20.0 (0.0-42.9) | 3.1 ( 0.0- 10.7) |
| APPE | ABER | NA | NA | NE | NE | 40.0 ( 0.0-100.0) | 33.3 ( 0.0- 70.1) |
| ABER | APPE | NA | NA | NE | NE | 13.3 (0.0-33.3) | 9.4 ( 0.0- 21.2) |
| APPE | PSYC | NA | NA | NE | NE | 33.3 ( 0.0-100.0) | 33.3 ( 0.0- 80.0) |
| PSYC | APPE | NA | NA | NE | NE | 6.7 (0.0-23.1) | 6.2 ( 0.0- 16.1) |
| APPE | DELU | NA | NA | NE | NE | 33.3 ( 0.0-100.0) | 33.3 ( 0.0- 80.0) |
| DELU | APPE | NA | NA | NE | NE | 6.7 (0.0-23.1) | 6.2 ( 0.0- 16.1) |
| APPE | HALL | NA | NA | NE | NE | NE | NE |
| HALL | APPE | NA | NA | NE | NE | NE | 3.1 ( 0.0- 11.1) |
| APPE | ELAT | NA | NA | NE | NE | 50.0 ( 0.0-100.0) | NE |
| ELAT | APPE | NA | NA | NE | NE | 6.7 (0.0-22.2) | NE |
| DISI | ABER | NE | 33.2 (NE-NE) | NE | 19.2 ( 4.8- 36.4) | 20.0 ( 0.0- 66.7) | 11.1 ( 0.0- 37.5) |
| ABER | DISI | NE | 46.1 (NE-NE) | NE | 45.5 (15.4-75.0) | 12.5 (0.0-42.9) | 25.0 ( 0.0-100.0) |
| DISI | PSYC | NE | 34.2 (NE-NE) | NE | 23.1 ( 7.1- 40.6) | 33.3 ( 0.0-100.0) | 33.3 ( 0.0- 80.0) |
| PSYC | DISI | NE | 54.5 (NE-NE) | NE | 54.5 (22.2-84.6) | 12.5 (0.0-40.0) | 50.0 ( 0.0-100.0) |
| DISI | DELU | NE | 33.1 (NE-NE) | NE | 22.7 ( 5.9- 41.7) | 33.3 ( 0.0-100.0) | 33.3 ( 0.0- 80.0) |
| DELU | DISI | NE | 49.1 (NE-NE) | NE | 45.5 (16.6-75.0) | 12.5 (0.0-40.0) | 50.0 ( 0.0-100.0) |
| DISI | HALL | NE | 39.3 (NE-NE) | NE | 30.0 ( 0.0- 62.5) | NE | NE |

| | | | | | | | |
|---|---|---|---|---|---|---|---|
| HALL | DISI | NE | 9.1 (NE-NE) | NE | 27.3 ( 0.0-57.1) | NE | NE |
| DISI | ELAT | NE | NE | NE | NE | NE | 25.0 ( 0.0-100.0) |
| ELAT | DISI | NE | NE | NE | NE | 25.0 (0.0-60.0) | 25.0 ( 0.0-100.0) |
| ABER | PSYC | NE | 53.6 (NE-NE) | NE | 34.6 (16.7- 53.9) | NE | 33.3 ( 0.0- 84.0) |
| PSYC | ABER | NE | 61.4 (NE-NE) | NE | 34.6 (16.7-53.6) | NE | 22.2 ( 0.0- 57.1) |
| ABER | DELU | NE | 56.0 (NE-NE) | NE | 36.4 (16.7- 57.9) | NE | 33.3 ( 0.0- 84.0) |
| DELU | ABER | NE | 59.8 (NE-NE) | NE | 30.8 (13.6-50.0) | NE | 22.2 ( 0.0- 57.1) |
| ABER | HALL | NE | 25.5 (NE-NE) | NE | 30.0 ( 0.0- 60.0) | NE | NE |
| HALL | ABER | NE | 4.3 (NE-NE) | NE | 11.5 ( 0.0-25.0) | NE | 11.1 ( 0.0- 37.5) |
| ABER | ELAT | NE | NE | NE | NE | 50.0 ( 0.0-100.0) | 25.0 ( 0.0-100.0) |
| ELAT | ABER | NE | NE | NE | NE | 20.0 (0.0-66.7) | 11.1 ( 0.0- 37.5) |
| PSYC | ELAT | NE | NE | NE | NE | NE | 50.0 ( 0.0-100.0) |
| ELAT | PSYC | NE | NE | NE | NE | NE | 33.3 ( 0.0- 80.0) |
| DELU | HALL | NE | 53.0 (NE-NE) | NE | 60.0 (25.0- 90.0) | NE | NE |
| HALL | DELU | NE | 8.3 (NE-NE) | NE | 27.3 ( 8.3-50.0) | NE | 16.7 ( 0.0- 57.1) |
| DELU | ELAT | NE | NE | NE | NE | NE | 50.0 ( 0.0-100.0) |
| ELAT | DELU | NE | NE | NE | NE | NE | 33.3 ( 0.0- 80.0) |
| HALL | ELAT | NE | NE | NE | NE | NE | NE |
| ELAT | HALL | NE | NE | NE | NE | NE | NE |

S1|S2, conditional prevalence of symptom S1 given presence of symptom S2.

Displayed are conditional prevalence (%) and 95% confidence intervals. Survey estimates for ADAMS are based on survey population weights, confidence intervals are not estimated.

NA, Not applicable; NE, Not estimable.

ABER, Aberrant motor behavior; AGIT, Agitation/aggression; ANXI, Anxiety; APAT, Apathy/indifference; APPE, Appetite/eating changes; DELU, Delusions; DEPR, Depression/dysphoria; DISI, Disinhibition; ELAT, Elation/euphoria; HALU, Hallucinations; IRRI, Irritability/lability; PSYC, Psychotic symptoms (hallucinations and/or delusions); SLEE, Nighttime behavioral disturbances.

### Supplement section 7. "Expected" vs "observed" prevalence of neuropsychiatric symptom pairs (hypothetical example)

Let's say, we have two independent samples and evaluate two symptoms, A and B, that are statistically independent random events.

In one sample of 1,000 subjects, prevalence of symptoms A and B is 40% and 10%, respectively, so that there are 400 subjects with symptom A and 100 subjects with symptom B.

In another sample of 10,000 subjects, prevalence of symptoms A and B is 1% and 2%, respectively, so that there are 100 subjects with symptom A and 200 subjects with symptom B.

As symptoms A and B are statistically independent random events, both symptoms will be presented in 4% (40% x 10%) of subjects in the first sample and in 0.02% (1% x 2%) of subjects in the second sample, or in 40 and 2 subjects, respectively.

Thus, across both samples combined, there are 11,000 subjects, symptom A is present in 500 subjects (400 + 100; hence, prevalence of 4.5%), symptom B is present in 300 subjects (100 + 200; hence, prevalence of 2.7%), and both symptoms are present in 42 subjects (40 + 2; hence, prevalence of **0.38**%). The latter "observed" estimate of the symptom pair prevalence is different from a value that one receives by multiplying prevalence of individual symptoms (4.5% x 2.7% = **0.12**%).

Supplemental Table 30. Example for "expected" vs "observed" prevalence

| | Sample 1 | Sample 2 | Samples 1 + 2 |
|---|---|---|---|
| Total N | 1,000 | 10,000 | 11,000 |
| Symptom A prevalence | 40%<br>n=400 | 1%<br>(n=100) | 4.5%<br>(n=500) |
| Symptom B prevalence | 10%<br>n=100 | 2%<br>(n=200) | 2.7%<br>(n=300) |
| "Expected" prevalence of the symptom pair (assuming statistical independence of A and B) | 4%<br>(40% x 10%)<br>n=40<br>(4% x 1,000) | 0.02%<br>(1% x 2%)<br>N=2<br>(0.02% x 10,000) | **0.12%**<br>(4.5% x 2.7%) |
| "Observed" prevalence of the symptom pair | | | **0.38%**<br>(40+2)*100 / 11,000 |
